# Long-term manifestations and modifiers of prevalence estimates of the post-COVID-19 syndrome: A systematic review and meta-analysis

**DOI:** 10.1101/2021.10.17.21265123

**Authors:** Daniel De-la-Rosa-Martinez, Marco Antonio Delaye-Martínez, Omar Yaxmehen Bello-Chavolla, Alejandro Sicilia-Andrade, Isaac David Juárez-Cruz, Carlos A. Fermín-Martínez, Alejandro Márquez-Salinas, Enrique Cañedo-Guerra, Luisa Fernández-Chirino, Mónica Martínez-Gutiérrez, Daniel Eduardo Sandoval-Colin, Diana Vilar-Compte

## Abstract

**Background:** Post-acute COVID-19 syndrome (PACS) is a multi-system disease comprising persistent symptomatology after the acute phase of infection. Long-term PACS effects significantly impact patient outcomes, but their incidence remains uncharacterized due to high heterogeneity between studies. Therefore, we aimed to summarize published data on PACS, characterizing the clinical presentation, prevalence, and modifiers of prevalence estimates.

**Method:** In this systematic review and meta-analysis, we research MEDLINE for original studies published from January 1st, 2020, to January 31st, 2021, that reported proportions of PACS manifestations. Studies were eligible for inclusion if they included patients aged ≥18 years with confirmed COVID-19 by RT-PCR or antigen testing and a minimum follow-up of 21 days. The prevalence of individual manifestations across studies was pooled using random-effects meta-analysis. For evaluating determinants of heterogeneity, meta-regression analysis was performed. This study was registered in PROSPERO (CRD42019125025).

**Results:** After screening 1,235 studies, we included 29 reports for analysis. Twenty-seven meta-analyses were performed, and 61 long-term manifestations were described. The pooled prevalence of PACS was 56% (95%CI 45-66%), with the most common manifestations being diminished health status, fatigue, asthenia, dyspnea, myalgias, hyposmia and dysgeusia. Most of the included studies presented high heterogeneity. After conducting the meta-regression analysis, we identified that age, gender, number of comorbidities, and reported symptoms significantly modify the prevalence estimation of PACS long-term manifestations.

**Conclusion:** PACS is inconsistently reported between studies, and population characteristics influence the prevalence estimates due to high heterogeneity. A systematized approach for the study of PACS is needed to characterize its impact adequately.

**Funding:** none

## INTRODUCTION

Post-acute COVID-19 syndrome (PACS), also known as Long Covid, is a multi-system disease that occurs after the acute phase of COVID-19. Most patients with COVID-19 develop a mild acute illness lasting 15 to 20 days [1, 2]; however, a variable proportion of patients persist with symptoms beyond four weeks from symptom onset. According to the World Health Organization, PACS “occurs in individuals with a history of probable or confirmed SARS CoV-2 infection, usually three months from the onset of COVID-19 with symptoms that last for at least two months and cannot be explained by an alternative diagnosis” [3]. Some studies have reported a prevalence of 10-33% of patients who undergo prolonged illness lasting from several weeks to months after acute COVID-19. Although these manifestations vary widely, the most common symptoms described are fatigue, headache, cough, breathlessness, joint pain, muscle and chest pain, weakness, gastrointestinal upset, rashes, chills, and olfactory and gustatory dysfunction. In addition, some neurocognitive impairments such as depression, anxiety, and post-traumatic stress syndrome have been highly reported.

Despite this growing available information, there is still uncertainty regarding the number and variety of the long-term effects attributable to PACS. Moreover, in agreement with previous observations in infections caused by Chikungunya, Ebola, Severe Acute Respiratory Syndrome Coronavirus (SARS-CoV-1), and Middle East respiratory syndrome (MERS) virus, these long-term manifestations could last for years after the acute infection [4–7]. Clinical characterization of PACS is still ongoing, and some risk factors have been preliminarily identified. Patients who developed severe COVID-19 are more likely to present long-term effects however, patients with an initial mild course are not free from developing new or persistent symptoms. In addition, some reports have placed women, patients admitted to the intensive care unit, and those with specific comorbidities as higher risk groups for developing long-term effects.

Recent data reporting the long-term effects of COVID-19 is showing a new landscape in SARS-CoV-2 infection. In addition, the proportion of patients with sequelae after COVID-19 who require specialized medical attention, rehabilitation programs, and absence from work is increasing, having profound medical, social, and economic implications which urgently need to be addressed by governments and other stakeholders. Here, we conducted a systematic review and metanalysis to evaluate data on the persistence of symptoms after acute COVID-19, characterizing the clinical presentation and modifiers of prevalence estimates in PACS.

## METHODS

### Search criteria

A sensitive reference search was conducted using MEDLINE (via PubMed) to include manuscripts from January 1st, 2020, to January 31st, 2021. A preliminary search was made in PubMed, looking for relevant studies reporting the clinical status of patients recovered from COVID-19 several weeks after the acute illness. We focused on keywords included in each article to establish a general landscape of how “persistent symptoms” were described. Keywords included in our search were: post-discharge symptoms; long-COVID; recovery; post-COVID syndrome; sequels; long-term consequences; persistent symptoms; post-COVID; SARS-CoV-2 and COVID-19. Post-acute COVID-19 syndrome (PACS) was not included as a keyword, as it has recently been described. With the selected keywords, we designed a search algorithm in PubMed that combined Boolean operators (AND/OR), search filters for language (English and Spanish), and year of publication (2020-2021).

### Screening and data extraction

Three authors (DDRM, MADM, IDJC) screened article titles and abstracts of the initial database search to identify those appropriate for complete text review. Pairs reviewed the full text of identified articles according to the following selection criteria:

1. Patients with confirmed COVID-19 by RT-PCR or rapid antigen test
2. Adult population (>18 years old)
3. Acute symptomatic illness during SARS-CoV-2 infection
4. A period of follow-up ≥ three weeks (21 days)
5. Reported the proportion of the specific persistent manifestations

Articles with confirmed reinfection cases were excluded from the analysis. For each included manuscript, data were extracted and summarized in a standardized database by reviewers.

### Quality assessment

Joanna Briggs Institute critical appraisal tools were used to evaluate quality of the studies and PRISMA check list was used for reporting the findings. A third reviewer solved any disagreement if the initial reviewers reported discordant results. This review was prospectively registered in the PROSPERO database (Project number CRD42021243541).

### Variables

We included demographic, clinical characteristics, and outcomes likely related to PACS in patients with previous COVID-19. Clinical signs and symptoms related to PACS were reported as proportions and described individually and classified according to the organ systems involved. Due to the nature of the pandemic and its rapid evolution, long-term manifestations associated with COVID-19 are not universally recognized or previously defined. Recently, the term PACS has been used as a reference in the literature. For this study and in agreement with the evidence of active viral infection and infectivity, PACS was defined as the persistence of symptoms ≥21 days after SARS-CoV-2 infection diagnosis [8].

### Statistical analysis

We calculated the pooled prevalence of symptoms for studies in which 1) the total number of patients with COVID-19 and the total number of those with persistent symptoms were reported by the authors and 2) at least two or more studies reported the specific long-term manifestation. We used a random-effects meta-analysis for calculating pooled prevalence estimates using the Hartung-Knapp-Sidik-Jonkman variance estimator, and the I² statistic test was calculated to quantify the heterogeneity of the studies [9]. I² values to establish the degree of heterogeneity were >75% for high; 50-75% for substantial; 25-50% for moderate and <25% for low heterogeneity. Forest plots were made to visualize the pooled prevalence of each long-term manifestation.

In order to diminish heterogeneity and improve meta-analytic pooling, we performed an outlier removing analysis as follows: We identified a study as an outlier if 1) its confidence interval did not overlap with the confidence interval of the pooled effect, 2) the upper bound of the 95%CI was lower than the lower bound of the pooled effect CI or 3) the lower bound of the 95% CI was higher than the upper bound of the pooled effect CI or 4)A covariance ratio (CovRatio) value <1 of each manuscript [10]. CovRatio was calculated by dividing the variance of the pooled effect without each manuscript by the variance of the initial pooled effect. As a secondary objective, we looked for modifiers of PACS prevalence estimates: patients’ age, gender, comorbidities, and the number of symptoms reported in each article. For this purpose, we performed a meta-regression analysis, and coefficients and 95% confidence intervals were calculated for those symptoms reported at least by three authors [11]. Statistical analyses were performed using the meta package in R software version 4.1.1.

## RESULTS

### Demographics characteristics

Twenty-nine articles were included in the analysis after the systematic review and selection criteria of all screened studies [12–40]. Twenty-two (75.8%), 3 (10.3%), 3 (10.3%) and 1 (3.4%) manuscripts were from Europe, America, Asia, and Africa, respectively. All articles were published during the second semester of 2020. The screening and selection process is shown in Figure 1. The number of patients included in the individual studies ranged from 23 to 4694, with a female proportion between 27 to 75%. The age range of participants was 27 to 70 years, and the period of follow-up since the onset of acute COVID-19 symptoms was 21 to 162 days. We did not identify studies with overlapping samples. The authors reported a wide range of comorbidities. Diabetes, hypertension, and obesity were reported in 17 (59%), 17 (59%), and 10 (37%) articles, respectively. Concerning the number of comorbidities, 6 (21%) manuscripts reported four or fewer, and 14 (48%) reported more than five comorbidities. No information on comorbidities was available for 8 (28%) manuscripts. Demographic and clinical characteristic of included studies are shown in *Table 1*

**Figure 1.**
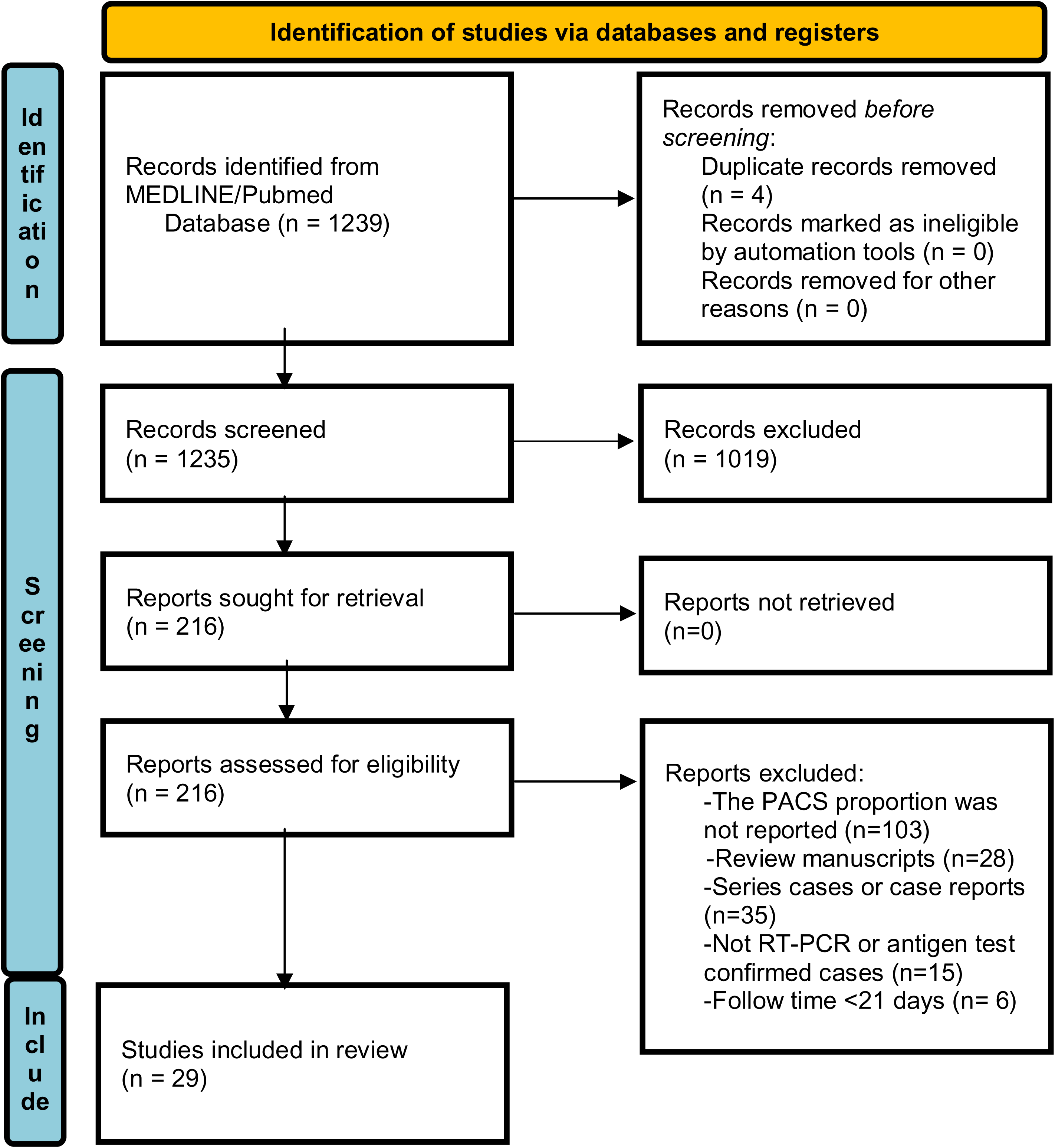
PRISMA flow diagram for search, screening, and manuscript selection.

**Table 1.** General and demographic characteristics of included studies.

| Author | Country | Design | Sample size (N) | Gender (% female) | Mean age years (SD or IQR) | Population | Comorbidities | PACS symptoms | Clinical spectrum (systems involved) | Follow-up (days) |
| --- | --- | --- | --- | --- | --- | --- | --- | --- | --- | --- |
| Konstantinidis, et al. | Greece | Cohort (prospective) | 79 | 51 | 30.7 (5.3) | Ambulatory | Not specified | Nasal obstruction, olfactory and gustatory dysfunctions, rhinitis and, rhinorrhea. | Neurological<br>Respiratory | 30 |
| Carvalho-Schneider, et al. | France | Cohort (prospective) | 150 | 56 | 49 (15) | Ambulatory and hospitalized | 0 comorbidities (46%), 1 comorbidity (35%), 2 comorbidities(19%) | Olfactory and gustatory dysfunctions, chest pain, diarrhea, weight loss, cutaneous signs, dyspnea, arthralgia, palpitations, fever, myalgia, cephalaea, asthenia and vomiting. | Neurological<br>Respiratory<br>Musculoskeletal<br>Gastrointestinal<br>Cardiovascular<br>Constitutional<br>Cutaneous | 30, 60 |
| Kamal, et al. | Egypt | Cross-sectional | 287 | 64 | 32.3(8.5) | Ambulatory and hospitalized | No comorbidities (70.7%), hypertension (7.7%), diabetes (5.2%), obesity (35.5%), dyslipidemia (1.4%), rheumatoid arthritis (1%), hypothyroidism (1%), asthma (1%), peptic ulcer (0.7%), arrhythmia (0.7%), smoking (9.8%) | Fatigue, anxiety, arthralgia, cephalaea, chest pain, depression, dyspnea, new onset diabetes, blurred vision, tinnitus, fever, obsessive-compulsive disorder, migraine, stroke, renal failure, myocarditis, arrhythmia. | Constitutional<br>Psychiatric<br>Musculoskeletal<br>Neurological<br>Respiratory<br>Endocrinological<br>Cardiovascular<br>Renal | 21 |
| Landi, et al. | Italy | Cohort (prospective) | 131 | 39 | 55.8 (14.8) | Hospitalized | Hypertension (29%), COPD (9.1%), heart failure (6.1%), diabetes (5.3%), renal failure (3.1%), smoking (9.8%) | Fatigue, dyspnea, arthralgia, cough, red eye, rhinitis, cephalaea, loss of appetite, sore throat, diarrhoea, and olfactory and gustatory dysfunctions. | Constitutional<br>Respiratory<br>Ocular<br>Musculoskeletal<br>Neurological<br>Gastrointestinal | 55 |
| Garrigues, et al. | France | Cross-sectional | 120 | 38 | 63.2 (15.7) | Hospitalized | Hypertension: (46.7%), overweight or obesity (47.5%), diabetes (21.7%) | Fatigue, dyspnea, memory loss, sleep disorders, attention disorder, hair loss, cough, chest pain, and olfactory and gustatory dysfunctions. | Constitutional<br>Respiratory<br>Neurological<br>Musculoskeletal | 110 |
| Tomasini, et al. | Italy | Cross-sectional | 105 | 27 | 55 (43-65) | Hospitalized | Not specified | Gastrointestinal symptoms, asthenia, anxiety, memory disorders, depression, burning pain, dyspnea, and olfactory and gustatory dysfunctions, chest pain, cephalgia, constipation, tinnitus, insomnia, palpitations, NSTEMI, cough, and sore throat. | Constitutional<br>Psychiatric<br>Neurological<br>Respiratory<br>Gastrointestinal<br>Musculoskeletal<br>Cardiovascular | 30-90 |
| Carfi, et al. | Italy | Cross-sectional | 143 | 37 | 56.5 (16.6) | Hospitalized | Hypertension (35%), thyroid disease (18.2%), immune disorders (11.2%), COPD (9.1%), diabetes (7%), cardiovascular disease (4.9%), cancer (3.5%), atrial fibrillation (2.8%), heart failure (2.8%), renal failure (2.1%), stroke (1.4%), active smoking (10.5%) | Fatigue, dyspnea, arthralgias, chest pain, cough, anosmia, sicca syndrome, rhinitis, red eyes, dysgeusia, headache, sputum, lack of appetite, sore throat, vertigo, myalgia and diarrhea. | Constitutional<br>Respiratory<br>Musculoskeletal<br>Neurological<br>Gastrointestinal | 60 |
| Vaira, et al. | Italy | Cohort (prospective) | 138 | 51 | 51.2 (8.8) | Ambulatory and hospitalized | Cardiovascular disease (26.8%), pulmonary disease (15.2%), diabetes (10.9%), obesity (29%) | Olfactory and gustatory dysfunction | Neurological | 60 |
| Halpin, et al. | UK | Cross-sectional | 100 | 46 | Ward patients: 70.5 (20-93)<br>ICU: 58.5 (34-84) | Hospitalized | Hypertension (41%), obesity (24%), diabetes (29%), gastrointestinal disease (25%), cancer (28%), heart failure (5%), mental health condition (19%), osteoarthritis (13%), rheumatologic disease (14%), CKD (15%), immunosuppressive condition (15%), asthma (13%), neurologic disease (12%), COPD (8%), obstructive sleep apnea (7%), hyperlipidemia (4%), ischemic heart disease (10%), tachyarrhythmias | Fatigue, dyspnea, post-traumatic stress syndrome, concentration and memory alterations, anxiety/depression, pain or discomfort, cough, continence problems, dysphagia, voice changes, appetite problems, and diminished mobility. | Constitutional<br>Respiratory<br>Psychiatric<br>Neurological<br>Urological<br>Gastrointestinal | 48 |
|  |  |  |  |  |  |  | (12%), valvular heart disease (3%), venous thromboembolism (5%), thyroid disease (5%), gynecological disease (3%), hematological disease (10%), infectious disease (6%) |  |  |  |
| Barón-Sánchez, et al. | Spain | Cross-sectional | 31 | 61 | 44.6 (13.08) | Ambulatory | Not specified. | Hyposmia. | Neurological | 38.47 |
| Villarreal, et al. | Spain | Cohort (retrospective) | 230 | 85 | 43 (18-62) | Ambulatory and hospitalized | Not specified. | Olfactory dysfunctions | Neurological | 30 |
| Zhao, et al. | China | Cohort (retrospective) | 55 | 42 | 47.7 (15.49) | Hospitalized | Hypertension (10.91%), diabetes (3.64%), cardiovascular disease (3.64%), smoking (3.64%), others (3.64%). | Gastrointestinal symptoms, cephalaea, fatigue, dyspnea, gustatory dysfunctions, cough, and sputum. | Gastrointestinal<br>Neurological<br>Constitutional<br>Respiratory | 64-93 |
| Pellaud, et al. | Switzerland | Cohort (retrospective) | 196 | N/E | 70 (60-80) | Hospitalized | Hypertension (60%), diabetes (27%), obesity (21%), coronary heart disease (13%), sleep apnea syndrome (13%), cardiac insufficiency (11%), COPD (8%), cancer (8%), immunosuppression (5%), smoking (29%). | Asthenia, respiratory symptoms, olfactory and gustatory dysfunction. | Constitutional<br>Respiratory<br>Neurological | 30 |
| Boscolo-Rizzo, et al. | Italy | Cross-sectional | 187 | 55 | 56 (20-89) | Ambulatory and hospitalized | Not specified. | Gustatory and olfactory dysfunction, cough, dyspnea, cephalaea, congested nose, myalgias/artralgias, loss of appetite, felt tired, sore throat, dizziness, diarrhea, sinonasal pain, abdominal pain, chest pain, fever, nausea. | Neurological<br>Respiratory<br>Musculoskeletal<br>Constitutional<br>Gastrointestinal | 30 |
| Park, et al. | Korea | Cohort (prospective) | 46 | 46 | 26 (18-57) | Hospitalized | Reflux (13%), gastritis (6.5%), irritable bowel disease (6.5%), fatty liver (2.1%), others (6.5%). | Sputum, cough, rhinorrhea, cephalaea, fatigue, chest pain, myalgia, nasal obstruction, anosmia, loss of appetite, fever. | Respiratory<br>Neurological<br>Constitutional<br>Musculoskeletal | 52 |
| Huang, et al. | China | Case-control | 26 | 62 | 38<br>(32-45) | Hospitalized | Hypertension (8%) | Precordial chest pain, palpitations, chest distress. | Cardiovascular | 47 |
| Caronna, et al. | Spain | Cohort<br>(prospective) | 130 | 51 | 53.9<br>(16.4) | Ambulatory<br>and<br>hospitalized | Migraine (70%) | Cephalea. | Neurological | 42 |
| Townsend, et al. | Ireland | Cross-sectional | 128 | 54 | 49.5<br>(15) | Ambulatory/<br>hospitalized | Anxiety/depression (7.8%) | Fatigue, not returning to full health. | Constitutional | 42 |
| Gambini, et al. | Italy | Cohort<br>(prospective) | 114 | 33 | 56.2<br>(14.2) | Hospitalized | Hypertension (43.8%), diabetes (26.6%), autoimmune disease (10.9%). | Reduction of near distance visual acuity, red eye | Ocular | 60.3 |
| Sonnweber, et al. | Austria | Cohort<br>(prospective) | 145 | 45 | 57<br>(14) | Ambulatory<br>and<br>hospitalized | Hypertension (30%), hypercholesterolemia (19%), diabetes (17%), cancer (12%), asthma (7%), CKD (7%), immunodeficiency (6%), COPD (6%), chronic liver disease (6%), interstitial lung disease (1%).. | Dyspnea, sleep disorders, night sweet, pain, olfactory dysfunctions, cough, diarrhea/vomiting. | Respiratory<br>Constitutional<br>Neurological<br>Gastrointestinal | 60-100 |
| Jacobs, et al. | USA | Cohort<br>(prospective) | 183 | 39 | 57<br>(48-68) | Hospitalized | Hypertension (47%), obesity (49.2%), overweight (36.1%), diabetes (28.4%), cardiac disease (27.9%), dyslipidemia (10.9%), asthma (10.4%), cancer (9.8%), immunodeficiency (4.4%), hypothyroidism (4.4%), psychiatric disorder (4.4%), COPD (3.8%), obstructive sleep apnea (3.3%), thromboembolic disease (1.6%). | Fatigue, dyspnea, cough, myalgias, cephalaea, phlegm, ageusia, olfactory dysfunction, confusion, eye irritation, diarrhea, ulcers, fever. | Constitutional<br>Respiratory<br>Musculoskeletal<br>Neurological<br>Ocular<br>Gastrointestinal<br>Cutaneous | 35 |
| Taboada, et al. | Spain | Cohort<br>(prospective) | 91 | 35 | 64.5<br>(10.4) | Hospitalized | Hypertension (55.5%), dislipidemia (44%), obesity (38.5%), diabetes (23.1%), cardiomyopathy (20.9%), COPD (8.8%), asthma (6.6%). | Dyspnea, myalgias, asthenia, insomnia, arthralgias, cough, olfactory dysfunctions, chest pain | Respiratory<br>Musculoskeletal<br>Constitutional<br>Neurological | 180 |
| Gerkin, et al. | USA | Cross- | 4694 | 75* | 40.6 | Not specified | Diabetes (2%), heart disease | Olfactory dysfunctions, | Neurological | 40 |
|  |  | sectional |  |  | (12)* |  | (0.3%), hypertension (8%), lung disease (0.5%), neurological disease (11%), obesity (1%), seasonal allergies (17%), smoking (63%), chronic sinus problems (3%), cancer (0.5%) | runny nose, sore throat, not recovered from respiratory symptom | Respiratory |  |
| Blair, et al. | USA | Cohort (prospective) | 118 | 57 | 56 (50-63) | Ambulatory and hospitalized | Hypertension (39%), smoking (26%), diabetes (16.1%), asthma or COPD (15.3%), solid tumor (15.3%), HIV (6.6%), hematologic malignancy (5.1%), CKD (3.4%), history of myocardial infarction (1.7%). | Fatigue, dry cough | Constitutional Respiratory | 28-60 |
| Niklassen, et al. | Germany, Italy | Cohort (prospective) | 111 | 46 | 44.5 (15) | Not specified | Not specified | Olfactory and gustatory dysfunctions. | Neurological | 28, 45, 77-162 |
| Raman, et al. | UK | Case-control | 88 | 40 | 55.4 (13.2)* | Hospitalized | Hypertension (37.9%), asthma (34.5%), diabetes mellitus (15.5%), depression (5.2%), COPD (5.2%), coronary artery disease (3.4%), cerebrovascular disease (1.7%), previous cancer (3.4%). | Dyspnea, fatigue. | Respiratory Constitutional | 60-90 |
| Townsend, et al. | Ireland | Cross-sectional | 153 | 57 | 48 (35-59) | Ambulatory and Hospitalized | Not specified | Fatigue, not returning to full health. | Constitutional | 75 |
| Myall, et al. | UK | Cohort (prospective) | 35 | 71 | 60.5 (10.7) | Hospitalized | Hypertension (31.4%), Obesity (25.7%), diabetes (22.9%), asthma (22.9%), CKD (5.8%), COPD (5.8%), HIV (2.9%). | Functional deficit. | Respiratory | 42 |
| Weerahandi, et al. | USA | Cohort (prospective) | 161 | 37 | 62 (50-67) | Hospitalized | Smoker (30.4%), CKD (8.1%), cancer (7.5%), arterial coronary disease (9.3%), diabetes (36.7%), cardiac failure (5%), hyperlipidemia (46.6%), hypertension (60.3%), asthma or COPD (24.2%), overweight (30.4%), obesity (41%). | Dyspnea, diminished general health status, diminished physical health, diminished mental health. | Respiratory Psychiatric | 37 |

\*From COVID-19 group.
Red eyes included: burning eye, foreign body sensation, watering eye, dry eye disease and eye irritation.
Olfactory dysfunctions include anosmia, hyposmia, dysosmia, phantosmia.
Gustatory dysfunctions include: ageusia, hypogeusia, dysgeusia
Olfactory and gustatory dysfunction include both olfactory and gustatory manifestations if they were reported together in the same article.
Chronic obstructive pulmonary disease (COPD), chronic kidney disease (CKD), Human Immunodeficiency Virus (HIV).
(N/E) Not specified

### Pooled prevalence of long-term PACS manifestations

For the pooled prevalence of persistent symptoms, 27 meta-analyses were performed, and 61 long-term manifestations as part of the PACS spectrum were described (Table 2). The general PACS prevalence was 56% (95%CI 45-66%), which included any new or persistent symptom after three weeks from COVID-19 onset. Fatigue (49%) and asthenia (32%) were the most prevalent constitutional symptoms after acute COVID-19. For cognitive manifestations, attention disorders and memory loss were reported in 27% and 23% of patients, respectively, while anxiety and post-traumatic stress disorders were reported in around 30%. The long-term prevalence and system involvement of PACS manifestations are shown in Figure 2.

**Figure 2.**
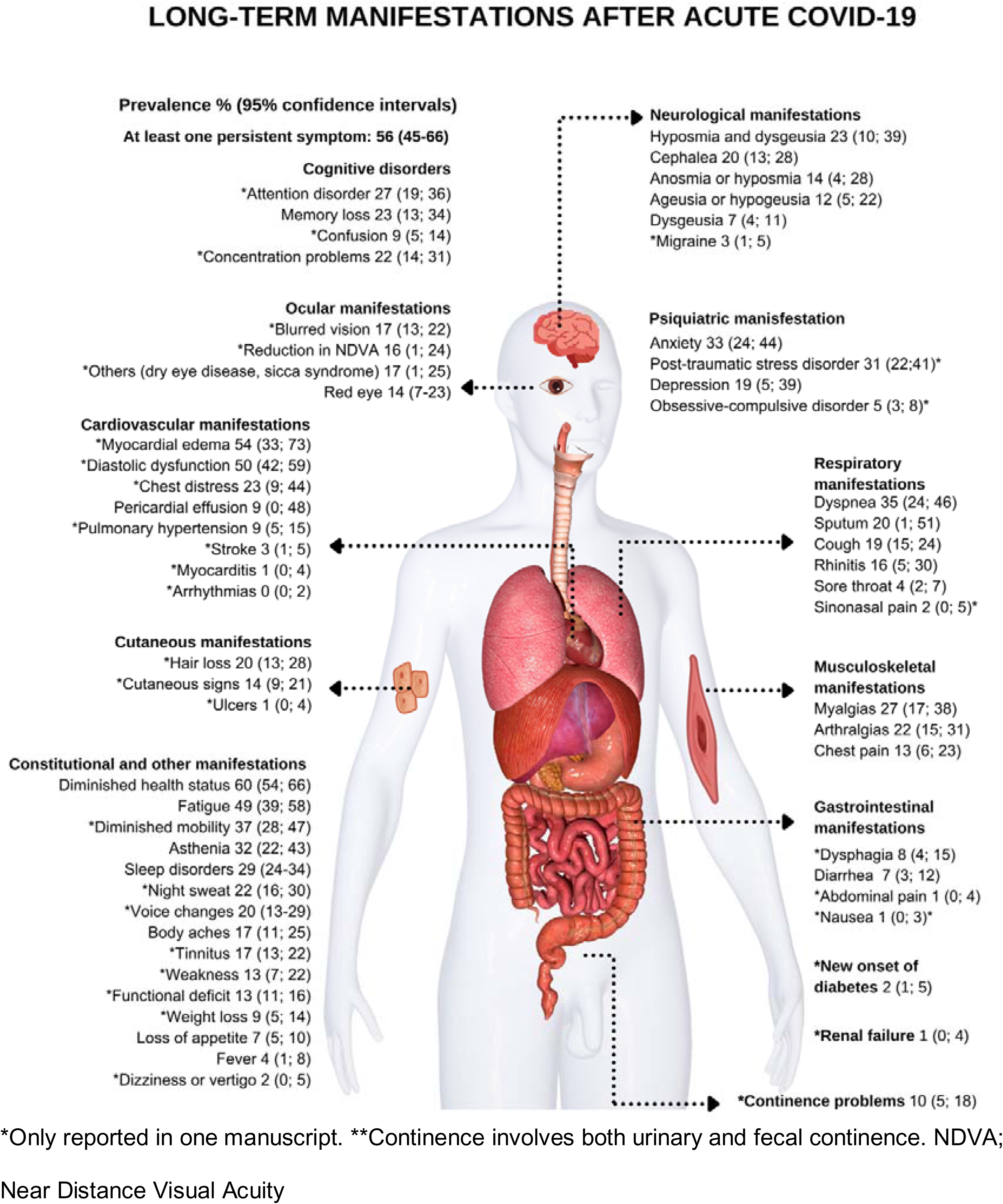
Pooled prevalence estimates of long-term manifestations after acute COVID-19

**Table 2.** Pooled prevalence estimates of new or persistent manifestation after acute COVID-19

| Symptom | Studies | Cases | Sample Size | Prevalence % (95 CI%) | I2 (%) | Symptom | Studies | Cases | Sample Size | Prevalence % (95 CI%) | I2 (%) |
| --- | --- | --- | --- | --- | --- | --- | --- | --- | --- | --- | --- |
| **PACS (at least one persistent symptom)** | 15 | 2923 | 5586 | 56 (45-66) | 98 | **Gastrointestinal manifestations** |  |  |  |  |  |
| **Constitutional manifestations** |  |  |  |  |  | Dysphagia | 1 | 8 | 100 | 8 (4; 15) | - |
| Fatigue | 12 | 828 | 1534 | 49 (39; 58) | 93 | Diarrhea | 5 | 59 | 774 | 7 (3; 12) | 82 |
| Asthenia | 5 | 199 | 649 | 32 (22; 43) | 89 | Abdominal pain | 1 | 2 | 187 | 1 (0; 4) | - |
| Night sweat | 1 | 32 | 145 | 22 (16; 30) | - | Nausea | 1 | 1 | 187 | 1 (0; 3) | - |
| Body aches | 3 | 62 | 350 | 17 (11; 25) | 68 | **Musculoskeletal manifestations** |  |  |  |  |  |
| Tinnitus | 1 | 48 | 287 | 17 (13; 22) | - | Myalgias | 4 | 133 | 470 | 27 (17; 38) | 84 |
| Weakness | 1 | 11 | 84 | 13 (7; 22) | - | Arthralgias | 6 | 225 | 963 | 22 (15; 31) | 88 |
| Weight loss | 1 | 13 | 150 | 9 (5; 14) | - | Chest pain | 8 | 174 | 1050 | 13 (6; 23) | 93 |
| Loss of appetite | 4 | 34 | 442 | 7 (5; 10) | 2 | **Endocrinological manifestations** |  |  |  |  |  |
| Fever | 5 | 45 | 853 | 4 (1; 8) | 86 | New onset diabetes | 1 | 7 | 287 | 2 (1; 5) | - |
| Dizziness or vertigo | 1 | 3 | 187 | 2 (0; 5) | - | **Renal manifestations** |  |  |  |  |  |
| **Neurological manifestations** |  |  |  |  |  | Renal failure | 1 | 4 | 287 | 1 (0; 4) | - |
| Hyposmia and dysgeusia | 4 | 99 | 426 | 23 (10; 39) | 92 | **Others** |  |  |  |  |  |
| Cephalaea | 8 | 240 | 1117 | 20 (13; 28) | 89 | Continence problems | 1 | 10 | 100 | 10 (5; 18) | - |
| Anosmia or hyposmia | 12 | 1747 | 4358 | 14 (4; 28) | 99 | Sleep disorders | 3 | 103 | 356 | 29 (24; 34) | 0 |
| Ageusia or hypogeusia | 4 | 77 | 591 | 12 (5; 22) | 91 | Voice changes | 1 | 20 | 100 | 20 (13; 29) | - |
| Dysgeusia | 3 | 19 | 269 | 7 (4; 11) | 18 | **General health perception** |  |  |  |  |  |
| Migraine | 1 | 8 | 287 | 3 (1; 5) | - | Diminished general health status. | 2 | 169 | 281 | 60 (54; 66) | 0 |
| **Cognitive disorders** |  |  |  |  |  | Diminished mobility | 1 | 37 | 100 | 37 (28;47) | - |
| Attention disorder | 1 | 32 | 120 | 27 (19; 36) | - | Functional deficit | 1 | 110 | 837 | 13 (11; 16) | - |
| Memory loss | 3 | 77 | 325 | 23 (13; 34) | 82 | **Respiratory manifestations** |  |  |  |  |  |
| Confusion | 1 | 16 | 183 | 9 (5; 14) | - | Dyspnea | 13 | 573 | 1733 | 35 (24; 46) | 96 |
| Concentration problems | 1 | 22 | 100 | 22 (14; 31) | - | Sputum | 3 | 54 | 284 | 20 (1; 51) | 96 |
| **Psychiatric manifestations** |  |  |  |  |  | Cough | 8 | 192 | 965 | 19 (15; 24) | 66 |
| Anxiety | 2 | 138 | 392 | 33 (24; 44) | 73 | Rhinitis** | 4 | 51 | 372 | 16 (5; 30) | 89 |
| Post-traumatic | 1 | 31 | 100 | 31 (22;41) | - | Sore throat | 2 | 13 | 296 | 4 (2; 7) | 0 |
| Depression | 2 | 93 | 392 | 19 (5; 39) | 94 | Sinonasal pain | 1 | 3 | 187 | 2 (0; 5) | - |
| Obsessive-compulsive disorder | 1 | 14 | 287 | 5 (3; 8) | - | **Cardiovascular manifestations** |  |  |  |  |  |
| **Ocular manifestations** |  |  |  |  |  | Myocardial edema | 1 | 14 | 26 | 54 (33; 73) | - |
| Blurred vision | 1 | 49 | 287 | 17 (13; 22) | - | Diastolic dysfunction | 1 | 73 | 145 | 50 (42; 59) | - |
| Reduction in NDVA | 1 | 18 | 114 | 16 (1; 24) | - | Chest distress | 1 | 6 | 26 | 23 (9; 44) | - |
| Others (dry eye disease, sicca syndrome) | 1 | 19 | 114 | 17 (1; 25) | - | Pericardial effusion | 2 | 8 | 171 | 9 (0; 48) | 95 |
| Red eye* | 3 | 55 | 406 | 14 (7-23) | 80 | Pulmonary hypertension | 1 | 13 | 145 | 9 (5; 15) | - |
| **Cutaneous manifestations** |  |  |  |  |  | Stroke | 1 | 8 | 287 | 3 (1; 5) | - |
| Hair loss | 1 | 24 | 120 | 20 (13; 28) | - | Arrhythmias | 1 | 1 | 287 | 0 (0; 2) | - |
| Cutaneous signs | 1 | 21 | 150 | 14 (9; 21) | - | Myocarditis | 1 | 4 | 287 | 1 (0; 4) | - |
| Ulcers | 1 | 2 | 183 | 1 (0; 4) | - |  |  |  |  |  |  |

\*Red eye includes eye irritation, foreign body sensation, burning eye, watering eye
\*\*Rhinitis includes congested nose and runny nose.
Near Distance Visual Acuity (NDVA); Non-ST segment elevation myocardial infarction (NSTEMI)

The most frequent long-term respiratory symptoms were dyspnea (35%), and sputum (20%), followed by cough (19%). Myocardial edema and diastolic dysfunction were present in 1 of 2 patients; nevertheless, this prevalence was reported only by one article. Gastrointestinal symptoms and rectal or urinary incontinence were reported in no more than 10% of patients. Ophthalmological sequelae occurred in <20% of cases. Around 1 in 4 patients had myalgias and arthralgias as part of musculoskeletal manifestations. For cutaneous signs, loss of hair was the most frequent finding. Sixty percent of the included participants reported a diminished overall health perception, and sleep disorders were observed in one-third. In addition, the report by Kamal et al. included new-onset diabetes mellitus and renal failure as entities related to COVID-19 long-term spectrum [24]. Forest plots for specific symptoms are found in *Supplementary material (figures).* Five (19%), 3 (11%), and 19 (70%) of meta-analyses reported low, substantial, and high heterogeneity, respectively. After outlier analysis, 11 (40%) pooled analysis shown low, 1 (4%) moderate, 6 substantial (22%), and 9 (33%) high heterogeneity (Table 3).

**Table 3.** Outlier removing analysis of pooled prevalence estimates for PACS and individual long-term manifestations.

| Symptom | Prevalence %<br>(95 CI%) | I<sup>2</sup> (%) | Outlier studies<br>excluded |
| --- | --- | --- | --- |
| PACS | 53 (47-69) | 82 | 7 |
| Fatigue | 48 (44-53) | 41 | 4 |
| Asthenia | 36 (32-41) | 0 | 1 |
| Body aches | 17 (11; 25) | 68 | 0 |
| Loss of appetite | 7 (5-10) | 2 | 0 |
| Fever | 2 (1;4) | 0 | 1 |
| Hyposmia and dysgeusia | 29 (24-35) | 0 | 1 |
| Cephalea | 20 (13; 28) | 89 | 0 |
| Anosmia or hyposmia | 13 (9-17) | 78 | 2 |
| Ageusia or hypogeusia | 8 (4-13) | 59 | 1 |
| Dysgeusia | 7 (4-11) | 18 | 0 |
| Memory loss | 18 (13-23) | 0 | 1 |
| Red eye* | 14 (7-23) | 80 | 0 |
| Diarrhea | 5 (3-7) | 12 | 1 |
| Myalgias | 31 (21-42) | 83 | 1 |
| Arthralgias | 26 (20-32) | 75 | 1 |
| Chest pain | 15 (10-19) | 51 | 2 |
| Sleep disorders | 29(24; 34) | 0 | 0 |
| Dyspnea | 36 (29-43) | 83 | 5 |
| Sputum | 7 (0-24) | 89 | 1 |
| Cough | 18 (14-22) | 55 | 1 |
| Rhinitis** | 9 (6-13) | 0 | 1 |

\*Red eye includes eye irritation, foreign body sensation, burning eye, watering eye
\*\*Rhinitis includes congested nose and runny nose.

### Meta-regression for modifiers of PACS prevalence

When evaluating modifying factors of PACS prevalence estimation, we found that age significantly raised prevalence estimation for fever, cough, and diarrhea within the evaluated manuscripts. Similarly, the proportion of female reported in the studies increased prevalence estimation of body aches, fever, and anosmia or hyposmia. Also, the prevalence estimation of arthralgias and red eye decreased with an increasing number of comorbidities. We also found that the general prevalence of PACS and cough increased according to the number of symptoms investigated by authors in each study, while the prevalence of asthenia and red-eye decreased. Coefficients with 95% confidence intervals of the metaregression are shown in *Supplementary Material (Table 1)*.

## DISCUSSION

Prevalence and variety of long-term clinical manifestations of COVID-19 are not fully characterized. This analysis described the prevalence of 61 new or persistent manifestations after 21 days from the onset of COVID-19 symptoms and reported modifiers of prevalence estimation, which may influence PACS spectrum. To date, there is not an accepted period for establishing long-term complications. Some institutions such as the Centers for Disease Control and Prevention (CDC) in the USA propose four weeks to define the long-term effects of COVID-19, while the guidelines of the National Institute for Health and Care Excellence (NICE) in England considers a more extended period of >12 weeks to classify the long-term manifestations. Notably, sequelae of COVID-19 persist for longer than six months and are higher than other respiratory infections, including influenza [41]. Definition of clinical stages during COVID-19 has proven to be challenging. However, Van Kampen, JJA et al, observed that shedding of SARS-CoV-2 virus has an average duration of 15 days, with a probability of <5% isolation of viable virus after three weeks from symptom onset. Based on this observation, a period of 21 days may reflect the conclusion of acute COVID-19 [42].

We found that 1 in 2 patients with COVID-19 will develop new or persistent clinical manifestations compatible with PACS. In a previous meta-analysis, Lopez-Leon S. et al. reported PACS prevalence as high as 80% [43]. In contrast, Taquet et al. found in a cohort of 273,618 COVID-19 survivors that 57% of patients had one or more long-COVID clinical manifestations during a 6-month period, although this number decreased to 36% after three months, this prevalence continues to be a large burden of long-term sequels [41]. Similar to our findings, Groff et al, reported a median PASC frequency at 1 month of 54.0% (45.0%-69.0%;); at 2-5 months, 55.0% (34.8%-65.5%); and at 6 or more months, 54.0% (31.0%-67.0%) [44]

Even though more reports are published on PACS, the heterogeneity of studies continues to limit the quality of evidence. In 19 (70%) of the pooled prevalence meta-analysis performed, heterogeneity was high; this large heterogeneity was expected first due to subjectivity involving specific clinical manifestations and second to the inability of standardized tools to evaluate some long-term manifestations. To deal with this high heterogeneity, an analysis to determine outlier manuscripts was performed, showing a 50% decrease of all high-level pooled prevalence analyses. This high heterogeneity could be explained to prevalence modifiers identified in the meta-regression analysis, including age, gender, comorbidities, and the number of evaluated symptoms [45]. By controlling sources of heterogeneity that influence prevalence estimates for PACS and its manifestations, future studies are more likely to provide unbiased prevalence estimates of PACS, which may better inform the follow-up of patients after acute COVID-19.

Little is known about the clinical characteristics of patients who develop PACS. A report by the CDC indicates that nearly one of five out-setting patients with no comorbidities and age from 18-34 years did not return to usual health within three weeks of testing due to the persistence of symptoms [46]. Interestingly, Dennis et al. observed that 70% of the individuals at low risk of COVID-19 related mortality developed impairment in one or more organs in the following four months after the onset of symptoms; most of those patients were young women with a low prevalence of comorbidities and ambulatory treatment for COVID-19 [47]. These characteristics are opposite of what is described in acute COVID-19, in which comorbid men have a higher susceptibility to develop severe COVID-19 [48]. Notably, we found that the percentage of women reported in the manuscripts analyzed increased the prevalence of body aches, fever, anosmia, or hyposmia, while reported age, modified prevalence in an inverse relation for fever, cough, and diarrhea. Thus, intrinsic patient characteristics and the design of previous studies may contribute to gender and age differences.

Immunity-modulated gender disparities may contribute to the differences found in the prognosis of COVID-19 and PACS onset. The infection response process follows a series of steps that initiate with the entrance of the SARS-CoV-2 virus to cells by attaching to ACE-2 receptor and its posterior recognition by Toll-like receptors (TLR) 3, 7, 8, and 9, RIG-I and MDA5, with a subsequent triggering of type I interferon (I-IFN) response and NLRP3 inflammasome activation, these signals culminate in pyroptosis and inflammation response [49]. Some reports establish an internal capacity of some coronaviruses to diminish and delay I-IFN response, leading to increased viral replication and severe illness [50]. Likewise, it has been proposed that the imbalance of this normal response may culminate in other disease manifestations such as multisystem inflammatory syndrome and post-acute COVID syndrome. In addition, higher titles of anti-IFN antibodies are found in men compared to women [51]. Thus, a more robust type I-IFN response in women and young individuals may partially explain the better immunological response during acute COVID-19 and posterior more adverse long-term effects in this population [52].

Precise estimates of PACS prevalence and its manifestations will be pivotal to planning public health policies aimed at mitigating the long-term socio-economic impact of Long COVID on public and private healthcare systems around the world [53]. Given the wide spectrum of PACS on different body systems, a multidisciplinary approach will be required to facilitate the recovery and rehabilitation of patients after COVID-19 [54]. Until now, there is no specific treatment or management to treat PACS manifestations; however, recent data on the impact of COVID-19 vaccinations on symptom recovery in PACS is promising and may provide a viable pathway to reduce its burden despite the emergence of new SARS-CoV-2 variants or breakthrough infections [55–57]

Our study has several limitations. Available data were primarily extracted from observational studies with no standardized symptoms in MEDLINE. Also, post-acute COVID-19 syndrome definitions were not available, contributing to increased risks of bias. Results should be interpreted with caution as high expected heterogeneity was found in most meta-analytic results. Despite that, a strength of our study was the meta-regression analysis conducted to explore for modifiers of prevalence estimates for post-acute COVID-19 syndrome. In addition, our findings are similar to those previously reported. Comorbidities were not available for eight manuscripts for meta-regression analysis, and one article did not specify the proportion of women in the population.

Our findings may help standardize populations to study PACS after acute COVID-19 in future studies and to provide better estimates of long-term manifestations, influencing PACS-related public policy. Longitudinal studies on incident COVID-19 cases require standardized approaches regarding the definition of PACS and long-term manifestations, consistent follow-up times across studies and populations with diverse clinical spectrums of COVID-19 to better understand the incidence of PACS, its burden and the impact on patients and healthcare systems. Data available on request from the authors.

## Supporting information

Prisma Checklist

Data quality

## Data Availability

Data is fully available within the manuscript

## Funding support

No specific founding was disclosed

## Conflict of interest’s disclosers

No conflict of interests was declared by the authors

---

**Supplementary table 1.** Meta Regression for modifiers of prevalence estimates for PACS and individual long-term manifestations.

|  | % of Female gender |  | Number of comorbidities |  | Age |  | Number of symptoms |  |
| --- | --- | --- | --- | --- | --- | --- | --- | --- |
| Symptom | Coefficient (CI 95%) | P value | Coefficient (CI 95%) | P value | Coefficient (CI 95%) | P value | Coefficient (CI 95%) | P value |
| **PACS** | -0.0035 (-0.0122-0.0052) | 0.425 | 0.0295 (-0.0111-0.0702 ) | 0.154 | 0.0016 (-0.0102- 0.0133) | 0.793 | 0.0207 ( 0.0021-0.0394 ) | 0.030 |
| **Fatigue** | 0.0031 (-0.0079; 0.0141) | 0.581 | 0.0016 (-0.0135; 0.0168) | 0.8318 | 0.0020 (-0.0061; 0.0102) | 0.625 | -0.000 (-0.0177; 0.0177) | 1.000 |
| **Asthenia** | -0.0034 (-0.0145; 0.0077) | 0.546 | - | - | 0.0059 (-0.0102; 0.0221) | 0.472 | -0.0195 (-0.0365; -0.0025) | 0.025 |
| **Body aches** | 0.0075 (0.0014; 0.0137) | 0.017 | - | - | 0.0038 (-0.0138; 0.0214) | 0.673 | 0.0966 (-0.0991; 0.2923) | 0.333 |
| **Loss of appetite** | -0.0022 (-0.0114; 0.0070) | 0.635 | 0.0024 (-0.0075; 0.0122) | 0.633 | 0.0026 (-0.0013; 0.0065) | 0.194 | 0.0007 (-0.0218; 0.0232) | 0.952 |
| **Fever** | 0.0081 (0.0026; 0.0136) | 0.004 | -0.0029 (-0.0390; 0.0332) | 0.874 | -0.0050 (-0.0099; -0.0000) | 0.049 | 0.0240 (-0.0057; 0.0538) | 0.113 |
| **Hyposmia and dysgeusia** | 0.0080 (-0.0058; 0.0217) | 0.254 | - | - | -0.0091 (-0.0188; 0.0006) | 0.067 | 0.0074 (-0.0265; 0.0412) | 0.670 |
| **Anosmia or hyposmia** | 0.0101 (0.0045; 0.0158) | <0.001 | 0.0099 (-0.0450; 0.0648) | 0.723 | -0.0027 (-0.0159; 0.0106) | 0.695 | -0.0044 (-0.0355; 0.0266) | 0.778 |
| **Ageusia or hypogeusia** | 0.0066 (-0.0125; 0.0256) | 0.499 | 0.0026 (-0.0186; 0.0239) | 0.809 | -0.0066 (-0.0342; 0.0209) | 0.636 | 0.0172 (-0.0093; 0.0437) | 0.203 |
| **Dysgeusia** | 0.0008 (-0.0137; 0.0154) | 0.912 | - | - | 0.0114 (-0.0082; 0.0309) | 0.254 | 0.0205 (-0.0058; 0.0468) | 0.127 |
| **Cephalaea** | 0.0078 (-0.0017; 0.0173) | 0.108 | -0.011 (-0.0315; 0.0095) | 0.292 | -0.0061 (-0.0136; 0.0013) | 0.107 | -0.0043 (-0.0237; 0.0151) | 0.664 |
| **Dyspnea** | -0.0064 (-0.0188; 0.0060) | 0.312 | 0.0022 (-0.0157; 0.0201) | 0.808 | 0.0128 (-0.0001; 0.0258) | 0.052 | -0.0210 (0.0467; 0.0047) | 0.109 |
| **Cough** | 0.0015 (-0.0063; 0.0094) | 0.698 | -0.0042 (-0.0208; 0.0124) | 0.620 | -0.0064 (-0.0113; -0.0015) | 0.011 | 0.012 (0.0005; 0.0236) | 0.041 |
| **Sputum** | 0.0697 (-0.0677; 0.2072) | 0.320 | -0.020 (-0.112; 0.072) | 0.671 | -0.0175 (-0.0431; 0.0082) | 0.183 | 0.0399 (-0.1094; 0.1893) | 0.600 |
| **Rhinitis\*** | -0.0093 (-0.0459; 0.0273) | 0.619 | - | - | -0.0079 (-0.0176; 0.0018) | 0.110 | 0.0038 (-0.0360; 0.0435) | 0.853 |
| **Diarrhea** | 0.0079 (-0.0020; 0.0178) | 0.117 | 0.0006 (-0.0299; 0.0310) | 0.971 | -0.025 (-0.0396; -0.0104) | <0.001 | -0.0091 (-0.0557; 0.0375) | 0.702 |
| **Myalgias** | 0.0010 (-0.0168; 0.0189) | 0.909 | 0.0062 (-0.0342; 0.0465) | 0.764 | 0.0058 (-0.0027; 0.0142) | 0.184 | -0.0158 (-0.0723; 0.0407) | 0.584 |
| **Arthralgias** | -0.0013 (-0.0112; 0.0086) | 0.794 | -0.0199 (-0.0388; -0.0010) | 0.039 | -0.0009 (-0.0110; 0.0091) | 0.854 | -0.0038 (-0.0279; 0.0203) | 0.759 |
| **Chest pain** | 0.0018 (-0.0102; 0.0137) | 0.772 | 0.0148 (-0.0070-0.0367) | 0.184 | -0.0047 (-0.0125; 0.0031) | 0.241 | -0.0018 (-0.0309; 0.0273) | 0.903 |
| **Red eye\*\*** | -0.0230 (-0.0528; 0.0068) | 0.130 | -0.0165 (-0.0267; -0.0062) | 0.002 | -0.1093 (-0.3159; 0.0973) | 0.300 | -0.0151 (-0.0278; -0.0025) | 0.019 |
| **Sleep disorders** | -0.0057 (-0.0179; 0.0065) | 0.361 | -0.0065 (-0.0216; 0.0085) | 0.394 | 0.0067 (-0.0076; 0.0209) | 0.357 | -0.0139 (-0.0733; 0.0455) | 0.646 |
| **Memory loss** | 0.0017 (-0.0207; 0.0240) | 0.8846 | - | - | 0.0012 (-0.0264; 0.0289) | 0.931 | 0.1041 (-0.2078; 0.4160) | 0.513 |

\*Red eye included: eye irritation, foreign body sensation, burning eye, watering eye
\*\*Rhinitis included: congested nose and runny nose.

## Supplementary figures

**Forest plots of long-term manifestation pooled prevalence estimates.**

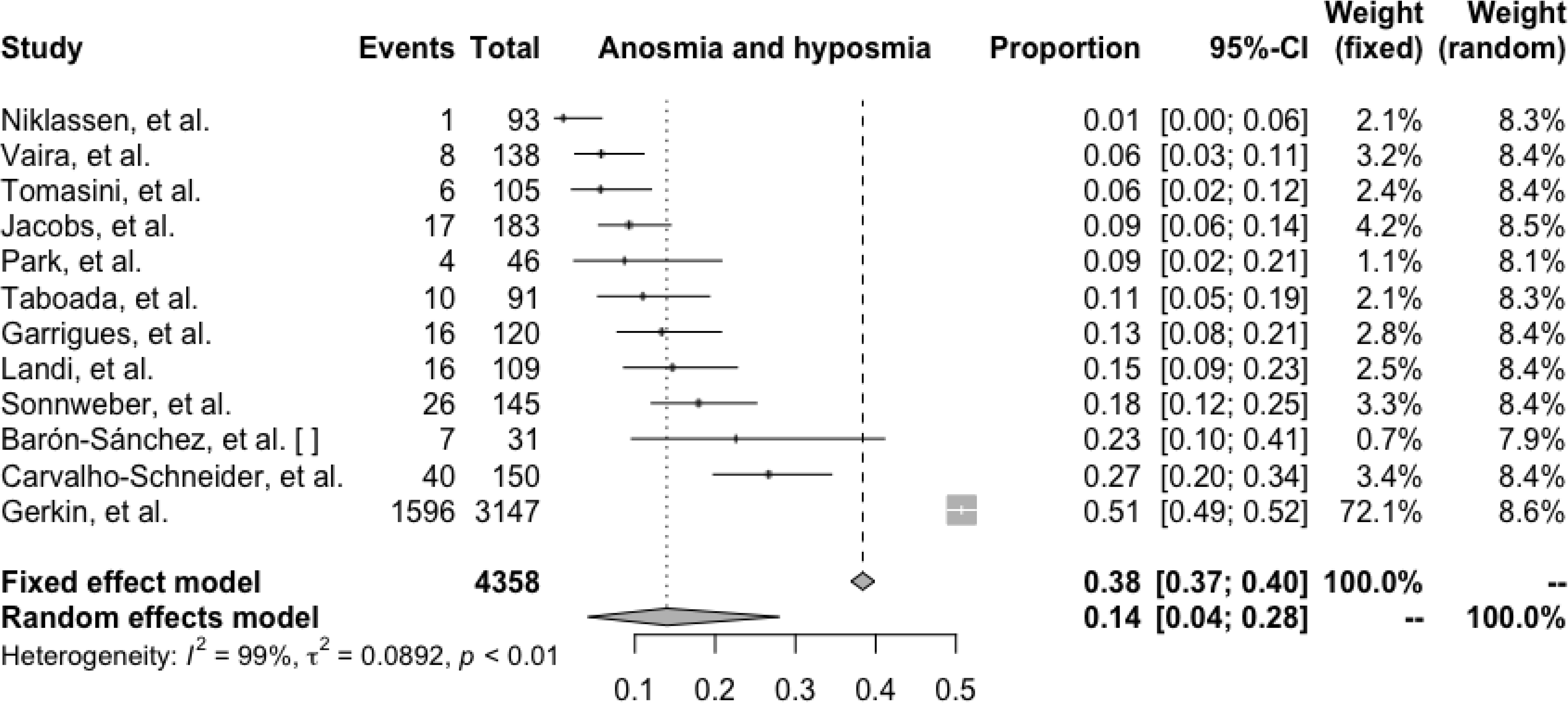
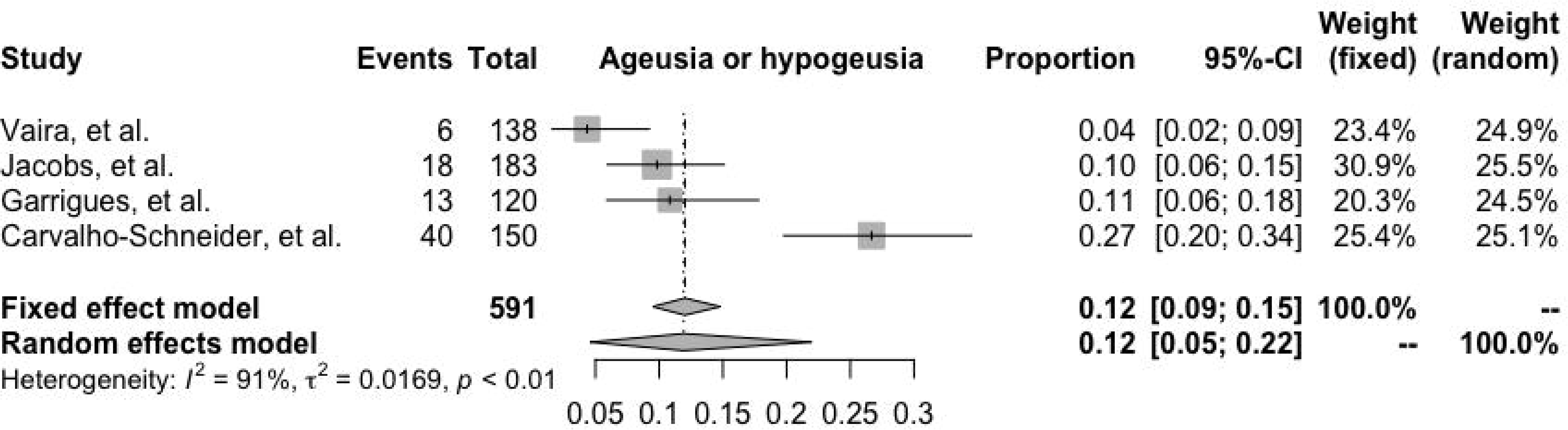
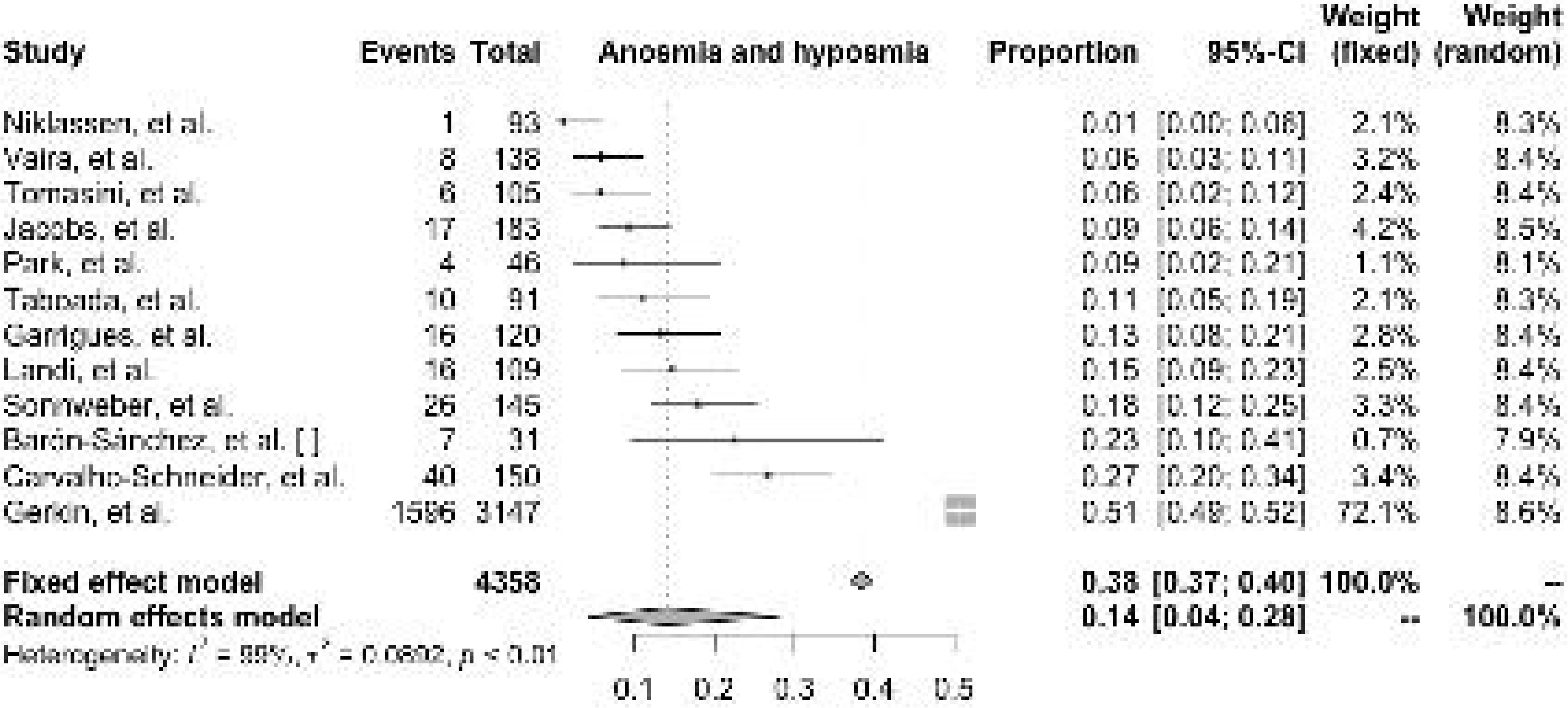
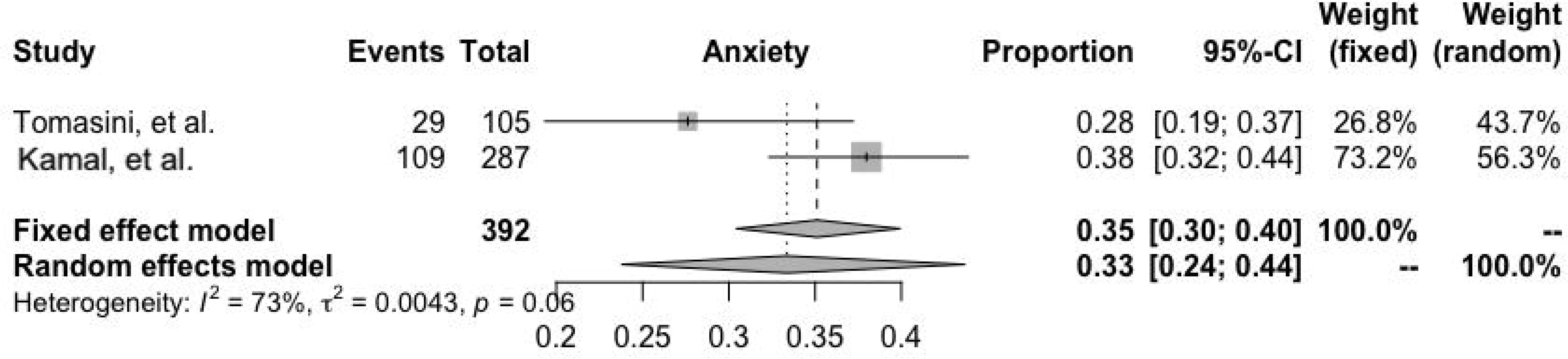
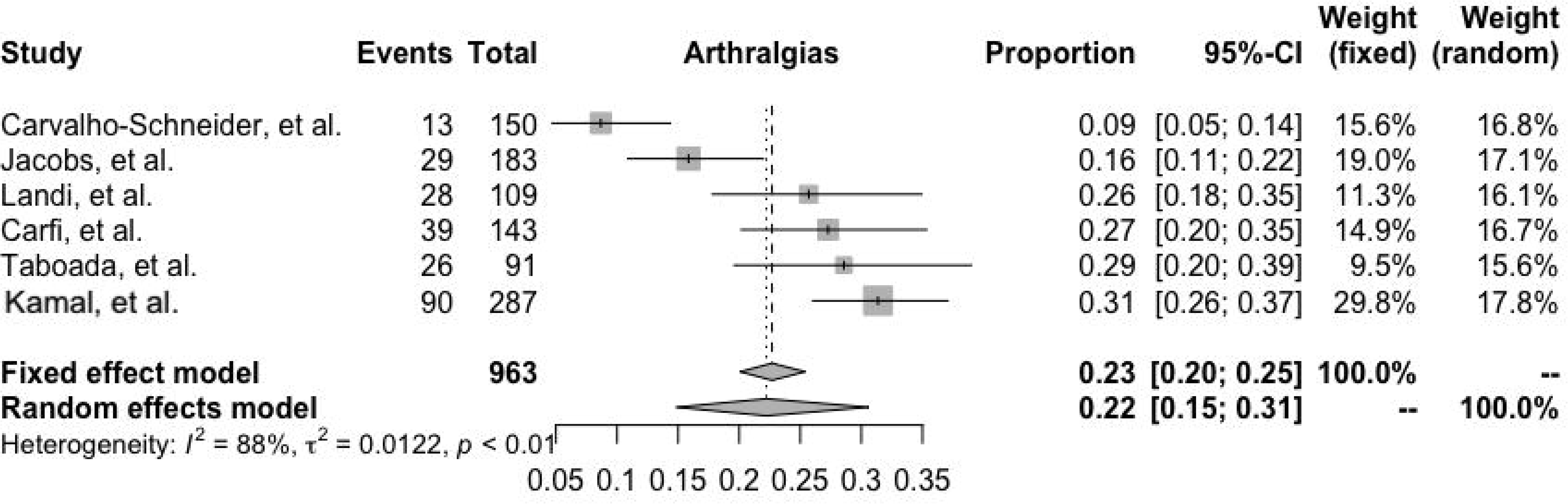
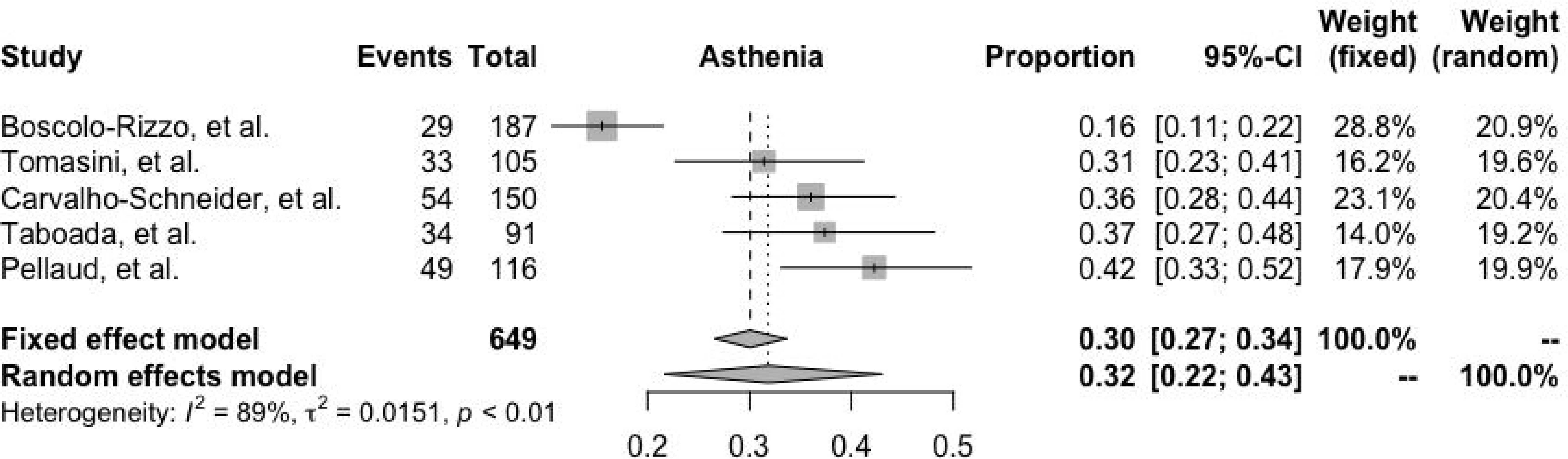
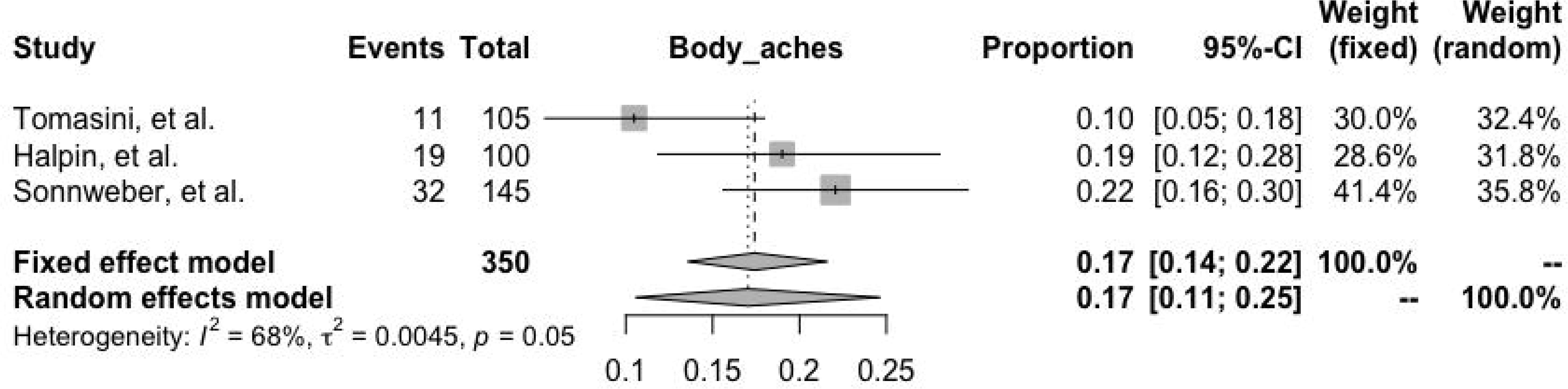
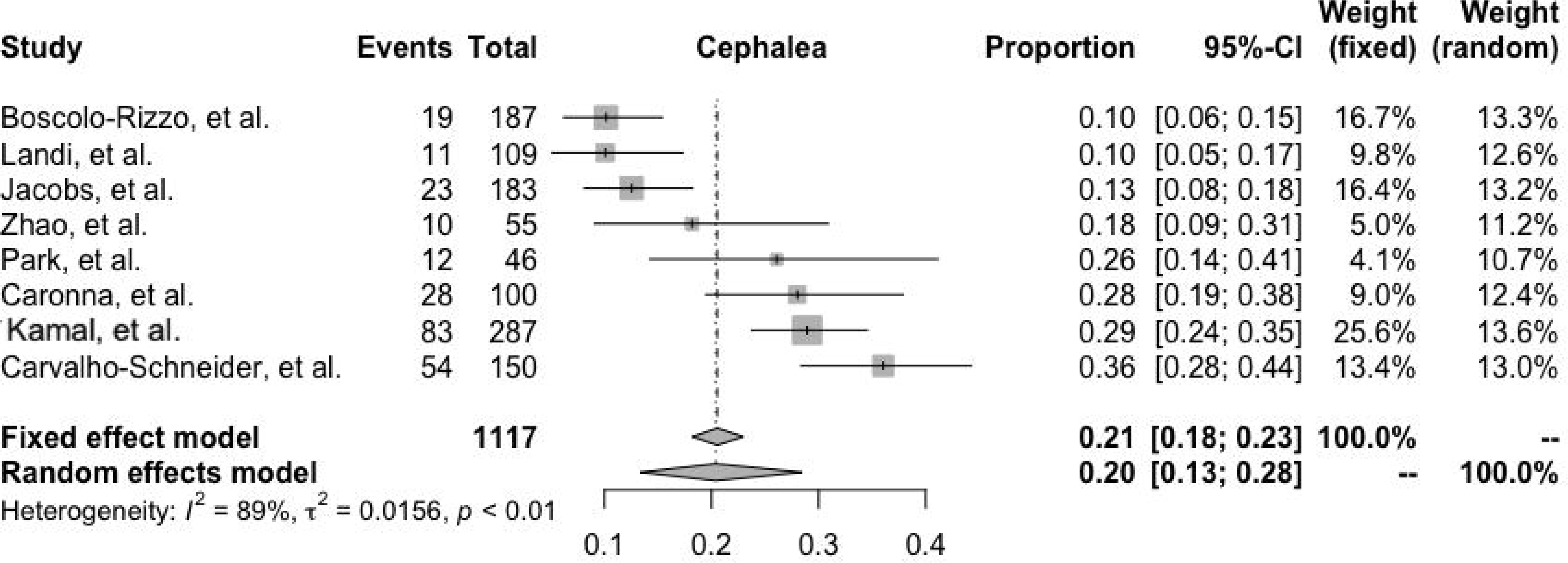
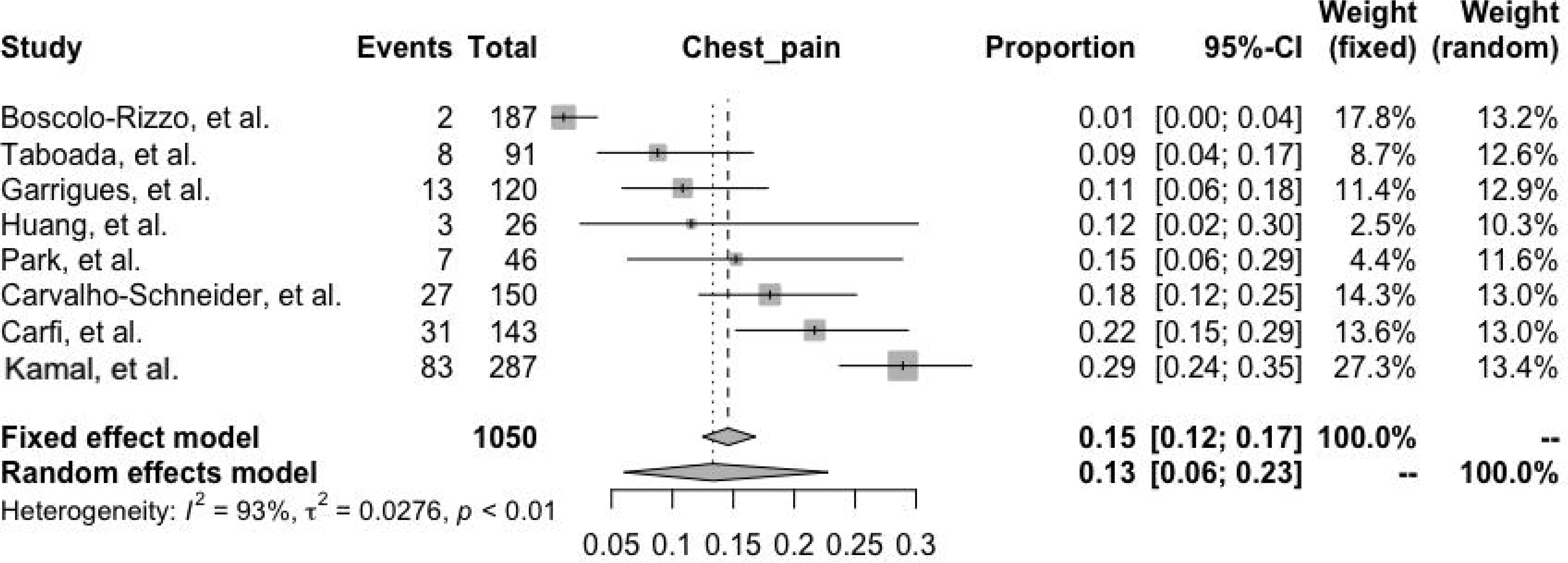
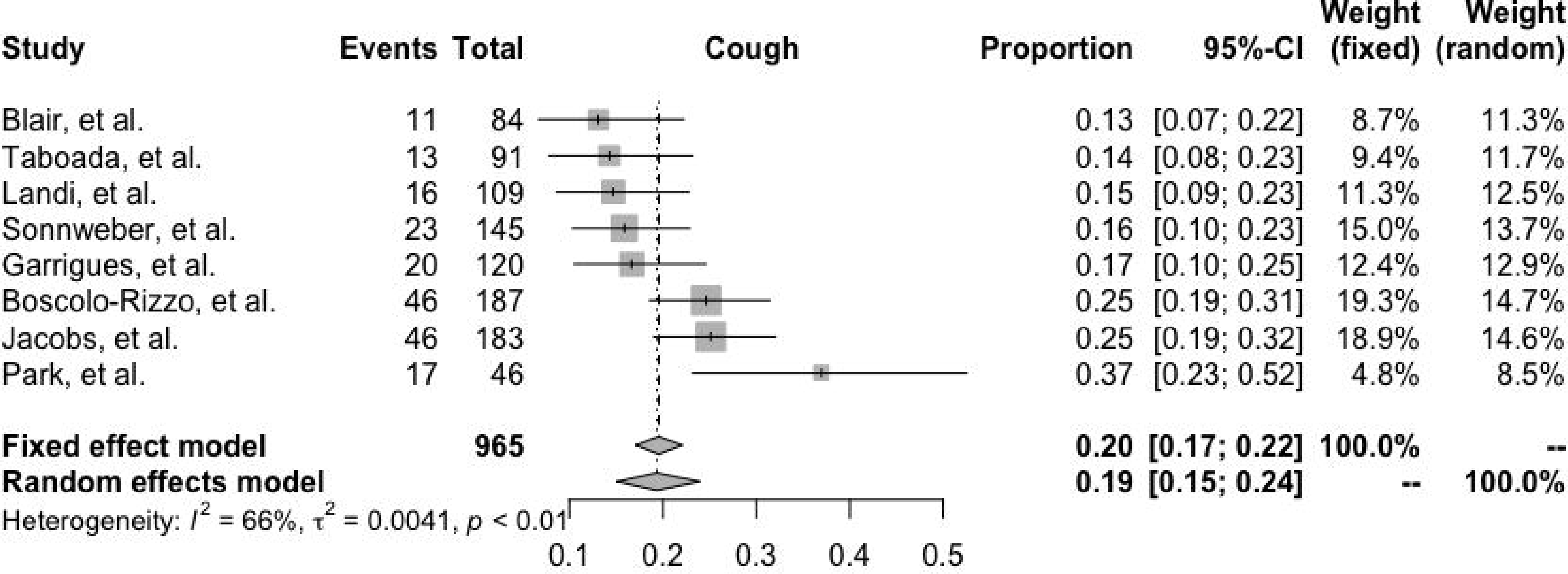
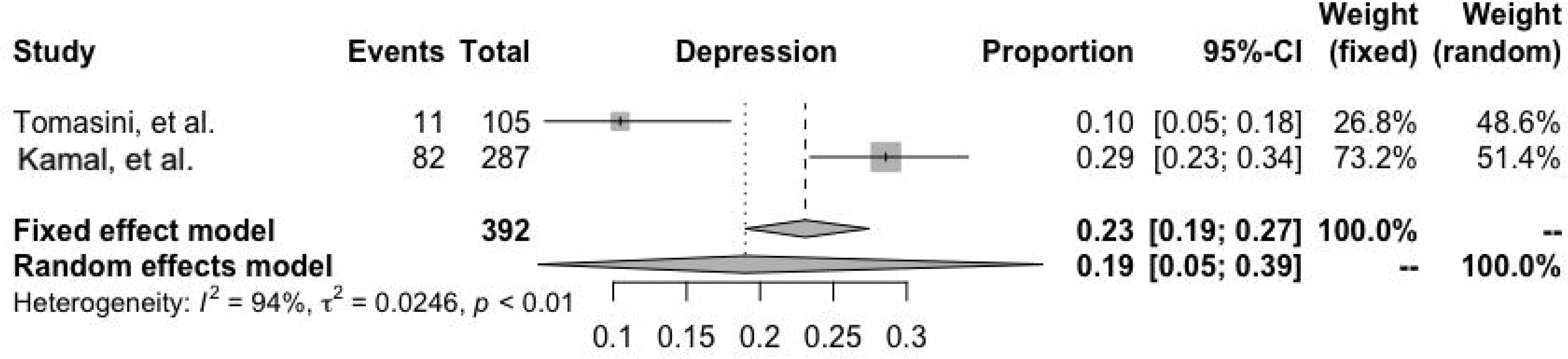
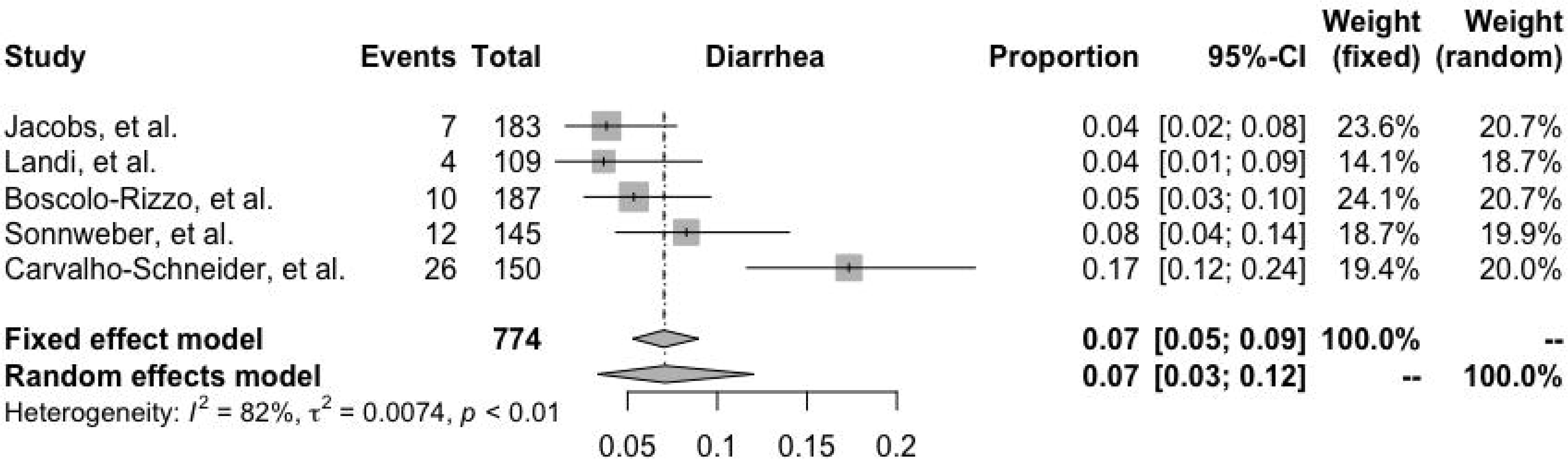
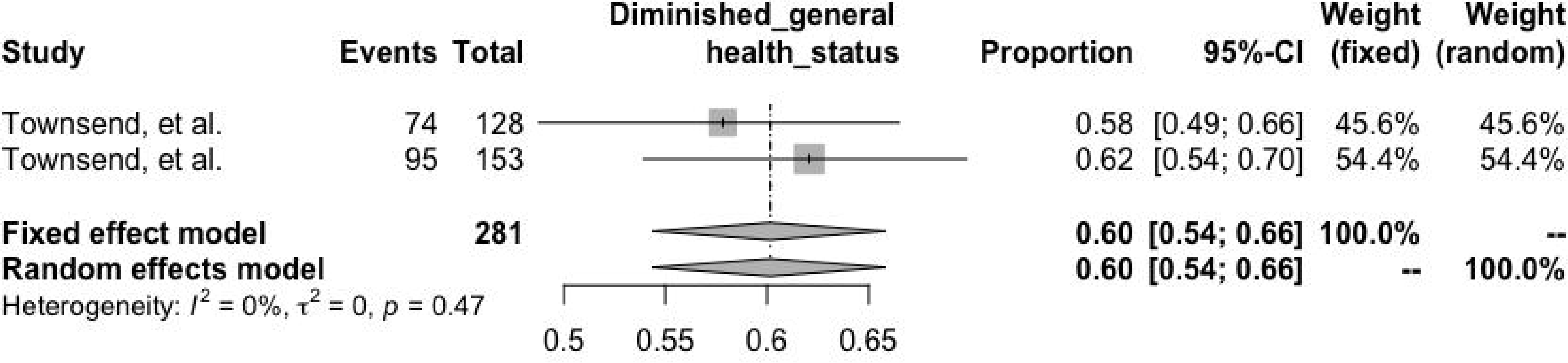
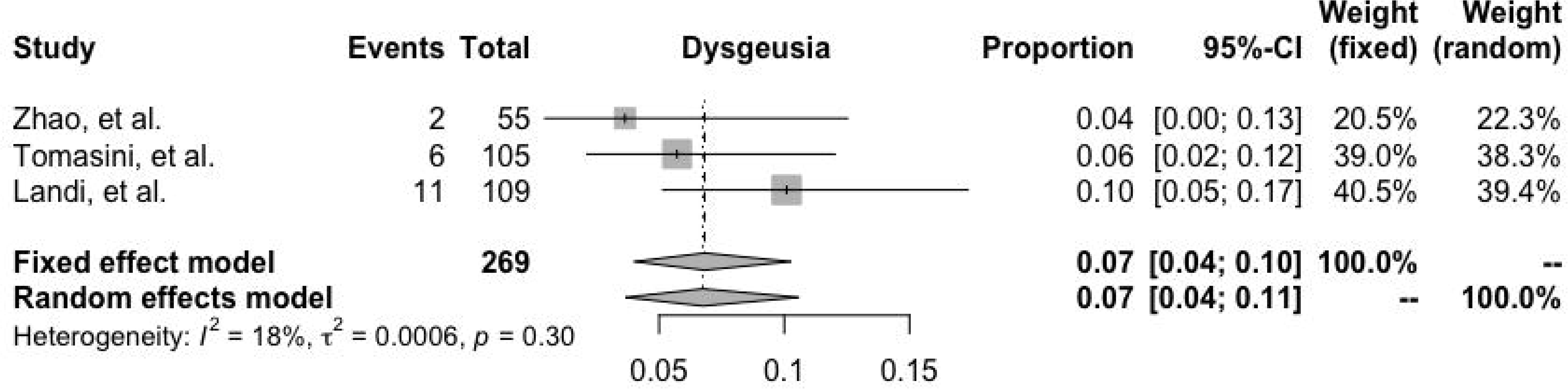
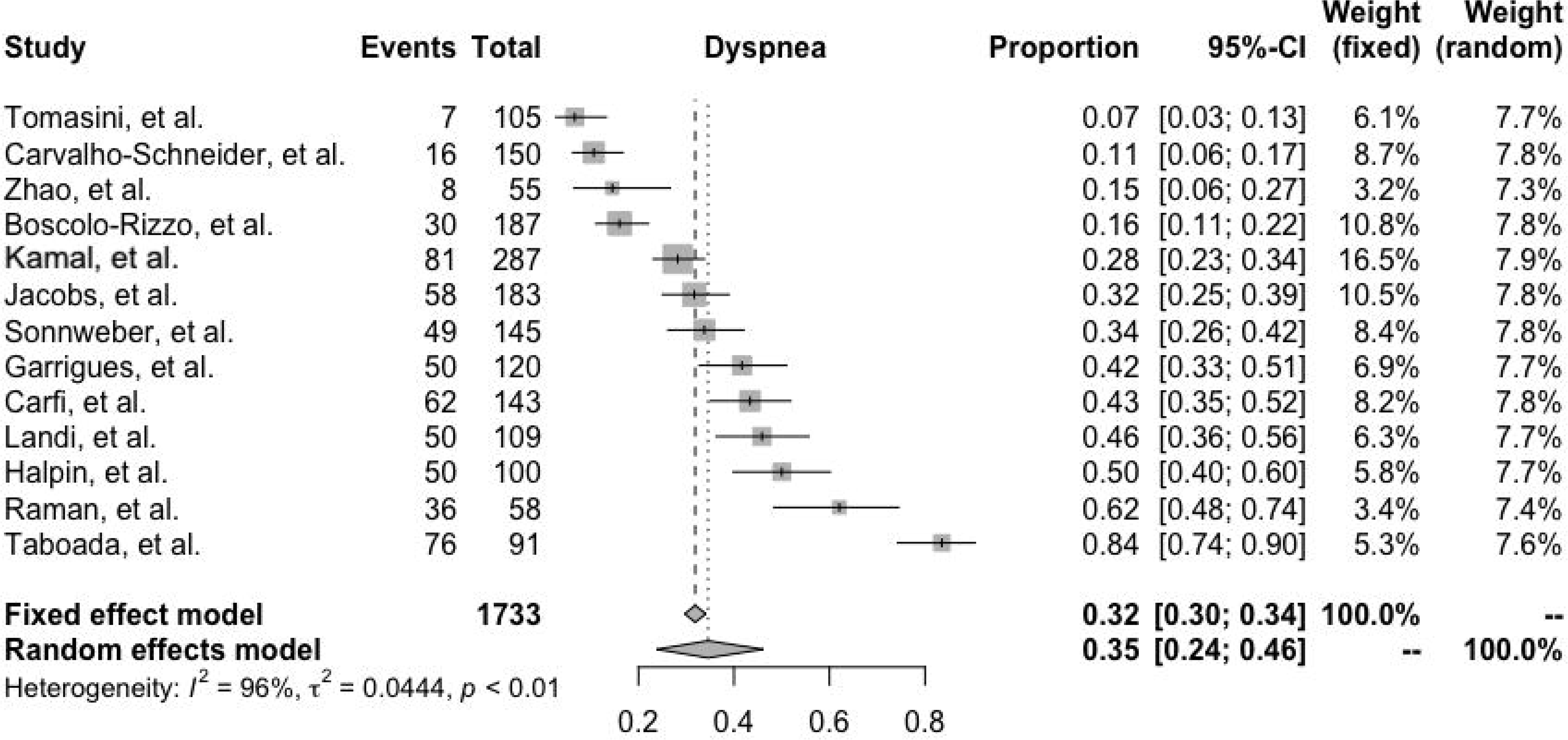
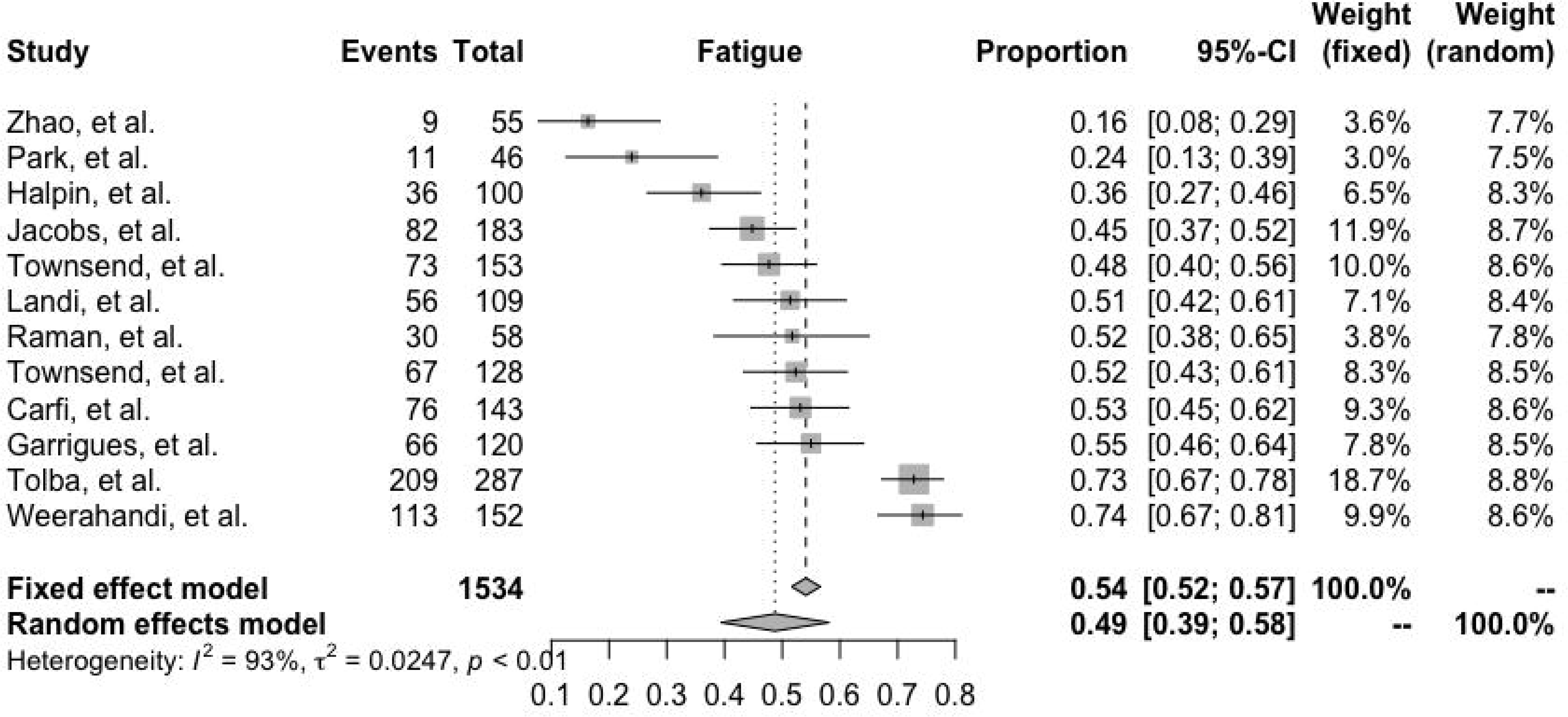
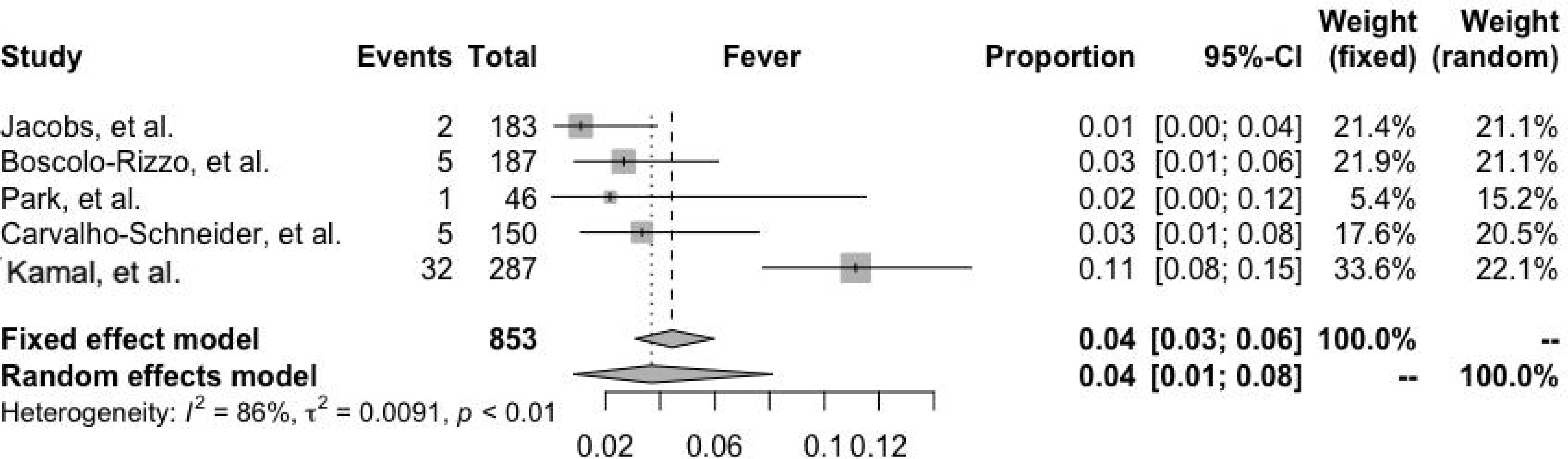
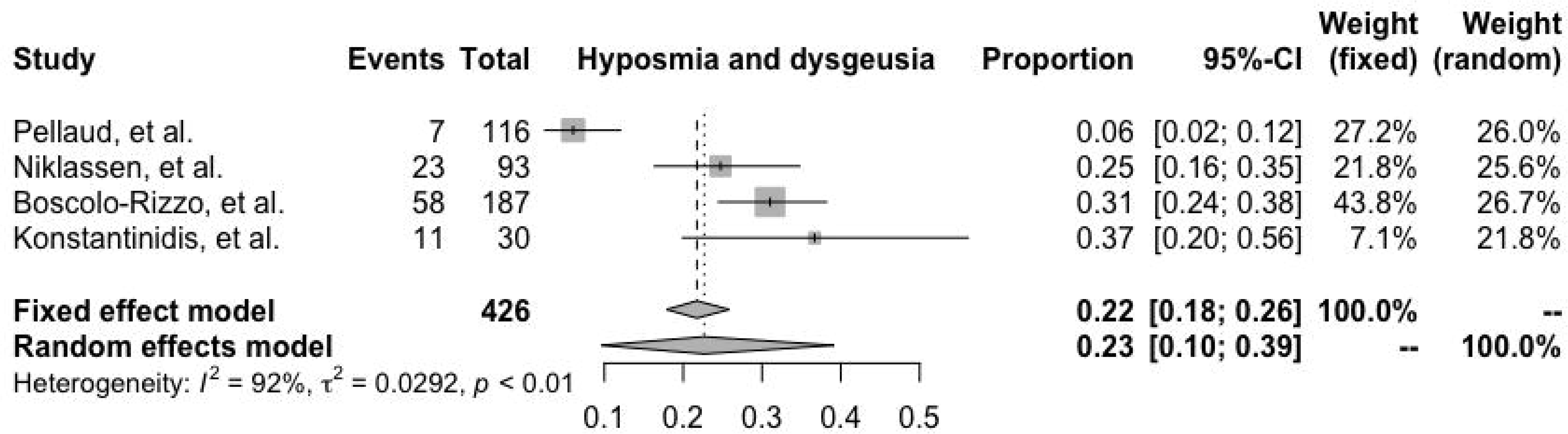
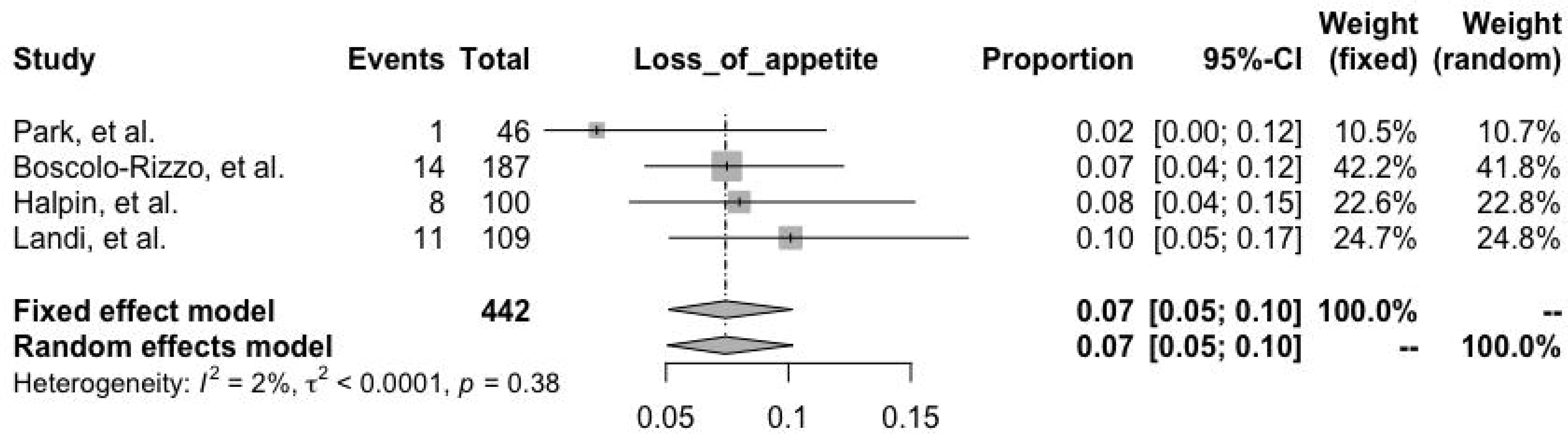
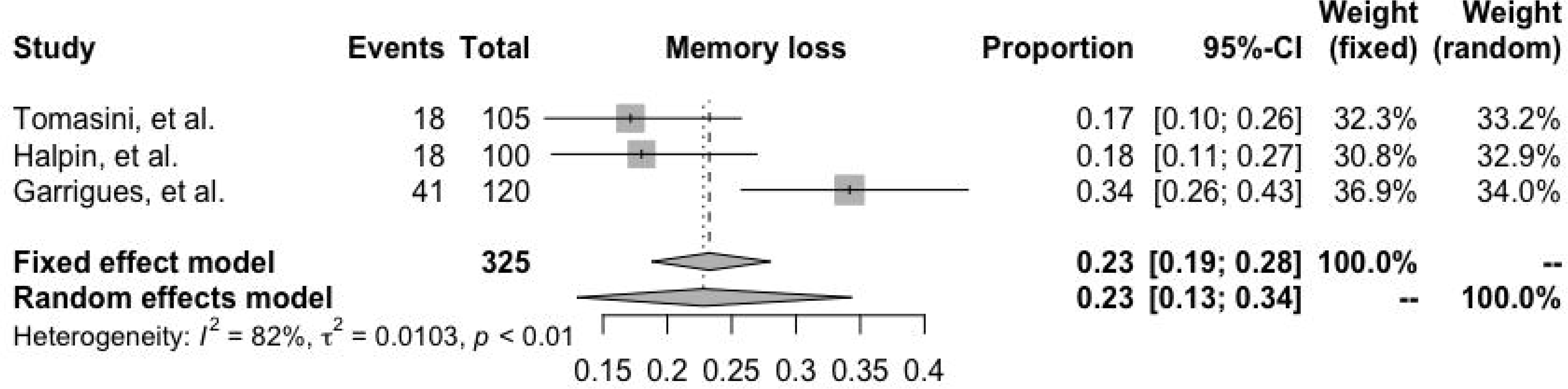
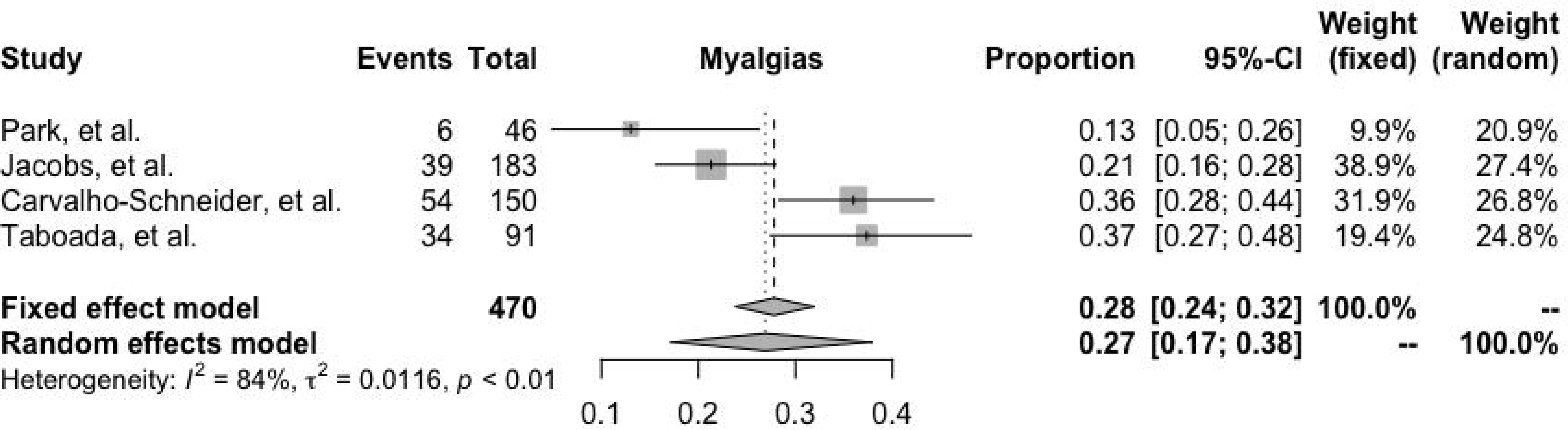
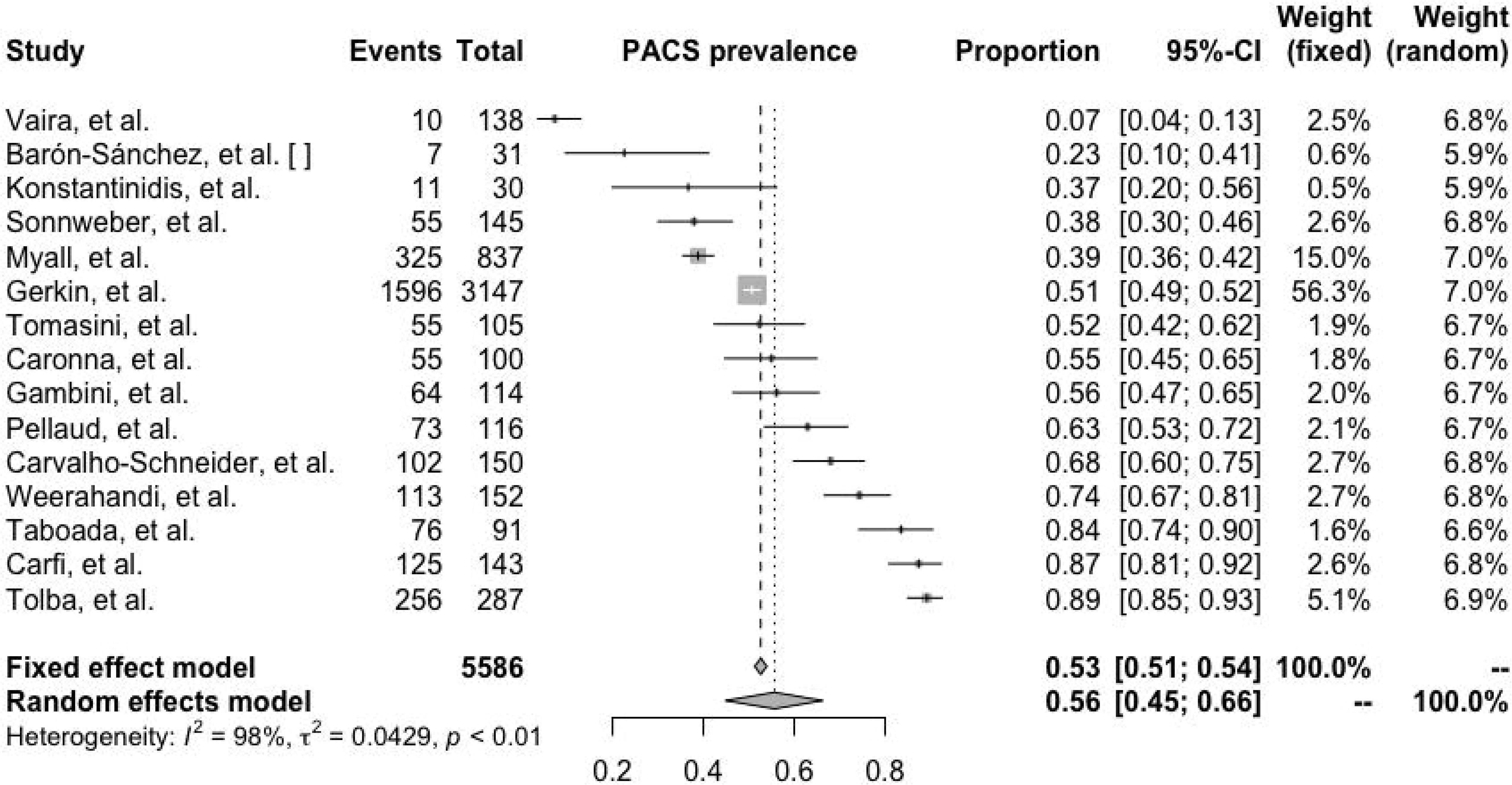
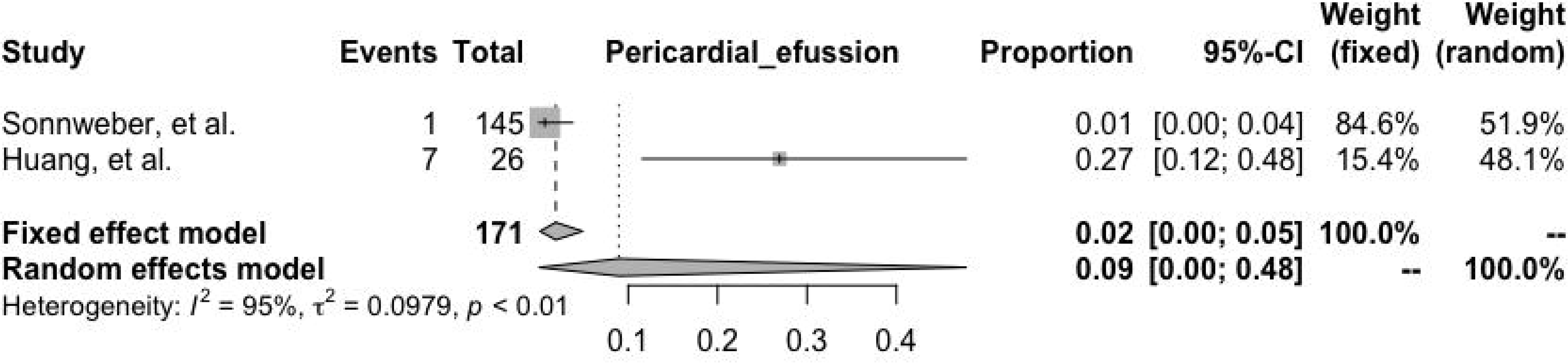
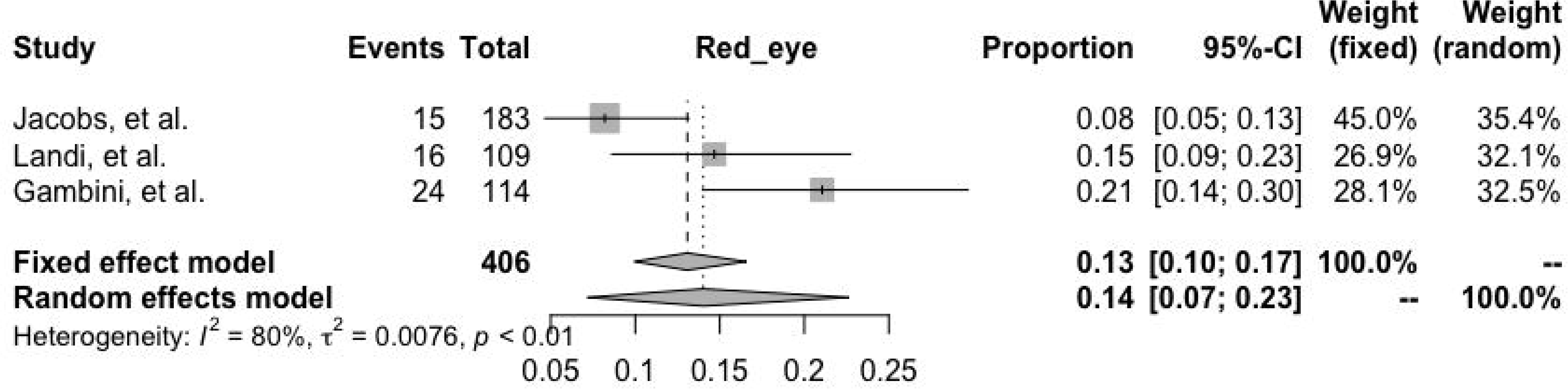
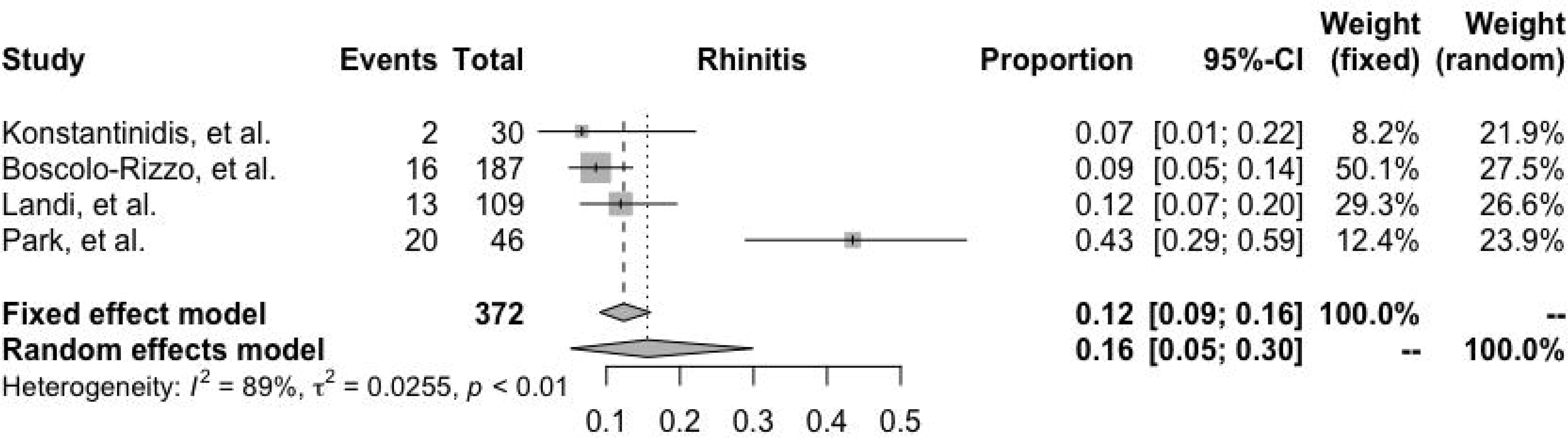
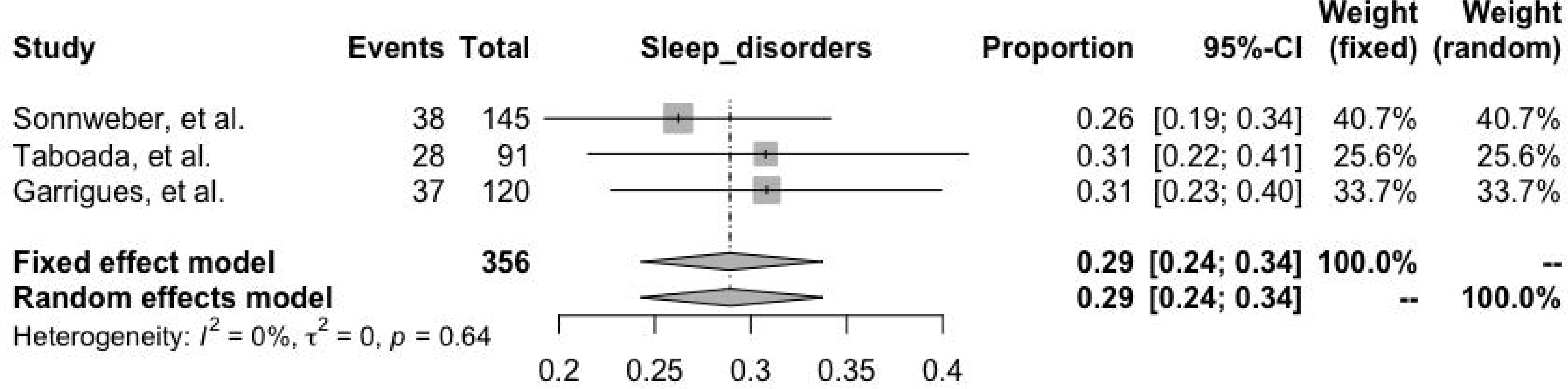
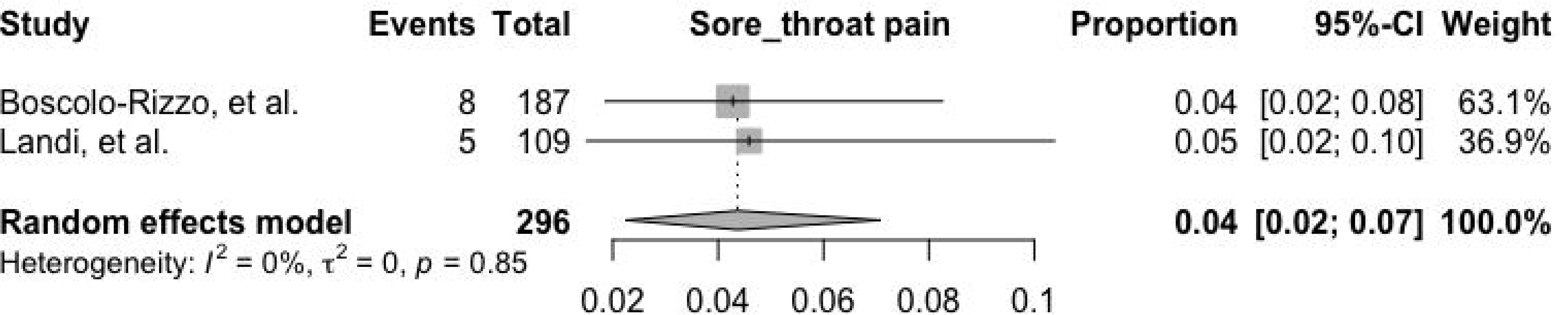
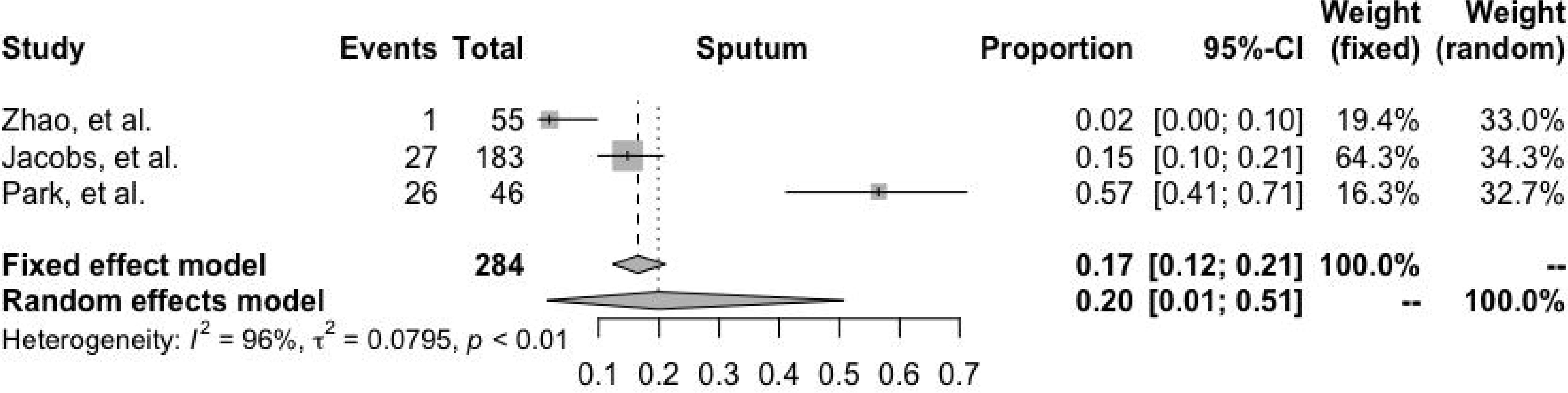

## Notes

### Competing Interest Statement

The authors have declared no competing interest.

### Funding Statement

No specific funding was received for this study

## REFERENCES

1. Cohen PA, Hall LE, John JN, Rapoport AB. The Early Natural History of SARS-CoV-2 Infection. Mayo Clinic Proceedings 2020; 95: 1124–6.

2. Wu Z, McGoogan JM. Characteristics of and Important Lessons From the Coronavirus Disease 2019 (COVID-19) Outbreak in China: Summary of a Report of 72 314 Cases From the Chinese Center for Disease Control and Prevention. JAMA 2020; 323: 1239.

3. . WHO. A clinical case definition of post COVID-19 condition by a Delphi consensus, 6 October 2021.

4. Moldofsky H, Patcai J. Chronic widespread musculoskeletal pain, fatigue, depression and disordered sleep in chronic post-SARS syndrome; a case-controlled study. BMC Neurol 2011; 11: 37.

5. Lam MH-B. Mental Morbidities and Chronic Fatigue in Severe Acute Respiratory Syndrome Survivors: Long-term Follow-up. Arch Intern Med 2009; 169: 2142.

6. Guillot X, Ribera A, Gasque P. Chikungunya-Induced Arthritis in Reunion Island: A Long-Term Observational Follow-Up Study Showing Frequently Persistent Joint Symptoms, Some Cases of Persistent Chikungunya Immunoglobulin M Positivity, and No Anticyclic Citrullinated Peptide Seroconversion After 13 Years. J Infect Dis 2020; 222: 1740–4.

7. Clark DV, Kibuuka H, Millard M, et al. Long-term sequelae after Ebola virus disease in Bundibugyo, Uganda: a retrospective cohort study. Lancet Infect Dis 2015; 15: 905–12.

8. Nalbandian A, Sehgal K, Gupta A, et al. Post-acute COVID-19 syndrome. Nat Med 2021; 27: 601– 15.

9. Langan D, Higgins JPT, Jackson D, et al. A comparison of heterogeneity variance estimators in simulated random-effects meta-analyses. Res Syn Meth 2019; 10: 83–98.

10. Viechtbauer W, Cheung MW-L. Outlier and influence diagnostics for meta-analysis. Res Synth Method 2010; 1: 112–25.

11. Viechtbauer W. Conducting Meta-Analyses in R with the metafor Package. J Stat Soft 2010; 36: 1–48.

12. Barón-Sánchez J, Santiago C, Goizueta-San Martín G, Arca R, Fernández R. Afectación del sentido del olfato y el gusto en la enfermedad leve por coronavirus (COVID-19) en pacientes españoles. Neurología 2020; 35: 633–8.

13. Blair PW, Brown DM, Jang M, et al. The Clinical Course of COVID-19 in the Outpatient Setting: A Prospective Cohort Study. Open Forum Infectious Diseases 2021; 8: ofab007.

14. Boscolo-Rizzo P, Borsetto D, Fabbris C, et al. Evolution of Altered Sense of Smell or Taste in Patients With Mildly Symptomatic COVID-19. JAMA Otolaryngol Head Neck Surg 2020; 146: 729.

15. Carfì A, Bernabei R, Landi F, for the Gemelli Against COVID-19 Post-Acute Care Study Group. Persistent Symptoms in Patients After Acute COVID-19. JAMA 2020; 324: 603.

16. Caronna E, Ballvé A, Llauradó A, et al. Headache: A striking prodromal and persistent symptom, predictive of COVID-19 clinical evolution. Cephalalgia 2020; 40: 1410–21.

17. Carvalho-Schneider C, Laurent E, Lemaignen A, et al. Follow-up of adults with noncritical COVID-19 two months after symptom onset. Clinical Microbiology and Infection 2021; 27: 258–63.

18. Gambini G, Savastano MC, Savastano A, et al. Ocular Surface Impairment After Coronavirus Disease 2019: A Cohort Study. Cornea 2021; 40: 477–83.

19. Garrigues E, Janvier P, Kherabi Y, et al. Post-discharge persistent symptoms and health-related quality of life after hospitalization for COVID-19. Journal of Infection 2020; 81: e4–6.

20. Gerkin RC, Ohla K, Veldhuizen MG, et al. Recent Smell Loss Is the Best Predictor of COVID-19 Among Individuals With Recent Respiratory Symptoms. Chemical Senses 2021; 46: bjaa081.

21. Halpin SJ, McIvor C, Whyatt G, et al. Postdischarge symptoms and rehabilitation needs in survivors of COVID-19 infection: A cross-sectional evaluation. J Med Virol 2021; 93: 1013–22.

22. Huang L, Zhao P, Tang D, et al. Cardiac Involvement in Patients Recovered From COVID-2019 Identified Using Magnetic Resonance Imaging. JACC: Cardiovascular Imaging 2020; 13: 2330–9.

23. Jacobs LG, Gourna Paleoudis E, Lesky-Di Bari D, et al. Persistence of symptoms and quality of life at 35 days after hospitalization for COVID-19 infection. Madeddu G, ed. PLoS ONE 2020; 15: e0243882.

24. Kamal M, Abo Omirah M, Hussein A, Saeed H. Assessment and characterisation of post-COVID-19 manifestations. Int J Clin Pract 2021; 75: e13746.

25. Konstantinidis I, Delides A, Tsakiropoulou E, Maragoudakis P, Sapounas S, Tsiodras S. Short-Term Follow-Up of Self-Isolated COVID-19 Patients with Smell and Taste Dysfunction in Greece: Two Phenotypes of Recovery. ORL J Otorhinolaryngol Relat Spec 2020; 82: 295–303.

26. Landi F, Carfì A, Benvenuto F, et al. Predictive Factors for a New Positive Nasopharyngeal Swab Among Patients Recovered From COVID-19. American Journal of Preventive Medicine 2021; 60: 13–9.

27. Myall KJ, Mukherjee B, Castanheira AM, et al. Persistent Post–COVID-19 Interstitial Lung Disease. An Observational Study of Corticosteroid Treatment. Annals ATS 2021; 18: 799–806.

28. Niklassen AS, Draf J, Huart C, et al. COVID-19: Recovery from Chemosensory Dysfunction. A Multicentre study on Smell and Taste. The Laryngoscope 2021; 131: 1095–100.

29. Park S, Lee C-W, Park D-I, et al. Detection of SARS-CoV-2 in Fecal Samples From Patients With Asymptomatic and Mild COVID-19 in Korea. Clinical Gastroenterology and Hepatology 2021; 19: 1387–1394.e2.

30. Pellaud C, Grandmaison G, Pham Huu Thien HP, et al. Characteristics, comorbidities, 30-day outcome and in-hospital mortality of patients hospitalised with COVID-19 in a Swiss area - a retrospective cohort study. Swiss Med Wkly 2020; 150: w20314.

31. Raman B, Cassar MP, Tunnicliffe EM, et al. Medium-term effects of SARS-CoV-2 infection on multiple vital organs, exercise capacity, cognition, quality of life and mental health, post-hospital discharge. EClinicalMedicine 2021; 31: 100683.

32. Sonnweber T, Sahanic S, Pizzini A, et al. Cardiopulmonary recovery after COVID-19: an observational prospective multicentre trial. Eur Respir J 2021; 57: 2003481.

33. Taboada M, Moreno E, Cariñena A, et al. Quality of life, functional status, and persistent symptoms after intensive care of COVID-19 patients. British Journal of Anaesthesia 2021; 126: e110–3.

34. Tomasoni D, Bai F, Castoldi R, et al. Anxiety and depression symptoms after virological clearance of COVID-19: A cross-sectional study in Milan, Italy. J Med Virol 2021; 93: 1175–9.

35. Townsend L, Dowds J, O’Brien K, et al. Persistent Poor Health after COVID-19 Is Not Associated with Respiratory Complications or Initial Disease Severity. Annals ATS 2021; 18: 997–1003.

36. Townsend L, Dyer AH, Jones K, et al. Persistent fatigue following SARS-CoV-2 infection is common and independent of severity of initial infection. Madeddu G, ed. PLoS ONE 2020; 15: e0240784.

37. Vaira LA, Hopkins C, Petrocelli M, et al. Smell and taste recovery in coronavirus disease 2019 patients: a 60-day objective and prospective study. J Laryngol Otol 2020; 134: 703–9.

38. Villarreal IM, Morato M, Martínez-RuizCoello M, et al. Olfactory and taste disorders in healthcare workers with COVID-19 infection. Eur Arch Otorhinolaryngol 2021; 278: 2123–7.

39. Weerahandi H, Hochman KA, Simon E, et al. Post-Discharge Health Status and Symptoms in Patients with Severe COVID-19. J GEN INTERN MED 2021; 36: 738–45.

40. Zhao Y, Shang Y, Song W, et al. Follow-up study of the pulmonary function and related physiological characteristics of COVID-19 survivors three months after recovery. EClinicalMedicine 2020; 25: 100463.

41. Taquet M, Dercon Q, Luciano S, Geddes JR, Husain M, Harrison PJ. Incidence, co-occurrence, and evolution of long-COVID features: A 6-month retrospective cohort study of 273,618 survivors of COVID-19. Kretzschmar MEE, ed. PLoS Med 2021; 18: e1003773.

42. van Kampen JJA, van de Vijver DAMC, Fraaij PLA, et al. Duration and key determinants of infectious virus shedding in hospitalized patients with coronavirus disease-2019 (COVID-19). Nat Commun 2021; 12: 267.

43. Lopez-Leon S, Wegman-Ostrosky T, Perelman C, et al. More than 50 long-term effects of COVID-19: a systematic review and meta-analysis. Sci Rep 2021; 11: 16144.

44. Groff D, Sun A, Ssentongo AE, et al. Short-term and Long-term Rates of Postacute Sequelae of SARS-CoV-2 Infection: A Systematic Review. JAMA Netw Open 2021; 4: e2128568.

45. Sudre CH, Murray B, Varsavsky T, et al. Attributes and predictors of long COVID. Nat Med 2021; 27: 626–31.

46. Tenforde MW, Kim SS, Lindsell CJ, et al. Symptom Duration and Risk Factors for Delayed Return to Usual Health Among Outpatients with COVID-19 in a Multistate Health Care Systems Network — United States, March–June 2020. MMWR Morb Mortal Wkly Rep 2020; 69: 993–8.

47. Dennis A, Wamil M, Alberts J, et al. Multiorgan impairment in low-risk individuals with post-COVID-19 syndrome: a prospective, community-based study. BMJ Open 2021; 11: e048391.

48. Brandi ML, Giustina A. Sexual Dimorphism of Coronavirus 19 Morbidity and Lethality. Trends in Endocrinology & Metabolism 2020; 31: 918–27.

49. Sette A, Crotty S. Adaptive immunity to SARS-CoV-2 and COVID-19. Cell 2021; 184: 861–80.

50. Schultze JL, Aschenbrenner AC. COVID-19 and the human innate immune system. Cell 2021; 184: 1671–92.

51. Bastard P, Rosen LB, Zhang Q, et al. Autoantibodies against type I IFNs in patients with life-threatening COVID-19. Science 2020; 370: eabd4585.

52. Brodin P. Immune determinants of COVID-19 disease presentation and severity. Nat Med 2021; 27: 28–33.

53. Menges D, Ballouz T, Anagnostopoulos A, et al. Burden of post-COVID-19 syndrome and implications for healthcare service planning: A population-based cohort study. Simuunza MC, ed. PLoS ONE 2021; 16: e0254523.

54. Wade DT. Rehabilitation after COVID-19: an evidence-based approach. Clin Med 2020; 20: 359–65.

55. Tran V-T, Perrodeau E, Saldanha J, Pane I, Ravaud P. Efficacy of COVID-19 Vaccination on the Symptoms of Patients With Long COVID: A Target Trial Emulation Using Data From the ComPaRe e-Cohort in France. SSRN Journal 2021.

56. Arnold DT, Milne A, Samms E, Stadon L, Maskell NA, Hamilton FW. Symptoms After COVID-19 Vaccination in Patients With Persistent Symptoms After Acute Infection: A Case Series. Ann Intern Med 2021; 174: 1334–6.

57. Antonelli M, Penfold RS, Merino J, et al. Risk factors and disease profile of post-vaccination SARS-CoV-2 infection in UK users of the COVID Symptom Study app: a prospective, community-based, nested, case-control study. The Lancet Infectious Diseases 2021: S1473309921004606.

