## Supplementary material for "Long-term manifestations and modifiers of prevalence estimates of the post-COVID-19 syndrome: A systematic review and meta-analysis": Data quality

### JBI CRITICAL APPRAISAL CHECKLIST FOR TEXT AND OPINION PAPERS

Reviewer Marco Antonio Delaye Martínez Date 14/11/20

Author Klitzman Robert L. Year 2020 Record Number 1_33103966_MADM

| | Yes | No | Unclear | Not applicable |
|---|---|---|---|---|
| 1. Is the source of the opinion clearly identified? | X | ☐ | ☐ | ☐ |
| 2. Does the source of opinion have standing in the field of expertise? | ☐ | X | ☐ | ☐ |
| 3. Are the interests of the relevant population the central focus of the opinion? | ☐ | X | ☐ | ☐ |
| 4. Is the stated position the result of an analytical process, and is there logic in the opinion expressed? | ☐ | X | ☐ | ☐ |
| 5. Is there reference to the extant literature? | ☐ | X | ☐ | ☐ |
| 6. Is any incongruence with the literature/sources logically defended? | ☐ | X | ☐ | ☐ |

Overall appraisal: Include ☐ Exclude X Seek further info ☐

Comments (Including reason for exclusion)

_______________________________________________

_______________________________________________

_______________________________________________

© JBI, 2020. All rights reserved. JBI grants use of these tools for research purposes only. All other enquiries should be sent to.

### JBI CRITICAL APPRAISAL CHECKLIST FOR CASE REPORTS

Reviewer Marco Antonio Delaye Martínez Date 15/11/20

Author Hoon Baang I., Smith C., Mirabelli C., et al. Year 2020 Record Number 2_33089317_MADM

| | Yes | No | Unclear | Not applicable |
|---|---|---|---|---|
| 1. Were patient's demographic characteristics clearly described? | ■ | ☐ | ☐ | ☐ |
| 2. Was the patient's history clearly described and presented as a timeline? | ■ | ☐ | ☐ | ☐ |
| 3. Was the current clinical condition of the patient on presentation clearly described? | ■ | ☐ | ☐ | ☐ |
| 4. Were diagnostic tests or assessment methods and the results clearly described? | ■ | ☐ | ☐ | ☐ |
| 5. Was the intervention(s) or treatment procedure(s) clearly described? | ■ | ☐ | ☐ | ☐ |
| 6. Was the post-intervention clinical condition clearly described? | ■ | ☐ | ☐ | ☐ |
| 7. Were adverse events (harms) or unanticipated events identified and described? | ☐ | ☐ | ■ | ☐ |
| 8. Does the case report provide takeaway lessons? | ■ | ☐ | ☐ | ☐ |

Overall appraisal: Include ☐ Exclude ■ Seek further info ☐

Comments (Including reason for exclusion)

No existe claridad en si los síntomas reportados son producto de COVID-19, la enfermedad de base del paciente (linfoma de células del manto) o el tratamiento administrado. Así mismo, no aporta información útil para los propósitos de nuestra revisión sistemática.

© JBI, 2020. All rights reserved. JBI grants use of these tools for research purposes only. All other enquiries should be sent to.

### JBI CRITICAL APPRAISAL CHECKLIST FOR COHORT STUDIES

Reviewer: Daniel de la Rosa  Date: ______

Author: Thomas Sonnweber et al  Year: 2020  Record Number: 3_33087116

| | Yes | No | Unclear | Not applicable |
|---|---|---|---|---|
| 1. Were the two groups similar and recruited from the same population? | ☑ | ☐ | ☐ | ☐ |
| 2. Were the exposures measured similarly to assign people to both exposed and unexposed groups? | ☑ | ☐ | ☐ | ☐ |
| 3. Was the exposure measured in a valid and reliable way? | ☑ | ☐ | ☐ | ☐ |
| 4. Were confounding factors identified? | ☐ | ☐ | ☑ | ☐ |
| 5. Were strategies to deal with confounding factors stated? | ☐ | ☐ | ☑ | ☐ |
| 6. Were the groups/participants free of the outcome at the start of the study (or at the moment of exposure)? | ☑ | ☐ | ☐ | ☐ |
| 7. Were the outcomes measured in a valid and reliable way? | ☑ | ☐ | ☐ | ☐ |
| 8. Was the follow up time reported and sufficient to be long enough for outcomes to occur? | ☐ | ☐ | ☑ | ☐ |
| 9. Was follow up complete, and if not, were the reasons to loss to follow up described and explored? | ☐ | ☐ | ☑ | ☐ |
| 10. Were strategies to address incomplete follow up utilized? | ☐ | ☐ | ☑ | ☐ |
| 11. Was appropriate statistical analysis used? | ☑ | ☐ | ☐ | ☐ |

Overall appraisal: Include ☐ Exclude ☑ Seek further info ☐

Comments (Including reason for exclusion)

30% insuficiencia de hierro, 13% deficiencia de hierro absoluta, 17% deficiencia de hierro funcional, anemia 9.2%. Se encontró una asociación positiva significativa entre anemia y enfermedad crítica por COVID-19, y anemia y citocinas proinflamatorias (IL-6, TNF-aplha e IL-10). El 38% de los pacientes presentaba hiperferritinemia (48% de los hombres y 23% de las mujeres). Los niveles de ferritina fueron mayores en pacientes con enfermedad severa (p=0.001) y la hiperferritinemia se asoció con un mayor CT-Score (p=0.001). No se encontró una asociaición entre hiperferritinemia y edad, género, comorbilidades.

© JBI, 2020. All rights reserved. JBI grants use of these tools for research purposes only. All other enquiries should be sent to.

### JBI CRITICAL APPRAISAL CHECKLIST FOR CASE REPORTS

Reviewer Marco Antonio Delaye Martínez Date 15/11/20

Author Buselli R., Corsi M., Necciari G., et al. Year 2020 Record Number 4_33078141_MADM

| | Yes | No | Unclear | Not applicable |
|---|---|---|---|---|
| 1. Were patient's demographic characteristics clearly described? | ● | ☐ | ☐ | ☐ |
| 2. Was the patient's history clearly described and presented as a timeline? | ● | ☐ | ☐ | ☐ |
| 3. Was the current clinical condition of the patient on presentation clearly described? | ◒ | ☐ | ☐ | ☐ |
| 4. Were diagnostic tests or assessment methods and the results clearly described? | ◗ | ☐ | ☐ | ☐ |
| 5. Was the intervention(s) or treatment procedure(s) clearly described? | ☐ | ◗ | ☐ | ☐ |
| 6. Was the post-intervention clinical condition clearly described? | ☐ | ● | ☐ | ☐ |
| 7. Were adverse events (harms) or unanticipated events identified and described? | ☐ | ● | ☐ | ☐ |
| 8. Does the case report provide takeaway lessons? | ☐ | ☐ | ◗ | ☐ |

Overall appraisal: Include ☐ Exclude ● Seek further info ☐

Comments (Including reason for exclusion)

En el articulo la disfonia está asociada a un trastorno por somatización que no sé explica por COVID-19. De incluirlo, la disfonia podría comportarse como un factor confusor.

© JBI, 2020. All rights reserved. JBI grants use of these tools for research purposes only. All other enquiries should be sent to.

### JBI CRITICAL APPRAISAL CHECKLIST FOR TEXT AND OPINION PAPERS

Reviewer Marco Antonio Delaye Martínez Date 14/11/20

Author Ricardo J. José, Manuel A. et. al. Year 2020 Record Number 5_33054432_MAD

| | Yes | No | Unclear | Not applicable |
|---|---|---|---|---|
| 1. Is the source of the opinion clearly identified? | ■ | □ | □ | □ |
| 2. Does the source of opinion have standing in the field of expertise? | ● | □ | □ | □ |
| 3. Are the interests of the relevant population the central focus of the opinion? | ● | □ | □ | □ |
| 4. Is the stated position the result of an analytical process, and is there logic in the opinion expressed? | ● | □ | □ | □ |
| 5. Is there reference to the extant literature? | ● | □ | □ | □ |
| 6. Is any incongruence with the literature/sources logically defended? | □ | ● | □ | □ |

Overall appraisal: Include □ Exclude ■ Seek further info □

Comments (Including reason for exclusion)

No aporta información concreta para los objetivos específicos de la presente revisión sistemática. Aún existen limitaciones en determinar si la presencia de bronquiectasias asociadas a COVID-19 representan un hallazgo consistente para "Long-COVID"

© JBI, 2020. All rights reserved. JBI grants use of these tools for research purposes only. All other enquiries should be sent to.

### JBI CRITICAL APPRAISAL CHECKLIST FOR COHORT STUDIES

Reviewer Marco Antonio Delaye Martínez Date 25/11/20

Author Bhattacharya B., Kumar R., Prakash V., et al. Year 2020 Record Number 6_33053181

| | Yes | No | Unclear | Not applicable |
|---|---|---|---|---|
| 1. Were the two groups similar and recruited from the same population? | ☐ | ☐ | ☐ | ■ |
| 2. Were the exposures measured similarly to assign people to both exposed and unexposed groups? | ☐ | ☐ | ☐ | ■ |
| 3. Was the exposure measured in a valid and reliable way? | ■ | ☐ | ☐ | ☐ |
| 4. Were confounding factors identified? | ☐ | ■ | ☐ | ☐ |
| 5. Were strategies to deal with confounding factors stated? | ☐ | ■ | ☐ | ☐ |
| 6. Were the groups/participants free of the outcome at the start of the study (or at the moment of exposure)? | ☐ | ■ | ☐ | ☐ |
| 7. Were the outcomes measured in a valid and reliable way? | ☐ | ■ | ☐ | ☐ |
| 8. Was the follow up time reported and sufficient to be long enough for outcomes to occur? | ☐ | ■ | ☐ | ☐ |
| 9. Was follow up complete, and if not, were the reasons to loss to follow up described and explored? | ■ | ☐ | ☐ | ☐ |
| 10. Were strategies to address incomplete follow up utilized? | ■ | ☐ | ☐ | ☐ |
| 11. Was appropriate statistical analysis used? | ☐ | ■ | ☐ | ☐ |

Overall appraisal: Include ☐ Exclude ■ Seek further info ☐

Comments (Including reason for exclusion)

Existen errores metodológicos en la presentación e interpretación de datos, lo que lo hace un artículo poco confiable. No reporta información relevante relacionada a longcovid.

© JBI, 2020. All rights reserved. JBI grants use of these tools for research purposes only. All other enquiries should be sent to.

### JBI CRITICAL APPRAISAL CHECKLIST FOR TEXT AND OPINION PAPERS

Reviewer Marco Antonio Delaye Martínez Date 23/11/20

Author Wijeratne T., Crewther S. Year 2020 Record Number 7_33070003

| | Yes | No | Unclear | Not applicable |
|---|---|---|---|---|
| 1. Is the source of the opinion clearly identified? | ● | ☐ | ☐ | ☐ |
| 2. Does the source of opinion have standing in the field of expertise? | ● | ☐ | ☐ | ☐ |
| 3. Are the interests of the relevant population the central focus of the opinion? | ● | ☐ | ☐ | ☐ |
| 4. Is the stated position the result of an analytical process, and is there logic in the opinion expressed? | ● | ☐ | ☐ | ☐ |
| 5. Is there reference to the extant literature? | ● | ☐ | ☐ | ☐ |
| 6. Is any incongruence with the literature/sources logically defended? | ☐ | ● | ☐ | ☐ |

Overall appraisal: Include ☐ Exclude ■ Seek further info ☐

Comments (Including reason for exclusion)

Por ser artículo de opinión, no se incluye dentro de la revisión sistemática, sin embargo, el artículo es útil para se incluido en la discusión o introducción.

© JBI, 2020. All rights reserved. JBI grants use of these tools for research purposes only. All other enquiries should be sent to.

### JBI CRITICAL APPRAISAL CHECKLIST FOR QUALITATIVE RESEARCH

Reviewer Marco Antonio Delaye Martínez Date 23/11/20

Author Kingstone T., Taylor A., O'Donell C., et al. Year 2020 Record Number 8_33051223

| | Yes | No | Unclear | Not applicable |
|---|---|---|---|---|
| 1. Is there congruity between the stated philosophical perspective and the research methodology? | ■ | □ | □ | □ |
| 2. Is there congruity between the research methodology and the research question or objectives? | ■ | □ | □ | □ |
| 3. Is there congruity between the research methodology and the methods used to collect data? | ■ | □ | □ | □ |
| 4. Is there congruity between the research methodology and the representation and analysis of data? | ■ | □ | □ | □ |
| 5. Is there congruity between the research methodology and the interpretation of results? | ■ | □ | □ | □ |
| 6. Is there a statement locating the researcher culturally or theoretically? | □ | □ | ■ | □ |
| 7. Is the influence of the researcher on the research, and vice- versa, addressed? | ■ | □ | □ | □ |
| 8. Are participants, and their voices, adequately represented? | ■ | □ | □ | □ |
| 9. Is the research ethical according to current criteria or, for recent studies, and is there evidence of ethical approval by an appropriate body? | ■ | □ | □ | □ |
| 10. Do the conclusions drawn in the research report flow from the analysis, or interpretation, of the data? | ■ | □ | □ | □ |

Overall appraisal: Include □ Exclude ■ Seek further info □

Comments (Including reason for exclusion)

No reporta estimadores en relación a la persistencia de síntomas persistentes, sin embargo, aporta información útil para incluir en discusión o análisis.

© JBI, 2020. All rights reserved. JBI grants use of these tools for research purposes only. All other enquiries should be sent to.

### JBI CRITICAL APPRAISAL CHECKLIST FOR COHORT STUDIES

Reviewer____DDMR______________________________Date____29 11 20__________

Author Konstantinidisa et al. ______Year_2020_ Record Number_ 9_33049753________

| | Yes | No | Unclear | Not applicable |
|---|---|---|---|---|
| 1. Were the two groups similar and recruited from the same population? | ☒ | ☐ | ☐ | ☐ |
| 2. Were the exposures measured similarly to assign people to both exposed and unexposed groups? | ☒ | ☐ | ☐ | ☐ |
| 3. Was the exposure measured in a valid and reliable way? | ☒ | ☐ | ☐ | ☐ |
| 4. Were confounding factors identified? | ☐ | ☐ | ☒ | ☐ |
| 5. Were strategies to deal with confounding factors stated? | ☒ | ☐ | ☐ | ☐ |
| 6. Were the groups/participants free of the outcome at the start of the study (or at the moment of exposure)? | ☒ | ☐ | ☐ | ☐ |
| 7. Were the outcomes measured in a valid and reliable way? | ☐ | ☐ | ☒ | ☐ |
| 8. Was the follow up time reported and sufficient to be long enough for outcomes to occur? | ☒ | ☐ | ☐ | ☐ |
| 9. Was follow up complete, and if not, were the reasons to loss to follow up described and explored? | ☒ | ☐ | ☐ | ☐ |
| 10. Were strategies to address incomplete follow up utilized? | ☒ | ☐ | ☐ | ☐ |
| 11. Was appropriate statistical analysis used? | ☒ | ☐ | ☐ | ☐ |

Overall appraisal: Include ☒ Exclude ☐ Seek further info ☐

Comments (Including reason for exclusion)

______________________________________________________________

______________________________________________________________

### JBI CRITICAL APPRAISAL CHECKLIST FOR SYSTEMATIC REVIEWS AND RESEARCH SYNTHESES

Reviewer Daniel de la Rosa Date 14-11-20

Author Franziska Weichmanna and Peter Rohdewaldb Year 2020 Record Number 10_33045354

| | | Yes | No | Unclear | Not applicable |
|---|---|---|---|---|---|
| 1. | Is the review question clearly and explicitly stated? | ☑ | ☐ | ☐ | ☐ |
| 2. | Were the inclusion criteria appropriate for the review question? | ☐ | ☐ | ☐ | ☑ |
| 3. | Was the search strategy appropriate? | ☐ | ☐ | ☐ | ☑ |
| 4. | Were the sources and resources used to search for studies adequate? | ☐ | ☐ | ☐ | ☑ |
| 5. | Were the criteria for appraising studies appropriate? | ☐ | ☐ | ☐ | ☑ |
| 6. | Was critical appraisal conducted by two or more reviewers independently? | ☐ | ☐ | ☐ | ☑ |
| 7. | Were there methods to minimize errors in data extraction? | ☐ | ☐ | ☐ | ☑ |
| 8. | Were the methods used to combine studies appropriate? | ☐ | ☐ | ☐ | ☑ |
| 9. | Was the likelihood of publication bias assessed? | ☐ | ☐ | ☐ | ☑ |
| 10. | Were recommendations for policy and/or practice supported by the reported data? | ☑ | ☐ | ☐ | ☐ |
| 11. | Were the specific directives for new research appropriate? | ☑ | ☐ | ☐ | ☐ |

Overall appraisal: Include ☐ Exclude ☑ Seek further info ☐

Comments (Including reason for exclusion)

### JBI Critical Appraisal Checklist for Case Series

Reviewer Marco Antonio Delaye Martínez Date 29/11/20

Author Wootton S., King M., Alison J., et al. Year 2020 Record Number 11_33042547

| | Yes | No | Unclear | Not applicable |
|---|---|---|---|---|
| • Were there clear criteria for inclusion in the case series? | ☐ | ● | ☐ | ☐ |
| • Was the condition measured in a standard, reliable way for all participants included in the case series? | ● | ☐ | ☐ | ☐ |
| • Were valid methods used for identification of the condition for all participants included in the case series? | ● | ☐ | ☐ | ☐ |
| • Did the case series have consecutive inclusion of participants? | ● | ☐ | ☐ | ☐ |
| • Did the case series have complete inclusion of participants? | ● | ☐ | ☐ | ☐ |
| • Was there clear reporting of the demographics of the participants in the study? | ● | ☐ | ☐ | ☐ |
| • Was there clear reporting of clinical information of the participants? | ● | ☐ | ☐ | ☐ |
| • Were the outcomes or follow up results of cases clearly reported? | ● | ☐ | ☐ | ☐ |
| • Was there clear reporting of the presenting site(s)/clinic(s) demographic information? | ☐ | ☐ | ● | ☐ |
| • Was statistical analysis appropriate? | ☐ | ● | ☐ | ☐ |

Overall appraisal: Include ☐ Exclude ● Seek further info ☐

Comments (Including reason for exclusion)

Se excluye por ser serie de casos. Sin embargo, contiene información útil para la discusión.

© JBI, 2020. All rights reserved. JBI grants use of these tools for research purposes only. All other enquiries should be sent to.

### JBI CRITICAL APPRAISAL CHECKLIST FOR COHORT STUDIES

Reviewer Marco Antonio Delaye Martinez Date 29/11/20

Author Afshar Y., Gaw S., Flaherman V., et al Year 2020 Record Number 12_33027186

| | Yes | No | Unclear | Not applicable |
|---|---|---|---|---|
| 1. Were the two groups similar and recruited from the same population? | ☐ | ☐ | ☐ | ☒ |
| 2. Were the exposures measured similarly to assign people to both exposed and unexposed groups? | ☐ | ☐ | ☐ | ☒ |
| 3. Was the exposure measured in a valid and reliable way? | ☒ | ☐ | ☐ | ☐ |
| 4. Were confounding factors identified? | ☒ | ☐ | ☐ | ☐ |
| 5. Were strategies to deal with confounding factors stated? | ☐ | ☐ | ☒ | ☐ |
| 6. Were the groups/participants free of the outcome at the start of the study (or at the moment of exposure)? | ☒ | ☐ | ☐ | ☐ |
| 7. Were the outcomes measured in a valid and reliable way? | ☒ | ☐ | ☐ | ☐ |
| 8. Was the follow up time reported and sufficient to be long enough for outcomes to occur? | ☒ | ☐ | ☐ | ☐ |
| 9. Was follow up complete, and if not, were the reasons to loss to follow up described and explored? | ☒ | ☐ | ☐ | ☐ |
| 10. Were strategies to address incomplete follow up utilized? | ☒ | ☐ | ☐ | ☐ |
| 11. Was appropriate statistical analysis used? | ☐ | ☐ | ☐ | ☒ |

Overall appraisal: Include ☐ Exclude ☒ Seek further info ☐

Comments (Including reason for exclusion)

No comments

© JBI, 2020. All rights reserved. JBI grants use of these tools for research purposes only. All other enquiries should be sent to.

### JBI CRITICAL APPRAISAL CHECKLIST FOR COHORT STUDIES

Reviewer Marco Antonio Delaye Martínez Date 29/11/20

Author Ramasamy K., Saniasiaya J., Abdul N. Year 2020 Record Number 13_33025798

| | Yes | No | Unclear | Not applicable |
|---|---|---|---|---|
| 1. Were the two groups similar and recruited from the same population? | ☐ | ☐ | ☐ | ● |
| 2. Were the exposures measured similarly to assign people to both exposed and unexposed groups? | ☐ | ☐ | ☐ | ● |
| 3. Was the exposure measured in a valid and reliable way? | ☐ | ● | ☐ | ☐ |
| 4. Were confounding factors identified? | ☐ | ● | ☐ | ☐ |
| 5. Were strategies to deal with confounding factors stated? | ☐ | ● | ☐ | ☐ |
| 6. Were the groups/participants free of the outcome at the start of the study (or at the moment of exposure)? | ● | ☐ | ☐ | ☐ |
| 7. Were the outcomes measured in a valid and reliable way? | ● | ☐ | ☐ | ☐ |
| 8. Was the follow up time reported and sufficient to be long enough for outcomes to occur? | ☐ | ● | ☐ | ☐ |
| 9. Was follow up complete, and if not, were the reasons to loss to follow up described and explored? | ● | ☐ | ☐ | ☐ |
| 10. Were strategies to address incomplete follow up utilized? | ☐ | ● | ☐ | ☐ |
| 11. Was appropriate statistical analysis used? | ● | ☐ | ☐ | ☐ |

Overall appraisal: Include ☐ Exclude ● Seek further info ☐

Comments (Including reason for exclusion)

El estudio se centra en determinar la prevalencia y el grado de severidad de anemia y agencia como síntomas que conforman el cuadro agudo de COVID-19. No aporta información útil para COVID prolongado; el tiempo de seguimiento es muy corto.

© JBI, 2020. All rights reserved. JBI grants use of these tools for research purposes only. All other enquiries should be sent to.

### JBI CRITICAL APPRAISAL CHECKLIST FOR COHORT STUDIES

Reviewer Marco Antonio Delaye Martínez Date 29/11/20

Author Carvalho-Schneider C., Laurent E., Lemaignen A., et al. Year 2020 Record Number 14_33031948

| | Yes | No | Unclear | Not applicable |
|---|---|---|---|---|
| 1. Were the two groups similar and recruited from the same population? | ☐ | ☐ | ☐ | ■ |
| 2. Were the exposures measured similarly to assign people to both exposed and unexposed groups? | ☐ | ☐ | ☐ | ■ |
| 3. Was the exposure measured in a valid and reliable way? | ■ | ☐ | ☐ | ☐ |
| 4. Were confounding factors identified? | ■ | ☐ | ☐ | ☐ |
| 5. Were strategies to deal with confounding factors stated? | ■ | ☐ | ☐ | ☐ |
| 6. Were the groups/participants free of the outcome at the start of the study (or at the moment of exposure)? | ■ | ☐ | ☐ | ☐ |
| 7. Were the outcomes measured in a valid and reliable way? | ■ | ☐ | ☐ | ☐ |
| 8. Was the follow up time reported and sufficient to be long enough for outcomes to occur? | ■ | ☐ | ☐ | ☐ |
| 9. Was follow up complete, and if not, were the reasons to loss to follow up described and explored? | ■ | ☐ | ☐ | ☐ |
| 10. Were strategies to address incomplete follow up utilized? | ■ | ☐ | ☐ | ☐ |
| 11. Was appropriate statistical analysis used? | ■ | ☐ | ☐ | ☐ |

Overall appraisal: Include ■ Exclude ☐ Seek further info ☐

Comments (Including reason for exclusion)

No comments.

© JBI, 2020. All rights reserved. JBI grants use of these tools for research purposes only. All other enquiries should be sent to.

### JBI CRITICAL APPRAISAL CHECKLIST FOR COHORT STUDIES

Reviewer Marco Antonio Delaye Martínez Date 30/11/20

Author S. Belli et al. Year 2020 Record Number 15_32764112

| | Yes | No | Unclear | Not applicable |
|---|---|---|---|---|
| 1. Were the two groups similar and recruited from the same population? | | | | X |
| 2. Were the exposures measured similarly to assign people to both exposed and unexposed groups? | | | | X |
| 3. Was the exposure measured in a valid and reliable way? | X | | | |
| 4. Were confounding factors identified? | | | X | |
| 5. Were strategies to deal with confounding factors stated? | | | X | |
| 6. Were the groups/participants free of the outcome at the start of the study (or at the moment of exposure)? | X | | | |
| 7. Were the outcomes measured in a valid and reliable way? | X | | | |
| 8. Was the follow up time reported and sufficient to be long enough for outcomes to occur? | X | | | |
| 9. Was follow up complete, and if not, were the reasons to loss to follow up described and explored? | X | | | |
| 10. Were strategies to address incomplete follow up utilized? | | X | | |
| 11. Was appropriate statistical analysis used? | X | | | |

Overall appraisal: Include ☐ Exclude ☒ Seek further info ☐

Comments (Including reason for exclusion)

No reporta la persistencia de síntomas asociados a COVID, según la definición de Long-COVID. Sin embargo, evalúa de forma de integral la disfunción física y capacidad para realizar las ADLs. Es útil para discusión.

© JBI, 2020. All rights reserved. JBI grants use of these tools for research purposes only. All other enquiries should be sent to.

### JBI CRITICAL APPRAISAL CHECKLIST FOR TEXT AND OPINION PAPERS

Reviewer Daniel de la Rosa Date 14/11/20

Author Jérôme René Lechien et al Year 2020 Record Number 16_33000300

| | Yes | No | Unclear | Not applicable |
|---|---|---|---|---|
| 1. Is the source of the opinion clearly identified? | ☒ | ☐ | ☐ | ☐ |
| 2. Does the source of opinion have standing in the field of expertise? | ☒ | ☐ | ☐ | ☐ |
| 3. Are the interests of the relevant population the central focus of the opinion? | ☒ | ☐ | ☐ | ☐ |
| 4. Is the stated position the result of an analytical process, and is there logic in the opinion expressed? | ☒ | ☐ | ☐ | ☐ |
| 5. Is there reference to the extant literature? | ☒ | ☐ | ☐ | ☐ |
| 6. Is any incongruence with the literature/sources logically defended? | ☐ | ☐ | ☒ | ☐ |

Overall appraisal: Include ☐ Exclude ☒ Seek further info ☐

Comments (Including reason for exclusion)

© JBI, 2020. All rights reserved. JBI grants use of these tools for research purposes only. All other enquiries should be sent to.

### JBI CRITICAL APPRAISAL CHECKLIST FOR TEXT AND OPINION PAPERS

Reviewer Daniel de la Rosa Date 20/11/20

Author Espen Lindholm Year 2020 Record Number 17_33070586_DDM

| | Yes | No | Unclear | Not applicable |
|---|---|---|---|---|
| 1. Is the source of the opinion clearly identified? | ☒ | ☐ | ☐ | ☐ |
| 2. Does the source of opinion have standing in the field of expertise? | ☒ | ☐ | ☐ | ☐ |
| 3. Are the interests of the relevant population the central focus of the opinion? | ☐ | ☐ | ☒ | ☐ |
| 4. Is the stated position the result of an analytical process, and is there logic in the opinion expressed? | ☐ | ☒ | ☐ | ☐ |
| 5. Is there reference to the extant literature? | ☒ | ☐ | ☐ | ☐ |
| 6. Is any incongruence with the literature/sources logically defended? | ☐ | ☐ | ☒ | ☐ |

Overall appraisal: Include ☐ Exclude ☒ Seek further info ☐

Comments (Including reason for exclusion)

© JBI, 2020. All rights reserved. JBI grants use of these tools for research purposes only. All other enquiries should be sent to.

### JBI CRITICAL APPRAISAL CHECKLIST FOR ANALYTICAL CROSS SECTIONAL STUDIES

Reviewer______Daniel de la Rosa Martinez______Date________29/11/20______________

Author_________Kamal et al____________Year__2020___ Record Number__ 18_32991035_______

| | Yes | No | Unclear | Not applicable |
|---|---|---|---|---|
| 1. Were the criteria for inclusion in the sample clearly defined? | ■ | □ | □ | □ |
| 2. Were the study subjects and the setting described in detail? | ■ | □ | □ | □ |
| 3. Was the exposure measured in a valid and reliable way? | ■ | □ | □ | □ |
| 4. Were objective, standard criteria used for measurement of the condition? | ■ | □ | □ | □ |
| 5. Were confounding factors identified? | □ | □ | ■ | □ |
| 6. Were strategies to deal with confounding factors stated? | □ | □ | ■ | □ |
| 7. Were the outcomes measured in a valid and reliable way? | ■ | □ | □ | □ |
| 8. Was appropriate statistical analysis used? | ■ | □ | □ | □ |

Overall appraisal: Include ■ Exclude □ Seek further info □

Comments (Including reason for exclusion)

______________________________________________

______________________________________________

______________________________________________

### EXPLANATION OF ANALYTICAL CROSS SECTIONAL STUDIES CRITICAL APPRAISAL

*How to cite:* Moola S, Munn Z, Tufanaru C, Aromataris E, Sears K, Sfetcu R, Currie M, Qureshi R, Mattis P, Lisy K, Mu P-F. Chapter 7: Systematic reviews of etiology and risk . In: Aromataris E, Munn Z (Editors). *JBI Manual for Evidence Synthesis*. JBI, 2020. Available from https://synthesismanual.jbi.global

**Analytical cross sectional studies Critical Appraisal Tool**

Answers: Yes, No, Unclear or Not/Applicable

#### 1. Were the criteria for inclusion in the sample clearly defined?

The authors should provide clear inclusion and exclusion criteria that they developed prior to recruitment of the study participants. The inclusion/exclusion criteria should be specified (e.g., risk, stage of disease progression) with sufficient detail and all the necessary information critical to the study.

#### 2. Were the study subjects and the setting described in detail?

The study sample should be described in sufficient detail so that other researchers can determine if it is comparable to the population of interest to them. The authors should provide a clear description of the population from which the study participants were selected or recruited, including demographics, location, and time period.

#### 3. Was the exposure measured in a valid and reliable way?

The study should clearly describe the method of measurement of exposure. Assessing validity requires that a 'gold standard' is available to which the measure can be compared. The validity of exposure measurement usually relates to whether a current measure is appropriate or whether a measure of past exposure is needed.

Reliability refers to the processes included in an epidemiological study to check repeatability of measurements of the exposures. These usually include intra-observer reliability and inter-observer reliability.

#### 4. Were objective, standard criteria used for measurement of the condition?

It is useful to determine if patients were included in the study based on either a specified diagnosis or definition. This is more likely to decrease the risk of bias. Characteristics are another useful approach to matching groups, and studies that did not use specified diagnostic methods or definitions should provide evidence on matching by key characteristics

#### 5. Were confounding factors identified?

Confounding has occurred where the estimated intervention exposure effect is biased by the presence of some difference between the comparison groups (apart from the exposure investigated/of interest). Typical confounders include baseline characteristics, prognostic factors, or concomitant exposures (e.g. smoking). A confounder is a difference between the comparison groups and it influences the direction of the study results. A high quality study at the level of cohort design will identify the potential confounders and measure them (where possible). This is difficult for studies where behavioral, attitudinal or lifestyle factors may impact on the results.

#### 6. Were strategies to deal with confounding factors stated?

Strategies to deal with effects of confounding factors may be dealt within the study design or in data analysis. By matching or stratifying sampling of participants, effects of confounding factors can be adjusted for. When dealing with adjustment in data analysis, assess the statistics used in the study. Most will be some form of multivariate regression analysis to account for the confounding factors measured.

© JBI, 2020. All rights reserved. JBI grants use of these tools for research purposes only. All other enquiries should be sent to.

#### 7. Were the outcomes measured in a valid and reliable way?

Read the methods section of the paper. If for e.g. lung cancer is assessed based on existing definitions or diagnostic criteria, then the answer to this question is likely to be yes. If lung cancer is assessed using observer reported, or self-reported scales, the risk of over- or under-reporting is increased, and objectivity is compromised. Importantly, determine if the measurement tools used were validated instruments as this has a significant impact on outcome assessment validity.

Having established the objectivity of the outcome measurement (e.g. lung cancer) instrument, it's important to establish how the measurement was conducted. Were those involved in collecting data trained or educated in the use of the instrument/s? (e.g. radiographers). If there was more than one data collector, were they similar in terms of level of education, clinical or research experience, or level of responsibility in the piece of research being appraised?

#### 8. Was appropriate statistical analysis used?

As with any consideration of statistical analysis, consideration should be given to whether there was a more appropriate alternate statistical method that could have been used. The methods section should be detailed enough for reviewers to identify which analytical techniques were used (in particular, regression or stratification) and how specific confounders were measured.

For studies utilizing regression analysis, it is useful to identify if the study identified which variables were included and how they related to the outcome. If stratification was the analytical approach used, were the strata of analysis defined by the specified variables? Additionally, it is also important to assess the appropriateness of the analytical strategy in terms of the assumptions associated with the approach as differing methods of analysis are based on differing assumptions about the data and how it will respond.

© JBI, 2020. All rights reserved. JBI grants use of these tools for research purposes only. All other enquiries should be sent to.

### JBI CRITICAL APPRAISAL CHECKLIST FOR SYSTEMATIC REVIEWS AND RESEARCH SYNTHESES

Reviewer Daniel de la Rosa Date 21-11-20

Author Dmitri Bougakov et al Year 2020 Record Number 19_32990925_DDM

| | | Yes | No | Unclear | Not applicable |
|---|---|---|---|---|---|
| 1. | Is the review question clearly and explicitly stated? | ☑ | ☐ | ☐ | ☐ |
| 2. | Were the inclusion criteria appropriate for the review question? | ☐ | ☐ | ☐ | ☑ |
| 3. | Was the search strategy appropriate? | ☐ | ☐ | ☐ | ☑ |
| 4. | Were the sources and resources used to search for studies adequate? | ☐ | ☐ | ☐ | ☑ |
| 5. | Were the criteria for appraising studies appropriate? | ☐ | ☐ | ☐ | ☑ |
| 6. | Was critical appraisal conducted by two or more reviewers independently? | ☐ | ☐ | ☐ | ☑ |
| 7. | Were there methods to minimize errors in data extraction? | ☐ | ☐ | ☐ | ☑ |
| 8. | Were the methods used to combine studies appropriate? | ☐ | ☐ | ☐ | ☑ |
| 9. | Was the likelihood of publication bias assessed? | ☐ | ☐ | ☐ | ☑ |
| 10. | Were recommendations for policy and/or practice supported by the reported data? | ☑ | ☐ | ☐ | ☐ |
| 11. | Were the specific directives for new research appropriate? | ☑ | ☐ | ☐ | ☐ |

Overall appraisal: Include ☐ Exclude ☑ Seek further info ☐

Comments (Including reason for exclusion)

___

___

___

### JBI CRITICAL APPRAISAL CHECKLIST FOR ANALYTICAL CROSS SECTIONAL STUDIES

Reviewer Daniel de la Rosa Date 20-11-20

Author Karimi-Galougahi et al Year 2020 Record Number 20_33014751

| | Yes | No | Unclear | Not applicable |
|---|---|---|---|---|
| 1. Were the criteria for inclusion in the sample clearly defined? | ☒ | ☐ | ☐ | ☐ |
| 2. Were the study subjects and the setting described in detail? | ☒ | ☐ | ☐ | ☐ |
| 3. Was the exposure measured in a valid and reliable way? | ☒ | ☐ | ☐ | ☐ |
| 4. Were objective, standard criteria used for measurement of the condition? | ☒ | ☐ | ☐ | ☐ |
| 5. Were confounding factors identified? | ☐ | ☐ | ☒ | ☐ |
| 6. Were strategies to deal with confounding factors stated? | ☐ | ☐ | ☒ | ☐ |
| 7. Were the outcomes measured in a valid and reliable way? | ☒ | ☐ | ☐ | ☐ |
| 8. Was appropriate statistical analysis used? | ☒ | ☐ | ☐ | ☐ |

Overall appraisal: Include ☐ Exclude ☒ Seek further info ☐

Comments (Including reason for exclusion)

Follow up time less than 21 days

### JBI CRITICAL APPRAISAL CHECKLIST FOR SYSTEMATIC REVIEWS AND RESEARCH SYNTHESES

Reviewer Daniel de la Rosa Date 21-11-20

Author Scott Rooney, et al Year 2020 Record Number 21_32737507

| | | Yes | No | Unclear | Not applicable |
|---|---|---|---|---|---|
| 1. | Is the review question clearly and explicitly stated? | ☒ | ☐ | ☐ | ☐ |
| 2. | Were the inclusion criteria appropriate for the review question? | ☐ | ☐ | ☐ | ☒ |
| 3. | Was the search strategy appropriate? | ☐ | ☐ | ☐ | ☒ |
| 4. | Were the sources and resources used to search for studies adequate? | ☐ | ☐ | ☐ | ☒ |
| 5. | Were the criteria for appraising studies appropriate? | ☐ | ☐ | ☐ | ☒ |
| 6. | Was critical appraisal conducted by two or more reviewers independently? | ☐ | ☐ | ☐ | ☒ |
| 7. | Were there methods to minimize errors in data extraction? | ☐ | ☐ | ☐ | ☒ |
| 8. | Were the methods used to combine studies appropriate? | ☐ | ☐ | ☐ | ☒ |
| 9. | Was the likelihood of publication bias assessed? | ☐ | ☐ | ☐ | ☒ |
| 10. | Were recommendations for policy and/or practice supported by the reported data? | ☒ | ☐ | ☐ | ☐ |
| 11. | Were the specific directives for new research appropriate? | ☒ | ☐ | ☐ | ☐ |

Overall appraisal: Include ☐ Exclude ☒ Seek further info ☐

Comments (Including reason for exclusion)

### JBI CRITICAL APPRAISAL CHECKLIST FOR ANALYTICAL CROSS SECTIONAL STUDIES

Reviewer: Daniel de la Rosa    Date: 22-11-20

Author: Antonio Rosales-Castillo+,    Year: 2020    Record Number: 22_33077167

| | Yes | No | Unclear | Not applicable |
|---|---|---|---|---|
| 1. Were the criteria for inclusion in the sample clearly defined? | ☐ | X | ☐ | ☐ |
| 2. Were the study subjects and the setting described in detail? | X | ☐ | ☐ | ☐ |
| 3. Was the exposure measured in a valid and reliable way? | X | ☐ | ☐ | ☐ |
| 4. Were objective, standard criteria used for measurement of the condition? | X | ☐ | ☐ | ☐ |
| 5. Were confounding factors identified? | ☐ | X | ☐ | ☐ |
| 6. Were strategies to deal with confounding factors stated? | ☐ | X | ☐ | ☐ |
| 7. Were the outcomes measured in a valid and reliable way? | X | ☐ | ☐ | ☐ |
| 8. Was appropriate statistical analysis used? | X | ☐ | ☐ | ☐ |

Overall appraisal: Include ☐ Exclude X Seek further info ☐

Comments (Including reason for exclusion)

"El tiempo de seguimiento no se especifica para todos los pacienes, ademas el metodo diagnostico no fue PCR para todos."

### JBI CRITICAL APPRAISAL CHECKLIST FOR CASE REPORTS

Reviewer Marco Antonio Delaye Martínez Date 31/12/20

Author Novak P. Year 2020 Record Number 23_32984564

| | Yes | No | Unclear | Not applicable |
|---|---|---|---|---|
| 1. Were patient's demographic characteristics clearly described? | X | | | |
| 2. Was the patient's history clearly described and presented as a timeline? | X | | | |
| 3. Was the current clinical condition of the patient on presentation clearly described? | X | | | |
| 4. Were diagnostic tests or assessment methods and the results clearly described? | X | | | |
| 5. Was the intervention(s) or treatment procedure(s) clearly described? | X | | | |
| 6. Was the post-intervention clinical condition clearly described? | | X | | |
| 7. Were adverse events (harms) or unanticipated events identified and described? | | X | | |
| 8. Does the case report provide takeaway lessons? | | | X | |

Overall appraisal: Include ☐ Exclude ☒ Seek further info ☐

Comments (Including reason for exclusion)

Por los antecedentes médicos del paciente es complicado determinar si la sintomatología corresponde a una exacerbacion de la enfermedades previas (OCHOS y SFN) o bien cora parte del síndrome post-COVID.

© JBI, 2020. All rights reserved. JBI grants use of these tools for research purposes only. All other enquiries should be sent to.

### JBI Critical Appraisal Checklist for Case Series

Reviewer Marco Antonio Delaye Martínez Date 31/12/20

Author Brogna B., Bignardi E., Brogna C., et al. Year 2020 Record Number 24_32998840

| | Yes | No | Unclear | Not applicable |
|---|---|---|---|---|
| • Were there clear criteria for inclusion in the case series? | ☐ | ☐ | ☐ | ■ |
| • Was the condition measured in a standard, reliable way for all participants included in the case series? | ■ | ☐ | ☐ | ☐ |
| • Were valid methods used for identification of the condition for all participants included in the case series? | ■ | ☐ | ☐ | ☐ |
| • Did the case series have consecutive inclusion of participants? | ☐ | ■ | ☐ | ☐ |
| • Did the case series have complete inclusion of participants? | ■ | ☐ | ☐ | ☐ |
| • Was there clear reporting of the demographics of the participants in the study? | ☐ | ■ | ☐ | ☐ |
| • Was there clear reporting of clinical information of the participants? | ■ | ☐ | ☐ | ☐ |
| • Were the outcomes or follow up results of cases clearly reported? | ■ | ☐ | ☐ | ☐ |
| • Was there clear reporting of the presenting site(s)/clinic(s) demographic information? | ☐ | ■ | ☐ | ☐ |
| • Was statistical analysis appropriate? | ☐ | ☐ | ☐ | ■ |

Overall appraisal: Include ☐ Exclude ■ Seek further info ☐

Comments (Including reason for exclusion)

El estudio no tiene por objetivo reportar la presencia de síntomas persistentes en los pacientes, más bien resalta el papel de la TAC de tórax para el diagnóstico de COVID-19 en pacientes con sintomatología sugestiva de la enfermedad y pruebas RT-PCR negativas .

© JBI, 2020. All rights reserved. JBI grants use of these tools for research purposes only. All other enquiries should be sent to.

### JBI CRITICAL APPRAISAL CHECKLIST FOR COHORT STUDIES

Reviewer Marco Antonio Delaye Martínez Date 31/12/20

Author Landi F., Carfi A., Benvenuto F., et al. Year 2020 Record Number 25_33041095

| | Yes | No | Unclear | Not applicable |
|---|---|---|---|---|
| 1. Were the two groups similar and recruited from the same population? | ■ | □ | □ | □ |
| 2. Were the exposures measured similarly to assign people to both exposed and unexposed groups? | ■ | □ | □ | □ |
| 3. Was the exposure measured in a valid and reliable way? | ■ | □ | □ | □ |
| 4. Were confounding factors identified? | ■ | □ | □ | □ |
| 5. Were strategies to deal with confounding factors stated? | ■ | □ | □ | □ |
| 6. Were the groups/participants free of the outcome at the start of the study (or at the moment of exposure)? | ■ | □ | □ | □ |
| 7. Were the outcomes measured in a valid and reliable way? | ■ | □ | □ | □ |
| 8. Was the follow up time reported and sufficient to be long enough for outcomes to occur? | ■ | □ | □ | □ |
| 9. Was follow up complete, and if not, were the reasons to loss to follow up described and explored? | ■ | □ | □ | □ |
| 10. Were strategies to address incomplete follow up utilized? | □ | □ | □ | ■ |
| 11. Was appropriate statistical analysis used? | ■ | □ | □ | □ |

Overall appraisal: Include ■ Exclude □ Seek further info □

Comments (Including reason for exclusion)

No comments.

© JBI, 2020. All rights reserved. JBI grants use of these tools for research purposes only. All other enquiries should be sent to.

### JBI CRITICAL APPRAISAL CHECKLIST FOR CASE REPORTS

Reviewer Marco Antonio Delaye Martínez Date 31/12/20

Author Carroll E., Neumann H., Aguero Rosenfield M., et al. Year 2020 Record Number 26_32944946

| | Yes | No | Unclear | Not applicable |
|---|---|---|---|---|
| 1. Were patient's demographic characteristics clearly described? | ☐ | ■ | ☐ | ☐ |
| 2. Was the patient's history clearly described and presented as a timeline? | ■ | ☐ | ☐ | ☐ |
| 3. Was the current clinical condition of the patient on presentation clearly described? | ■ | ☐ | ☐ | ☐ |
| 4. Were diagnostic tests or assessment methods and the results clearly described? | ■ | ☐ | ☐ | ☐ |
| 5. Was the intervention(s) or treatment procedure(s) clearly described? | ■ | ☐ | ☐ | ☐ |
| 6. Was the post-intervention clinical condition clearly described? | ■ | ☐ | ☐ | ☐ |
| 7. Were adverse events (harms) or unanticipated events identified and described? | ■ | ☐ | ☐ | ☐ |
| 8. Does the case report provide takeaway lessons? | ■ | ☐ | ☐ | ☐ |

Overall appraisal: Include ☐ Exclude ■ Seek further info ☐

Comments (Including reason for exclusion)

Caso poco específico para long-COVID

© JBI, 2020. All rights reserved. JBI grants use of these tools for research purposes only. All other enquiries should be sent to.

#### JBI Critical Appraisal Checklist for Case Series

Reviewer Marco Antonio Delaye Martínez Date 31/12/20

Author Masset C., Ville S., Halary F., et al. Year 2020 Record Number 27_32939955

| | Yes | No | Unclear | Not applicable |
|---|---|---|---|---|
| Were there clear criteria for inclusion in the case series? | ☐ | ■ | ☐ | ☐ |
| Was the condition measured in a standard, reliable way for all participants included in the case series? | ■ | ☐ | ☐ | ☐ |
| Were valid methods used for identification of the condition for all participants included in the case series? | ■ | ☐ | ☐ | ☐ |
| Did the case series have consecutive inclusion of participants? | ☐ | ☐ | ■ | ☐ |
| Did the case series have complete inclusion of participants? | ■ | ☐ | ☐ | ☐ |
| Was there clear reporting of the demographics of the participants in the study? | ☐ | ■ | ☐ | ☐ |
| Was there clear reporting of clinical information of the participants? | ■ | ☐ | ☐ | ☐ |
| Were the outcomes or follow up results of cases clearly reported? | ■ | ☐ | ☐ | ☐ |
| Was there clear reporting of the presenting site(s)/clinic(s) demographic information? | ☐ | ■ | ☐ | ☐ |
| Was statistical analysis appropriate? | ☐ | ☐ | ☐ | ■ |

Overall appraisal: Include ☐ Exclude ■ Seek further info ☐

Comments (Including reason for exclusion)

Los síntomas reportados no cumplen con el criterio de temporalidad para síntomas persistentes.

© JBI, 2020. All rights reserved. JBI grants use of these tools for research purposes only. All other enquiries should be sent to.

### JBI CRITICAL APPRAISAL CHECKLIST FOR ANALYTICAL CROSS SECTIONAL STUDIES

Reviewer Marco Antonio Delaye Martínez Date 31/12/20

Author W. Vaes A., Machado F., Meys R., et al. Year 2020 Record Number 28_32932582

| | Yes | No | Unclear | Not applicable |
|---|---|---|---|---|
| 1. Were the criteria for inclusion in the sample clearly defined? | ☐ | ☐ | ☒ | ☐ |
| 2. Were the study subjects and the setting described in detail? | ☒ | ☐ | ☐ | ☐ |
| 3. Was the exposure measured in a valid and reliable way? | ☒ | ☐ | ☐ | ☐ |
| 4. Were objective, standard criteria used for measurement of the condition? | ☒ | ☐ | ☐ | ☐ |
| 5. Were confounding factors identified? | ☒ | ☐ | ☐ | ☐ |
| 6. Were strategies to deal with confounding factors stated? | ☐ | ☐ | ☒ | ☐ |
| 7. Were the outcomes measured in a valid and reliable way? | ☒ | ☐ | ☐ | ☐ |
| 8. Was appropriate statistical analysis used? | ☒ | ☐ | ☐ | ☐ |

Overall appraisal: Include ☐ Exclude ☒ Seek further info ☐

Comments (Including reason for exclusion)

La proporción de pacientes con diagnóstico confirmado de COVID por RT-PCR es tan sólo del 17%; el resto fue diagnosticado por síntomas o no tenía diagnóstico al momento del estudio. Sin embargo por la información que aporta el estudio es útil para la discusión.

© JBI, 2020. All rights reserved. JBI grants use of these tools for research purposes only. All other enquiries should be sent to.

### JBI CRITICAL APPRAISAL CHECKLIST FOR COHORT STUDIES

Reviewer Marco Antonio Delaye Martínez Date 04/01/21

Author Pizzini A., Aichner M., Sahanic S., et al. Year 2020 Record Number 29_32932831

| | Yes | No | Unclear | Not applicable |
|---|---|---|---|---|
| 1. Were the two groups similar and recruited from the same population? | ■ | □ | □ | □ |
| 2. Were the exposures measured similarly to assign people to both exposed and unexposed groups? | ■ | □ | □ | □ |
| 3. Was the exposure measured in a valid and reliable way? | ■ | □ | □ | □ |
| 4. Were confounding factors identified? | ■ | □ | □ | □ |
| 5. Were strategies to deal with confounding factors stated? | □ | □ | ■ | □ |
| 6. Were the groups/participants free of the outcome at the start of the study (or at the moment of exposure)? | □ | ■ | □ | □ |
| 7. Were the outcomes measured in a valid and reliable way? | ■ | □ | □ | □ |
| 8. Was the follow up time reported and sufficient to be long enough for outcomes to occur? | ■ | □ | □ | □ |
| 9. Was follow up complete, and if not, were the reasons to loss to follow up described and explored? | ■ | □ | □ | □ |
| 10. Were strategies to address incomplete follow up utilized? | □ | ■ | □ | □ |
| 11. Was appropriate statistical analysis used? | ■ | □ | □ | □ |

Overall appraisal: Include □ Exclude ■ Seek further info □

Comments (Including reason for exclusion)

No se reportan síntomas persistentes por COVID-19, sin embargo se descarta la asociación de deficiencia de vitamina D y el desarrollo de sí tomas persistentes. El artículo es útil para discusión.

© JBI, 2020. All rights reserved. JBI grants use of these tools for research purposes only. All other enquiries should be sent to.

### JBI CRITICAL APPRAISAL CHECKLIST FOR SYSTEMATIC REVIEWS AND RESEARCH SYNTHESES

Reviewer: Daniel de la Rosa  Date: 30-11-20

Author: Lawrence B. Afrina et al.  Year: 2020  Record Number: 30_32920235_DDM

| | | Yes | No | Unclear | Not applicable |
|---|---|---|---|---|---|
| 1. | Is the review question clearly and explicitly stated? | ☒ | ☐ | ☐ | ☐ |
| 2. | Were the inclusion criteria appropriate for the review question? | ☐ | ☐ | ☐ | ☒ |
| 3. | Was the search strategy appropriate? | ☐ | ☐ | ☐ | ☒ |
| 4. | Were the sources and resources used to search for studies adequate? | ☐ | ☐ | ☐ | ☒ |
| 5. | Were the criteria for appraising studies appropriate? | ☐ | ☐ | ☐ | ☒ |
| 6. | Was critical appraisal conducted by two or more reviewers independently? | ☐ | ☐ | ☐ | ☒ |
| 7. | Were there methods to minimize errors in data extraction? | ☐ | ☐ | ☐ | ☒ |
| 8. | Were the methods used to combine studies appropriate? | ☐ | ☐ | ☐ | ☒ |
| 9. | Was the likelihood of publication bias assessed? | ☐ | ☐ | ☐ | ☒ |
| 10. | Were recommendations for policy and/or practice supported by the reported data? | ☒ | ☐ | ☐ | ☐ |
| 11. | Were the specific directives for new research appropriate? | ☒ | ☐ | ☐ | ☐ |

Overall appraisal: Include ☐ Exclude ☒ Seek further info ☐

Comments (Including reason for exclusion)

### JBI CRITICAL APPRAISAL CHECKLIST FOR COHORT STUDIES

Reviewer Marco Antonio Delaye Martínez Date 04/01/21

Author Chen J., Xu X., Hu J., et al. Year 2020 Record Number 31_32909961

| | Yes | No | Unclear | Not applicable |
|---|---|---|---|---|
| 1. Were the two groups similar and recruited from the same population? | ■ | □ | □ | □ |
| 2. Were the exposures measured similarly to assign people to both exposed and unexposed groups? | ■ | □ | □ | □ |
| 3. Was the exposure measured in a valid and reliable way? | ■ | □ | □ | □ |
| 4. Were confounding factors identified? | ■ | □ | □ | □ |
| 5. Were strategies to deal with confounding factors stated? | ■ | □ | □ | □ |
| 6. Were the groups/participants free of the outcome at the start of the study (or at the moment of exposure)? | ■ | □ | □ | □ |
| 7. Were the outcomes measured in a valid and reliable way? | ■ | □ | □ | □ |
| 8. Was the follow up time reported and sufficient to be long enough for outcomes to occur? | ■ | □ | □ | □ |
| 9. Was follow up complete, and if not, were the reasons to loss to follow up described and explored? | ■ | □ | □ | □ |
| 10. Were strategies to address incomplete follow up utilized? | □ | ■ | □ | □ |
| 11. Was appropriate statistical analysis used? | ■ | □ | □ | □ |

Overall appraisal: Include □ Exclude ■ Seek further info □

Comments (Including reason for exclusion)

No se reporta la persistencia de síntomas por COVID-19.

© JBI, 2020. All rights reserved. JBI grants use of these tools for research purposes only. All other enquiries should be sent to.

### JBI CRITICAL APPRAISAL CHECKLIST FOR ANALYTICAL CROSS SECTIONAL STUDIES

Reviewer Daniel de la Rosa Date 23-11-20

Author Helen O'Brien Year 2020 Record Number 32_32894436

| | Yes | No | Unclear | Not applicable |
|---|---|---|---|---|
| 1. Were the criteria for inclusion in the sample clearly defined? | ☐ | ☒ | ☐ | ☐ |
| 2. Were the study subjects and the setting described in detail? | ☒ | ☐ | ☐ | ☐ |
| 3. Was the exposure measured in a valid and reliable way? | ☐ | ☐ | ☐ | ☒ |
| 4. Were objective, standard criteria used for measurement of the condition? | ☐ | ☐ | ☐ | ☒ |
| 5. Were confounding factors identified? | ☐ | ☐ | ☐ | ☒ |
| 6. Were strategies to deal with confounding factors stated? | ☐ | ☐ | ☐ | ☒ |
| 7. Were the outcomes measured in a valid and reliable way? | ☐ | ☐ | ☐ | ☒ |
| 8. Was appropriate statistical analysis used? | ☐ | ☐ | ☐ | ☒ |

Overall appraisal: Include ☐ Exclude ☒ Seek further info ☐

Comments (Including reason for exclusion)

"El tiempo de seguimiento no se especifica para todos los pacienes, ademas el metodo diagnostico no fue PCR para todos."

### JBI CRITICAL APPRAISAL CHECKLIST FOR TEXT AND OPINION PAPERS

Reviewer Marco Antonio Delaye Martínez Date 04/01/21

Author Davido B., Seang S., Barizien N., et al. Year 2020 Record Number 33_32898712

| | Yes | No | Unclear | Not applicable |
|---|---|---|---|---|
| 1. Is the source of the opinion clearly identified? | ■ | □ | □ | □ |
| 2. Does the source of opinion have standing in the field of expertise? | ■ | □ | □ | □ |
| 3. Are the interests of the relevant population the central focus of the opinion? | ■ | □ | □ | □ |
| 4. Is the stated position the result of an analytical process, and is there logic in the opinion expressed? | □ | ■ | □ | □ |
| 5. Is there reference to the extant literature? | ■ | □ | □ | □ |
| 6. Is any incongruence with the literature/sources logically defended? | ■ | □ | □ | □ |

Overall appraisal: Include □ Exclude ■ Seek further info □

Comments (Including reason for exclusion)

El artículo no es útil para los propósitos de la presente revisión

© JBI, 2020. All rights reserved. JBI grants use of these tools for research purposes only. All other enquiries should be sent to.

### JBI CRITICAL APPRAISAL CHECKLIST FOR CASE REPORTS

Reviewer Marco Antonio Delaye Martínez Date 31/12/20

Author Wang M., Li T., Qiao F., et al. Year 2020 Record Number 34_32898993

| | Yes | No | Unclear | Not applicable |
|---|---|---|---|---|
| 1. Were patient's demographic characteristics clearly described? | ☐ | ■ | ☐ | ☐ |
| 2. Was the patient's history clearly described and presented as a timeline? | ■ | ☐ | ☐ | ☐ |
| 3. Was the current clinical condition of the patient on presentation clearly described? | ■ | ☐ | ☐ | ☐ |
| 4. Were diagnostic tests or assessment methods and the results clearly described? | ■ | ☐ | ☐ | ☐ |
| 5. Was the intervention(s) or treatment procedure(s) clearly described? | ■ | ☐ | ☐ | ☐ |
| 6. Was the post-intervention clinical condition clearly described? | ■ | ☐ | ☐ | ☐ |
| 7. Were adverse events (harms) or unanticipated events identified and described? | ☐ | ■ | ☐ | ☐ |
| 8. Does the case report provide takeaway lessons? | ■ | ☐ | ☐ | ☐ |

Overall appraisal: Include ☐ Exclude ■ Seek further info ☐

Comments (Including reason for exclusion)

Dado que es un reporte de caso, el artículo se excluye de la revisión sistemática. Sin embargo aporta información relevante que puede ser utilizada en la discusión.

© JBI, 2020. All rights reserved. JBI grants use of these tools for research purposes only. All other enquiries should be sent to.

### JBI CRITICAL APPRAISAL CHECKLIST FOR TEXT AND OPINION PAPERS

Reviewer Marco Antonio Delaye Martínez Date 04/01/21

Author Miglis M., Goodman B., Chémali K., et al. Year 2020 Record Number 35_32891765

| | Yes | No | Unclear | Not applicable |
|---|---|---|---|---|
| 1. Is the source of the opinion clearly identified? | ☒ | ☐ | ☐ | ☐ |
| 2. Does the source of opinion have standing in the field of expertise? | ☒ | ☐ | ☐ | ☐ |
| 3. Are the interests of the relevant population the central focus of the opinion? | ☒ | ☐ | ☐ | ☐ |
| 4. Is the stated position the result of an analytical process, and is there logic in the opinion expressed? | ☒ | ☐ | ☐ | ☐ |
| 5. Is there reference to the extant literature? | ☒ | ☐ | ☐ | ☐ |
| 6. Is any incongruence with the literature/sources logically defended? | ☐ | ☒ | ☐ | ☐ |

Overall appraisal: Include ☐ Exclude ☒ Seek further info ☐

Comments (Including reason for exclusion)

El artículo carece de información relevante para incluirlo en la discusión . Además no cumple con el criterio de inclusión en cuanto al tipo de artículos aceptados para la revisión.

© JBI, 2020. All rights reserved. JBI grants use of these tools for research purposes only. All other enquiries should be sent to.

### JBI CRITICAL APPRAISAL CHECKLIST FOR SYSTEMATIC REVIEWS AND RESEARCH SYNTHESES

Reviewer Daniel de la Rosa Date 24-12-20

Author Mathew G Wilson, et al Year 2020 Record Number 21_32737507

| | | Yes | No | Unclear | Not applicable |
|---|---|---|---|---|---|
| 1. | Is the review question clearly and explicitly stated? | ☒ | ☐ | ☐ | ☐ |
| 2. | Were the inclusion criteria appropriate for the review question? | ☐ | ☐ | ☐ | ☒ |
| 3. | Was the search strategy appropriate? | ☐ | ☐ | ☐ | ☒ |
| 4. | Were the sources and resources used to search for studies adequate? | ☐ | ☐ | ☐ | ☒ |
| 5. | Were the criteria for appraising studies appropriate? | ☐ | ☐ | ☐ | ☒ |
| 6. | Was critical appraisal conducted by two or more reviewers independently? | ☐ | ☐ | ☐ | ☒ |
| 7. | Were there methods to minimize errors in data extraction? | ☐ | ☐ | ☐ | ☒ |
| 8. | Were the methods used to combine studies appropriate? | ☐ | ☐ | ☐ | ☒ |
| 9. | Was the likelihood of publication bias assessed? | ☐ | ☐ | ☐ | ☒ |
| 10. | Were recommendations for policy and/or practice supported by the reported data? | ☒ | ☐ | ☐ | ☐ |
| 11. | Were the specific directives for new research appropriate? | ☒ | ☐ | ☐ | ☐ |

Overall appraisal: Include ☐ Exclude ☒ Seek further info ☐

Comments (Including reason for exclusion)

______________________________________________

______________________________________________

______________________________________________

### JBI CRITICAL APPRAISAL CHECKLIST FOR COHORT STUDIES

Reviewer Marco Antonio Delaye Martínez Date 04/01/21

Author Krisztian S., Palmer J., Chen D., et al. Year 2020 Record Number 37_33091285

| | Yes | No | Unclear | Not applicable |
|---|---|---|---|---|
| 1. Were the two groups similar and recruited from the same population? | ■ | ☐ | ☐ | ☐ |
| 2. Were the exposures measured similarly to assign people to both exposed and unexposed groups? | ☐ | ☐ | ☐ | ■ |
| 3. Was the exposure measured in a valid and reliable way? | ☐ | ☐ | ☐ | ■ |
| 4. Were confounding factors identified? | ☐ | ☐ | ■ | ☐ |
| 5. Were strategies to deal with confounding factors stated? | ☐ | ☐ | ■ | ☐ |
| 6. Were the groups/participants free of the outcome at the start of the study (or at the moment of exposure)? | ■ | ☐ | ☐ | ☐ |
| 7. Were the outcomes measured in a valid and reliable way? | ■ | ☐ | ☐ | ☐ |
| 8. Was the follow up time reported and sufficient to be long enough for outcomes to occur? | ☐ | ■ | ☐ | ☐ |
| 9. Was follow up complete, and if not, were the reasons to loss to follow up described and explored? | ■ | ☐ | ☐ | ☐ |
| 10. Were strategies to address incomplete follow up utilized? | ☐ | ■ | ☐ | ☐ |
| 11. Was appropriate statistical analysis used? | ■ | ☐ | ☐ | ☐ |

Overall appraisal: Include ☐ Exclude ■ Seek further info ☐

Comments (Including reason for exclusion)

Los sintomas persistentes que reportan no cumplen con los criterios de temporalidad. Además, no sé reporta la proporción de cada síntoma.

© JBI, 2020. All rights reserved. JBI grants use of these tools for research purposes only. All other enquiries should be sent to.

### JBI CRITICAL APPRAISAL CHECKLIST FOR CASE REPORTS

Reviewer Marco Antonio Delaye Martínez Date 04/01/21

Author Iancu G., Salomon A., Birlutiu V. Year 2020 Record Number 38_32871902

| | Yes | No | Unclear | Not applicable |
|---|---|---|---|---|
| 1. Were patient's demographic characteristics clearly described? | ☐ | ■ | ☐ | ☐ |
| 2. Was the patient's history clearly described and presented as a timeline? | ■ | ☐ | ☐ | ☐ |
| 3. Was the current clinical condition of the patient on presentation clearly described? | ■ | ☐ | ☐ | ☐ |
| 4. Were diagnostic tests or assessment methods and the results clearly described? | ■ | ☐ | ☐ | ☐ |
| 5. Was the intervention(s) or treatment procedure(s) clearly described? | ■ | ☐ | ☐ | ☐ |
| 6. Was the post-intervention clinical condition clearly described? | ■ | ☐ | ☐ | ☐ |
| 7. Were adverse events (harms) or unanticipated events identified and described? | ☐ | ☐ | ☐ | ■ |
| 8. Does the case report provide takeaway lessons? | ■ | ☐ | ☐ | ☐ |

Overall appraisal: Include ☐ Exclude ■ Seek further info ☐

Comments (Including reason for exclusion)

Por el diseño del estudio no puede ser incluido en la revisión sistemática, sin embargo aporta información útil para ser incluido en la discusión.

© JBI, 2020. All rights reserved. JBI grants use of these tools for research purposes only. All other enquiries should be sent to.

### JBI CRITICAL APPRAISAL CHECKLIST FOR TEXT AND OPINION PAPERS

Reviewer Marco Antonio Delaye Martínez Date 04/01/21

Author Garg P., Arora U., Kumar A., et al. Year 2020 Record Number 39_32852801

| | Yes | No | Unclear | Not applicable |
|---|---|---|---|---|
| 1. Is the source of the opinion clearly identified? | ■ | □ | □ | □ |
| 2. Does the source of opinion have standing in the field of expertise? | □ | ■ | □ | □ |
| 3. Are the interests of the relevant population the central focus of the opinion? | ■ | □ | □ | □ |
| 4. Is the stated position the result of an analytical process, and is there logic in the opinion expressed? | ■ | □ | □ | □ |
| 5. Is there reference to the extant literature? | ■ | □ | □ | □ |
| 6. Is any incongruence with the literature/sources logically defended? | □ | ■ | □ | □ |

Overall appraisal: Include □ Exclude ■ Seek further info □

Comments (Including reason for exclusion)

Los síntomas persistentes que reportan corresponden a otros estudios incluidos contemplados para la revisión en la misma base de datos, por lo que el artículo queda rechazado.

© JBI, 2020. All rights reserved. JBI grants use of these tools for research purposes only. All other enquiries should be sent to.

### JBI Critical Appraisal Checklist for Case Series

Reviewer Marco Antonio Delaye Martínez Date 04/01/21

Author Roth C., Berat N., Schnider P., et al. Year 2020 Record Number 40_32852173

| | Yes | No | Unclear | Not applicable |
|---|---|---|---|---|
| 1. Were there clear criteria for inclusion in the case series? | ☐ | ■ | ☐ | ☐ |
| 2. Was the condition measured in a standard, reliable way for all participants included in the case series? | ■ | ☐ | ☐ | ☐ |
| 3. Were valid methods used for identification of the condition for all participants included in the case series? | ■ | ☐ | ☐ | ☐ |
| 4. Did the case series have consecutive inclusion of participants? | ■ | ☐ | ☐ | ☐ |
| 5. Did the case series have complete inclusion of participants? | ■ | ☐ | ☐ | ☐ |
| 6. Was there clear reporting of the demographics of the participants in the study? | ☐ | ■ | ☐ | ☐ |
| 7. Was there clear reporting of clinical information of the participants? | ■ | ☐ | ☐ | ☐ |
| 8. Were the outcomes or follow up results of cases clearly reported? | ■ | ☐ | ☐ | ☐ |
| 9. Was there clear reporting of the presenting site(s)/clinic(s) demographic information? | ☐ | ■ | ☐ | ☐ |
| 10. Was statistical analysis appropriate? | ☐ | ■ | ☐ | ☐ |

Overall appraisal: Include ☐ Exclude ■ Seek further info ☐

Comments (Including reason for exclusion)

Por las caracteristicasbdel cuadro clínico de los pacientes, el artículo ofrece información relevante para la discusión. Sin embargo, se excluye de l revisión sistemática por el diseño del estudio.

© JBI, 2020. All rights reserved. JBI grants use of these tools for research purposes only. All other enquiries should be sent to.

### JBI CRITICAL APPRAISAL CHECKLIST FOR SYSTEMATIC REVIEWS AND RESEARCH SYNTHESES

Reviewer: Daniel de la Rosa    Date: 21-12-20

Author: Piero Portincasa, et al    Year: 2020    Record Number: 41_32589264

| | | Yes | No | Unclear | Not applicable |
|---|---|---|---|---|---|
| 1. | Is the review question clearly and explicitly stated? | ☒ | ☐ | ☐ | ☐ |
| 2. | Were the inclusion criteria appropriate for the review question? | ☐ | ☐ | ☒ | ☐ |
| 3. | Was the search strategy appropriate? | ☐ | ☐ | ☐ | ☒ |
| 4. | Were the sources and resources used to search for studies adequate? | ☐ | ☐ | ☒ | ☐ |
| 5. | Were the criteria for appraising studies appropriate? | ☐ | ☐ | ☐ | ☒ |
| 6. | Was critical appraisal conducted by two or more reviewers independently? | ☐ | ☐ | ☐ | ☒ |
| 7. | Were there methods to minimize errors in data extraction? | ☐ | ☐ | ☐ | ☒ |
| 8. | Were the methods used to combine studies appropriate? | ☐ | ☐ | ☐ | ☒ |
| 9. | Was the likelihood of publication bias assessed? | ☐ | ☐ | ☐ | ☒ |
| 10. | Were recommendations for policy and/or practice supported by the reported data? | ☒ | ☐ | ☐ | ☐ |
| 11. | Were the specific directives for new research appropriate? | ☒ | ☐ | ☐ | ☐ |

Overall appraisal: Include ☐ Exclude ☒ Seek further info ☐

Comments (Including reason for exclusion)

### JBI CRITICAL APPRAISAL CHECKLIST FOR SYSTEMATIC REVIEWS AND RESEARCH SYNTHESES

Reviewer Daniel de la Rosa Date 21-12-20

Author Chanu Rhee, et al. Year 2020 Record Number 42_33029620

| | Yes | No | Unclear | Not applicable |
|---|---|---|---|---|
| 1. Is the review question clearly and explicitly stated? | ☒ | ☐ | ☐ | ☐ |
| 2. Were the inclusion criteria appropriate for the review question? | ☐ | ☐ | ☐ | ☒ |
| 3. Was the search strategy appropriate? | ☐ | ☐ | ☐ | ☒ |
| 4. Were the sources and resources used to search for studies adequate? | ☐ | ☐ | ☐ | ☒ |
| 5. Were the criteria for appraising studies appropriate? | ☐ | ☐ | ☐ | ☒ |
| 6. Was critical appraisal conducted by two or more reviewers independently? | ☐ | ☐ | ☐ | ☒ |
| 7. Were there methods to minimize errors in data extraction? | ☐ | ☐ | ☐ | ☒ |
| 8. Were the methods used to combine studies appropriate? | ☐ | ☐ | ☐ | ☒ |
| 9. Was the likelihood of publication bias assessed? | ☐ | ☐ | ☐ | ☒ |
| 10. Were recommendations for policy and/or practice supported by the reported data? | ☒ | ☐ | ☐ | ☐ |
| 11. Were the specific directives for new research appropriate? | ☒ | ☐ | ☐ | ☐ |

Overall appraisal: Include ☐ Exclude ☒ Seek further info ☐

Comments (Including reason for exclusion)

### JBI CRITICAL APPRAISAL CHECKLIST FOR TEXT AND OPINION PAPERS

Reviewer Marco Antonio Delaye Martínez Date 05/01/21

Author Sebio R. Year 2020 Record Number 43_32844317

| | Yes | No | Unclear | Not applicable |
|---|---|---|---|---|
| 1. Is the source of the opinion clearly identified? | ■ | □ | □ | □ |
| 2. Does the source of opinion have standing in the field of expertise? | □ | ■ | □ | □ |
| 3. Are the interests of the relevant population the central focus of the opinion? | ■ | □ | □ | □ |
| 4. Is the stated position the result of an analytical process, and is there logic in the opinion expressed? | ■ | □ | □ | □ |
| 5. Is there reference to the extant literature? | ■ | □ | □ | □ |
| 6. Is any incongruence with the literature/sources logically defended? | □ | ■ | □ | □ |

Overall appraisal: Include □ Exclude ■ Seek further info □

Comments (Including reason for exclusion)

El articulo no proporciona información útil más allá de la ya consultada para los propósitos de la presente revisión sistemática.

© JBI, 2020. All rights reserved. JBI grants use of these tools for research purposes only. All other enquiries should be sent to.

### JBI CRITICAL APPRAISAL CHECKLIST FOR ANALYTICAL CROSS SECTIONAL STUDIES

Reviewer Marco Antonio Delaye Martínez Date 05/01/21

Author Garrigues E., Janvier P., Kherabi Y., et al. Year 2020 Record Number 44_32853602

| | Yes | No | Unclear | Not applicable |
|---|---|---|---|---|
| 1. Were the criteria for inclusion in the sample clearly defined? | ☐ | ■ | ☐ | ☐ |
| 2. Were the study subjects and the setting described in detail? | ☐ | ■ | ☐ | ☐ |
| 3. Was the exposure measured in a valid and reliable way? | ■ | ☐ | ☐ | ☐ |
| 4. Were objective, standard criteria used for measurement of the condition? | ■ | ☐ | ☐ | ☐ |
| 5. Were confounding factors identified? | ☐ | ■ | ☐ | ☐ |
| 6. Were strategies to deal with confounding factors stated? | ☐ | ■ | ☐ | ☐ |
| 7. Were the outcomes measured in a valid and reliable way? | ■ | ☐ | ☐ | ☐ |
| 8. Was appropriate statistical analysis used? | ■ | ☐ | ☐ | ☐ |

Overall appraisal: Include ☐ Exclude ■ Seek further info ☐

Comments (Including reason for exclusion)

Las datos que aquí reportan pertenecen a un estudio transversal contenido dentro de una carta al editor. Por la información descrita, el artículo es útil para los propósitos de la revisión sistemática. Sin embargo, Su aceptación al estudio está a reserva de discusión con los otros revisores.

© JBI, 2020. All rights reserved. JBI grants use of these tools for research purposes only. All other enquiries should be sent to.

### JBI CRITICAL APPRAISAL CHECKLIST FOR ANALYTICAL CROSS SECTIONAL STUDIES

Reviewer Marco Antonio Delaye Martínez Date 05/01/21

Author Tomasoni D., Bai F., Castoldi R., et al. Year 2020 Record Number 45_32841387

| | Yes | No | Unclear | Not applicable |
|---|---|---|---|---|
| 1. Were the criteria for inclusion in the sample clearly defined? | ☐ | ☐ | ☐ | ☐ |
| 2. Were the study subjects and the setting described in detail? | ■ | ☐ | ☐ | ☐ |
| 3. Was the exposure measured in a valid and reliable way? | ■ | ☐ | ☐ | ☐ |
| 4. Were objective, standard criteria used for measurement of the condition? | ■ | ☐ | ☐ | ☐ |
| 5. Were confounding factors identified? | ☐ | ☐ | ■ | ☐ |
| 6. Were strategies to deal with confounding factors stated? | ☐ | ☐ | ■ | ☐ |
| 7. Were the outcomes measured in a valid and reliable way? | ■ | ☐ | ☐ | ☐ |
| 8. Was appropriate statistical analysis used? | ■ | ☐ | ☐ | ☐ |

Overall appraisal: Include ■ Exclude ☐ Seek further info ☐

Comments (Including reason for exclusion)

No comments.

© JBI, 2020. All rights reserved. JBI grants use of these tools for research purposes only. All other enquiries should be sent to.

### JBI CRITICAL APPRAISAL CHECKLIST FOR CASE CONTROL STUDIES

Reviewer: Daniel de la Rosa  Date: 06-01-20

Author: Deborah H. L. Ng, et al.  Year: 2020  Record Number: 46_32999893

| | Yes | No | Unclear | Not applicable |
|---|---|---|---|---|
| 1. Were the groups comparable other than the presence of disease in cases or the absence of disease in controls? | ☑ | ☐ | ☐ | ☐ |
| 2. Were cases and controls matched appropriately? | ☑ | ☐ | ☐ | ☐ |
| 3. Were the same criteria used for identification of cases and controls? | ☑ | ☐ | ☐ | ☐ |
| 4. Was exposure measured in a standard, valid and reliable way? | ☑ | ☐ | ☐ | ☐ |
| 5. Was exposure measured in the same way for cases and controls? | ☑ | ☐ | ☐ | ☐ |
| 6. Were confounding factors identified? | ☑ | ☐ | ☐ | ☐ |
| 7. Were strategies to deal with confounding factors stated? | ☑ | ☐ | ☐ | ☐ |
| 8. Were outcomes assessed in a standard, valid and reliable way for cases and controls? | ☑ | ☐ | ☐ | ☐ |
| 9. Was the exposure period of interest long enough to be meaningful? | ☐ | ☐ | ☐ | ☑ |
| 10. Was appropriate statistical analysis used? | ☑ | ☐ | ☐ | ☐ |

Overall appraisal: Include ☐ Exclude ☑ Seek further info ☐

Comments (Including reason for exclusion)

They don't report relevant information regarding persistent symptoms

### JBI CRITICAL APPRAISAL CHECKLIST FOR TEXT AND OPINION PAPERS

Reviewer Daniel de la Rosa Date 14/11/20

Author Arsun Bektas et al Year 2020 Record Number 47_32849908

| | Yes | No | Unclear | Not applicable |
|---|---|---|---|---|
| 1. Is the source of the opinion clearly identified? | ☒ | ☐ | ☐ | ☐ |
| 2. Does the source of opinion have standing in the field of expertise? | ☒ | ☐ | ☐ | ☐ |
| 3. Are the interests of the relevant population the central focus of the opinion? | ☒ | ☐ | ☐ | ☐ |
| 4. Is the stated position the result of an analytical process, and is there logic in the opinion expressed? | ☒ | ☐ | ☐ | ☐ |
| 5. Is there reference to the extant literature? | ☒ | ☐ | ☐ | ☐ |
| 6. Is any incongruence with the literature/sources logically defended? | ☒ | ☐ | ☐ | ☐ |

Overall appraisal: Include ☐ Exclude ☒ Seek further info ☐

Comments (Including reason for exclusion)

Review

© JBI, 2020. All rights reserved. JBI grants use of these tools for research purposes only. All other enquiries should be sent to.

### JBI CRITICAL APPRAISAL CHECKLIST FOR COHORT STUDIES

Reviewer Daniel de la Rosa Date 05-01-21

Author Sachie Ikegami et al. Year ________ Record Number 48_32840002

| | Yes | No | Unclear | Not applicable |
|---|---|---|---|---|
| 1. Were the two groups similar and recruited from the same population? | ☒ | ☐ | ☐ | ☐ |
| 2. Were the exposures measured similarly to assign people to both exposed and unexposed groups? | ☒ | ☐ | ☐ | ☐ |
| 3. Was the exposure measured in a valid and reliable way? | ☐ | ☐ | ☒ | ☐ |
| 4. Were confounding factors identified? | ☒ | ☐ | ☐ | ☐ |
| 5. Were strategies to deal with confounding factors stated? | ☒ | ☐ | ☐ | ☐ |
| 6. Were the groups/participants free of the outcome at the start of the study (or at the moment of exposure)? | ☐ | ☐ | ☒ | ☐ |
| 7. Were the outcomes measured in a valid and reliable way? | ☒ | ☐ | ☐ | ☐ |
| 8. Was the follow up time reported and sufficient to be long enough for outcomes to occur? | ☐ | ☒ | ☐ | ☐ |
| 9. Was follow up complete, and if not, were the reasons to loss to follow up described and explored? | ☐ | ☐ | ☒ | ☐ |
| 10. Were strategies to address incomplete follow up utilized? | ☐ | ☒ | ☐ | ☐ |
| 11. Was appropriate statistical analysis used? | ☒ | ☐ | ☐ | ☐ |

Overall appraisal: Include ☐ Exclude ☒ Seek further info ☐

Comments (Including reason for exclusion)

No comments.

© JBI, 2020. All rights reserved. JBI grants use of these tools for research purposes only. All other enquiries should be sent to.

### JBI CRITICAL APPRAISAL CHECKLIST FOR CASE REPORTS

Reviewer Marco Antonio Delaye Martínez Date 05/01/21

Author Wang J., Hang X., Wei B., et al. Year 2020 Record Number 49_32846839

| | Yes | No | Unclear | Not applicable |
|---|---|---|---|---|
| 1. Were patient's demographic characteristics clearly described? | ☐ | ● | ☐ | ☐ |
| 2. Was the patient's history clearly described and presented as a timeline? | ● | ☐ | ☐ | ☐ |
| 3. Was the current clinical condition of the patient on presentation clearly described? | ● | ☐ | ☐ | ☐ |
| 4. Were diagnostic tests or assessment methods and the results clearly described? | ● | ☐ | ☐ | ☐ |
| 5. Was the intervention(s) or treatment procedure(s) clearly described? | ● | ☐ | ☐ | ☐ |
| 6. Was the post-intervention clinical condition clearly described? | ● | ☐ | ☐ | ☐ |
| 7. Were adverse events (harms) or unanticipated events identified and described? | ☐ | ● | ☐ | ☐ |
| 8. Does the case report provide takeaway lessons? | ● | ☐ | ☐ | ☐ |

Overall appraisal: Include ☐ Exclude ● Seek further info ☐

Comments (Including reason for exclusion)

Debido que no sé reporta la persistencia de síntomas por COVID-19 a profundidad, el artículo queda excluido de la revisión sistemática. Así mismo, no cumple con el criterio de diseño para su inclusión.

© JBI, 2020. All rights reserved. JBI grants use of these tools for research purposes only. All other enquiries should be sent to.

### JBI CRITICAL APPRAISAL CHECKLIST FOR ANALYTICAL CROSS SECTIONAL STUDIES

Reviewer Daniel de la Rosa Date 10-01-21

Author Peng Xia et al Year 2020 Record Number 50_32826326

| | Yes | No | Unclear | Not applicable |
|---|---|---|---|---|
| 1. Were the criteria for inclusion in the sample clearly defined? | ☒ | ☐ | ☐ | ☐ |
| 2. Were the study subjects and the setting described in detail? | ☒ | ☐ | ☐ | ☐ |
| 3. Was the exposure measured in a valid and reliable way? | ☒ | ☐ | ☐ | ☐ |
| 4. Were objective, standard criteria used for measurement of the condition? | ☐ | ☐ | ☒ | ☐ |
| 5. Were confounding factors identified? | ☐ | ☐ | ☒ | ☐ |
| 6. Were strategies to deal with confounding factors stated? | ☐ | ☐ | ☒ | ☐ |
| 7. Were the outcomes measured in a valid and reliable way? | ☐ | ☐ | ☒ | ☐ |
| 8. Was appropriate statistical analysis used? | ☒ | ☐ | ☐ | ☐ |

Overall appraisal: Include ☐ Exclude ☒ Seek further info ☐

Comments (Including reason for exclusion)

No information on long term manifestation of COVID-19

### JBI CRITICAL APPRAISAL CHECKLIST FOR COHORT STUDIES

Reviewer: Daniel de la Rosa Date: 05-01-21

Author: Maria A. Corcorran, et al. Year: 2020 Record Number: 51_32827597

| | Yes | No | Unclear | Not applicable |
|---|---|---|---|---|
| 1. Were the two groups similar and recruited from the same population? | ☒ | ☐ | ☐ | ☐ |
| 2. Were the exposures measured similarly to assign people to both exposed and unexposed groups? | ☒ | ☐ | ☐ | ☐ |
| 3. Was the exposure measured in a valid and reliable way? | ☒ | ☐ | ☐ | ☐ |
| 4. Were confounding factors identified? | ☐ | ☒ | ☐ | ☐ |
| 5. Were strategies to deal with confounding factors stated? | ☐ | ☒ | ☐ | ☐ |
| 6. Were the groups/participants free of the outcome at the start of the study (or at the moment of exposure)? | ☒ | ☐ | ☐ | ☐ |
| 7. Were the outcomes measured in a valid and reliable way? | ☒ | ☐ | ☐ | ☐ |
| 8. Was the follow up time reported and sufficient to be long enough for outcomes to occur? | ☐ | ☒ | ☐ | ☐ |
| 9. Was follow up complete, and if not, were the reasons to loss to follow up described and explored? | ☐ | ☐ | ☒ | ☐ |
| 10. Were strategies to address incomplete follow up utilized? | ☐ | ☒ | ☐ | ☐ |
| 11. Was appropriate statistical analysis used? | ☒ | ☐ | ☐ | ☐ |

Overall appraisal: Include ☐ Exclude ☒ Seek further info ☐

Comments (Including reason for exclusion)

No comments.

© JBI, 2020. All rights reserved. JBI grants use of these tools for research purposes only. All other enquiries should be sent to.

### CHECKLIST FOR ANALYTICAL CROSS SECTIONAL STUDIES

Critical Appraisal tools for use in JBI Systematic Reviews

### INTRODUCTION

JBI is an JBI is an international research organisation based in the Faculty of Health and Medical Sciences at the University of Adelaide, South Australia. JBI develops and delivers unique evidence-based information, software, education and training designed to improve healthcare practice and health outcomes. With over 70 Collaborating Entities, servicing over 90 countries, JBI is a recognised global leader in evidence-based healthcare.

#### JBI Systematic Reviews

The core of evidence synthesis is the systematic review of literature of a particular intervention, condition or issue. The systematic review is essentially an analysis of the available literature (that is, evidence) and a judgment of the effectiveness or otherwise of a practice, involving a series of complex steps. JBI takes a particular view on what counts as evidence and the methods utilised to synthesise those different types of evidence. In line with this broader view of evidence, JBI has developed theories, methodologies and rigorous processes for the critical appraisal and synthesis of these diverse forms of evidence in order to aid in clinical decision-making in healthcare. There now exists JBI guidance for conducting reviews of effectiveness research, qualitative research, prevalence/incidence, etiology/risk, economic evaluations, text/opinion, diagnostic test accuracy, mixed-methods, umbrella reviews and scoping reviews. Further information regarding JBI systematic reviews can be found in the JBI Evidence Synthesis Manual.

#### JBI Critical Appraisal Tools

All systematic reviews incorporate a process of critique or appraisal of the research evidence. The purpose of this appraisal is to assess the methodological quality of a study and to determine the extent to which a study has addressed the possibility of bias in its design, conduct and analysis. All papers selected for inclusion in the systematic review (that is – those that meet the inclusion criteria described in the protocol) need to be subjected to rigorous appraisal by two critical appraisers. The results of this appraisal can then be used to inform synthesis and interpretation of the results of the study. JBI Critical appraisal tools have been developed by the JBI and collaborators and approved by the JBI Scientific Committee following extensive peer review. Although designed for use in systematic reviews, JBI critical appraisal tools can also be used when creating Critically Appraised Topics (CAT), in journal clubs and as an educational tool.

© JBI, 2020. All rights reserved. JBI grants use of these tools for research purposes only. All other enquiries should be sent to.

### JBI CRITICAL APPRAISAL CHECKLIST FOR ANALYTICAL CROSS SECTIONAL STUDIES

**Reviewer: Daniel de la Rosa Martinez** **Date: 27 de abril 2020**

Author: Year: 2020 Record Number: 52_32839787_DDM

| | Yes | No | Unclear | Not applicable |
|---|---|---|---|---|
| 1. Were the criteria for inclusion in the sample clearly defined? | ■ | ☐ | ☐ | ☐ |
| 2. Were the study subjects and the setting described in detail? | ■ | ☐ | ☐ | ☐ |
| 3. Was the exposure measured in a valid and reliable way? | ☐ | ☐ | ■ | ☐ |
| 4. Were objective, standard criteria used for measurement of the condition? | ☐ | ☐ | ■ | ☐ |
| 5. Were confounding factors identified? | ■ | ☐ | ☐ | ☐ |
| 6. Were strategies to deal with confounding factors stated? | ■ | ☐ | ☐ | ☐ |
| 7. Were the outcomes measured in a valid and reliable way? | ■ | ☐ | ☐ | ☐ |
| 8. Was appropriate statistical analysis used? | ■ | ☐ | ☐ | ☐ |

Overall appraisal: Include ☐ Exclude ■ Seek further info ☐

Comments (Including reason for exclusion)

_No se mencionan sintomas persistentes, se hace una exploracion de la clinica pero sin el seguimiento suficiente para explorar secuelas o long covid

© JBI, 2020. All rights reserved. JBI grants use of these tools for research purposes only. All other enquiries should be sent to.

### EXPLANATION OF ANALYTICAL CROSS SECTIONAL STUDIES CRITICAL APPRAISAL

*How to cite:* Moola S, Munn Z, Tufanaru C, Aromataris E, Sears K, Sfetcu R, Currie M, Qureshi R, Mattis P, Lisy K, Mu P-F. Chapter 7: Systematic reviews of etiology and risk . In: Aromataris E, Munn Z (Editors). *JBI Manual for Evidence Synthesis*. JBI, 2020. Available from https://synthesismanual.jbi.global

**Analytical cross sectional studies Critical Appraisal Tool**

Answers: Yes, No, Unclear or Not/Applicable

#### 1. Were the criteria for inclusion in the sample clearly defined?

The authors should provide clear inclusion and exclusion criteria that they developed prior to recruitment of the study participants. The inclusion/exclusion criteria should be specified (e.g., risk, stage of disease progression) with sufficient detail and all the necessary information critical to the study.

#### 2. Were the study subjects and the setting described in detail?

The study sample should be described in sufficient detail so that other researchers can determine if it is comparable to the population of interest to them. The authors should provide a clear description of the population from which the study participants were selected or recruited, including demographics, location, and time period.

#### 3. Was the exposure measured in a valid and reliable way?

The study should clearly describe the method of measurement of exposure. Assessing validity requires that a 'gold standard' is available to which the measure can be compared. The validity of exposure measurement usually relates to whether a current measure is appropriate or whether a measure of past exposure is needed.

Reliability refers to the processes included in an epidemiological study to check repeatability of measurements of the exposures. These usually include intra-observer reliability and inter-observer reliability.

#### 4. Were objective, standard criteria used for measurement of the condition?

It is useful to determine if patients were included in the study based on either a specified diagnosis or definition. This is more likely to decrease the risk of bias. Characteristics are another useful approach to matching groups, and studies that did not use specified diagnostic methods or definitions should provide evidence on matching by key characteristics

#### 5. Were confounding factors identified?

Confounding has occurred where the estimated intervention exposure effect is biased by the presence of some difference between the comparison groups (apart from the exposure investigated/of interest). Typical confounders include baseline characteristics, prognostic factors, or concomitant exposures (e.g. smoking). A confounder is a difference between the comparison groups and it influences the direction of the study results. A high quality study at the level of cohort design will identify the potential confounders and measure them (where possible). This is difficult for studies where behavioral, attitudinal or lifestyle factors may impact on the results.

#### 6. Were strategies to deal with confounding factors stated?

Strategies to deal with effects of confounding factors may be dealt within the study design or in data analysis. By matching or stratifying sampling of participants, effects of confounding factors can be adjusted for. When dealing with adjustment in data analysis, assess the statistics used in the study. Most will be some form of multivariate regression analysis to account for the confounding factors measured.

© JBI, 2020. All rights reserved. JBI grants use of these tools for research purposes only. All other enquiries should be sent to.

#### 7. Were the outcomes measured in a valid and reliable way?

Read the methods section of the paper. If for e.g. lung cancer is assessed based on existing definitions or diagnostic criteria, then the answer to this question is likely to be yes. If lung cancer is assessed using observer reported, or self-reported scales, the risk of over- or under-reporting is increased, and objectivity is compromised. Importantly, determine if the measurement tools used were validated instruments as this has a significant impact on outcome assessment validity.

Having established the objectivity of the outcome measurement (e.g. lung cancer) instrument, it's important to establish how the measurement was conducted. Were those involved in collecting data trained or educated in the use of the instrument/s? (e.g. radiographers). If there was more than one data collector, were they similar in terms of level of education, clinical or research experience, or level of responsibility in the piece of research being appraised?

#### 8. Was appropriate statistical analysis used?

As with any consideration of statistical analysis, consideration should be given to whether there was a more appropriate alternate statistical method that could have been used. The methods section should be detailed enough for reviewers to identify which analytical techniques were used (in particular, regression or stratification) and how specific confounders were measured.

For studies utilizing regression analysis, it is useful to identify if the study identified which variables were included and how they related to the outcome. If stratification was the analytical approach used, were the strata of analysis defined by the specified variables? Additionally, it is also important to assess the appropriateness of the analytical strategy in terms of the assumptions associated with the approach as differing methods of analysis are based on differing assumptions about the data and how it will respond.

© JBI, 2020. All rights reserved. JBI grants use of these tools for research purposes only. All other enquiries should be sent to.

### JBI CRITICAL APPRAISAL CHECKLIST FOR CASE REPORTS

Reviewer: Daniel de la Rosa Martinez Date: 15-01-21

Author: Tale et al Year: 2020 Record Number: 53_32814978_DDM

| | Yes | No | Unclear | Not applicable |
|---|---|---|---|---|
| 1. Were patient's demographic characteristics clearly described? | ☐ | ☐ | ☒ | ☐ |
| 2. Was the patient's history clearly described and presented as a timeline? | ☒ | ☐ | ☐ | ☐ |
| 3. Was the current clinical condition of the patient on presentation clearly described? | ☒ | ☐ | ☐ | ☐ |
| 4. Were diagnostic tests or assessment methods and the results clearly described? | ☒ | ☐ | ☐ | ☐ |
| 5. Was the intervention(s) or treatment procedure(s) clearly described? | ☐ | ☐ | ☐ | ☒ |
| 6. Was the post-intervention clinical condition clearly described? | ☐ | ☐ | ☐ | ☒ |
| 7. Were adverse events (harms) or unanticipated events identified and described? | ☐ | ☐ | ☒ | ☐ |
| 8. Does the case report provide takeaway lessons? | ☒ | ☐ | ☐ | ☐ |

Overall appraisal: Include ☐ Exclude ☒ Seek further info ☐

Comments (Including reason for exclusion)

Case report

© JBI, 2020. All rights reserved. JBI grants use of these tools for research purposes only. All other enquiries should be sent to.

### JBI CRITICAL APPRAISAL CHECKLIST FOR SYSTEMATIC REVIEWS AND RESEARCH SYNTHESES

Reviewer: Daniel de la Rosa    Date: 21-11-20

Author: J. R. Kelly et al    Year: 2020    Record Number: 54_32811575

| | | Yes | No | Unclear | Not applicable |
|---|---|---|---|---|---|
| 1. | Is the review question clearly and explicitly stated? | ☒ | ☐ | ☐ | ☐ |
| 2. | Were the inclusion criteria appropriate for the review question? | ☐ | ☐ | ☐ | ☒ |
| 3. | Was the search strategy appropriate? | ☐ | ☐ | ☐ | ☒ |
| 4. | Were the sources and resources used to search for studies adequate? | ☐ | ☐ | ☐ | ☒ |
| 5. | Were the criteria for appraising studies appropriate? | ☐ | ☐ | ☐ | ☒ |
| 6. | Was critical appraisal conducted by two or more reviewers independently? | ☐ | ☐ | ☐ | ☒ |
| 7. | Were there methods to minimize errors in data extraction? | ☐ | ☐ | ☐ | ☒ |
| 8. | Were the methods used to combine studies appropriate? | ☐ | ☐ | ☐ | ☒ |
| 9. | Was the likelihood of publication bias assessed? | ☐ | ☐ | ☐ | ☒ |
| 10. | Were recommendations for policy and/or practice supported by the reported data? | ☐ | ☐ | ☒ | ☐ |
| 11. | Were the specific directives for new research appropriate? | ☒ | ☐ | ☐ | ☐ |

Overall appraisal: Include ☐ Exclude ☒ Seek further info ☐

Comments (Including reason for exclusion)

Review - Excludes

### JBI CRITICAL APPRAISAL CHECKLIST FOR ANALYTICAL CROSS SECTIONAL STUDIES

Reviewer: Daniel de la Rosa  Date: 16-01-21

Author: Christine Miaskowski et al  Year: 2020  Record Number: 55_32809060

| | Yes | No | Unclear | Not applicable |
|---|---|---|---|---|
| 1. Were the criteria for inclusion in the sample clearly defined? | ☒ | ☐ | ☐ | ☐ |
| 2. Were the study subjects and the setting described in detail? | ☒ | ☐ | ☐ | ☐ |
| 3. Was the exposure measured in a valid and reliable way? | ☒ | ☐ | ☐ | ☐ |
| 4. Were objective, standard criteria used for measurement of the condition? | ☒ | ☐ | ☐ | ☐ |
| 5. Were confounding factors identified? | ☐ | ☒ | ☐ | ☐ |
| 6. Were strategies to deal with confounding factors stated? | ☐ | ☒ | ☐ | ☐ |
| 7. Were the outcomes measured in a valid and reliable way? | ☒ | ☐ | ☐ | ☐ |
| 8. Was appropriate statistical analysis used? | ☒ | ☐ | ☐ | ☐ |

Overall appraisal: Include ☐ Exclude ☒ Seek further info ☐

Comments (Including reason for exclusion)

No information about PACS

### JBI CRITICAL APPRAISAL CHECKLIST FOR ANALYTICAL CROSS SECTIONAL STUDIES

Reviewer Daniel de la Rosa Date 20-01-21

Author Ding Shi, et al Year 2020 Record Number 56_32614392_DDM

| | Yes | No | Unclear | Not applicable |
|---|---|---|---|---|
| 1. Were the criteria for inclusion in the sample clearly defined? | ☒ | ☐ | ☐ | ☐ |
| 2. Were the study subjects and the setting described in detail? | ☒ | ☐ | ☐ | ☐ |
| 3. Was the exposure measured in a valid and reliable way? | ☒ | ☐ | ☐ | ☐ |
| 4. Were objective, standard criteria used for measurement of the condition? | ☒ | ☐ | ☐ | ☐ |
| 5. Were confounding factors identified? | ☒ | ☐ | ☐ | ☐ |
| 6. Were strategies to deal with confounding factors stated? | ☒ | ☐ | ☐ | ☐ |
| 7. Were the outcomes measured in a valid and reliable way? | ☒ | ☐ | ☐ | ☐ |
| 8. Was appropriate statistical analysis used? | ☒ | ☐ | ☐ | ☐ |

Overall appraisal: Include ☐ Exclude ☒ Seek further info ☐

Comments (Including reason for exclusion)

No information about long-term manifestations

### JBI CRITICAL APPRAISAL CHECKLIST FOR ANALYTICAL CROSS SECTIONAL STUDIES

Reviewer: Daniel de la Rosa Date: 22-11-20

Author: Martijn A. Spruit et al Year: 2020 Record Number: 58_32817258

| | Yes | No | Unclear | Not applicable |
|---|---|---|---|---|
| 1. Were the criteria for inclusion in the sample clearly defined? | ☐ | ☐ | ☒ | ☐ |
| 2. Were the study subjects and the setting described in detail? | ☐ | ☐ | ☐ | ☒ |
| 3. Was the exposure measured in a valid and reliable way? | ☐ | ☐ | ☐ | ☒ |
| 4. Were objective, standard criteria used for measurement of the condition? | ☐ | ☐ | ☐ | ☒ |
| 5. Were confounding factors identified? | ☐ | ☐ | ☐ | ☒ |
| 6. Were strategies to deal with confounding factors stated? | ☐ | ☐ | ☐ | ☒ |
| 7. Were the outcomes measured in a valid and reliable way? | ☐ | ☐ | ☐ | ☒ |
| 8. Was appropriate statistical analysis used? | ☐ | ☐ | ☐ | ☒ |

Overall appraisal: Include ☐ Exclude ☒ Seek further info ☐

Comments (Including reason for exclusion)

Guideliness based on expert opinion

### JBI Critical Appraisal Checklist for Case Series

Reviewer Daniel de la Rosa Martinez Date 04/01/21

Author Martin A. Vilela-Estradaa Year 2020 Record Number 59_32804922

| | Yes | No | Unclear | Not applicable |
|---|---|---|---|---|
| • Were there clear criteria for inclusion in the case series? | ☐ | ✓ | ☐ | ☐ |
| • Was the condition measured in a standard, reliable way for all participants included in the case series? | ✓ | ☐ | ☐ | ☐ |
| • Were valid methods used for identification of the condition for all participants included in the case series? | ✓ | ☐ | ☐ | ☐ |
| • Did the case series have consecutive inclusion of participants? | ✓ | ☐ | ☐ | ☐ |
| • Did the case series have complete inclusion of participants? | ✓ | ☐ | ☐ | ☐ |
| • Was there clear reporting of the demographics of the participants in the study? | ✓ | ☐ | ☐ | ☐ |
| • Was there clear reporting of clinical information of the participants? | ✓ | ☐ | ☐ | ☐ |
| • Were the outcomes or follow up results of cases clearly reported? | ✓ | ☐ | ☐ | ☐ |
| • Was there clear reporting of the presenting site(s)/clinic(s) demographic information? | ✓ | ☐ | ☐ | ☐ |
| • Was statistical analysis appropriate? | ☐ | ☐ | ☐ | ✓ |

Overall appraisal: Include ☐ Exclude ✓ Seek further info ☐

Comments (Including reason for exclusion)

© JBI, 2020. All rights reserved. JBI grants use of these tools for research purposes only. All other enquiries should be sent to.

### JBI CRITICAL APPRAISAL CHECKLIST FOR CASE REPORTS

Reviewer: Daniel de la Rosa  Date: 21-01-21

Author: Pedro Augusto Sampaio Rocha-Filho,  Year: 2020  Record Number: 60_32790179

| | Yes | No | Unclear | Not applicable |
|---|---|---|---|---|
| 1. Were patient's demographic characteristics clearly described? | ☑ | ☐ | ☐ | ☐ |
| 2. Was the patient's history clearly described and presented as a timeline? | ☑ | ☐ | ☐ | ☐ |
| 3. Was the current clinical condition of the patient on presentation clearly described? | ☑ | ☐ | ☐ | ☐ |
| 4. Were diagnostic tests or assessment methods and the results clearly described? | ☑ | ☐ | ☐ | ☐ |
| 5. Was the intervention(s) or treatment procedure(s) clearly described? | ☑ | ☐ | ☐ | ☐ |
| 6. Was the post-intervention clinical condition clearly described? | ☑ | ☐ | ☐ | ☐ |
| 7. Were adverse events (harms) or unanticipated events identified and described? | ☑ | ☐ | ☐ | ☐ |
| 8. Does the case report provide takeaway lessons? | ☑ | ☐ | ☐ | ☐ |

Overall appraisal: Include ☐ Exclude ☑ Seek further info ☐

Comments (Including reason for exclusion)

Case report

© JBI, 2020. All rights reserved. JBI grants use of these tools for research purposes only. All other enquiries should be sent to.

### JBI CRITICAL APPRAISAL CHECKLIST FOR ANALYTICAL CROSS SECTIONAL STUDIES

Reviewer Daniel de la Rosa Date 26-01-21

Author Salma Batool-Anwar, Year 2020 Record Number 61_32780011

| | Yes | No | Unclear | Not applicable |
|---|---|---|---|---|
| 1. Were the criteria for inclusion in the sample clearly defined? | ☒ | ☐ | ☐ | ☐ |
| 2. Were the study subjects and the setting described in detail? | ☒ | ☐ | ☐ | ☐ |
| 3. Was the exposure measured in a valid and reliable way? | ☒ | ☐ | ☐ | ☐ |
| 4. Were objective, standard criteria used for measurement of the condition? | ☒ | ☐ | ☐ | ☐ |
| 5. Were confounding factors identified? | ☐ | ☒ | ☐ | ☐ |
| 6. Were strategies to deal with confounding factors stated? | ☐ | ☒ | ☐ | ☐ |
| 7. Were the outcomes measured in a valid and reliable way? | ☒ | ☐ | ☐ | ☐ |
| 8. Was appropriate statistical analysis used? | ☒ | ☐ | ☐ | ☐ |

Overall appraisal: Include ☐ Exclude ☒ Seek further info ☐

Comments (Including reason for exclusion)

No information about PACS

### JBI CRITICAL APPRAISAL CHECKLIST FOR ANALYTICAL CROSS SECTIONAL STUDIES

Reviewer______Daniel de la Rosa Martinez______Date________29/11/20______________

Author_________Carfi____________ Year__2020___ Record Number__ 62_32644129_______

| | Yes | No | Unclear | Not applicable |
|---|---|---|---|---|
| 1. Were the criteria for inclusion in the sample clearly defined? | ☒ | ☐ | ☐ | ☐ |
| 2. Were the study subjects and the setting described in detail? | ☒ | ☐ | ☐ | ☐ |
| 3. Was the exposure measured in a valid and reliable way? | ☒ | ☐ | ☐ | ☐ |
| 4. Were objective, standard criteria used for measurement of the condition? | ☒ | ☐ | ☐ | ☐ |
| 5. Were confounding factors identified? | ☐ | ☐ | ☒ | ☐ |
| 6. Were strategies to deal with confounding factors stated? | ☐ | ☐ | ☒ | ☐ |
| 7. Were the outcomes measured in a valid and reliable way? | ☒ | ☐ | ☐ | ☐ |
| 8. Was appropriate statistical analysis used? | ☒ | ☐ | ☐ | ☐ |

Overall appraisal: Include ☒ Exclude ☐ Seek further info ☐

Comments (Including reason for exclusion)

______________________________________________

______________________________________________

______________________________________________

### EXPLANATION OF ANALYTICAL CROSS SECTIONAL STUDIES CRITICAL APPRAISAL

*How to cite:* Moola S, Munn Z, Tufanaru C, Aromataris E, Sears K, Sfetcu R, Currie M, Qureshi R, Mattis P, Lisy K, Mu P-F. Chapter 7: Systematic reviews of etiology and risk . In: Aromataris E, Munn Z (Editors). *JBI Manual for Evidence Synthesis*. JBI, 2020. Available from https://synthesismanual.jbi.global

**Analytical cross sectional studies Critical Appraisal Tool**

Answers: Yes, No, Unclear or Not/Applicable

#### 1. Were the criteria for inclusion in the sample clearly defined?

The authors should provide clear inclusion and exclusion criteria that they developed prior to recruitment of the study participants. The inclusion/exclusion criteria should be specified (e.g., risk, stage of disease progression) with sufficient detail and all the necessary information critical to the study.

#### 2. Were the study subjects and the setting described in detail?

The study sample should be described in sufficient detail so that other researchers can determine if it is comparable to the population of interest to them. The authors should provide a clear description of the population from which the study participants were selected or recruited, including demographics, location, and time period.

#### 3. Was the exposure measured in a valid and reliable way?

The study should clearly describe the method of measurement of exposure. Assessing validity requires that a 'gold standard' is available to which the measure can be compared. The validity of exposure measurement usually relates to whether a current measure is appropriate or whether a measure of past exposure is needed.

Reliability refers to the processes included in an epidemiological study to check repeatability of measurements of the exposures. These usually include intra-observer reliability and inter-observer reliability.

#### 4. Were objective, standard criteria used for measurement of the condition?

It is useful to determine if patients were included in the study based on either a specified diagnosis or definition. This is more likely to decrease the risk of bias. Characteristics are another useful approach to matching groups, and studies that did not use specified diagnostic methods or definitions should provide evidence on matching by key characteristics

#### 5. Were confounding factors identified?

Confounding has occurred where the estimated intervention exposure effect is biased by the presence of some difference between the comparison groups (apart from the exposure investigated/of interest). Typical confounders include baseline characteristics, prognostic factors, or concomitant exposures (e.g. smoking). A confounder is a difference between the comparison groups and it influences the direction of the study results. A high quality study at the level of cohort design will identify the potential confounders and measure them (where possible). This is difficult for studies where behavioral, attitudinal or lifestyle factors may impact on the results.

#### 6. Were strategies to deal with confounding factors stated?

Strategies to deal with effects of confounding factors may be dealt within the study design or in data analysis. By matching or stratifying sampling of participants, effects of confounding factors can be adjusted for. When dealing with adjustment in data analysis, assess the statistics used in the study. Most will be some form of multivariate regression analysis to account for the confounding factors measured.

© JBI, 2020. All rights reserved. JBI grants use of these tools for research purposes only. All other enquiries should be sent to.

#### 7. Were the outcomes measured in a valid and reliable way?

Read the methods section of the paper. If for e.g. lung cancer is assessed based on existing definitions or diagnostic criteria, then the answer to this question is likely to be yes. If lung cancer is assessed using observer reported, or self-reported scales, the risk of over- or under-reporting is increased, and objectivity is compromised. Importantly, determine if the measurement tools used were validated instruments as this has a significant impact on outcome assessment validity.

Having established the objectivity of the outcome measurement (e.g. lung cancer) instrument, it's important to establish how the measurement was conducted. Were those involved in collecting data trained or educated in the use of the instrument/s? (e.g. radiographers). If there was more than one data collector, were they similar in terms of level of education, clinical or research experience, or level of responsibility in the piece of research being appraised?

#### 8. Was appropriate statistical analysis used?

As with any consideration of statistical analysis, consideration should be given to whether there was a more appropriate alternate statistical method that could have been used. The methods section should be detailed enough for reviewers to identify which analytical techniques were used (in particular, regression or stratification) and how specific confounders were measured.

For studies utilizing regression analysis, it is useful to identify if the study identified which variables were included and how they related to the outcome. If stratification was the analytical approach used, were the strata of analysis defined by the specified variables? Additionally, it is also important to assess the appropriateness of the analytical strategy in terms of the assumptions associated with the approach as differing methods of analysis are based on differing assumptions about the data and how it will respond.

© JBI, 2020. All rights reserved. JBI grants use of these tools for research purposes only. All other enquiries should be sent to.

### JBI CRITICAL APPRAISAL CHECKLIST FOR COHORT STUDIES

Reviewer: Daniel de la Rosa    Date: 30-01-21

Author: Vallari Shah, et al    Year: 2020    Record Number: 63_32526039

| | Yes | No | Unclear | Not applicable |
|---|---|---|---|---|
| 1. Were the two groups similar and recruited from the same population? | ☒ | ☐ | ☐ | ☐ |
| 2. Were the exposures measured similarly to assign people to both exposed and unexposed groups? | ☒ | ☐ | ☐ | ☐ |
| 3. Was the exposure measured in a valid and reliable way? | ☒ | ☐ | ☐ | ☐ |
| 4. Were confounding factors identified? | ☐ | ☐ | ☒ | ☐ |
| 5. Were strategies to deal with confounding factors stated? | ☐ | ☐ | ☒ | ☐ |
| 6. Were the groups/participants free of the outcome at the start of the study (or at the moment of exposure)? | ☐ | ☐ | ☒ | ☐ |
| 7. Were the outcomes measured in a valid and reliable way? | ☒ | ☐ | ☐ | ☐ |
| 8. Was the follow up time reported and sufficient to be long enough for outcomes to occur? | ☒ | ☐ | ☐ | ☐ |
| 9. Was follow up complete, and if not, were the reasons to loss to follow up described and explored? | ☐ | ☐ | ☐ | ☒ |
| 10. Were strategies to address incomplete follow up utilized? | ☐ | ☐ | ☐ | ☒ |
| 11. Was appropriate statistical analysis used? | ☒ | ☐ | ☐ | ☐ |

Overall appraisal: Include ☐ Exclude ☒ Seek further info ☐

Comments (Including reason for exclusion)

No information about PACS

© JBI, 2020. All rights reserved. JBI grants use of these tools for research purposes only. All other enquiries should be sent to.

### JBI CRITICAL APPRAISAL CHECKLIST FOR CASE REPORTS

Reviewer Daniel de la Rosa Date 31-01-21

Author Sam K. Touisserkani et al Year 2020 Record Number 64_32802540

| | Yes | No | Unclear | Not applicable |
|---|---|---|---|---|
| 1. Were patient's demographic characteristics clearly described? | ☐ | ☒ | ☐ | ☐ |
| 2. Was the patient's history clearly described and presented as a timeline? | ☐ | ☒ | ☐ | ☐ |
| 3. Was the current clinical condition of the patient on presentation clearly described? | ☒ | ☐ | ☐ | ☐ |
| 4. Were diagnostic tests or assessment methods and the results clearly described? | ☒ | ☐ | ☐ | ☐ |
| 5. Was the intervention(s) or treatment procedure(s) clearly described? | ☒ | ☐ | ☐ | ☐ |
| 6. Was the post-intervention clinical condition clearly described? | ☐ | ☐ | ☐ | ☒ |
| 7. Were adverse events (harms) or unanticipated events identified and described? | ☐ | ☐ | ☐ | ☒ |
| 8. Does the case report provide takeaway lessons? | ☒ | ☐ | ☐ | ☐ |

Overall appraisal: Include ☐ Exclude ☒ Seek further info ☐

Comments (Including reason for exclusion)

© JBI, 2020. All rights reserved. JBI grants use of these tools for research purposes only. All other enquiries should be sent to.

### JBI CRITICAL APPRAISAL CHECKLIST FOR COHORT STUDIES

Reviewer: Daniel de la Rosa Martinez  Date: 31-01-21

Author: Carlo Gaspardone, et al.  Year: 2020  Record Number: 65_32770687

| | Yes | No | Unclear | Not applicable |
|---|---|---|---|---|
| 1. Were the two groups similar and recruited from the same population? | ☑ | ☐ | ☐ | ☐ |
| 2. Were the exposures measured similarly to assign people to both exposed and unexposed groups? | ☑ | ☐ | ☐ | ☐ |
| 3. Was the exposure measured in a valid and reliable way? | ☑ | ☐ | ☐ | ☐ |
| 4. Were confounding factors identified? | ☐ | ☑ | ☐ | ☐ |
| 5. Were strategies to deal with confounding factors stated? | ☐ | ☑ | ☐ | ☐ |
| 6. Were the groups/participants free of the outcome at the start of the study (or at the moment of exposure)? | ☐ | ☐ | ☐ | ☑ |
| 7. Were the outcomes measured in a valid and reliable way? | ☑ | ☐ | ☐ | ☐ |
| 8. Was the follow up time reported and sufficient to be long enough for outcomes to occur? | ☐ | ☐ | ☑ | ☐ |
| 9. Was follow up complete, and if not, were the reasons to loss to follow up described and explored? | ☐ | ☐ | ☑ | ☐ |
| 10. Were strategies to address incomplete follow up utilized? | ☐ | ☑ | ☐ | ☐ |
| 11. Was appropriate statistical analysis used? | ☑ | ☐ | ☐ | ☐ |

Overall appraisal: Include ☐ Exclude ☑ Seek further info ☐

Comments (Including reason for exclusion)

No comments.

© JBI, 2020. All rights reserved. JBI grants use of these tools for research purposes only. All other enquiries should be sent to.

### JBI CRITICAL APPRAISAL CHECKLIST FOR CASE REPORTS

Reviewer Daniel de la Rosa Date 31-01-20

Author Xuemin Guo et al Year________ Record Number 66_32849650

| | Yes | No | Unclear | Not applicable |
|---|---|---|---|---|
| 1. Were patient's demographic characteristics clearly described? | ☑ | ☐ | ☐ | ☐ |
| 2. Was the patient's history clearly described and presented as a timeline? | ☑ | ☐ | ☐ | ☐ |
| 3. Was the current clinical condition of the patient on presentation clearly described? | ☑ | ☐ | ☐ | ☐ |
| 4. Were diagnostic tests or assessment methods and the results clearly described? | ☑ | ☐ | ☐ | ☐ |
| 5. Was the intervention(s) or treatment procedure(s) clearly described? | ☑ | ☐ | ☐ | ☐ |
| 6. Was the post-intervention clinical condition clearly described? | ☑ | ☐ | ☐ | ☐ |
| 7. Were adverse events (harms) or unanticipated events identified and described? | ☑ | ☐ | ☐ | ☐ |
| 8. Does the case report provide takeaway lessons? | ☑ | ☐ | ☐ | ☐ |

Overall appraisal: Include ☐ Exclude ☑ Seek further info ☐

Comments (Including reason for exclusion)

Case report

© JBI, 2020. All rights reserved. JBI grants use of these tools for research purposes only. All other enquiries should be sent to.

### JBI CRITICAL APPRAISAL CHECKLIST FOR QUASI-EXPERIMENTAL STUDIES

Reviewer: Isaac David Juárez Cruz  Date: 14/11/2020

Author: Supeng Wang, Yang Pan, Quanyi Wang, Huangyu Miao, Ashley N. Brown, Libin Rong  Year: 2020  Record Number: 67_32771304_IDJC

| | Yes | No | Unclear | Not applicable |
|---|---|---|---|---|
| 1. Is it clear in the study what is the 'cause' and what is the 'effect' (i.e. there is no confusion about which variable comes first)? | ☑ | ☐ | ☐ | ☐ |
| 2. Were the participants included in any comparisons similar? | ☐ | ☐ | ☑ | ☐ |
| 3. Were the participants included in any comparisons receiving similar treatment/care, other than the exposure or intervention of interest? | ☑ | ☐ | ☐ | ☐ |
| 4. Was there a control group? | ☐ | ☐ | ☐ | ☑ |
| 5. Were there multiple measurements of the outcome both pre and post the intervention/exposure? | ☑ | ☐ | ☐ | ☐ |
| 6. Was follow up complete and if not, were differences between groups in terms of their follow up adequately described and analyzed? | ☑ | ☐ | ☐ | ☐ |
| 7. Were the outcomes of participants included in any comparisons measured in the same way? | ☑ | ☐ | ☐ | ☐ |
| 8. Were outcomes measured in a reliable way? | ☑ | ☐ | ☐ | ☐ |
| 9. Was appropriate statistical analysis used? | ☑ | ☐ | ☐ | ☐ |

Overall appraisal: Include ☐ Exclude ☑ Seek further info ☐

Comments (Including reason for exclusion)

Se excluye este Artículo, no por su cálidad, sino porque no cumple con la información de interés para la revisión. No hace mención a datos o información que nos sean útiles por el momento.

© JBI, 2020. All rights reserved. JBI grants use of these tools for research purposes only. All other enquiries should be sent to.

### JBI CRITICAL APPRAISAL CHECKLIST FOR ANALYTICAL CROSS SECTIONAL STUDIES

Reviewer Daniel de la Rosa Date 31-01-21

Author Vaira LA, et al Year 2020 Record Number 68_32762737

| | Yes | No | Unclear | Not applicable |
|---|---|---|---|---|
| 1. Were the criteria for inclusion in the sample clearly defined? | ☒ | ☐ | ☐ | ☐ |
| 2. Were the study subjects and the setting described in detail? | ☒ | ☐ | ☐ | ☐ |
| 3. Was the exposure measured in a valid and reliable way? | ☒ | ☐ | ☐ | ☐ |
| 4. Were objective, standard criteria used for measurement of the condition? | ☒ | ☐ | ☐ | ☐ |
| 5. Were confounding factors identified? | ☐ | ☒ | ☐ | ☐ |
| 6. Were strategies to deal with confounding factors stated? | ☐ | ☒ | ☐ | ☐ |
| 7. Were the outcomes measured in a valid and reliable way? | ☒ | ☐ | ☐ | ☐ |
| 8. Was appropriate statistical analysis used? | ☒ | ☐ | ☐ | ☐ |

Overall appraisal: Include ☐ Exclude ☒ Seek further info ☐

Comments (Including reason for exclusion)

Follow up time less than 21 days

### JBI CRITICAL APPRAISAL CHECKLIST FOR ANALYTICAL CROSS SECTIONAL STUDIES

Reviewer: Daniel de la Rosa  Date: 31-01-21

Author: Cristina Corsini Campiolia, et al.  Year: 2020  Record Number: 69_32777762

| | Yes | No | Unclear | Not applicable |
|---|---|---|---|---|
| 1. Were the criteria for inclusion in the sample clearly defined? | ☒ | ☐ | ☐ | ☐ |
| 2. Were the study subjects and the setting described in detail? | ☒ | ☐ | ☐ | ☐ |
| 3. Was the exposure measured in a valid and reliable way? | ☒ | ☐ | ☐ | ☐ |
| 4. Were objective, standard criteria used for measurement of the condition? | ☒ | ☐ | ☐ | ☐ |
| 5. Were confounding factors identified? | ☐ | ☒ | ☐ | ☐ |
| 6. Were strategies to deal with confounding factors stated? | ☐ | ☒ | ☐ | ☐ |
| 7. Were the outcomes measured in a valid and reliable way? | ☒ | ☐ | ☐ | ☐ |
| 8. Was appropriate statistical analysis used? | ☒ | ☐ | ☐ | ☐ |

Overall appraisal: Include ☐ Exclude ☒ Seek further info ☐

Comments (Including reason for exclusion)

Follow up time less than 21 days

### JBI CRITICAL APPRAISAL CHECKLIST FOR COHORT STUDIES

Reviewer: Isaac David Juárez Cruz  Date: 14/11/20

Author: Pon et-Megemont L, Paris P, Tronchere A, Salazar JP, Pereira B, Dallel R, Aumeran C, Beytout J, Jacome C, Laurichesse H, Lessens O, Mrozeck N, Vidal M, Moisés X.  Year: 2020  Record Number: 70_32757419_IDJC

| | Yes | No | Unclear | Not applicable |
|---|---|---|---|---|
| 1. Were the two groups similar and recruited from the same population? | ✓ | ☐ | ☐ | ☐ |
| 2. Were the exposures measured similarly to assign people to both exposed and unexposed groups? | ✓ | ☐ | ☐ | ☐ |
| 3. Was the exposure measured in a valid and reliable way? | ☐ | ☐ | ✓ | ☐ |
| 4. Were confounding factors identified? | ☐ | ✓ | ☐ | ☐ |
| 5. Were strategies to deal with confounding factors stated? | ☐ | ✓ | ☐ | ☐ |
| 6. Were the groups/participants free of the outcome at the start of the study (or at the moment of exposure)? | ✓ | ☐ | ☐ | ☐ |
| 7. Were the outcomes measured in a valid and reliable way? | ☐ | ☐ | ✓ | ☐ |
| 8. Was the follow up time reported and sufficient to be long enough for outcomes to occur? | ✓ | ☐ | ☐ | ☐ |
| 9. Was follow up complete, and if not, were the reasons to loss to follow up described and explored? | ✓ | ☐ | ☐ | ☐ |
| 10. Were strategies to address incomplete follow up utilized? | ☐ | ☐ | ☐ | ✓ |
| 11. Was appropriate statistical analysis used? | ✓ | ☐ | ☐ | ☐ |

Overall appraisal: Include ☐ Exclude ✓ Seek further info ☐

Comments (Including reason for exclusion)

Se incluye. Aporta información sobre la persistencia de síntomas neurologicos, su relación con cefales y la prevalencia de dolores de cabeza durante fase aguda y post infección.

© JBI, 2020. All rights reserved. JBI grants use of these tools for research purposes only. All other enquiries should be sent to.

### JBI CRITICAL APPRAISAL CHECKLIST FOR SYSTEMATIC REVIEWS AND RESEARCH SYNTHESES

Reviewer Isaac David Juárez Cruz Date 15/11/20

Author A. Demeo, N. Marorta, M. Barletta, I. Pino, C. Marinero, A. Petraroli, L. Moggio y Antonio Amendolia. Year 2020 Record Number 71_32840156_IDJC

| | Yes | No | Unclear | Not applicable |
|---|---|---|---|---|
| 1. Is the review question clearly and explicitly stated? | ☑ | ☐ | ☐ | ☐ |
| 2. Were the inclusion criteria appropriate for the review question? | ☑ | ☐ | ☐ | ☐ |
| 3. Was the search strategy appropriate? | ☑ | ☐ | ☐ | ☐ |
| 4. Were the sources and resources used to search for studies adequate? | ☑ | ☐ | ☐ | ☐ |
| 5. Were the criteria for appraising studies appropriate? | ☑ | ☐ | ☐ | ☐ |
| 6. Was critical appraisal conducted by two or more reviewers independently? | ☑ | ☐ | ☐ | ☐ |
| 7. Were there methods to minimize errors in data extraction? | ☐ | ☐ | ☑ | ☐ |
| 8. Were the methods used to combine studies appropriate? | ☑ | ☐ | ☐ | ☐ |
| 9. Was the likelihood of publication bias assessed? | ☑ | ☐ | ☐ | ☐ |
| 10. Were recommendations for policy and/or practice supported by the reported data? | ☑ | ☐ | ☐ | ☐ |
| 11. Were the specific directives for new research appropriate? | ☑ | ☐ | ☐ | ☐ |

Overall appraisal: Include ☐ Exclude ☑ Seek further info ☐

Comments (Including reason for exclusion)

Se excluye, si bien es una buena revisión, no hace mención de síntomas persistentes o algún dato de utilidad para nuestra revisión, por el hecho de que es una revisión muy temprana, lo hay muchos datos disponibles.

© JBI, 2020. All rights reserved. JBI grants use of these tools for research purposes only. All other enquiries should be sent to.

### CHECKLIST FOR ANALYTICAL CROSS SECTIONAL STUDIES

Critical Appraisal tools for use in JBI Systematic Reviews

### INTRODUCTION

JBI is an JBI is an international research organisation based in the Faculty of Health and Medical Sciences at the University of Adelaide, South Australia. JBI develops and delivers unique evidence-based information, software, education and training designed to improve healthcare practice and health outcomes. With over 70 Collaborating Entities, servicing over 90 countries, JBI is a recognised global leader in evidence-based healthcare.

#### JBI Systematic Reviews

The core of evidence synthesis is the systematic review of literature of a particular intervention, condition or issue. The systematic review is essentially an analysis of the available literature (that is, evidence) and a judgment of the effectiveness or otherwise of a practice, involving a series of complex steps. JBI takes a particular view on what counts as evidence and the methods utilised to synthesise those different types of evidence. In line with this broader view of evidence, JBI has developed theories, methodologies and rigorous processes for the critical appraisal and synthesis of these diverse forms of evidence in order to aid in clinical decision-making in healthcare. There now exists JBI guidance for conducting reviews of effectiveness research, qualitative research, prevalence/incidence, etiology/risk, economic evaluations, text/opinion, diagnostic test accuracy, mixed-methods, umbrella reviews and scoping reviews. Further information regarding JBI systematic reviews can be found in the JBI Evidence Synthesis Manual.

#### JBI Critical Appraisal Tools

All systematic reviews incorporate a process of critique or appraisal of the research evidence. The purpose of this appraisal is to assess the methodological quality of a study and to determine the extent to which a study has addressed the possibility of bias in its design, conduct and analysis. All papers selected for inclusion in the systematic review (that is – those that meet the inclusion criteria described in the protocol) need to be subjected to rigorous appraisal by two critical appraisers. The results of this appraisal can then be used to inform synthesis and interpretation of the results of the study. JBI Critical appraisal tools have been developed by the JBI and collaborators and approved by the JBI Scientific Committee following extensive peer review. Although designed for use in systematic reviews, JBI critical appraisal tools can also be used when creating Critically Appraised Topics (CAT), in journal clubs and as an educational tool.

© JBI, 2020. All rights reserved. JBI grants use of these tools for research purposes only. All other enquiries should be sent to.

### JBI CRITICAL APPRAISAL CHECKLIST FOR ANALYTICAL CROSS SECTIONAL STUDIES

Reviewer: Daniel de la Rosa Martinez. Date: lunes 26 de Abril 2021

Author: Anika Singanayagam, et al Year: 2020 Record Number: 72_32794447_DDM

| | Yes | No | Unclear | Not applicable |
|---|---|---|---|---|
| 1. Were the criteria for inclusion in the sample clearly defined? | ☐ | ■ | ☐ | ☐ |
| 2. Were the study subjects and the setting described in detail? | ☐ | ■ | ☐ | ☐ |
| 3. Was the exposure measured in a valid and reliable way? | ■ | ☐ | ☐ | ☐ |
| 4. Were objective, standard criteria used for measurement of the condition? | ☐ | ☐ | ■ | ☐ |
| 5. Were confounding factors identified? | ☐ | ■ | ☐ | ☐ |
| 6. Were strategies to deal with confounding factors stated? | ☐ | ■ | ☐ | ☐ |
| 7. Were the outcomes measured in a valid and reliable way? | ■ | ☐ | ☐ | ☐ |
| 8. Was appropriate statistical analysis used? | ■ | ☐ | ☐ | ☐ |

Overall appraisal: Include ☐ Exclude ■ Seek further info ☐

Comments (Including reason for exclusion)

No aporta datos en relacion a sintomas persistentes, hace referencia a niveles de CT y su asociacion a desenlaces clinicos

© JBI, 2020. All rights reserved. JBI grants use of these tools for research purposes only. All other enquiries should be sent to.

### EXPLANATION OF ANALYTICAL CROSS SECTIONAL STUDIES CRITICAL APPRAISAL

*How to cite:* Moola S, Munn Z, Tufanaru C, Aromataris E, Sears K, Sfetcu R, Currie M, Qureshi R, Mattis P, Lisy K, Mu P-F. Chapter 7: Systematic reviews of etiology and risk . In: Aromataris E, Munn Z (Editors). *JBI Manual for Evidence Synthesis*. JBI, 2020. Available from https://synthesismanual.jbi.global

**Analytical cross sectional studies Critical Appraisal Tool**

Answers: Yes, No, Unclear or Not/Applicable

#### 1. Were the criteria for inclusion in the sample clearly defined?

The authors should provide clear inclusion and exclusion criteria that they developed prior to recruitment of the study participants. The inclusion/exclusion criteria should be specified (e.g., risk, stage of disease progression) with sufficient detail and all the necessary information critical to the study.

#### 2. Were the study subjects and the setting described in detail?

The study sample should be described in sufficient detail so that other researchers can determine if it is comparable to the population of interest to them. The authors should provide a clear description of the population from which the study participants were selected or recruited, including demographics, location, and time period.

#### 3. Was the exposure measured in a valid and reliable way?

The study should clearly describe the method of measurement of exposure. Assessing validity requires that a 'gold standard' is available to which the measure can be compared. The validity of exposure measurement usually relates to whether a current measure is appropriate or whether a measure of past exposure is needed.

Reliability refers to the processes included in an epidemiological study to check repeatability of measurements of the exposures. These usually include intra-observer reliability and inter-observer reliability.

#### 4. Were objective, standard criteria used for measurement of the condition?

It is useful to determine if patients were included in the study based on either a specified diagnosis or definition. This is more likely to decrease the risk of bias. Characteristics are another useful approach to matching groups, and studies that did not use specified diagnostic methods or definitions should provide evidence on matching by key characteristics

#### 5. Were confounding factors identified?

Confounding has occurred where the estimated intervention exposure effect is biased by the presence of some difference between the comparison groups (apart from the exposure investigated/of interest). Typical confounders include baseline characteristics, prognostic factors, or concomitant exposures (e.g. smoking). A confounder is a difference between the comparison groups and it influences the direction of the study results. A high quality study at the level of cohort design will identify the potential confounders and measure them (where possible). This is difficult for studies where behavioral, attitudinal or lifestyle factors may impact on the results.

#### 6. Were strategies to deal with confounding factors stated?

Strategies to deal with effects of confounding factors may be dealt within the study design or in data analysis. By matching or stratifying sampling of participants, effects of confounding factors can be adjusted for. When dealing with adjustment in data analysis, assess the statistics used in the study. Most will be some form of multivariate regression analysis to account for the confounding factors measured.

© JBI, 2020. All rights reserved. JBI grants use of these tools for research purposes only. All other enquiries should be sent to.

#### 7. Were the outcomes measured in a valid and reliable way?

Read the methods section of the paper. If for e.g. lung cancer is assessed based on existing definitions or diagnostic criteria, then the answer to this question is likely to be yes. If lung cancer is assessed using observer reported, or self-reported scales, the risk of over- or under-reporting is increased, and objectivity is compromised. Importantly, determine if the measurement tools used were validated instruments as this has a significant impact on outcome assessment validity.

Having established the objectivity of the outcome measurement (e.g. lung cancer) instrument, it's important to establish how the measurement was conducted. Were those involved in collecting data trained or educated in the use of the instrument/s? (e.g. radiographers). If there was more than one data collector, were they similar in terms of level of education, clinical or research experience, or level of responsibility in the piece of research being appraised?

#### 8. Was appropriate statistical analysis used?

As with any consideration of statistical analysis, consideration should be given to whether there was a more appropriate alternate statistical method that could have been used. The methods section should be detailed enough for reviewers to identify which analytical techniques were used (in particular, regression or stratification) and how specific confounders were measured.

For studies utilizing regression analysis, it is useful to identify if the study identified which variables were included and how they related to the outcome. If stratification was the analytical approach used, were the strata of analysis defined by the specified variables? Additionally, it is also important to assess the appropriateness of the analytical strategy in terms of the assumptions associated with the approach as differing methods of analysis are based on differing assumptions about the data and how it will respond.

© JBI, 2020. All rights reserved. JBI grants use of these tools for research purposes only. All other enquiries should be sent to.

### JBI CRITICAL APPRAISAL CHECKLIST FOR CASE CONTROL STUDIES

Reviewer: Isaac David Juárez Cruz   Date: 18/11/20

Author: Vaira L, Hopkins C, Petrocelli M, Lechien J R, Chiesa-EstombaC M, Salzano G, Cucurullo M, Salzano F A, Saussez S, Boscolo-Rizzo P, Biglioli FDe Riu G.   Year: 2020   Record Number: 73_32782030_IDJC

| | Yes | No | Unclear | Not applicable |
|---|---|---|---|---|
| 1. Were the groups comparable other than the presence of disease in cases or the absence of disease in controls? | ☑ | ☐ | ☐ | ☐ |
| 2. Were cases and controls matched appropriately? | ☑ | ☐ | ☐ | ☐ |
| 3. Were the same criteria used for identification of cases and controls? | ☑ | ☐ | ☐ | ☐ |
| 4. Was exposure measured in a standard, valid and reliable way? | ☑ | ☐ | ☐ | ☐ |
| 5. Was exposure measured in the same way for cases and controls? | ☑ | ☐ | ☐ | ☐ |
| 6. Were confounding factors identified? | ☐ | ☐ | ☑ | ☐ |
| 7. Were strategies to deal with confounding factors stated? | ☐ | ☐ | ☑ | ☐ |
| 8. Were outcomes assessed in a standard, valid and reliable way for cases and controls? | ☑ | ☐ | ☐ | ☐ |
| 9. Was the exposure period of interest long enough to be meaningful? | ☑ | ☐ | ☐ | ☐ |
| 10. Was appropriate statistical analysis used? | ☑ | ☐ | ☐ | ☐ |

Overall appraisal: Include ☑ Exclude ☐ Seek further info ☐

Comments (Including reason for exclusion)

Se incluye, aporta mucha información de utilidad, asi como hace alusión a la frecuencia de sintomas persistentes. Sin embargo, reporta no encontrar relación o predictores en personas que presentaran sintomas persistentes

© JBI, 2020. All rights reserved. JBI grants use of these tools for research purposes only. All other enquiries should be sent to.

### JBI CRITICAL APPRAISAL CHECKLIST FOR ANALYTICAL CROSS SECTIONAL STUDIES

Reviewer Daniel Eduardo Sandoval Colin Date 03/04/2021

Author Alexander Wieck Fjaeldstad Year 2020 Record Number 74_32741438_DESC

| | Yes | No | Unclear | Not applicable |
|---|---|---|---|---|
| 1. Were the criteria for inclusion in the sample clearly defined? | ☐ | ☒ | ☐ | ☐ |
| 2. Were the study subjects and the setting described in detail? | ☐ | ☐ | ☒ | ☐ |
| 3. Was the exposure measured in a valid and reliable way? | ☐ | ☒ | ☐ | ☐ |
| 4. Were objective, standard criteria used for measurement of the condition? | ☐ | ☒ | ☐ | ☐ |
| 5. Were confounding factors identified? | ☐ | ☒ | ☐ | ☐ |
| 6. Were strategies to deal with confounding factors stated? | ☐ | ☒ | ☐ | ☐ |
| 7. Were the outcomes measured in a valid and reliable way? | ☐ | ☒ | ☐ | ☐ |
| 8. Was appropriate statistical analysis used? | ☐ | ☐ | ☒ | ☐ |

Overall appraisal: Include ☐ Exclude ☒ Seek further info ☐

Comments (Including reason for exclusion)

No incluye sujetos con prueba (PCR o antígenos) positiva para SARS-CoV2.

### JBI CRITICAL APPRAISAL CHECKLIST FOR TEXT AND OPINION PAPERS

Reviewer Daniel Eduardo Sandoval Colin Date 03/04/2021

Author Manuela Pennisi, Giuseppe Lanza, Luca Falzone et al Year 2020 Record Number 75_32751841_DESC

| | Yes | No | Unclear | Not applicable |
|---|---|---|---|---|
| 1. Is the source of the opinion clearly identified? | ☐ | ☐ | ☐ | ☐ |
| 2. Does the source of opinion have standing in the field of expertise? | ☐ | ☐ | ☐ | ☐ |
| 3. Are the interests of the relevant population the central focus of the opinion? | ☐ | ☐ | ☐ | ☐ |
| 4. Is the stated position the result of an analytical process, and is there logic in the opinion expressed? | ☐ | ☐ | ☐ | ☐ |
| 5. Is there reference to the extant literature? | ☐ | ☐ | ☐ | ☐ |
| 6. Is any incongruence with the literature/sources logically defended? | ☐ | ☐ | ☐ | ☐ |

Overall appraisal: Include ☐ Exclude ☒ Seek further info ☐

Comments (Including reason for exclusion)

No se realiza cuestionario porque no cumple con los criterios de inclusión.

Revisión sobre manifestaciones neurologicas agudas en pacientes con COVID-19

### JBI CRITICAL APPRAISAL CHECKLIST FOR ANALYTICAL CROSS SECTIONAL STUDIES

Reviewer Daniel Eduardo Sandoval Colin Date 03/04/2021

Author Mark W. Tenforde, Sara S. Kim, Christopher J. Lindsell, et al. Year 2020 Record Number 76_32730238_DESC

| | Yes | No | Unclear | Not applicable |
|---|---|---|---|---|
| 1. Were the criteria for inclusion in the sample clearly defined? | X | | | |
| 2. Were the study subjects and the setting described in detail? | X | | | |
| 3. Was the exposure measured in a valid and reliable way? | X | | | |
| 4. Were objective, standard criteria used for measurement of the condition? | X | | | |
| 5. Were confounding factors identified? | | | | X |
| 6. Were strategies to deal with confounding factors stated? | | | | X |
| 7. Were the outcomes measured in a valid and reliable way? | X | | | |
| 8. Was appropriate statistical analysis used? | X | | | |

Overall appraisal: Include ☐ Exclude ☐ Seek further info ☒

Comments (Including reason for exclusion)

Solicitar más información sobre las frecuencias de los síntomas no mencionados en el artículo.

OXFORD

*QJM: An International Journal of Medicine*, 2020, 837–838

doi: 10.1093/qjmed/hcaa255
Advance Access Publication Date: 19 August 2020
Clinical picture

CLINICAL PICTURE

### Post-COVID-19 pneumonia pulmonary fibrosis

#### Case

A 48-year-old male patient with no prior co-morbidities presented to the emergency department with complaints of dry cough, fever and throat pain for the preceding 5 days followed by shortness of breath for the past 2 days. He had no significant travel or contact history. His vitals were stable except for mild hypoxemia on room air ($SpO_2$ of 92%). On laboratory investigation he had lymphopenia, elevated Lactate dehydrogenase (LDH), raised C-reactive protein and elevated D-dimer levels. Initial chest radiograph revealed peripheral multiple inhomogeneous opacities in bilateral lung fields. His reverse transcriptase-polymerase chain reaction nasal and oropharyngeal swab for COVID-19 came out be positive and he was managed conservatively with supplemental oxygen, low molecular weight heparin, dexamethasone and antipyretics. He was symptomatically improved but continued to have hypoxemia even after 3 weeks of treatment, so a high resolution computed tomography of the chest was performed and it showed architectural distortion, interlobar septal thickening and traction bronchiectasis features (shown in Figure 1A and B) suggestive of fibrotic lung disease. He was discharged on home oxygen therapy and planned to enroll him in antifibrotic therapy trail during the subsequent follow-up.

#### Discussion

Clinical manifestations of Corona virus disease-2019 (COVID-19) have ranged from asymptomatic/mild symptoms to severe illness and mortality.[1] Most of the mild and moderate cases are recovered completely but a small proportion of severe cases with acute respiratory distress syndrome continued to remain hypoxemic despite adequate treatment. Chest imaging of this subset of patients revealed fibrotic changes in the form of traction bronchiectasis, architectural distortion and septal thickening similar to the changes seen in other fibrotic lung diseases.[2] The pathogenesis of post-infective pulmonary fibrosis includes dysregulated release of matrix metalloproteinases during the inflammatory phase of adult respiratory distress syndrome (ARDS) causing epithelial and endothelial injury with unchecked fibroproliferation. There is also a vascular dysfunction which is a key component of the switch from ARDS to fibrosis, with vascular endothelial growth factor and cytokines such as interleukin-6 and tumor necrosis factor alpha being implicated.[3] Although the role of presently available antifibrotic drugs (pirfenidone and nintedanib) for fibrotic lung diseases beyond idiopathic pulmonary fibrosis have been evaluated by some authors[4] their role in post-COVID-19 pneumonia pulmonary fibrosis need further research in the present pandemic.

Photographs and text from: S. Tale, Department of Pulmonary Medicine, All India Institute of Medical Sciences, Rishikesh 249203, India; S. Ghosh, Post Graduate Institute of Medical Education and Research, Chandigarh 160012, India; S.P. Meitei, All India Institute of Medical Sciences, Rishikesh 249203, India;

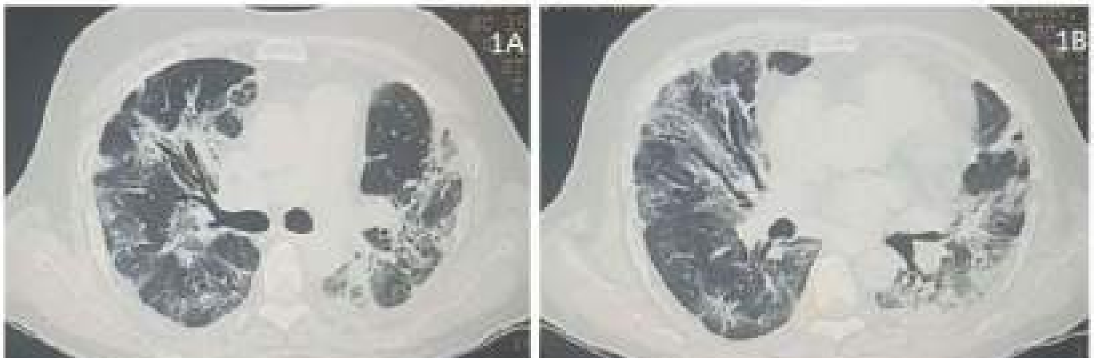


Figure 1. (A and B) Traction bronchiectasis, architectural distortion and interlobar septal thickening suggestive of pulmonary fibrosis.

© The Author(s) 2020. Published by Oxford University Press on behalf of the Association of Physicians. All rights reserved.
For permissions, please



Downloaded from https://academic.oup.com/qjmed/article/113/11/837/5894556 by guest on 24 January 2021

M. Kolli, Gandhi Medical College, Secunderabad 500025, India; A.K. Garbhapu, Dr. RML Medical College, Delhi 110001, India; S. Pudi, All India Institute of Medical Sciences, Rishikesh, India.

*Conflict of interest*: None declared.

#### References

1. Guan WJ, Ni ZY, Hu Y, Liang WH, Ou CQ, He ZX, et al. Clinical characteristics of coronavirus disease 2019 in China. *N Engl J Med* 2020; **382**:1708–20.
2. Kalchiem DO, Galvin JR, Burke AP, Atamas SP, Todd NW. Interstitial lung disease and pulmonary fibrosis: a practical approach for general medicine physicians with focus on the medical history. *J Clin Med* 2018; **7**:476.
3. Collins BF, Raghu G. Antifibrotic therapy for fibrotic lung disease beyond idiopathic pulmonary fibrosis. *Eur Respir Rev* 2019; **28**:190022.
4. Burnham EL, Janssen WJ, Riches DW, Moss M, Downey GP. The fibroproliferative response in acute respiratory distress syndrome: mechanisms and clinical significance. *Eur Respir J* 2014; **43**:276–85.

Downloaded from https://academic.oup.com/qjmed/article/113/11/837/5894556 by guest on 24 January 2021

### JBI CRITICAL APPRAISAL CHECKLIST FOR COHORT STUDIES

Reviewer: Isaac David Juárez Cruz  Date: 24/11/20

Author: Tingting Guo, Xiaoming Liu, Cihao Xu, Jiazheng Wang, Lian Yang, Heshui Shi, Meng Dai.  Year: 2020  Record Number: 77_32922173_IDJC

| | Yes | No | Unclear | Not applicable |
|---|---|---|---|---|
| 1. Were the two groups similar and recruited from the same population? | ☑ | ☐ | ☐ | ☐ |
| 2. Were the exposures measured similarly to assign people to both exposed and unexposed groups? | ☑ | ☐ | ☐ | ☐ |
| 3. Was the exposure measured in a valid and reliable way? | ☑ | ☐ | ☐ | ☐ |
| 4. Were confounding factors identified? | ☐ | ☐ | ☑ | ☐ |
| 5. Were strategies to deal with confounding factors stated? | ☐ | ☐ | ☑ | ☐ |
| 6. Were the groups/participants free of the outcome at the start of the study (or at the moment of exposure)? | ☑ | ☐ | ☐ | ☐ |
| 7. Were the outcomes measured in a valid and reliable way? | ☑ | ☐ | ☐ | ☐ |
| 8. Was the follow up time reported and sufficient to be long enough for outcomes to occur? | ☑ | ☐ | ☐ | ☐ |
| 9. Was follow up complete, and if not, were the reasons to loss to follow up described and explored? | ☑ | ☐ | ☐ | ☐ |
| 10. Were strategies to address incomplete follow up utilized? | ☑ | ☐ | ☐ | ☐ |
| 11. Was appropriate statistical analysis used? | ☑ | ☐ | ☐ | ☐ |

Overall appraisal: Include ☐ Exclude ☐ Seek further info ☑

Comments (Including reason for exclusion)

No estoy seguro de la inclusión o exclusión debido a que maneja únicamente datos de CT a lo largo se 4 semanas despues de inicio de los sintomas, pero no menciona síntomas persistentes

© JBI, 2020. All rights reserved. JBI grants use of these tools for research purposes only. All other enquiries should be sent to.

### JBI CRITICAL APPRAISAL CHECKLIST FOR QUALITATIVE RESEARCH

Reviewer Isaac David Juárez Cruz Date 25/11/20

Author Pratas Diogo, Silva Jorge Year 2020 Record Number 78_32730589_IDJC

| | Yes | No | Unclear | Not applicable |
|---|---|---|---|---|
| 1. Is there congruity between the stated philosophical perspective and the research methodology? | ☑ | ☐ | ☐ | ☐ |
| 2. Is there congruity between the research methodology and the research question or objectives? | ☑ | ☐ | ☐ | ☐ |
| 3. Is there congruity between the research methodology and the methods used to collect data? | ☑ | ☐ | ☐ | ☐ |
| 4. Is there congruity between the research methodology and the representation and analysis of data? | ☑ | ☐ | ☐ | ☐ |
| 5. Is there congruity between the research methodology and the interpretation of results? | ☑ | ☐ | ☐ | ☐ |
| 6. Is there a statement locating the researcher culturally or theoretically? | ☐ | ☐ | ☑ | ☐ |
| 7. Is the influence of the researcher on the research, and vice- versa, addressed? | ☐ | ☐ | ☑ | ☐ |
| 8. Are participants, and their voices, adequately represented? | ☐ | ☐ | ☐ | ☑ |
| 9. Is the research ethical according to current criteria or, for recent studies, and is there evidence of ethical approval by an appropriate body? | ☐ | ☐ | ☑ | ☐ |
| 10. Do the conclusions drawn in the research report flow from the analysis, or interpretation, of the data? | ☑ | ☐ | ☐ | ☐ |

Overall appraisal: Include ☐ Exclude ☑ Seek further info ☐

Comments (Including reason for exclusion)

Se excluye, debido a que no propociona información que empate xon lo que buscamos
Se centra en el reconocimiento de secuencias de ADN, de proteinas y como esto puede auxiliar en el desarrollo de tratamientos y en las pruebas RT-PCR, entre otras cosas

© JBI, 2020. All rights reserved. JBI grants use of these tools for research purposes only. All other enquiries should be sent to.

### JBI CRITICAL APPRAISAL CHECKLIST FOR ANALYTICAL CROSS SECTIONAL STUDIES

Reviewer Daniel Eduardo Sandoval Colin Date 31/03/2021

Author Stephen J Halpin, Claire McIvor, Gemma Whyatt et al. Year 2020 Record Number 79_32729939_DESC

| | Yes | No | Unclear | Not applicable |
|---|---|---|---|---|
| 1. Were the criteria for inclusion in the sample clearly defined? | X | | | |
| 2. Were the study subjects and the setting described in detail? | X | | | |
| 3. Was the exposure measured in a valid and reliable way? | X | | | |
| 4. Were objective, standard criteria used for measurement of the condition? | X | | | |
| 5. Were confounding factors identified? | X | | | |
| 6. Were strategies to deal with confounding factors stated? | | X | | |
| 7. Were the outcomes measured in a valid and reliable way? | X | | | |
| 8. Was appropriate statistical analysis used? | | X | | |

Overall appraisal: Include [X] Exclude [ ] Seek further info [ ]

Comments (Including reason for exclusion)

### JBI Critical Appraisal Checklist for Case Series

Reviewer Daniel Eduardo Sandoval Colin Date 31/03/2021

Author E Guedj, M Million, P Dudouet et al. Year 2020 Record Number 80_32728799_DESC

| | Yes | No | Unclear | Not applicable |
|---|---|---|---|---|
| • Were there clear criteria for inclusion in the case series? | ☐ | ☐ | ☐ | ☐ |
| • Was the condition measured in a standard, reliable way for all participants included in the case series? | ☐ | ☐ | ☐ | ☐ |
| • Were valid methods used for identification of the condition for all participants included in the case series? | ☐ | ☐ | ☐ | ☐ |
| • Did the case series have consecutive inclusion of participants? | ☐ | ☐ | ☐ | ☐ |
| • Did the case series have complete inclusion of participants? | ☐ | ☐ | ☐ | ☐ |
| • Was there clear reporting of the demographics of the participants in the study? | ☐ | ☐ | ☐ | ☐ |
| • Was there clear reporting of clinical information of the participants? | ☐ | ☐ | ☐ | ☐ |
| • Were the outcomes or follow up results of cases clearly reported? | ☐ | ☐ | ☐ | ☐ |
| • Was there clear reporting of the presenting site(s)/clinic(s) demographic information? | ☐ | ☐ | ☐ | ☐ |
| • Was statistical analysis appropriate? | ☐ | ☐ | ☐ | ☐ |

Overall appraisal: Include ☐ Exclude ☒ Seek further info ☐

Comments (Including reason for exclusion)

No se realiza cuestionario porque no cumple con los criterios de inclusión.

Reporte de hipometabolismo en áreas cerebrales por F-FDG PET de 2 casos en etapa postviral por COVID-19

### JBI CRITICAL APPRAISAL CHECKLIST FOR ANALYTICAL CROSS SECTIONAL STUDIES

Reviewer Carlos Alberto Fermin Martínez Date 02/04/2021

Author Barón-Sánchez, et al. Year 2020 Record Number 81_32900532_CAFM

| | Yes | No | Unclear | Not applicable |
|---|---|---|---|---|
| 1. Were the criteria for inclusion in the sample clearly defined? | ☒ | ☐ | ☐ | ☐ |
| 2. Were the study subjects and the setting described in detail? | ☒ | ☐ | ☐ | ☐ |
| 3. Was the exposure measured in a valid and reliable way? | ☐ | ☐ | ☒ | ☐ |
| 4. Were objective, standard criteria used for measurement of the condition? | ☒ | ☐ | ☐ | ☐ |
| 5. Were confounding factors identified? | ☐ | ☒ | ☐ | ☐ |
| 6. Were strategies to deal with confounding factors stated? | ☐ | ☒ | ☐ | ☐ |
| 7. Were the outcomes measured in a valid and reliable way? | ☒ | ☐ | ☐ | ☐ |
| 8. Was appropriate statistical analysis used? | ☒ | ☐ | ☐ | ☐ |

Overall appraisal: Include ☒ Exclude ☐ Seek further info ☐

Comments (Including reason for exclusion)

No hay información sobre la duración de los síntomas además de anosmia e hiposmia. Se describen los resultados de pacientes con "PCR positiva" pero esta información se obtuvo mediante una encuesta en línea o telefónica.

© JBI, 2020. All rights reserved. JBI grants use of these tools for research purposes only. All other enquiries should be sent to.

### JBI CRITICAL APPRAISAL CHECKLIST FOR COHORT STUDIES

Reviewer: Isaac David Juárez Cruz  Date: 28/11/2020

Author: *  Year: 2020  Record Number: 82_32725271_IDJC

| | Yes | No | Unclear | Not applicable |
|---|---|---|---|---|
| 1. Were the two groups similar and recruited from the same population? | ☑ | ☐ | ☐ | ☐ |
| 2. Were the exposures measured similarly to assign people to both exposed and unexposed groups? | ☑ | ☐ | ☐ | ☐ |
| 3. Was the exposure measured in a valid and reliable way? | ☑ | ☐ | ☐ | ☐ |
| 4. Were confounding factors identified? | ☐ | ☐ | ☑ | ☐ |
| 5. Were strategies to deal with confounding factors stated? | ☐ | ☐ | ☑ | ☐ |
| 6. Were the groups/participants free of the outcome at the start of the study (or at the moment of exposure)? | ☐ | ☑ | ☐ | ☐ |
| 7. Were the outcomes measured in a valid and reliable way? | ☑ | ☐ | ☐ | ☐ |
| 8. Was the follow up time reported and sufficient to be long enough for outcomes to occur? | ☑ | ☐ | ☐ | ☐ |
| 9. Was follow up complete, and if not, were the reasons to loss to follow up described and explored? | ☑ | ☐ | ☐ | ☐ |
| 10. Were strategies to address incomplete follow up utilized? | ☐ | ☐ | ☐ | ☑ |
| 11. Was appropriate statistical analysis used? | ☑ | ☐ | ☐ | ☐ |

Overall appraisal: Include ☑ Exclude ☐ Seek further info ☐

Comments (Including reason for exclusion)

Estudio de HCW, mencionado que las OTD son prevalentes en este personal, de igual forma desglosa en que esoecialidades medicas es más común encontrar estos desordenes.

© JBI, 2020. All rights reserved. JBI grants use of these tools for research purposes only. All other enquiries should be sent to.

*

Ithzel Maria Villarreal, Marta Morato,
Mar Martínez-RuizCoello, Andrés Navarro,
Raimon Garcia-Chillerón, Álvaro Ruiz,
Isabella Vacallanos de Almeida, Luis Mazón,
Guillermo Plaza

### JBI CRITICAL APPRAISAL CHECKLIST FOR COHORT STUDIES

Reviewer Carlos Alberto Fermin Martinez Date 02/04/2021

Author Xu K, et al. Year 2020 Record Number 83_32271376_CAFM

| | Yes | No | Unclear | Not applicable |
|---|---|---|---|---|
| 1. Were the two groups similar and recruited from the same population? | ☑ | ☐ | ☐ | ☐ |
| 2. Were the exposures measured similarly to assign people<br>3. to both exposed and unexposed groups? | ☑ | ☐ | ☐ | ☐ |
| 4. Was the exposure measured in a valid and reliable way? | ☑ | ☐ | ☐ | ☐ |
| 5. Were confounding factors identified? | ☑ | ☐ | ☐ | ☐ |
| 6. Were strategies to deal with confounding factors stated? | ☑ | ☐ | ☐ | ☐ |
| 7. Were the groups/participants free of the outcome at the start of the study (or at the moment of exposure)? | ☐ | ☐ | ☐ | ☑ |
| 8. Were the outcomes measured in a valid and reliable way? | ☑ | ☐ | ☐ | ☐ |
| 9. Was the follow up time reported and sufficient to be long enough for outcomes to occur? | ☑ | ☐ | ☐ | ☐ |
| 10. Was follow up complete, and if not, were the reasons to loss to follow up described and explored? | ☑ | ☐ | ☐ | ☐ |
| 11. Were strategies to address incomplete follow up utilized? | ☐ | ☑ | ☐ | ☐ |
| 12. Was appropriate statistical analysis used? | ☑ | ☐ | ☐ | ☐ |

Overall appraisal: Include ☐ Exclude ☑ Seek further info ☐

Comments (Including reason for exclusion)

Aunque el artículo tiene una metodología robusta y detallada, la información que provee no entra en el alcance de nuestro estudio.

© JBI, 2020. All rights reserved. JBI grants use of these tools for research purposes only. All other enquiries should be sent to.

### JBI CRITICAL APPRAISAL CHECKLIST FOR SYSTEMATIC REVIEWS AND RESEARCH SYNTHESES

Reviewer Daniel de la Rosa Date 01-02-21

Author Maria Gavriatopoulou et al Year 2020 Record Number 84_32720223

Review

| | Yes | No | Unclear | Not applicable |
|---|---|---|---|---|
| 1. Is the review question clearly and explicitly stated? | X | | | |
| 2. Were the inclusion criteria appropriate for the review question? | | | | X |
| 3. Was the search strategy appropriate? | | | | X |
| 4. Were the sources and resources used to search for studies adequate? | | | | X |
| 5. Were the criteria for appraising studies appropriate? | | | | X |
| 6. Was critical appraisal conducted by two or more reviewers independently? | | | | X |
| 7. Were there methods to minimize errors in data extraction? | | | | X |
| 8. Were the methods used to combine studies appropriate? | | | | X |
| 9. Was the likelihood of publication bias assessed? | | | | X |
| 10. Were recommendations for policy and/or practice supported by the reported data? | X | | | |
| 11. Were the specific directives for new research appropriate? | X | | | |

Overall appraisal: Include ☐ Exclude ☒ Seek further info ☐

Comments (Including reason for exclusion)

### JBI CRITICAL APPRAISAL CHECKLIST FOR TEXT AND OPINION PAPERS

Reviewer: Isaac David Juárez Cruz  Date: 29/11/20

Author: Antonelli M, Donelli D.  Year: 2020  Record Number: 85_32710297_IDJC

| | Yes | No | Unclear | Not applicable |
|---|---|---|---|---|
| 1. Is the source of the opinion clearly identified? | ☑ | ☐ | ☐ | ☐ |
| 2. Does the source of opinion have standing in the field of expertise? | ☐ | ☐ | ☑ | ☐ |
| 3. Are the interests of the relevant population the central focus of the opinion? | ☑ | ☐ | ☐ | ☐ |
| 4. Is the stated position the result of an analytical process, and is there logic in the opinion expressed? | ☑ | ☐ | ☐ | ☐ |
| 5. Is there reference to the extant literature? | ☑ | ☐ | ☐ | ☐ |
| 6. Is any incongruence with the literature/sources logically defended? | ☑ | ☐ | ☐ | ☐ |

Overall appraisal: Include ☐ Exclude ☑ Seek further info ☐

Comments (Including reason for exclusion)

Excluido, solo menciona que es recomendable utilizar la rehabilitación en Spa's con tecnicas ya utlizadas anteriormente para fibrosis pulmonar entre otras. No hace mención de datos relevantes.

© JBI, 2020. All rights reserved. JBI grants use of these tools for research purposes only. All other enquiries should be sent to.

### JBI CRITICAL APPRAISAL CHECKLIST FOR COHORT STUDIES

Reviewer: Isaac David Juárez Cruz Date: 29/11/20

Author: * Year: 2020 Record Number: 86_32929402_IDJC

| | Yes | No | Unclear | Not applicable |
|---|---|---|---|---|
| 1. Were the two groups similar and recruited from the same population? | ☑ | ☐ | ☐ | ☐ |
| 2. Were the exposures measured similarly to assign people to both exposed and unexposed groups? | ☑ | ☐ | ☐ | ☐ |
| 3. Was the exposure measured in a valid and reliable way? | ☑ | ☐ | ☐ | ☐ |
| 4. Were confounding factors identified? | ☐ | ☐ | ☑ | ☐ |
| 5. Were strategies to deal with confounding factors stated? | ☐ | ☐ | ☑ | ☐ |
| 6. Were the groups/participants free of the outcome at the start of the study (or at the moment of exposure)? | ☐ | ☐ | ☐ | ☑ |
| 7. Were the outcomes measured in a valid and reliable way? | ☑ | ☐ | ☐ | ☐ |
| 8. Was the follow up time reported and sufficient to be long enough for outcomes to occur? | ☑ | ☐ | ☐ | ☐ |
| 9. Was follow up complete, and if not, were the reasons to loss to follow up described and explored? | ☑ | ☐ | ☐ | ☐ |
| 10. Were strategies to address incomplete follow up utilized? | ☐ | ☐ | ☐ | ☑ |
| 11. Was appropriate statistical analysis used? | ☑ | ☐ | ☐ | ☐ |

Overall appraisal: Include ☐ Exclude ☑ Seek further info ☐

Comments (Including reason for exclusion): Unicamente hace alusión a si ntomas agudos, menciona que es recomendable el seguimiento de pacientes que han sido dados de alta, sin embargo no habla más de ello. Se mencionan las medidas que deberian de tomar los pacientes que han sido dads de alta para evitar el contagio debido a que aun son portadores del virus.

© JBI, 2020. All rights reserved. JBI grants use of these tools for research purposes only. All other enquiries should be sent to.

*

Lu Xia, Jun Chen, Thomas Friedemann, Zongguo Yang, Yun Ling, Xuhui Liu, Shuihua Lu, Tao Li, Zhigang Song, Wei Huang, Yunfei Lu, Sven Schröder, and Hongzhou Lu

### JBI CRITICAL APPRAISAL CHECKLIST FOR COHORT STUDIES

Reviewer Isaac David Juárez Cruz Date 29/11/20

Author Davido B, Seang S, Tubiana R, Truchis P. Year 2020 Record Number 87_32712242_IDJC

| | Yes | No | Unclear | Not applicable |
|---|---|---|---|---|
| 1. Were the two groups similar and recruited from the same population? | ✓ | | | |
| 2. Were the exposures measured similarly to assign people to both exposed and unexposed groups? | | | ✓ | |
| 3. Was the exposure measured in a valid and reliable way? | | | ✓ | |
| 4. Were confounding factors identified? | | | ✓ | |
| 5. Were strategies to deal with confounding factors stated? | | | | ✓ |
| 6. Were the groups/participants free of the outcome at the start of the study (or at the moment of exposure)? | ✓ | | | |
| 7. Were the outcomes measured in a valid and reliable way? | ✓ | | | |
| 8. Was the follow up time reported and sufficient to be long enough for outcomes to occur? | ✓ | | | |
| 9. Was follow up complete, and if not, were the reasons to loss to follow up described and explored? | | | ✓ | |
| 10. Were strategies to address incomplete follow up utilized? | | | ✓ | |
| 11. Was appropriate statistical analysis used? | | | ✓ | |

Overall appraisal: Include ☑ Exclude ☐ Seek further info ☐

Comments (Including reason for exclusion)

© JBI, 2020. All rights reserved. JBI grants use of these tools for research purposes only. All other enquiries should be sent to.

### JBI CRITICAL APPRAISAL CHECKLIST FOR SYSTEMATIC REVIEWS AND RESEARCH SYNTHESES

Reviewer: Isaac David Juárez Cruz  Date: 10/12/20

Author: Weiss A, Jellingsø, Morten S.  Year: 2020  Record Number: 88_32711256_IDJC

| | Yes | No | Unclear | Not applicable |
|---|---|---|---|---|
| 1. Is the review question clearly and explicitly stated? | ☑ | ☐ | ☐ | ☐ |
| 2. Were the inclusion criteria appropriate for the review question? | ☑ | ☐ | ☐ | ☐ |
| 3. Was the search strategy appropriate? | ☑ | ☐ | ☐ | ☐ |
| 4. Were the sources and resources used to search for studies adequate? | ☑ | ☐ | ☐ | ☐ |
| 5. Were the criteria for appraising studies appropriate? | ☑ | ☐ | ☐ | ☐ |
| 6. Was critical appraisal conducted by two or more reviewers independently? | ☐ | ☐ | ☑ | ☐ |
| 7. Were there methods to minimize errors in data extraction? | ☑ | ☐ | ☐ | ☐ |
| 8. Were the methods used to combine studies appropriate? | ☑ | ☐ | ☐ | ☐ |
| 9. Was the likelihood of publication bias assessed? | ☐ | ☐ | ☑ | ☐ |
| 10. Were recommendations for policy and/or practice supported by the reported data? | ☑ | ☐ | ☐ | ☐ |
| 11. Were the specific directives for new research appropriate? | ☑ | ☐ | ☐ | ☐ |

Overall appraisal: Include ☐ Exclude ☑ Seek further info ☐

Comments (Including reason for exclusion)

No aporta información o datos relevantes de la fase post aguda de la enfermedad, hace un analisis muy detallado unicamente de la fase aguda.

© JBI, 2020. All rights reserved. JBI grants use of these tools for research purposes only. All other enquiries should be sent to.

### JBI CRITICAL APPRAISAL CHECKLIST FOR TEXT AND OPINION PAPERS

Reviewer Isaac David Juárez Cruz Date 30/11/20

Author David H & Rosalina G Year 2020 Record Number 89_32700223_IDJC

| | Yes | No | Unclear | Not applicable |
|---|---|---|---|---|
| 1. Is the source of the opinion clearly identified? | ☐ | ☐ | ☐ | ☑ |
| 2. Does the source of opinion have standing in the field of expertise? | ☐ | ☐ | ☐ | ☑ |
| 3. Are the interests of the relevant population the central focus of the opinion? | ☑ | ☐ | ☐ | ☐ |
| 4. Is the stated position the result of an analytical process, and is there logic in the opinion expressed? | ☑ | ☐ | ☐ | ☐ |
| 5. Is there reference to the extant literature? | ☑ | ☐ | ☐ | ☐ |
| 6. Is any incongruence with the literature/sources logically defended? | ☐ | ☐ | ☐ | ☑ |

Overall appraisal: Include ☑ Exclude ☐ Seek further info ☐

Comments (Including reason for exclusion)

© JBI, 2020. All rights reserved. JBI grants use of these tools for research purposes only. All other enquiries should be sent to.

### JBI CRITICAL APPRAISAL CHECKLIST FOR CASE REPORTS

Reviewer Isaac David Juárez Cruz Date 30/11/20

Author Soon Tjin Lim, Benjamin Janaway, Harry Costello, Anand Trip and Gary Price Year 2020 Record Number 90_3269735_IDJC

| | Yes | No | Unclear | Not applicable |
|---|---|---|---|---|
| 1. Were patient's demographic characteristics clearly described? | ☑ | ☐ | ☐ | ☐ |
| 2. Was the patient's history clearly described and presented as a timeline? | ☑ | ☐ | ☐ | ☐ |
| 3. Was the current clinical condition of the patient on presentation clearly described? | ☑ | ☐ | ☐ | ☐ |
| 4. Were diagnostic tests or assessment methods and the results clearly described? | ☑ | ☐ | ☐ | ☐ |
| 5. Was the intervention(s) or treatment procedure(s) clearly described? | ☑ | ☐ | ☐ | ☐ |
| 6. Was the post-intervention clinical condition clearly described? | ☑ | ☐ | ☐ | ☐ |
| 7. Were adverse events (harms) or unanticipated events identified and described? | ☐ | ☐ | ☑ | ☐ |
| 8. Does the case report provide takeaway lessons? | ☑ | ☐ | ☐ | ☐ |

Overall appraisal: Include ☐ Exclude ☑ Seek further info ☐

Comments (Including reason for exclusion)

© JBI, 2020. All rights reserved. JBI grants use of these tools for research purposes only. All other enquiries should be sent to.

### JBI CRITICAL APPRAISAL CHECKLIST FOR COHORT STUDIES

Reviewer ALEJANDRO MÁRQUEZ SALINAS Date 31/03/2021

Author Querini-Rovere P, et al. Year 2020 Record Number 91_32701913_AMS

| | Yes | No | Unclear | Not applicable |
|---|---|---|---|---|
| 1. Were the two groups similar and recruited from the same population? | ☐ | ☐ | ☐ | ☑ |
| 2. Were the exposures measured similarly to assign people<br>3. to both exposed and unexposed groups? | ☐ | ☐ | ☐ | ☑ |
| 4. Was the exposure measured in a valid and reliable way? | ☐ | ☐ | ☑ | ☐ |
| 5. Were confounding factors identified? | ☑ | ☐ | ☐ | ☐ |
| 6. Were strategies to deal with confounding factors stated? | ☐ | ☐ | ☑ | ☐ |
| 7. Were the groups/participants free of the outcome at the start of the study (or at the moment of exposure)? | ☐ | ☐ | ☑ | ☐ |
| 8. Were the outcomes measured in a valid and reliable way? | ☐ | ☐ | ☐ | ☑ |
| 9. Was the follow up time reported and sufficient to be long enough for outcomes to occur? | ☑ | ☐ | ☐ | ☐ |
| 10. Was follow up complete, and if not, were the reasons to loss to follow up described and explored? | ☐ | ☐ | ☑ | ☐ |
| 11. Were strategies to address incomplete follow up utilized? | ☐ | ☐ | ☑ | ☐ |
| 12. Was appropriate statistical analysis used? | ☐ | ☐ | ☑ | ☐ |

Overall appraisal: Include ☐ Exclude ☐ Seek further info ☑

Comments (Including reason for exclusion)

Esta articulo solamente describe brevemente a la cohorte que fue seguida sin presentar resultados relevantes para nuestro análisis, por lo que se excluye del estudio. Podría valer la pena revisar si se ha publicado información adicional utilizando esta cohorte de pacientes.

© JBI, 2020. All rights reserved. JBI grants use of these tools for research purposes only. All other enquiries should be sent to.

### JBI CRITICAL APPRAISAL CHECKLIST FOR TEXT AND OPINION PAPERS

Reviewer Daniel de la Rosa Date 02-02-21

Author Mary Daval et al Year 2020 Record Number 92_32690074

| | Yes | No | Unclear | Not applicable |
|---|---|---|---|---|
| 1. Is the source of the opinion clearly identified? | ☐ | ☐ | ☐ | ☒ |
| 2. Does the source of opinion have standing in the field of expertise? | ☐ | ☐ | ☐ | ☒ |
| 3. Are the interests of the relevant population the central focus of the opinion? | ☐ | ☐ | ☐ | ☒ |
| 4. Is the stated position the result of an analytical process, and is there logic in the opinion expressed? | ☒ | ☐ | ☐ | ☐ |
| 5. Is there reference to the extant literature? | ☐ | ☐ | ☐ | ☒ |
| 6. Is any incongruence with the literature/sources logically defended? | ☐ | ☒ | ☐ | ☐ |

Overall appraisal: Include ☐ Exclude ☒ Seek further info ☐

Comments (Including reason for exclusion)

© JBI, 2020. All rights reserved. JBI grants use of these tools for research purposes only. All other enquiries should be sent to.

### JBI CRITICAL APPRAISAL CHECKLIST FOR COHORT STUDIES

Reviewer ALEJANDRO MÁRQUEZ SALINAS Date 31/03/2021

Author Chiesa-Estomba CM, et al Year 2020 Record Number 93_32677329_AMS

| | Yes | No | Unclear | Not applicable |
|---|---|---|---|---|
| 1. Were the two groups similar and recruited from the same population? | ☐ | ☐ | ☑ | ☐ |
| 2. Were the exposures measured similarly to assign people<br>3. to both exposed and unexposed groups? | ☐ | ☐ | ☑ | ☐ |
| 4. Was the exposure measured in a valid and reliable way? | ☑ | ☐ | ☐ | ☐ |
| 5. Were confounding factors identified? | ☐ | ☑ | ☐ | ☐ |
| 6. Were strategies to deal with confounding factors stated? | ☐ | ☑ | ☐ | ☐ |
| 7. Were the groups/participants free of the outcome at the start of the study (or at the moment of exposure)? | ☑ | ☐ | ☐ | ☐ |
| 8. Were the outcomes measured in a valid and reliable way? | ☑ | ☐ | ☐ | ☐ |
| 9. Was the follow up time reported and sufficient to be long enough for outcomes to occur? | ☑ | ☐ | ☐ | ☐ |
| 10. Was follow up complete, and if not, were the reasons to loss to follow up described and explored? | ☐ | ☑ | ☐ | ☐ |
| 11. Were strategies to address incomplete follow up utilized? | ☐ | ☑ | ☐ | ☐ |
| 12. Was appropriate statistical analysis used? | ☐ | ☑ | ☐ | ☐ |

Overall appraisal: Include ☐ Exclude ☑ Seek further info ☐

Comments (Including reason for exclusion)

En la metodología del estudio se dice que se incluyeron pacientes con PCR o con serología positiva, pero no se muestra información desagregada al respecto. Además, no todos los pacientes tuvieron el mismo tiempo de seguimiento (rango de seguimiento de 30 a 71 días), lo cual complica la evaluación de persistencia de síntomas. Por último, la descripción y presentación del análisis estadístico es confusa.

© JBI, 2020. All rights reserved. JBI grants use of these tools for research purposes only. All other enquiries should be sent to.

### JBI CRITICAL APPRAISAL CHECKLIST FOR COHORT STUDIES

Reviewer ALEJANDRO MÁRQUEZ SALINAS Date 31/03/2021

Author Zhao Y, et al. Year 2020 Record Number 94_32838236_AMS

| | Yes | No | Unclear | Not applicable |
|---|---|---|---|---|
| 1. Were the two groups similar and recruited from the same population? | ☑ | ☐ | ☐ | ☐ |
| 2. Were the exposures measured similarly to assign people<br>3. to both exposed and unexposed groups? | ☑ | ☐ | ☐ | ☐ |
| 4. Was the exposure measured in a valid and reliable way? | ☑ | ☐ | ☐ | ☐ |
| 5. Were confounding factors identified? | ☑ | ☐ | ☐ | ☐ |
| 6. Were strategies to deal with confounding factors stated? | ☑ | ☐ | ☐ | ☐ |
| 7. Were the groups/participants free of the outcome at the start of the study (or at the moment of exposure)? | ☑ | ☐ | ☐ | ☐ |
| 8. Were the outcomes measured in a valid and reliable way? | ☑ | ☐ | ☐ | ☐ |
| 9. Was the follow up time reported and sufficient to be long enough for outcomes to occur? | ☑ | ☐ | ☐ | ☐ |
| 10. Was follow up complete, and if not, were the reasons to loss to follow up described and explored? | ☑ | ☐ | ☐ | ☐ |
| 11. Were strategies to address incomplete follow up utilized? | ☑ | ☐ | ☐ | ☐ |
| 12. Was appropriate statistical analysis used? | ☑ | ☐ | ☐ | ☐ |

Overall appraisal: Include ☑ Exclude ☐ Seek further info ☐

Comments (Including reason for exclusion)

Aunque no se analizan los factores de riesgo determinantes de persistencia de síntomas, sí se reporta la prevalencia de algunos síntomas persistentes. Además, se analizan los factores asociados con persistencia en la afección tomográfica y de la función pulmonar.

© JBI, 2020. All rights reserved. JBI grants use of these tools for research purposes only. All other enquiries should be sent to.

### JBI CRITICAL APPRAISAL CHECKLIST FOR TEXT AND OPINION PAPERS

Reviewer Alejandro Sicilia Andrade Date 30/04/2021

Author Megan M. Hosey, Dale M Needham Year 2020 Record Number 95_32569623_ASA

| | Yes | No | Unclear | Not applicable |
|---|---|---|---|---|
| 1. Is the source of the opinion clearly identified? | ☑ | ☐ | ☐ | ☐ |
| 2. Does the source of opinion have standing in the field of expertise? | ☑ | ☐ | ☐ | ☐ |
| 3. Are the interests of the relevant population the central focus of the opinion? | ☑ | ☐ | ☐ | ☐ |
| 4. Is the stated position the result of an analytical process, and is there logic in the opinion expressed? | ☑ | ☐ | ☐ | ☐ |
| 5. Is there reference to the extant literature? | ☑ | ☐ | ☐ | ☐ |
| 6. Is any incongruence with the literature/sources logically defended? | ☑ | ☐ | ☐ | ☐ |

Overall appraisal: Include ☐ Exclude ☑ Seek further info ☐

Comments (Including reason for exclusion)

A pesar de que es un buen artículo, se excluye por tipo de estudio (No se incluyen opiniones)

© JBI, 2020. All rights reserved. JBI grants use of these tools for research purposes only. All other enquiries should be sent to.

### JBI CRITICAL APPRAISAL CHECKLIST FOR CASE CONTROL STUDIES

Reviewer Alejandro Sicilia Andrade Date 30/04/2021

Author Zhao Fu, Ningning Tang et al. Year 2020 Record Number 96_32665630_ASA

| | Yes | No | Unclear | Not applicable |
|---|---|---|---|---|
| 1. Were the groups comparable other than the presence of disease in cases or the absence of disease in controls? | ☑ | ☐ | ☐ | ☐ |
| 2. Were cases and controls matched appropriately? | ☑ | ☐ | ☐ | ☐ |
| 3. Were the same criteria used for identification of cases and controls? | ☑ | ☐ | ☐ | ☐ |
| 4. Was exposure measured in a standard, valid and reliable way? | ☑ | ☐ | ☐ | ☐ |
| 5. Was exposure measured in the same way for cases and controls? | ☑ | ☐ | ☐ | ☐ |
| 6. Were confounding factors identified? | ☐ | ☑ | ☐ | ☐ |
| 7. Were strategies to deal with confounding factors stated? | ☐ | ☑ | ☐ | ☐ |
| 8. Were outcomes assessed in a standard, valid and reliable way for cases and controls? | ☑ | ☐ | ☐ | ☐ |
| 9. Was the exposure period of interest long enough to be meaningful? | ☑ | ☐ | ☐ | ☐ |
| 10. Was appropriate statistical analysis used? | ☑ | ☐ | ☐ | ☐ |

Overall appraisal: Include ☐ Exclude ☑ Seek further info ☐

Comments (Including reason for exclusion)

It does not report the follow-up time

© JBI, 2020. All rights reserved. JBI grants use of these tools for research purposes only. All other enquiries should be sent to.

### JBI CRITICAL APPRAISAL CHECKLIST FOR COHORT STUDIES

Reviewer Alejandro Sicilia Andrade Date 30 / 04 / 2021

Author Pellaud Charlotte, Grandmaison Gaël , et al. Year 2020 Record Number 97_32902869_ASA

| | Yes | No | Unclear | Not applicable |
|---|---|---|---|---|
| 1. Were the two groups similar and recruited from the same population? | ☑ | ☐ | ☐ | ☐ |
| 2. Were the exposures measured similarly to assign people<br>3. to both exposed and unexposed groups? | ☑ | ☐ | ☐ | ☐ |
| 4. Was the exposure measured in a valid and reliable way? | ☑ | ☐ | ☐ | ☐ |
| 5. Were confounding factors identified? | ☑ | ☐ | ☐ | ☐ |
| 6. Were strategies to deal with confounding factors stated? | ☑ | ☐ | ☐ | ☐ |
| 7. Were the groups/participants free of the outcome at the start of the study (or at the moment of exposure)? | ☑ | ☐ | ☐ | ☐ |
| 8. Were the outcomes measured in a valid and reliable way? | ☑ | ☐ | ☐ | ☐ |
| 9. Was the follow up time reported and sufficient to be long enough for outcomes to occur? | ☑ | ☐ | ☐ | ☐ |
| 10. Was follow up complete, and if not, were the reasons to loss to follow up described and explored? | ☑ | ☐ | ☐ | ☐ |
| 11. Were strategies to address incomplete follow up utilized? | ☑ | ☐ | ☐ | ☐ |
| 12. Was appropriate statistical analysis used? | ☐ | ☑ | ☐ | ☐ |

Overall appraisal: Include ☑ Exclude ☐ Seek further info ☐

Comments (Including reason for exclusion)

© JBI, 2020. All rights reserved. JBI grants use of these tools for research purposes only. All other enquiries should be sent to.

### JBI CRITICAL APPRAISAL CHECKLIST FOR CASE CONTROL STUDIES

Reviewer Isaac David Juárez Cruz Date 17/12/20

Author * Year 2020 Record Number 98_32662745_IDJC

| | Yes | No | Unclear | Not applicable |
|---|---|---|---|---|
| 1. Were the groups comparable other than the presence of disease in cases or the absence of disease in controls? | ✓ | ☐ | ☐ | ☐ |
| 2. Were cases and controls matched appropriately? | ☐ | ☐ | ✓ | ☐ |
| 3. Were the same criteria used for identification of cases and controls? | ☐ | ✓ | ☐ | ☐ |
| 4. Was exposure measured in a standard, valid and reliable way? | ✓ | ☐ | ☐ | ☐ |
| 5. Was exposure measured in the same way for cases and controls? | ✓ | ☐ | ☐ | ☐ |
| 6. Were confounding factors identified? | ☐ | ✓ | ☐ | ☐ |
| 7. Were strategies to deal with confounding factors stated? | ☐ | ☐ | ☐ | ✓ |
| 8. Were outcomes assessed in a standard, valid and reliable way for cases and controls? | ✓ | ☐ | ☐ | ☐ |
| 9. Was the exposure period of interest long enough to be meaningful? | ✓ | ☐ | ☐ | ☐ |
| 10. Was appropriate statistical analysis used? | ✓ | ☐ | ☐ | ☐ |

Overall appraisal: Include ☐ Exclude ✓ Seek further info ☐

Comments (Including reason for exclusion)

© JBI, 2020. All rights reserved. JBI grants use of these tools for research purposes only. All other enquiries should be sent to.

*

Luca D'Ascanio, Manlio Pandolfini, Cristina Cingolani, Gino Latini, Paolo Gradoni, Maria Capalbo, Gabriele Frausini, Massimo Maranzano, Michael J. Brenner, and Arianna Di Stadio.

### JBI Critical Appraisal Checklist for Case Series

Reviewer Alejandro Sicilia Andrade Date 30/04/2021

Author Abdoulaye Keita, Hamza Abdou, Ibrahima Diallo, Alseny Camara Year 2020 Record Number 99_32952820_ASA

| | Yes | No | Unclear | Not applicable |
|---|---|---|---|---|
| • Were there clear criteria for inclusion in the case series? | ☑ | ☐ | ☐ | ☐ |
| • Was the condition measured in a standard, reliable way for all participants included in the case series? | ☑ | ☐ | ☐ | ☐ |
| • Were valid methods used for identification of the condition for all participants included in the case series? | ☑ | ☐ | ☐ | ☐ |
| • Did the case series have consecutive inclusion of participants? | ☐ | ☑ | ☐ | ☐ |
| • Did the case series have complete inclusion of participants? | ☑ | ☐ | ☐ | ☐ |
| • Was there clear reporting of the demographics of the participants in the study? | ☑ | ☐ | ☐ | ☐ |
| • Was there clear reporting of clinical information of the participants? | ☑ | ☐ | ☐ | ☐ |
| • Were the outcomes or follow up results of cases clearly reported? | ☑ | ☐ | ☐ | ☐ |
| • Was there clear reporting of the presenting site(s)/clinic(s) demographic information? | ☑ | ☐ | ☐ | ☐ |
| • Was statistical analysis appropriate? | ☐ | ☐ | ☐ | ☑ |

Overall appraisal: Include ☐ Exclude ☑ Seek further info ☐

Comments (Including reason for exclusion)

Good evidence, but case series studies are not to be included according to our protocol

© JBI, 2020. All rights reserved. JBI grants use of these tools for research purposes only. All other enquiries should be sent to.

### JBI Critical Appraisal Checklist for Case Series

Reviewer Alejandro Sicilia Andrade Date 30/04/2021

Author Lan Lin, Shanshan-Luo, Renjo Qin, Mengling Yang, Xiaobei Wang, et al. Year 2020 Record Number 100_32659373_ASA

| | Yes | No | Unclear | Not applicable |
|---|---|---|---|---|
| • Were there clear criteria for inclusion in the case series? | ☑ | ☐ | ☐ | ☐ |
| • Was the condition measured in a standard, reliable way for all participants included in the case series? | ☑ | ☐ | ☐ | ☐ |
| • Were valid methods used for identification of the condition for all participants included in the case series? | ☑ | ☐ | ☐ | ☐ |
| • Did the case series have consecutive inclusion of participants? | ☐ | ☑ | ☐ | ☐ |
| • Did the case series have complete inclusion of participants? | ☑ | ☐ | ☐ | ☐ |
| • Was there clear reporting of the demographics of the participants in the study? | ☑ | ☐ | ☐ | ☐ |
| • Was there clear reporting of clinical information of the participants? | ☐ | ☐ | ☑ | ☐ |
| • Were the outcomes or follow up results of cases clearly reported? | ☑ | ☐ | ☐ | ☐ |
| • Was there clear reporting of the presenting site(s)/clinic(s) demographic information? | ☑ | ☐ | ☐ | ☐ |
| • Was statistical analysis appropriate? | ☑ | ☐ | ☐ | ☐ |

Overall appraisal: Include ☐ Exclude ☑ Seek further info ☐

Comments (Including reason for exclusion)

Quality of the study is good, but case series are not to be included in our study

© JBI, 2020. All rights reserved. JBI grants use of these tools for research purposes only. All other enquiries should be sent to.

### JBI CRITICAL APPRAISAL CHECKLIST FOR CASE REPORTS

Reviewer: Alejandro Sicilia Andrade  Date: 30/04/21

Author: Alberto Aiolfi, Barbara Bruni, Tullio Biraghi, Andrea Montisci, et al.  Year: 2020  Record Number: 101_32664116_ASA

| | Yes | No | Unclear | Not applicable |
|---|---|---|---|---|
| 1. Were patient's demographic characteristics clearly described? | ☑ | ☐ | ☐ | ☐ |
| 2. Was the patient's history clearly described and presented as a timeline? | ☑ | ☐ | ☐ | ☐ |
| 3. Was the current clinical condition of the patient on presentation clearly described? | ☑ | ☐ | ☐ | ☐ |
| 4. Were diagnostic tests or assessment methods and the results clearly described? | ☑ | ☐ | ☐ | ☐ |
| 5. Was the intervention(s) or treatment procedure(s) clearly described? | ☑ | ☐ | ☐ | ☐ |
| 6. Was the post-intervention clinical condition clearly described? | ☑ | ☐ | ☐ | ☐ |
| 7. Were adverse events (harms) or unanticipated events identified and described? | ☐ | ☐ | ☐ | ☑ |
| 8. Does the case report provide takeaway lessons? | ☑ | ☐ | ☐ | ☐ |

Overall appraisal: Include ☐ Exclude ☑ Seek further info ☐

Comments (Including reason for exclusion)

Our study does not include case reports

© JBI, 2020. All rights reserved. JBI grants use of these tools for research purposes only. All other enquiries should be sent to.

### JBI CRITICAL APPRAISAL CHECKLIST FOR TEXT AND OPINION PAPERS

Reviewer Enrique Cañedo Guerra Date 29/03/2021

Author Mouna Asly, Asmaa Hazim Year 2020 Record Number 102_32952812_ECG

| | Yes | No | Unclear | Not applicable |
|---|---|---|---|---|
| 1. Is the source of the opinion clearly identified? | ☐ | ☐ | ☐ | ☑ |
| 2. Does the source of opinion have standing in the field of expertise? | ☐ | ☐ | ☐ | ☑ |
| 3. Are the interests of the relevant population the central focus of the opinion? | ☑ | ☐ | ☐ | ☐ |
| 4. Is the stated position the result of an analytical process, and is there logic in the opinion expressed? | ☑ | ☐ | ☐ | ☐ |
| 5. Is there reference to the extant literature? | ☐ | ☐ | ☐ | ☑ |
| 6. Is any incongruence with the literature/sources logically defended? | ☐ | ☑ | ☐ | ☐ |

Overall appraisal: Include ☐ Exclude ☑ Seek further info ☐

Comments (Including reason for exclusion)

© JBI, 2020. All rights reserved. JBI grants use of these tools for research purposes only. All other enquiries should be sent to.

### JBI CRITICAL APPRAISAL CHECKLIST FOR CASE CONTROL STUDIES

Reviewer Isaac David Juárez Cruz Date 17/12/20

Author Feng Pan, Chuansheng Zheng, TianheYe, Lingli Li, Dehan Liu, Lin Li, Richard L. Hesketh & LianYang Year 2020 Record Number 103_32647307_IDJC

| | Yes | No | Unclear | Not applicable |
|---|---|---|---|---|
| 1. Were the groups comparable other than the presence of disease in cases or the absence of disease in controls? | ✓ | ☐ | ☐ | ☐ |
| 2. Were cases and controls matched appropriately? | ☐ | ☐ | ✓ | ☐ |
| 3. Were the same criteria used for identification of cases and controls? | ✓ | ☐ | ☐ | ☐ |
| 4. Was exposure measured in a standard, valid and reliable way? | ✓ | ☐ | ☐ | ☐ |
| 5. Was exposure measured in the same way for cases and controls? | ✓ | ☐ | ☐ | ☐ |
| 6. Were confounding factors identified? | ☐ | ✓ | ☐ | ☐ |
| 7. Were strategies to deal with confounding factors stated? | ☐ | ☐ | ☐ | ✓ |
| 8. Were outcomes assessed in a standard, valid and reliable way for cases and controls? | ✓ | ☐ | ☐ | ☐ |
| 9. Was the exposure period of interest long enough to be meaningful? | ✓ | ☐ | ☐ | ☐ |
| 10. Was appropriate statistical analysis used? | ✓ | ☐ | ☐ | ☐ |

Overall appraisal: Include ☐ Exclude ✓ Seek further info ☐

Comments (Including reason for exclusion)

© JBI, 2020. All rights reserved. JBI grants use of these tools for research purposes only. All other enquiries should be sent to.

### JBI CRITICAL APPRAISAL CHECKLIST FOR COHORT STUDIES

Reviewer Enrique Cañedo Guerra Date 29/03/2021

Author Tong-Zeng Li, Zhen-Huan Cao, Yu Chen, Miao-Tian Cai, Long-Yu Zhang, Hui Xu, Jia-Ying Zhang, Chun-Hua Ma, Yang Liu, Li-Juan Gao, Zhong-Hui Duan, Dan-Lei Mou, Lian-Chun Liang Year 2020 Record Number 104_32644223_ECG

| | Yes | No | Unclear | Not applicable |
|---|---|---|---|---|
| 1. Were the two groups similar and recruited from the same population? | ☑ | ☐ | ☐ | ☐ |
| 2. Were the exposures measured similarly to assign people<br>3. to both exposed and unexposed groups? | ☑ | ☐ | ☐ | ☐ |
| 4. Was the exposure measured in a valid and reliable way? | ☑ | ☐ | ☐ | ☐ |
| 5. Were confounding factors identified? | ☑ | ☐ | ☐ | ☐ |
| 6. Were strategies to deal with confounding factors stated? | ☐ | ☐ | ☑ | ☐ |
| 7. Were the groups/participants free of the outcome at the start of the study (or at the moment of exposure)? | ☐ | ☐ | ☐ | ☑ |
| 8. Were the outcomes measured in a valid and reliable way? | ☑ | ☐ | ☐ | ☐ |
| 9. Was the follow up time reported and sufficient to be long enough for outcomes to occur? | ☑ | ☐ | ☐ | ☐ |
| 10. Was follow up complete, and if not, were the reasons to loss to follow up described and explored? | ☑ | ☐ | ☐ | ☐ |
| 11. Were strategies to address incomplete follow up utilized? | ☑ | ☐ | ☐ | ☐ |
| 12. Was appropriate statistical analysis used? | ☑ | ☐ | ☐ | ☐ |

Overall appraisal: Include ☐ Exclude ☑ Seek further info ☐

Comments (Including reason for exclusion)

No hay seguimiento clínico, el objetivo del estudio es ver replicación viral

© JBI, 2020. All rights reserved. JBI grants use of these tools for research purposes only. All other enquiries should be sent to.

### JBI CRITICAL APPRAISAL CHECKLIST FOR TEXT AND OPINION PAPERS

Reviewer: Enrique Cañedo Guerra  Date: 27/03/2021

Author: Tahir Jameel , Mukhtiar Baig , Zohair J. Gazzaz  Year: 2020  Record Number: 105_32782867_ECG

| | Yes | No | Unclear | Not applicable |
|---|---|---|---|---|
| 1. Is the source of the opinion clearly identified? | ☐ | ☐ | ☐ | ☑ |
| 2. Does the source of opinion have standing in the field of expertise? | ☐ | ☐ | ☐ | ☑ |
| 3. Are the interests of the relevant population the central focus of the opinion? | ☑ | ☐ | ☐ | ☐ |
| 4. Is the stated position the result of an analytical process, and is there logic in the opinion expressed? | ☑ | ☐ | ☐ | ☐ |
| 5. Is there reference to the extant literature? | ☐ | ☐ | ☐ | ☑ |
| 6. Is any incongruence with the literature/sources logically defended? | ☐ | ☑ | ☐ | ☐ |

Overall appraisal: Include ☐ Exclude ☑ Seek further info ☐

Comments (Including reason for exclusion)

© JBI, 2020. All rights reserved. JBI grants use of these tools for research purposes only. All other enquiries should be sent to.

### JBI CRITICAL APPRAISAL CHECKLIST FOR SYSTEMATIC REVIEWS AND RESEARCH SYNTHESES

Reviewer Isaac David Juárez Cruz Date 25/12/20

Author Bell C, Crabtree S, Hall E, Sabdage S. Year 2020 Record Number 106_32837330_IDJC

| | | Yes | No | Unclear | Not applicable |
|---|---|---|---|---|---|
| 1. | Is the review question clearly and explicitly stated? | ✓ | | | |
| 2. | Were the inclusion criteria appropriate for the review question? | | | | ✓ |
| 3. | Was the search strategy appropriate? | | | | ✓ |
| 4. | Were the sources and resources used to search for studies adequate? | | | | ✓ |
| 5. | Were the criteria for appraising studies appropriate? | | | | ✓ |
| 6. | Was critical appraisal conducted by two or more reviewers independently? | | | | ✓ |
| 7. | Were there methods to minimize errors in data extraction? | | | | ✓ |
| 8. | Were the methods used to combine studies appropriate? | ✓ | | | |
| 9. | Was the likelihood of publication bias assessed? | | | ✓ | |
| 10. | Were recommendations for policy and/or practice supported by the reported data? | ✓ | | | |
| 11. | Were the specific directives for new research appropriate? | ✓ | | | |

Overall appraisal: Include ☐ Exclude ✓ Seek further info ☐

Comments (Including reason for exclusion)

© JBI, 2020. All rights reserved. JBI grants use of these tools for research purposes only. All other enquiries should be sent to.

### JBI Critical Appraisal Checklist for Case Series

Reviewer: Isaac David Juárez Cruz    Date: 03/01/21

Author: *    Year: 2020    Record Number: 107_32614442_IDJC

| | Yes | No | Unclear | Not applicable |
|---|---|---|---|---|
| Were there clear criteria for inclusion in the case series? | ✓ | ☐ | ☐ | ☐ |
| Was the condition measured in a standard, reliable way for all participants included in the case series? | ✓ | ☐ | ☐ | ☐ |
| Were valid methods used for identification of the condition for all participants included in the case series? | ✓ | ☐ | ☐ | ☐ |
| Did the case series have consecutive inclusion of participants? | ☐ | ✓ | ☐ | ☐ |
| Did the case series have complete inclusion of participants? | ☐ | ✓ | ☐ | ☐ |
| Was there clear reporting of the demographics of the participants in the study? | ✓ | ☐ | ☐ | ☐ |
| Was there clear reporting of clinical information of the participants? | ✓ | ☐ | ☐ | ☐ |
| Were the outcomes or follow up results of cases clearly reported? | ✓ | ☐ | ☐ | ☐ |
| Was there clear reporting of the presenting site(s)/clinic(s) demographic information? | ✓ | ☐ | ☐ | ☐ |
| Was statistical analysis appropriate? | ✓ | ☐ | ☐ | ☐ |

Overall appraisal: Include ✓ Exclude ☐ Seek further info ☐

Comments (Including reason for exclusion)

© JBI, 2020. All rights reserved. JBI grants use of these tools for research purposes only. All other enquiries should be sent to.

*

Paolo Boscolo-Rizzo, MD; Daniele Borsetto, MD; Cristoforo Fabbris, MD; Giacomo Spinato, MD;
Daniele Frezza, MD; Anna Menegaldo, MD; Francesca Mularoni, MD; Piergiorgio Gaudioso, MD;
Diego Cazzador, MD; Silvia Marciani, MD; Samuele Frasconi, MD; Maria Ferraro, MD; Cecilia Berro, MD; Chiara Varago, MD; Piero Nicolai, MD; Giancarlo Tirelli, MD; Maria Cristina Da Mosto, MD;
Rupert Obholzer, MA, MBBS; Roberto Rigoli, MD; Jerry Polesel, ScD; Claire Hopkins, MBBS

### JBI CRITICAL APPRAISAL CHECKLIST FOR SYSTEMATIC REVIEWS AND RESEARCH SYNTHESES

Reviewer: Enrique Cañedo Guerrax Date: 30/03/2021

Author: Luz Elena Ojeda Carmona, Mar a Del Carmen C rdoba Nieto , Alvaro Luis Diaz Alvarado Year: 2020 Record Number: 108_32618463_ECGdd d

| | | Yes | No | Unclear | Not applicable |
|---|---|---|---|---|---|
| 1. | Is the review question clearly and explicitly stated? | ☒ | ☐ | ☐ | ☐ |
| 2. | Were the inclusion criteria appropriate for the review question? | ☒ | ☐ | ☐ | ☐ |
| 3. | Was the search strategy appropriate? | ☒ | ☐ | ☐ | ☐ |
| 4. | Were the sources and resources used to search for studies adequate? | ☒ | ☐ | ☐ | ☐ |
| 5. | Were the criteria for appraising studies appropriate? | ☐ | ☐ | ☒ | ☐ |
| 6. | Was critical appraisal conducted by two or more reviewers independently? | ☐ | ☐ | ☒ | ☐ |
| 7. | Were there methods to minimize errors in data extraction? | ☐ | ☐ | ☒ | ☐ |
| 8. | Were the methods used to combine studies appropriate? | ☒ | ☐ | ☐ | ☐ |
| 9. | Was the likelihood of publication bias assessed? | ☐ | ☐ | ☒ | ☐ |
| 10. | Were recommendations for policy and/or practice supported by the reported data? | ☒ | ☐ | ☐ | ☐ |
| 11. | Were the specific directives for new research appropriate? | ☒ | ☐ | ☐ | ☐ |

Overall appraisal: Include ☐ Exclude ☒ Seek further info ☐

Comments (Including reason for exclusion)

No incluye información precisa sobre síntomas de long COVID.

© JBI, 2020. All rights reserved. JBI grants use of these tools for research purposes only. All other enquires should be sent to

### JBI CRITICAL APPRAISAL CHECKLIST FOR CASE REPORTS

Reviewer Isaac David Juárez Cruz Date 05/0121

Author * Year 2020 Record Number 109_32706216_IDJC

| | Yes | No | Unclear | Not applicable |
|---|---|---|---|---|
| 1. Were patient's demographic characteristics clearly described? | ☑ | ☐ | ☐ | ☐ |
| 2. Was the patient's history clearly described and presented as a timeline? | ☑ | ☐ | ☐ | ☐ |
| 3. Was the current clinical condition of the patient on presentation clearly described? | ☑ | ☐ | ☐ | ☐ |
| 4. Were diagnostic tests or assessment methods and the results clearly described? | ☑ | ☐ | ☐ | ☐ |
| 5. Was the intervention(s) or treatment procedure(s) clearly described? | ☑ | ☐ | ☐ | ☐ |
| 6. Was the post-intervention clinical condition clearly described? | ☑ | ☐ | ☐ | ☐ |
| 7. Were adverse events (harms) or unanticipated events identified and described? | ☐ | ☐ | ☑ | ☐ |
| 8. Does the case report provide takeaway lessons? | ☑ | ☐ | ☐ | ☐ |

Overall appraisal: Include ☐ Exclude ☑ Seek further info ☐

Comments (Including reason for exclusion)

© JBI, 2020. All rights reserved. JBI grants use of these tools for research purposes only. All other enquiries should be sent to.

### JBI CRITICAL APPRAISAL CHECKLIST FOR TEXT AND OPINION PAPERS

Reviewer Mónica Itzel Martínez Gutiérrez Date 03/04/2021

Author Perrin R, Riste L, Hann M, Walther A, Mukherjee A, Heald A. Year 2020 Record Number 110_32758891_MIMG

| | Yes | No | Unclear | Not applicable |
|---|---|---|---|---|
| 1. Is the source of the opinion clearly identified? | ☑ | ☐ | ☐ | ☐ |
| 2. Does the source of opinion have standing in the field of expertise? | ☑ | ☐ | ☐ | ☐ |
| 3. Are the interests of the relevant population the central focus of the opinion? | ☑ | ☐ | ☐ | ☐ |
| 4. Is the stated position the result of an analytical process, and is there logic in the opinion expressed? | ☑ | ☐ | ☐ | ☐ |
| 5. Is there reference to the extant literature? | ☑ | ☐ | ☐ | ☐ |
| 6. Is any incongruence with the literature/sources logically defended? | ☑ | ☐ | ☐ | ☐ |

Overall appraisal: Include ☐ Exclude ☑ Seek further info ☐

Comments (Including reason for exclusion)

Es una carta al editor, basado en un reporte de caso, que remarca la posibilidad de un sindrome post –viral por COVID-19

© JBI, 2020. All rights reserved. JBI grants use of these tools for research purposes only. All other enquiries should be sent to.

### JBI CRITICAL APPRAISAL CHECKLIST FOR SYSTEMATIC REVIEWS AND RESEARCH SYNTHESES

Reviewer: Isaac David Juárez Cruz  Date: 05/01/21

Author: Raul D. Mitrani, Nitika Dabas, Jeffrey J. Goldberger  Year: 2020  Record Number: 111_32599178_IDJC

| | Yes | No | Unclear | Not applicable |
|---|---|---|---|---|
| 1. Is the review question clearly and explicitly stated? | ✓ | ☐ | ☐ | ☐ |
| 2. Were the inclusion criteria appropriate for the review question? | ☐ | ☐ | ✓ | ☐ |
| 3. Was the search strategy appropriate? | ☐ | ☐ | ✓ | ☐ |
| 4. Were the sources and resources used to search for studies adequate? | ☐ | ☐ | ✓ | ☐ |
| 5. Were the criteria for appraising studies appropriate? | ☐ | ☐ | ✓ | ☐ |
| 6. Was critical appraisal conducted by two or more reviewers independently? | ☐ | ☐ | ✓ | ☐ |
| 7. Were there methods to minimize errors in data extraction? | ☐ | ☐ | ✓ | ☐ |
| 8. Were the methods used to combine studies appropriate? | ☐ | ☐ | ✓ | ☐ |
| 9. Was the likelihood of publication bias assessed? | ☐ | ☐ | ✓ | ☐ |
| 10. Were recommendations for policy and/or practice supported by the reported data? | ✓ | ☐ | ☐ | ☐ |
| 11. Were the specific directives for new research appropriate? | ✓ | ☐ | ☐ | ☐ |

Overall appraisal: Include ☐ Exclude ✓ Seek further info ☐

Comments (Including reason for exclusion)

© JBI, 2020. All rights reserved. JBI grants use of these tools for research purposes only. All other enquiries should be sent to.

### JBI CRITICAL APPRAISAL CHECKLIST FOR COHORT STUDIES

Reviewer Isaac David Juárez Cruz Date 09/01/2021

Author ★ Year 2020 Record Number 112_32586839_IDJC

| | Yes | No | Unclear | Not applicable |
|---|---|---|---|---|
| 1. Were the two groups similar and recruited from the same population? | ☐ | ☐ | ☐ | ☑ |
| 2. Were the exposures measured similarly to assign people to both exposed and unexposed groups? | ☐ | ☐ | ☐ | ☑ |
| 3. Was the exposure measured in a valid and reliable way? | ☑ | ☐ | ☐ | ☐ |
| 4. Were confounding factors identified? | ☐ | ☐ | ☑ | ☐ |
| 5. Were strategies to deal with confounding factors stated? | ☐ | ☐ | ☑ | ☐ |
| 6. Were the groups/participants free of the outcome at the start of the study (or at the moment of exposure)? | ☐ | ☐ | ☐ | ☑ |
| 7. Were the outcomes measured in a valid and reliable way? | ☑ | ☐ | ☐ | ☐ |
| 8. Was the follow up time reported and sufficient to be long enough for outcomes to occur? | ☑ | ☐ | ☐ | ☐ |
| 9. Was follow up complete, and if not, were the reasons to loss to follow up described and explored? | ☑ | ☐ | ☐ | ☐ |
| 10. Were strategies to address incomplete follow up utilized? | ☐ | ☐ | ☑ | ☐ |
| 11. Was appropriate statistical analysis used? | ☑ | ☐ | ☐ | ☐ |

Overall appraisal: Include ☐ Exclude ☑ Seek further info ☐

Comments (Including reason for exclusion)

© JBI, 2020. All rights reserved. JBI grants use of these tools for research purposes only. All other enquiries should be sent to.

*

Daoyuan Si, Beibei Du, Lujia Ni, Bo Yang MD, Huan Sun, Nan Jiang,
Guohui Liu, Stéphane Massé, Lina Jin, Jared Nanthakumar, Abhishek Bhaskaran,
Ping Yang, Kumaraswamy Nanthakumar

### JBI CRITICAL APPRAISAL CHECKLIST FOR SYSTEMATIC REVIEWS AND RESEARCH SYNTHESES

Reviewer: Isaac David Juárez Cruz  Date: 09/01/21

Author: *  Year: 2020  Record Number: 113_32574165_IDJC

| | Yes | No | Unclear | Not applicable |
|---|---|---|---|---|
| 1. Is the review question clearly and explicitly stated? | ☑ | ☐ | ☐ | ☐ |
| 2. Were the inclusion criteria appropriate for the review question? | ☑ | ☐ | ☐ | ☐ |
| 3. Was the search strategy appropriate? | ☑ | ☐ | ☐ | ☐ |
| 4. Were the sources and resources used to search for studies adequate? | ☑ | ☐ | ☐ | ☐ |
| 5. Were the criteria for appraising studies appropriate? | ☑ | ☐ | ☐ | ☐ |
| 6. Was critical appraisal conducted by two or more reviewers independently? | ☑ | ☐ | ☐ | ☐ |
| 7. Were there methods to minimize errors in data extraction? | ☑ | ☐ | ☐ | ☐ |
| 8. Were the methods used to combine studies appropriate? | ☑ | ☐ | ☐ | ☐ |
| 9. Was the likelihood of publication bias assessed? | ☑ | ☐ | ☐ | ☐ |
| 10. Were recommendations for policy and/or practice supported by the reported data? | ☑ | ☐ | ☐ | ☐ |
| 11. Were the specific directives for new research appropriate? | ☑ | ☐ | ☐ | ☐ |

Overall appraisal: Include ☐ Exclude ☑ Seek further info ☐

Comments (Including reason for exclusion)

No menciona proporción de algun sintoma persistente, solo menciona 1 vez que se ha detectado tos persistente pero no habla mas al respecto.

© JBI, 2020. All rights reserved. JBI grants use of these tools for research purposes only. All other enquiries should be sent to.

*

Michael C. Grant, Luke Geoghegan, Marc Arbyn, Zakaria Mohammed, Luke McGuinness, Emily L. Clarke, Ryckie G. Wade

### JBI CRITICAL APPRAISAL CHECKLIST FOR COHORT STUDIES

Reviewer Isaac David Juárez Cruz Date 10/01/21

Author * Year 2020 Record Number 114_32574840_IDJC

| | Yes | No | Unclear | Not applicable |
|---|---|---|---|---|
| 1. Were the two groups similar and recruited from the same population? | ☑ | ☐ | ☐ | ☐ |
| 2. Were the exposures measured similarly to assign people to both exposed and unexposed groups? | ☑ | ☐ | ☐ | ☐ |
| 3. Was the exposure measured in a valid and reliable way? | ☑ | ☐ | ☐ | ☐ |
| 4. Were confounding factors identified? | ☐ | ☐ | ☑ | ☐ |
| 5. Were strategies to deal with confounding factors stated? | ☐ | ☐ | ☑ | ☐ |
| 6. Were the groups/participants free of the outcome at the start of the study (or at the moment of exposure)? | ☐ | ☐ | ☐ | ☑ |
| 7. Were the outcomes measured in a valid and reliable way? | ☑ | ☐ | ☐ | ☐ |
| 8. Was the follow up time reported and sufficient to be long enough for outcomes to occur? | ☑ | ☐ | ☐ | ☐ |
| 9. Was follow up complete, and if not, were the reasons to loss to follow up described and explored? | ☑ | ☐ | ☐ | ☐ |
| 10. Were strategies to address incomplete follow up utilized? | ☑ | ☐ | ☐ | ☐ |
| 11. Was appropriate statistical analysis used? | ☑ | ☐ | ☐ | ☐ |

Overall appraisal: Include ☐ Exclude ☑ Seek further info ☐

Comments (Including reason for exclusion)

Sin información de utilidad para la revisión

© JBI, 2020. All rights reserved. JBI grants use of these tools for research purposes only. All other enquiries should be sent to.

*

De Chang, Peng Zhao, Dawei Zhang, Jing-Hui Dong, Zhe Xu, Guang Yang, Bo-Yu Li, Hong-Xia Liu, Bo-An Li, Cheng-Feng Qin, Xiao-Hua Peng, Fu-Sheng Wang, Li-Xin Xie, Zhu Chen, Charles S. Dela Cruz, Lokesh Sharma, and En-Qiang Qin

### JBI Critical Appraisal Checklist for Case Series

Reviewer Isaac David Juárez Cruz Date 10/01/21

Author * Year 2020 Record Number 115_32549526_IDJC

| | Yes | No | Unclear | Not applicable |
|---|---|---|---|---|
| Were there clear criteria for inclusion in the case series? | ✓ | ☐ | ☐ | ☐ |
| Was the condition measured in a standard, reliable way for all participants included in the case series? | ✓ | ☐ | ☐ | ☐ |
| Were valid methods used for identification of the condition for all participants included in the case series? | ✓ | ☐ | ☐ | ☐ |
| Did the case series have consecutive inclusion of participants? | ☐ | ✓ | ☐ | ☐ |
| Did the case series have complete inclusion of participants? | ☐ | ☐ | ✓ | ☐ |
| Was there clear reporting of the demographics of the participants in the study? | ✓ | ☐ | ☐ | ☐ |
| Was there clear reporting of clinical information of the participants? | ✓ | ☐ | ☐ | ☐ |
| Were the outcomes or follow up results of cases clearly reported? | ✓ | ☐ | ☐ | ☐ |
| Was there clear reporting of the presenting site(s)/clinic(s) demographic information? | ✓ | ☐ | ☐ | ☐ |
| Was statistical analysis appropriate? | ✓ | ☐ | ☐ | ☐ |

Overall appraisal: Include ☐ Exclude ✓ Seek further info ☐

Comments (Including reason for exclusion)

© JBI, 2020. All rights reserved. JBI grants use of these tools for research purposes only. All other enquiries should be sent to.

*

Se Yoon Park, Soon Gyu Yun, Jeong Won Shin, Bo Young Lee, Hyo-Ju Son, Seungjae Lee, Eunjung Lee, and Tae Hyong Kim

### JBI CRITICAL APPRAISAL CHECKLIST FOR CASE REPORTS

Reviewer Isaac David Juárez Cruz Date 11/01/21

Author Sadaf Ali, Smitha Mathew, Joseph M. Pappachan Year 2020 Record Number 116_32544670_IDJC

| | Yes | No | Unclear | Not applicable |
|---|---|---|---|---|
| 1. Were patient's demographic characteristics clearly described? | ☑ | ☐ | ☐ | ☐ |
| 2. Was the patient's history clearly described and presented as a timeline? | ☑ | ☐ | ☐ | ☐ |
| 3. Was the current clinical condition of the patient on presentation clearly described? | ☑ | ☐ | ☐ | ☐ |
| 4. Were diagnostic tests or assessment methods and the results clearly described? | ☑ | ☐ | ☐ | ☐ |
| 5. Was the intervention(s) or treatment procedure(s) clearly described? | ☑ | ☐ | ☐ | ☐ |
| 6. Was the post-intervention clinical condition clearly described? | ☑ | ☐ | ☐ | ☐ |
| 7. Were adverse events (harms) or unanticipated events identified and described? | ☑ | ☐ | ☐ | ☐ |
| 8. Does the case report provide takeaway lessons? | ☑ | ☐ | ☐ | ☐ |

Overall appraisal: Include ☐ Exclude ☑ Seek further info ☐

Comments (Including reason for exclusion)

© JBI, 2020. All rights reserved. JBI grants use of these tools for research purposes only. All other enquiries should be sent to.

### JBI CRITICAL APPRAISAL CHECKLIST FOR COHORT STUDIES

Reviewer: Isaac David Juárez Cruz Date: 11/01/04

Author: William R. Aaron S, Joseph P. Year: 2020 Record Number: 117_32533556_IDJC

| | Yes | No | Unclear | Not applicable |
|---|---|---|---|---|
| 1. Were the two groups similar and recruited from the same population? | ☑ | ☐ | ☐ | ☐ |
| 2. Were the exposures measured similarly to assign people to both exposed and unexposed groups? | ☑ | ☐ | ☐ | ☐ |
| 3. Was the exposure measured in a valid and reliable way? | ☑ | ☐ | ☐ | ☐ |
| 4. Were confounding factors identified? | ☐ | ☐ | ☑ | ☐ |
| 5. Were strategies to deal with confounding factors stated? | ☐ | ☐ | ☑ | ☐ |
| 6. Were the groups/participants free of the outcome at the start of the study (or at the moment of exposure)? | ☐ | ☐ | ☐ | ☑ |
| 7. Were the outcomes measured in a valid and reliable way? | ☑ | ☐ | ☐ | ☐ |
| 8. Was the follow up time reported and sufficient to be long enough for outcomes to occur? | ☑ | ☐ | ☐ | ☐ |
| 9. Was follow up complete, and if not, were the reasons to loss to follow up described and explored? | ☑ | ☐ | ☐ | ☐ |
| 10. Were strategies to address incomplete follow up utilized? | ☑ | ☐ | ☐ | ☐ |
| 11. Was appropriate statistical analysis used? | ☑ | ☐ | ☐ | ☐ |

Overall appraisal: Include ☐ Exclude ☑ Seek further info ☐

Comments (Including reason for exclusion)

© JBI, 2020. All rights reserved. JBI grants use of these tools for research purposes only. All other enquiries should be sent to.

### JBI CRITICAL APPRAISAL CHECKLIST FOR CASE REPORTS

Reviewer Fernández Chirino Luisa Date 08/04/2021

Author Tung-Chen, Y. Blanco-Alonso, S., Antón-Huguet, B., Figueras-López, C., Ugueto-Rodrigo, C. Year 2020 Record Number 118_32680679_LFC

| | Yes | No | Unclear | Not applicable |
|---|---|---|---|---|
| 1. Were patient's demographic characteristics clearly described? | ☒ | ☐ | ☐ | ☐ |
| 2. Was the patient's history clearly described and presented as a timeline? | ☒ | ☐ | ☐ | ☐ |
| 3. Was the current clinical condition of the patient on presentation clearly described? | ☒ | ☐ | ☐ | ☐ |
| 4. Were diagnostic tests or assessment methods and the results clearly described? | ☒ | ☐ | ☐ | ☐ |
| 5. Was the intervention(s) or treatment procedure(s) clearly described? | ☒ | ☐ | ☐ | ☐ |
| 6. Was the post-intervention clinical condition clearly described? | ☒ | ☐ | ☐ | ☐ |
| 7. Were adverse events (harms) or unanticipated events identified and described? | ☒ | ☐ | ☐ | ☐ |
| 8. Does the case report provide takeaway lessons? | ☐ | ☐ | ☒ | ☐ |

Overall appraisal: Include ☐ Exclude ☒ Seek further info ☐

Comments (Including reason for exclusion)

Insufficient description

### JBI CRITICAL APPRAISAL CHECKLIST FOR TEXT AND OPINION PAPERS

Reviewer: Isaac David Juárez Cruz    Date: 15/01/21

Author: Gemelli against COVID-19 post accute carestudy group    Year: 2020    Record Number: 119_32529595_IDJC

| | Yes | No | Unclear | Not applicable |
|---|---|---|---|---|
| 1. Is the source of the opinion clearly identified? | ☐ | ☐ | ☐ | ☑ |
| 2. Does the source of opinion have standing in the field of expertise? | ☐ | ☐ | ☐ | ☑ |
| 3. Are the interests of the relevant population the central focus of the opinion? | ☑ | ☐ | ☐ | ☐ |
| 4. Is the stated position the result of an analytical process, and is there logic in the opinion expressed? | ☑ | ☐ | ☐ | ☐ |
| 5. Is there reference to the extant literature? | ☐ | ☐ | ☐ | ☑ |
| 6. Is any incongruence with the literature/sources logically defended? | ☐ | ☐ | ☐ | ☑ |

Overall appraisal: Include ☐ Exclude ☑ Seek further info ☐

Comments (Including reason for exclusion)

© JBI, 2020. All rights reserved. JBI grants use of these tools for research purposes only. All other enquiries should be sent to.

### JBI CRITICAL APPRAISAL CHECKLIST FOR COHORT STUDIES

Reviewer Fermández Chirino Luisa Date 08/04/2021

Author Park, S., Lee, C., Park, D., Hee-Yeon, W., Cheong, H. S. Ahn, K., Kwon, M., Joo, E. Year 2020 Record Number 120_32534042_LFC

| | Yes | No | Unclear | Not applicable |
|---|---|---|---|---|
| 1. Were the two groups similar and recruited from the same population? | ☒ | ☐ | ☐ | ☐ |
| 2. Were the exposures measured similarly to assign people<br>3. to both exposed and unexposed groups? | ☒ | ☐ | ☐ | ☐ |
| 4. Was the exposure measured in a valid and reliable way? | ☒ | ☐ | ☐ | ☐ |
| 5. Were confounding factors identified? | ☐ | ☐ | ☒ | ☐ |
| 6. Were strategies to deal with confounding factors stated? | ☐ | ☐ | ☒ | ☐ |
| 7. Were the groups/participants free of the outcome at the start of the study (or at the moment of exposure)? | ☐ | ☒ | ☐ | ☐ |
| 8. Were the outcomes measured in a valid and reliable way? | ☒ | ☐ | ☐ | ☐ |
| 9. Was the follow up time reported and sufficient to be long enough for outcomes to occur? | ☐ | ☒ | ☐ | ☐ |
| 10. Was follow up complete, and if not, were the reasons to loss to follow up described and explored? | ☒ | ☐ | ☐ | ☐ |
| 11. Were strategies to address incomplete follow up utilized? | ☒ | ☐ | ☐ | ☐ |
| 12. Was appropriate statistical analysis used? | ☒ | ☐ | ☐ | ☐ |

Overall appraisal: Include ☒ Exclude ☐ Seek further info ☐

Comments (including reason for exclusion)

### JBI CRITICAL APPRAISAL CHECKLIST FOR ANALYTICAL CROSS SECTIONAL STUDIES

Reviewer Fernández Chirino Luisa Date 08/04/2021

Author Yan, C., Prajapati, D., Ritter, M., DeConde, A. Year 2020 Record Number 121_32513065_LFC

| | Yes | No | Unclear | Not applicable |
|---|---|---|---|---|
| 1. Were the criteria for inclusion in the sample clearly defined? | ☐ | ☒ | ☐ | ☐ |
| 2. Were the study subjects and the setting described in detail? | ☒ | ☐ | ☐ | ☐ |
| 3. Was the exposure measured in a valid and reliable way? | ☒ | ☐ | ☐ | ☐ |
| 4. Were objective, standard criteria used for measurement of the condition? | ☒ | ☐ | ☐ | ☐ |
| 5. Were confounding factors identified? | ☐ | ☐ | ☒ | ☐ |
| 6. Were strategies to deal with confounding factors stated? | ☐ | ☐ | ☒ | ☐ |
| 7. Were the outcomes measured in a valid and reliable way? | ☒ | ☐ | ☐ | ☐ |
| 8. Was appropriate statistical analysis used? | ☒ | ☐ | ☐ | ☐ |

Overall appraisal: Include ☐ Exclude ☒ Seek further info ☐

Comments (Including reason for exclusion)

______________________________________________

______________________________________________

______________________________________________

### JBI CRITICAL APPRAISAL CHECKLIST FOR CASE REPORTS

Reviewer Fernández Chirino Luisa Date 08/04/2021

Author Pavon, A. G., Meier, D., Samim, D., Rotzinger, D., Fournier, S., Marquis, P., Monney, P., Muller, O., Schwitter, J. Year 2020 Record Number 122_32522523_LFC

| | Yes | No | Unclear | Not applicable |
|---|---|---|---|---|
| 1. Were patient's demographic characteristics clearly described? | ☒ | ☐ | ☐ | ☐ |
| 2. Was the patient's history clearly described and presented as a timeline? | ☒ | ☐ | ☐ | ☐ |
| 3. Was the current clinical condition of the patient on presentation clearly described? | ☒ | ☐ | ☐ | ☐ |
| 4. Were diagnostic tests or assessment methods and the results clearly described? | ☒ | ☐ | ☐ | ☐ |
| 5. Was the intervention(s) or treatment procedure(s) clearly described? | ☒ | ☐ | ☐ | ☐ |
| 6. Was the post-intervention clinical condition clearly described? | ☒ | ☐ | ☐ | ☐ |
| 7. Were adverse events (harms) or unanticipated events identified and described? | ☒ | ☐ | ☐ | ☐ |
| 8. Does the case report provide takeaway lessons? | ☐ | ☐ | ☒ | ☐ |

Overall appraisal: Include ☐ Exclude ☒ Seek further info ☐

Comments (Including reason for exclusion)

The article talks about persostent COVID not Long COVID, as the patient remained positive.

### JBI CRITICAL APPRAISAL CHECKLIST FOR CASE CONTROL STUDIES

Reviewer Fermández Chirino Luisa Date 08/04/2021

Author Chung, T. W., Sridhar, S., Zhang, A., J., Chan, K., et al. Year 2020 Record Number 123_32548209_LFC

| | Yes | No | Unclear | Not applicable |
|---|---|---|---|---|
| 1. Were the groups comparable other than the presence of disease in cases or the absence of disease in controls? | ☒ | ☐ | ☐ | ☐ |
| 2. Were cases and controls matched appropriately? | ☒ | ☐ | ☐ | ☐ |
| 3. Were the same criteria used for identification of cases and controls? | ☒ | ☐ | ☐ | ☐ |
| 4. Was exposure measured in a standard, valid and reliable way? | ☒ | ☐ | ☐ | ☐ |
| 5. Was exposure measured in the same way for cases and controls? | ☒ | ☐ | ☐ | ☐ |
| 6. Were confounding factors identified? | ☐ | ☐ | ☒ | ☐ |
| 7. Were strategies to deal with confounding factors stated? | ☐ | ☐ | ☒ | ☐ |
| 8. Were outcomes assessed in a standard, valid and reliable way for cases and controls? | ☒ | ☐ | ☐ | ☐ |
| 9. Was the exposure period of interest long enough to be meaningful? | ☒ | ☐ | ☐ | ☐ |
| 10. Was appropriate statistical analysis used? | ☒ | ☐ | ☐ | ☐ |

Overall appraisal: Include ☐ Exclude ☒ Seek further info ☐

Comments (including reason for exclusion)

### EXPLANATION OF CASE CONTROL STUDIES CRITICAL APPRAISAL

How to cite: *Moola S, Munn Z, Tufanaru C, Aromataris E, Sears K, Sfetcu R, Currie M, Qureshi R, Mattis P, Lisy K, Mu P-F. Chapter 7: Systematic reviews of etiology and risk . In: Aromataris E, Munn Z (Editors). JBI Manual for Evidence Synthesis.* JBI, 2020. Available from https://synthesismanual.jbi.global

**Case Control Studies Critical Appraisal Tool**

Answers: Yes, No, Unclear or Not/Applicable

1. **Were the groups comparable other than presence of disease in cases or absence of disease in controls?**

   The control group should be representative of the source population that produced the cases. This is usually done by individual matching; wherein controls are selected for each case on the basis of similarity with respect to certain characteristics other than the exposure of interest. Frequency or group matching is an alternative method. Selection bias may result if the groups are not comparable.

2. **Were cases and controls matched appropriately?**

   As in item 1, the study should include clear definitions of the source population. Sources from which cases and controls were recruited should be carefully looked at. For example, cancer registries may be used to recruit participants in a study examining risk factors for lung cancer, which typify population-based case control studies. Study participants may be selected from the target population, the source population, or from a pool of eligible participants (such as in hospital-based case control studies).

3. **Were the same criteria used for identification of cases and controls?**

   It is useful to determine if patients were included in the study based on either a specified diagnosis or definition. This is more likely to decrease the risk of bias. Characteristics are another useful approach to matching groups, and studies that did not use specified diagnostic methods or definitions should provide evidence on matching by key characteristics. A case should be defined clearly. It is also important that controls must fulfil all the eligibility criteria defined for the cases except for those relating to diagnosis of the disease.

4. **Was exposure measured in a standard, valid and reliable way?**

   The study should clearly describe the method of measurement of exposure. Assessing validity requires that a 'gold standard' is available to which the measure can be compared. The validity of exposure measurement usually relates to whether a current measure is appropriate or whether a measure of past exposure is needed.

   Case control studies may investigate many different 'exposures' that may or may not be associated with the condition. In these cases, reviewers should use the main exposure of interest for their review to answer this question when using this tool at the study level.

   Reliability refers to the processes included in an epidemiological study to check repeatability of measurements of the exposures. These usually include intra-observer reliability and inter-observer reliability.

5. **Was exposure measured in the same way for cases and controls?**

   As in item 4, the study should clearly describe the method of measurement of exposure. The exposure measures should be clearly defined and described in detail. Assessment of exposure or

© JBI, 2020. All rights reserved. JBI grants use of these tools for research purposes only. All other enquiries should be sent to.

risk factors should have been carried out according to same procedures or protocols for both cases and controls.

##### 6. Were confounding factors identified?

Confounding has occurred where the estimated intervention exposure effect is biased by the presence of some difference between the comparison groups (apart from the exposure investigated/of interest). Typical confounders include baseline characteristics, prognostic factors, or concomitant exposures (e.g. smoking). A confounder is a difference between the comparison groups and it influences the direction of the study results. A high quality study at the level of case control design will identify the potential confounders and measure them (where possible). This is difficult for studies where behavioral, attitudinal or lifestyle factors may impact on the results.

##### 7. Were strategies to deal with confounding factors stated?

Strategies to deal with effects of confounding factors may be dealt within the study design or in data analysis. By matching or stratifying sampling of participants, effects of confounding factors can be adjusted for. When dealing with adjustment in data analysis, assess the statistics used in the study. Most will be some form of multivariate regression analysis to account for the confounding factors measured. Look out for a description of statistical methods as regression methods such as logistic regression are usually employed to deal with confounding factors/ variables of interest.

##### 8. Were outcomes assessed in a standard, valid and reliable way for cases and controls?

Read the methods section of the paper. If for e.g. lung cancer is assessed based on existing definitions or diagnostic criteria, then the answer to this question is likely to be yes. If lung cancer is assessed using observer reported, or self-reported scales, the risk of over- or under-reporting is increased, and objectivity is compromised. Importantly, determine if the measurement tools used were validated instruments as this has a significant impact on outcome assessment validity.

Having established the objectivity of the outcome measurement (e.g. lung cancer) instrument, it's important to establish how the measurement was conducted. Were those involved in collecting data trained or educated in the use of the instrument/s? (e.g. radiographers). If there was more than one data collector, were they similar in terms of level of education, clinical or research experience, or level of responsibility in the piece of research being appraised?

##### 9. Was the exposure period of interest long enough to be meaningful?

It is particularly important in a case control study that the exposure time was sufficient enough to show an association between the exposure and the outcome. It may be that the exposure period may be too short or too long to influence the outcome.

##### 10. Was appropriate statistical analysis used?

As with any consideration of statistical analysis, consideration should be given to whether there was a more appropriate alternate statistical method that could have been used. The methods section should be detailed enough for reviewers to identify which analytical techniques were used (in particular, regression or stratification) and how specific confounders were measured.

For studies utilizing regression analysis, it is useful to identify if the study identified which variables were included and how they related to the outcome. If stratification was the analytical approach used, were the strata of analysis defined by the specified variables? Additionally, it is also important to assess the appropriateness of the analytical strategy in terms of the assumptions associated with the approach as differing methods of analysis are based on differing assumptions about the data and how it will respond.

© JBI, 2020. All rights reserved. JBI grants use of these tools for research purposes only. All other enquiries should be sent to.

© JBI, 2020. All rights reserved. JBI grants use of these tools for research purposes only. All other enquiries should be sent to.

### JBI CRITICAL APPRAISAL CHECKLIST FOR TEXT AND OPINION PAPERS

Reviewer Isaac David Juárez Cruz Date 29/01/21

Author ★ Year 2020 Record Number 124_30505489_IDJC

| | Yes | No | Unclear | Not applicable |
|---|---|---|---|---|
| 1. Is the source of the opinion clearly identified? | ☐ | ☐ | ☐ | ☑ |
| 2. Does the source of opinion have standing in the field of expertise? | ☐ | ☐ | ☐ | ☑ |
| 3. Are the interests of the relevant population the central focus of the opinion? | ☑ | ☐ | ☐ | ☐ |
| 4. Is the stated position the result of an analytical process, and is there logic in the opinion expressed? | ☐ | ☐ | ☐ | ☑ |
| 5. Is there reference to the extant literature? | ☐ | ☐ | ☐ | ☑ |
| 6. Is any incongruence with the literature/sources logically defended? | ☐ | ☐ | ☐ | ☑ |

Overall appraisal: Include ☐ Exclude ☑ Seek further info ☐

Comments (Including reason for exclusion)

© JBI, 2020. All rights reserved. JBI grants use of these tools for research purposes only. All other enquiries should be sent to.

*

Sandro Iannaccone, Paola Castellazzi, Andrea Tettamanti, Elise Houdayer, Luigia Brugliera, Francesco de Blasio, Paolo Cimino, Marco Ripa, Carlo Meloni, Federica Alemanno, Paolo Scarpellini

### JBI CRITICAL APPRAISAL CHECKLIST FOR CASE REPORTS

Reviewer Juárez Cruz Isaac David Date 29/01/21

Author Jieqing Jessica Xu, Daniel Samaha, Suhas Mondhe, David MassicotteAzarniouch, Gregory Knoll, Marcel Ruzicka Year 2020 Record Number 125_32483909_IDJC

| | Yes | No | Unclear | Not applicable |
|---|---|---|---|---|
| 1. Were patient's demographic characteristics clearly described? | ☑ | ☐ | ☐ | ☐ |
| 2. Was the patient's history clearly described and presented as a timeline? | ☑ | ☐ | ☐ | ☐ |
| 3. Was the current clinical condition of the patient on presentation clearly described? | ☑ | ☐ | ☐ | ☐ |
| 4. Were diagnostic tests or assessment methods and the results clearly described? | ☑ | ☐ | ☐ | ☐ |
| 5. Was the intervention(s) or treatment procedure(s) clearly described? | ☑ | ☐ | ☐ | ☐ |
| 6. Was the post-intervention clinical condition clearly described? | ☑ | ☐ | ☐ | ☐ |
| 7. Were adverse events (harms) or unanticipated events identified and described? | ☑ | ☐ | ☐ | ☐ |
| 8. Does the case report provide takeaway lessons? | ☑ | ☐ | ☐ | ☐ |

Overall appraisal: Include ☑ Exclude ☐ Seek further info ☐

Comments (Including reason for exclusion)

© JBI, 2020. All rights reserved. JBI grants use of these tools for research purposes only. All other enquiries should be sent to.

### JBI CRITICAL APPRAISAL CHECKLIST FOR ANALYTICAL CROSS SECTIONAL STUDIES

Reviewer Isaac David Juárez Cruz Date 16/01/21

Author * Year 2020 Record Number 126_32381497_IDJC

| | Yes | No | Unclear | Not applicable |
|---|---|---|---|---|
| 1. Were the criteria for inclusion in the sample clearly defined? | ☑ | ☐ | ☐ | ☐ |
| 2. Were the study subjects and the setting described in detail? | ☐ | ☑ | ☐ | ☐ |
| 3. Was the exposure measured in a valid and reliable way? | ☑ | ☐ | ☐ | ☐ |
| 4. Were objective, standard criteria used for measurement of the condition? | ☑ | ☐ | ☐ | ☐ |
| 5. Were confounding factors identified? | ☐ | ☑ | ☐ | ☐ |
| 6. Were strategies to deal with confounding factors stated? | ☐ | ☐ | ☑ | ☐ |
| 7. Were the outcomes measured in a valid and reliable way? | ☑ | ☐ | ☐ | ☐ |
| 8. Was appropriate statistical analysis used? | ☑ | ☐ | ☐ | ☐ |

Overall appraisal: Include ☐ Exclude ☑ Seek further info ☐

Comments (Including reason for exclusion)

© JBI, 2020. All rights reserved. JBI grants use of these tools for research purposes only. All other enquiries should be sent to.

### JBI CRITICAL APPRAISAL CHECKLIST FOR ANALYTICAL CROSS SECTIONAL STUDIES

Reviewer: Isaac David Juárez Cruz  Date: 16/01/21

Author: *  Year: 2020  Record Number: 126_32381497_IDJC

| | Yes | No | Unclear | Not applicable |
|---|---|---|---|---|
| 1. Were the criteria for inclusion in the sample clearly defined? | ☑ | ☐ | ☐ | ☐ |
| 2. Were the study subjects and the setting described in detail? | ☐ | ☑ | ☐ | ☐ |
| 3. Was the exposure measured in a valid and reliable way? | ☑ | ☐ | ☐ | ☐ |
| 4. Were objective, standard criteria used for measurement of the condition? | ☑ | ☐ | ☐ | ☐ |
| 5. Were confounding factors identified? | ☐ | ☑ | ☐ | ☐ |
| 6. Were strategies to deal with confounding factors stated? | ☐ | ☐ | ☑ | ☐ |
| 7. Were the outcomes measured in a valid and reliable way? | ☑ | ☐ | ☐ | ☐ |
| 8. Was appropriate statistical analysis used? | ☑ | ☐ | ☐ | ☐ |

Overall appraisal: Include ☐ Exclude ☑ Seek further info ☐

Comments (Including reason for exclusion)

© JBI, 2020. All rights reserved. JBI grants use of these tools for research purposes only. All other enquiries should be sent to.

### JBI CRITICAL APPRAISAL CHECKLIST FOR SYSTEMATIC REVIEWS AND RESEARCH SYNTHESES

Reviewer Isaac David Juárez Cruz Date 30/01/21

Author * Year 2020 Record Number 127_3224475821_IDJC

| | Yes | No | Unclear | Not applicable |
|---|---|---|---|---|
| 1. Is the review question clearly and explicitly stated? | ☑ | ☐ | ☐ | ☐ |
| 2. Were the inclusion criteria appropriate for the review question? | ☑ | ☐ | ☐ | ☐ |
| 3. Was the search strategy appropriate? | ☑ | ☐ | ☐ | ☐ |
| 4. Were the sources and resources used to search for studies adequate? | ☑ | ☐ | ☐ | ☐ |
| 5. Were the criteria for appraising studies appropriate? | ☑ | ☐ | ☐ | ☐ |
| 6. Was critical appraisal conducted by two or more reviewers independently? | ☑ | ☐ | ☐ | ☐ |
| 7. Were there methods to minimize errors in data extraction? | ☑ | ☐ | ☐ | ☐ |
| 8. Were the methods used to combine studies appropriate? | ☑ | ☐ | ☐ | ☐ |
| 9. Was the likelihood of publication bias assessed? | ☑ | ☐ | ☐ | ☐ |
| 10. Were recommendations for policy and/or practice supported by the reported data? | ☑ | ☐ | ☐ | ☐ |
| 11. Were the specific directives for new research appropriate? | ☑ | ☐ | ☐ | ☐ |

Overall appraisal: Include Exclude ☑ Seek further info ☐

Comments (Including reason for exclusion)

© JBI, 2020. All rights reserved. JBI grants use of these tools for research purposes only. All other enquiries should be sent to.

*

Robert M Barker-Davies, Oliver O'Sullivan, Kahawalage Pumi Prathima Senaratne , Polly Baker, Mark Cranley, Shreshth Dharm-Datta, Henrietta Ellis, Duncan Goodall,4,7 Michael Gough, Sarah Lewis, Jonathan Norman, Theodora Papadopoulou,4,8 David Roscoe, Daniel Sherwood, Philippa Turner, Tammy Walker, Alan Mistlin, Rhodri Phillip, Alastair M Nicol, Alexander N Bennett, Sardar Bahadur

### JBI CRITICAL APPRAISAL CHECKLIST FOR STUDIES REPORTING PREVALENCE DATA

Reviewer Isaac David Juárez Cruz Date 30/01/21

Author Saurabh Gombara, Marcello Chang, Catherine A. Hogan, James Zehnder, Scott Boyda, Benjamin A. Pinsky, Nigam H. Shah Year 2020 Record Number 128_32505778_IDJC

| | Yes | No | Unclear | Not applicable |
|---|---|---|---|---|
| 1. Was the sample frame appropriate to address the target population? | ☐ | ☐ | ☐ | ✓ |
| 2. Were study participants sampled in an appropriate way? | ✓ | ☐ | ☐ | ☐ |
| 3. Was the sample size adequate? | ✓ | ☐ | ☐ | ☐ |
| 4. Were the study subjects and the setting described in detail? | ✓ | ☐ | ☐ | ☐ |
| 5. Was the data analysis conducted with sufficient coverage of the identified sample? | ☐ | ☐ | ✓ | ☐ |
| 6. Were valid methods used for the identification of the condition? | ✓ | ☐ | ☐ | ☐ |
| 7. Was the condition measured in a standard, reliable way for all participants? | ✓ | ☐ | ☐ | ☐ |
| 8. Was there appropriate statistical analysis? | ✓ | ☐ | ☐ | ☐ |
| 9. Was the response rate adequate, and if not, was the low response rate managed appropriately? | ☐ | ☐ | ☐ | ✓ |

Overall appraisal: Include ☐ Exclude ✓ Seek further info ☐

Comments (Including reason for exclusion)

© JBI, 2020. All rights reserved. JBI grants use of these tools for research purposes only. All other enquiries should be sent to.

### JBI CRITICAL APPRAISAL CHECKLIST FOR TEXT AND OPINION PAPERS

Reviewer: Isaac David Juárez Cruz  Date: 30/01/21

Author: Felix Inchausti, Angus MacBeth, Ilanit Hasson-Ohayon, Giancarlo Dimaggio  Year: 2020  Record Number: 129_32836375_IDJC

| | Yes | No | Unclear | Not applicable |
|---|---|---|---|---|
| 1. Is the source of the opinion clearly identified? | ☑ | ☐ | ☐ | ☐ |
| 2. Does the source of opinion have standing in the field of expertise? | ☑ | ☐ | ☐ | ☐ |
| 3. Are the interests of the relevant population the central focus of the opinion? | ☑ | ☐ | ☐ | ☐ |
| 4. Is the stated position the result of an analytical process, and is there logic in the opinion expressed? | ☑ | ☐ | ☐ | ☐ |
| 5. Is there reference to the extant literature? | ☑ | ☐ | ☐ | ☐ |
| 6. Is any incongruence with the literature/sources logically defended? | ☐ | ☐ | ☑ | ☐ |

Overall appraisal: Include ☐ Exclude ☑ Seek further info ☐

Comments (Including reason for exclusion)

© JBI, 2020. All rights reserved. JBI grants use of these tools for research purposes only. All other enquiries should be sent to.

### JBI CRITICAL APPRAISAL CHECKLIST FOR COHORT STUDIES

Reviewer Marco Antonio Delaye Martínez Date 24/04/21

Author Huang L., Zhao P., Tang D., et al. Year 2020 Record Number 130_32763118

| | Yes | No | Unclear | Not applicable |
|---|---|---|---|---|
| 1. Were the two groups similar and recruited from the same population? | ☑ | ☐ | ☐ | ☐ |
| 2. Were the exposures measured similarly to assign people to both exposed and unexposed groups? | ☑ | ☐ | ☐ | ☐ |
| 3. Was the exposure measured in a valid and reliable way? | ☑ | ☐ | ☐ | ☐ |
| 4. Were confounding factors identified? | ☑ | ☐ | ☐ | ☐ |
| 5. Were strategies to deal with confounding factors stated? | ☑ | ☐ | ☐ | ☐ |
| 6. Were the groups/participants free of the outcome at the start of the study (or at the moment of exposure)? | ☑ | ☐ | ☐ | ☐ |
| 7. Were the outcomes measured in a valid and reliable way? | ☑ | ☐ | ☐ | ☐ |
| 8. Was the follow up time reported and sufficient to be long enough for outcomes to occur? | ☑ | ☐ | ☐ | ☐ |
| 9. Was follow up complete, and if not, were the reasons to loss to follow up described and explored? | ☑ | ☐ | ☐ | ☐ |
| 10. Were strategies to address incomplete follow up utilized? | ☑ | ☐ | ☐ | ☐ |
| 11. Was appropriate statistical analysis used? | ☑ | ☐ | ☐ | ☐ |

Overall appraisal: Include ☑ Exclude ☐ Seek further info ☐

Comments (Including reason for exclusion)

No comments.

© JBI, 2020. All rights reserved. JBI grants use of these tools for research purposes only. All other enquiries should be sent to.

### JBI CRITICAL APPRAISAL CHECKLIST FOR COHORT STUDIES

Reviewer Isaac David Juárez Cruz Date 30/01/21

Author Claire Hopkins, Pavol Surda, Emily Whitehead and B. Nirmal Kumar Year 2020 Record Number 131_32366299_IDJC

| | Yes | No | Unclear | Not applicable |
|---|---|---|---|---|
| 1. Were the two groups similar and recruited from the same population? | ☐ | ☐ | ☐ | ☑ |
| 2. Were the exposures measured similarly to assign people to both exposed and unexposed groups? | ☐ | ☐ | ☐ | ☑ |
| 3. Was the exposure measured in a valid and reliable way? | ☐ | ☐ | ☑ | ☐ |
| 4. Were confounding factors identified? | ☐ | ☑ | ☐ | ☐ |
| 5. Were strategies to deal with confounding factors stated? | ☐ | ☑ | ☐ | ☐ |
| 6. Were the groups/participants free of the outcome at the start of the study (or at the moment of exposure)? | ☐ | ☑ | ☐ | ☐ |
| 7. Were the outcomes measured in a valid and reliable way? | ☐ | ☐ | ☑ | ☐ |
| 8. Was the follow up time reported and sufficient to be long enough for outcomes to occur? | ☑ | ☐ | ☐ | ☐ |
| 9. Was follow up complete, and if not, were the reasons to loss to follow up described and explored? | ☑ | ☐ | ☐ | ☐ |
| 10. Were strategies to address incomplete follow up utilized? | ☑ | ☐ | ☐ | ☐ |
| 11. Was appropriate statistical analysis used? | ☑ | ☐ | ☐ | ☐ |

Overall appraisal: Include ☐ Exclude ☑ Seek further info ☐

Comments (Including reason for exclusion)

No a todos los participantes se les realizo RT-PCR por lo que los datos carecen de validez

© JBI, 2020. All rights reserved. JBI grants use of these tools for research purposes only. All other enquiries should be sent to.

### JBI CRITICAL APPRAISAL CHECKLIST FOR ANALYTICAL CROSS SECTIONAL STUDIES

Reviewer Marco Antonio Delaye Martínez Date 7/04/21

Author Yan C., Prajapati D., Either M., et al. Year 2020 Record Number 132_32513065

| | Yes | No | Unclear | Not applicable |
|---|---|---|---|---|
| 1. Were the criteria for inclusion in the sample clearly defined? | ☐ | ☑ | ☐ | ☐ |
| 2. Were the study subjects and the setting described in detail? | ☐ | ☑ | ☐ | ☐ |
| 3. Was the exposure measured in a valid and reliable way? | ☑ | ☐ | ☐ | ☐ |
| 4. Were objective, standard criteria used for measurement of the condition? | ☐ | ☑ | ☐ | ☐ |
| 5. Were confounding factors identified? | ☐ | ☑ | ☐ | ☐ |
| 6. Were strategies to deal with confounding factors stated? | ☐ | ☑ | ☐ | ☐ |
| 7. Were the outcomes measured in a valid and reliable way? | ☑ | ☐ | ☐ | ☐ |
| 8. Was appropriate statistical analysis used? | ☐ | ☑ | ☐ | ☐ |

Overall appraisal: Include ☐ Exclude ● Seek further info ☐

Comments (Including reason for exclusion)

El estudio no aporta información completa sobre las características demográficas y clínicas de los pacientes. No se puede establecer de forma concreta el tiempo de seguimiento. Cabe la posibilidad de que algunos pacientesnfueran entrevistados antes de los 21 días de seguimiento.

© JBI, 2020. All rights reserved. JBI grants use of these tools for research purposes only. All other enquiries should be sent to.

### JBI CRITICAL APPRAISAL CHECKLIST FOR ANALYTICAL CROSS SECTIONAL STUDIES

Reviewer Marco Antonio Delaye Martínez Date 7/04/21

Author Barón-Sánchez J., Santiago C., Goizueta G., et al. Year 2020 Record Number 133_32900532

| | Yes | No | Unclear | Not applicable |
|---|---|---|---|---|
| 1. Were the criteria for inclusion in the sample clearly defined? | ☑ | ☐ | ☐ | ☐ |
| 2. Were the study subjects and the setting described in detail? | ☑ | ☐ | ☐ | ☐ |
| 3. Was the exposure measured in a valid and reliable way? | ☐ | ☑ | ☐ | ☐ |
| 4. Were objective, standard criteria used for measurement of the condition? | ☑ | ☐ | ☐ | ☐ |
| 5. Were confounding factors identified? | ☐ | ☑ | ☐ | ☐ |
| 6. Were strategies to deal with confounding factors stated? | ☐ | ☑ | ☐ | ☐ |
| 7. Were the outcomes measured in a valid and reliable way? | ☑ | ☐ | ☐ | ☐ |
| 8. Was appropriate statistical analysis used? | ☑ | ☐ | ☐ | ☐ |

Overall appraisal: Include ☐ Exclude ● Seek further info ☐

Comments (Including reason for exclusion)

Sólo 31 pacientes contaban con prueba PCR positiva para SARS-CoV-2. No se reportan caracteristicas claras de relevancia para los fines de la presente revisuon sistematica.

© JBI, 2020. All rights reserved. JBI grants use of these tools for research purposes only. All other enquiries should be sent to.

### JBI CRITICAL APPRAISAL CHECKLIST FOR ANALYTICAL CROSS SECTIONAL STUDIES

Reviewer: Daniel de la Rosa  Date: 22-11-20

Author: Louis Poncet-Megemont, et al  Year: 2020  Record Number: 135_32757419

| | Yes | No | Unclear | Not applicable |
|---|---|---|---|---|
| 1. Were the criteria for inclusion in the sample clearly defined? | ☒ | ☐ | ☐ | ☐ |
| 2. Were the study subjects and the setting described in detail? | ☒ | ☐ | ☐ | ☐ |
| 3. Was the exposure measured in a valid and reliable way? | ☒ | ☐ | ☐ | ☐ |
| 4. Were objective, standard criteria used for measurement of the condition? | ☒ | ☐ | ☐ | ☐ |
| 5. Were confounding factors identified? | ☐ | ☒ | ☐ | ☐ |
| 6. Were strategies to deal with confounding factors stated? | ☐ | ☒ | ☐ | ☐ |
| 7. Were the outcomes measured in a valid and reliable way? | ☒ | ☐ | ☐ | ☐ |
| 8. Was appropriate statistical analysis used? | ☒ | ☐ | ☐ | ☐ |

Overall appraisal: Include ☐ Exclude ☒ Seek further info ☐

Comments (Including reason for exclusion)

The diagnostic method was not PCR for all participants

### JBI CRITICAL APPRAISAL CHECKLIST FOR COHORT STUDIES

Reviewer Marco Antonio Delaye Martinez Date 07/04/21

Author Gong Y., Guan L., Jin Z.m et al. Year 2020 Record Number 136_32441786

| | Yes | No | Unclear | Not applicable |
|---|---|---|---|---|
| 1. Were the two groups similar and recruited from the same population? | ☑ | ☐ | ☐ | ☐ |
| 2. Were the exposures measured similarly to assign people to both exposed and unexposed groups? | ☑ | ☐ | ☐ | ☐ |
| 3. Was the exposure measured in a valid and reliable way? | ☑ | ☐ | ☐ | ☐ |
| 4. Were confounding factors identified? | ☐ | ☑ | ☐ | ☐ |
| 5. Were strategies to deal with confounding factors stated? | ☐ | ☑ | ☐ | ☐ |
| 6. Were the groups/participants free of the outcome at the start of the study (or at the moment of exposure)? | ☑ | ☐ | ☐ | ☐ |
| 7. Were the outcomes measured in a valid and reliable way? | ☑ | ☐ | ☐ | ☐ |
| 8. Was the follow up time reported and sufficient to be long enough for outcomes to occur? | ☑ | ☐ | ☐ | ☐ |
| 9. Was follow up complete, and if not, were the reasons to loss to follow up described and explored? | ☑ | ☐ | ☐ | ☐ |
| 10. Were strategies to address incomplete follow up utilized? | ☐ | ☐ | ☑ | ☐ |
| 11. Was appropriate statistical analysis used? | ☑ | ☐ | ☐ | ☐ |

Overall appraisal: Include ☐ Exclude ☑ Seek further info ☐

Comments (Including reason for exclusion)

El articulo no cumple con los criterios básicos de seleccciónny no aporta informacion útil para los objetivos de la presente revisión sistemática.

© JBI, 2020. All rights reserved. JBI grants use of these tools for research purposes only. All other enquiries should be sent to.

### JBI CRITICAL APPRAISAL CHECKLIST FOR ANALYTICAL CROSS SECTIONAL STUDIES

Reviewer Marco Antonio Delaye Martínez Date 7/04/21

Author Akter F., Mannan A., Mehedi H., et al. Year 2020 Record Number 137_33113469

| | Yes | No | Unclear | Not applicable |
|---|---|---|---|---|
| 1. Were the criteria for inclusion in the sample clearly defined? | ✔ | ☐ | ☐ | ☐ |
| 2. Were the study subjects and the setting described in detail? | ✔ | ☐ | ☐ | ☐ |
| 3. Was the exposure measured in a valid and reliable way? | ✔ | ☐ | ☐ | ☐ |
| 4. Were objective, standard criteria used for measurement of the condition? | ✔ | ☐ | ☐ | ☐ |
| 5. Were confounding factors identified? | ☐ | ☐ | ✔ | ☐ |
| 6. Were strategies to deal with confounding factors stated? | ☐ | ☐ | ✔ | ☐ |
| 7. Were the outcomes measured in a valid and reliable way? | ✔ | ☐ | ☐ | ☐ |
| 8. Was appropriate statistical analysis used? | ✔ | ☐ | ☐ | ☐ |

Overall appraisal: Include ☐ Exclude ✔ Seek further info ☐

Comments (Including reason for exclusion)

© JBI, 2020. All rights reserved. JBI grants use of these tools for research purposes only. All other enquiries should be sent to.

### JBI CRITICAL APPRAISAL CHECKLIST FOR TEXT AND OPINION PAPERS

Reviewer Marco Antonio Delaye Martinez Date 07/04/20

Author Klitzman R. Year 2020 Record Number 138_33103966

| | Yes | No | Unclear | Not applicable |
|---|---|---|---|---|
| 1. Is the source of the opinion clearly identified? | ☑ | ☐ | ☐ | ☐ |
| 2. Does the source of opinion have standing in the field of expertise? | ☑ | ☐ | ☐ | ☐ |
| 3. Are the interests of the relevant population the central focus of the opinion? | ☑ | ☐ | ☐ | ☐ |
| 4. Is the stated position the result of an analytical process, and is there logic in the opinion expressed? | ☑ | ☐ | ☐ | ☐ |
| 5. Is there reference to the extant literature? | ☑ | ☐ | ☐ | ☐ |
| 6. Is any incongruence with the literature/sources logically defended? | ☑ | ☐ | ☐ | ☐ |

Overall appraisal: Include ☐ Exclude ☑ Seek further info ☐

Comments (Including reason for exclusion)

El artículo no cuenta con las características definidas para ser incluido en la presente revisión sistemática. No importa información más allá de la ya documentada en artículos seleccionados.

© JBI, 2020. All rights reserved. JBI grants use of these tools for research purposes only. All other enquiries should be sent to.

### JBI CRITICAL APPRAISAL CHECKLIST FOR COHORT STUDIES

Reviewer Marco Antonio Delaye Martínez Date 07/04/21

Author Caronna E., Ballvé A., Llauradó A., et al. Year 2020 Record Number 139_33146036

| | Yes | No | Unclear | Not applicable |
|---|---|---|---|---|
| 1. Were the two groups similar and recruited from the same population? | ☑ | ☐ | ☐ | ☐ |
| 2. Were the exposures measured similarly to assign people to both exposed and unexposed groups? | ☑ | ☐ | ☐ | ☐ |
| 3. Was the exposure measured in a valid and reliable way? | ☑ | ☐ | ☐ | ☐ |
| 4. Were confounding factors identified? | ☐ | ☑ | ☐ | ☐ |
| 5. Were strategies to deal with confounding factors stated? | ☐ | ☑ | ☐ | ☐ |
| 6. Were the groups/participants free of the outcome at the start of the study (or at the moment of exposure)? | ☑ | ☐ | ☐ | ☐ |
| 7. Were the outcomes measured in a valid and reliable way? | ☑ | ☐ | ☐ | ☐ |
| 8. Was the follow up time reported and sufficient to be long enough for outcomes to occur? | ☑ | ☐ | ☐ | ☐ |
| 9. Was follow up complete, and if not, were the reasons to loss to follow up described and explored? | ☐ | ☑ | ☐ | ☐ |
| 10. Were strategies to address incomplete follow up utilized? | ☑ | ☐ | ☐ | ☐ |
| 11. Was appropriate statistical analysis used? | ☑ | ☐ | ☐ | ☐ |

Overall appraisal: Include ☑ Exclude ☐ Seek further info ☐

Comments (Including reason for exclusion)

No comments.

© JBI, 2020. All rights reserved. JBI grants use of these tools for research purposes only. All other enquiries should be sent to.

### CHECKLIST FOR ANALYTICAL CROSS SECTIONAL STUDIES

Critical Appraisal tools for use in JBI Systematic Reviews

### INTRODUCTION

JBI is an JBI is an international research organisation based in the Faculty of Health and Medical Sciences at the University of Adelaide, South Australia. JBI develops and delivers unique evidence-based information, software, education and training designed to improve healthcare practice and health outcomes. With over 70 Collaborating Entities, servicing over 90 countries, JBI is a recognised global leader in evidence-based healthcare.

#### JBI Systematic Reviews

The core of evidence synthesis is the systematic review of literature of a particular intervention, condition or issue. The systematic review is essentially an analysis of the available literature (that is, evidence) and a judgment of the effectiveness or otherwise of a practice, involving a series of complex steps. JBI takes a particular view on what counts as evidence and the methods utilised to synthesise those different types of evidence. In line with this broader view of evidence, JBI has developed theories, methodologies and rigorous processes for the critical appraisal and synthesis of these diverse forms of evidence in order to aid in clinical decision-making in healthcare. There now exists JBI guidance for conducting reviews of effectiveness research, qualitative research, prevalence/incidence, etiology/risk, economic evaluations, text/opinion, diagnostic test accuracy, mixed-methods, umbrella reviews and scoping reviews. Further information regarding JBI systematic reviews can be found in the JBI Evidence Synthesis Manual.

#### JBI Critical Appraisal Tools

All systematic reviews incorporate a process of critique or appraisal of the research evidence. The purpose of this appraisal is to assess the methodological quality of a study and to determine the extent to which a study has addressed the possibility of bias in its design, conduct and analysis. All papers selected for inclusion in the systematic review (that is – those that meet the inclusion criteria described in the protocol) need to be subjected to rigorous appraisal by two critical appraisers. The results of this appraisal can then be used to inform synthesis and interpretation of the results of the study. JBI Critical appraisal tools have been developed by the JBI and collaborators and approved by the JBI Scientific Committee following extensive peer review. Although designed for use in systematic reviews, JBI critical appraisal tools can also be used when creating Critically Appraised Topics (CAT), in journal clubs and as an educational tool.

© JBI, 2020. All rights reserved. JBI grants use of these tools for research purposes only. All other enquiries should be sent to.

### JBI CRITICAL APPRAISAL CHECKLIST FOR ANALYTICAL CROSS SECTIONAL STUDIES

Reviewer: Daniel de la Rosa Martinez. Date: Martes 13 de Abril 2021

Author: Rossana Bussan, et al. Year: 2020 Record Number: 140_33158808_DDM

| | Yes | No | Unclear | Not applicable |
|---|---|---|---|---|
| 1. Were the criteria for inclusion in the sample clearly defined? | ■ | □ | □ | □ |
| 2. Were the study subjects and the setting described in detail? | ■ | □ | □ | □ |
| 3. Was the exposure measured in a valid and reliable way? | ■ | □ | □ | □ |
| 4. Were objective, standard criteria used for measurement of the condition? | ■ | □ | □ | □ |
| 5. Were confounding factors identified? | □ | ■ | □ | □ |
| 6. Were strategies to deal with confounding factors stated? | □ | ■ | □ | □ |
| 7. Were the outcomes measured in a valid and reliable way? | ■ | □ | □ | □ |
| 8. Was appropriate statistical analysis used? | □ | □ | ■ | □ |

Overall appraisal: Include □ Exclude ■ Seek further info □

Comments (Including reason for exclusion)

**No aporta información relevante para los fines de la revisión**

© JBI, 2020. All rights reserved. JBI grants use of these tools for research purposes only. All other enquiries should be sent to.

**Explanation of analytical cross sectional studies critical appraisal**

*How to cite:* Moola S, Munn Z, Tufanaru C, Aromataris E, Sears K, Sfetcu R, Currie M, Qureshi R, Mattis P, Lisy K, Mu P-F. Chapter 7: Systematic reviews of etiology and risk . In: Aromataris E, Munn Z (Editors). *JBI Manual for Evidence Synthesis*. JBI, 2020. Available from https://synthesismanual.jbi.global

**Analytical cross sectional studies Critical Appraisal Tool**

Answers: Yes, No, Unclear or Not/Applicable

#### 1. Were the criteria for inclusion in the sample clearly defined?

The authors should provide clear inclusion and exclusion criteria that they developed prior to recruitment of the study participants. The inclusion/exclusion criteria should be specified (e.g., risk, stage of disease progression) with sufficient detail and all the necessary information critical to the study.

#### 2. Were the study subjects and the setting described in detail?

The study sample should be described in sufficient detail so that other researchers can determine if it is comparable to the population of interest to them. The authors should provide a clear description of the population from which the study participants were selected or recruited, including demographics, location, and time period.

#### 3. Was the exposure measured in a valid and reliable way?

The study should clearly describe the method of measurement of exposure. Assessing validity requires that a 'gold standard' is available to which the measure can be compared. The validity of exposure measurement usually relates to whether a current measure is appropriate or whether a measure of past exposure is needed.

Reliability refers to the processes included in an epidemiological study to check repeatability of measurements of the exposures. These usually include intra-observer reliability and inter-observer reliability.

#### 4. Were objective, standard criteria used for measurement of the condition?

It is useful to determine if patients were included in the study based on either a specified diagnosis or definition. This is more likely to decrease the risk of bias. Characteristics are another useful approach to matching groups, and studies that did not use specified diagnostic methods or definitions should provide evidence on matching by key characteristics

#### 5. Were confounding factors identified?

Confounding has occurred where the estimated intervention exposure effect is biased by the presence of some difference between the comparison groups (apart from the exposure investigated/of interest). Typical confounders include baseline characteristics, prognostic factors, or concomitant exposures (e.g. smoking). A confounder is a difference between the comparison groups and it influences the direction of the study results. A high quality study at the level of cohort design will identify the potential confounders and measure them (where possible). This is difficult for studies where behavioral, attitudinal or lifestyle factors may impact on the results.

#### 6. Were strategies to deal with confounding factors stated?

Strategies to deal with effects of confounding factors may be dealt within the study design or in data analysis. By matching or stratifying sampling of participants, effects of confounding factors can be adjusted for. When dealing with adjustment in data analysis, assess the statistics used in the study. Most will be some form of multivariate regression analysis to account for the confounding factors measured.

#### 7. Were the outcomes measured in a valid and reliable way?

© JBI, 2020. All rights reserved. JBI grants use of these tools for research purposes only. All other enquiries should be sent to.

Read the methods section of the paper. If for e.g. lung cancer is assessed based on existing definitions or diagnostic criteria, then the answer to this question is likely to be yes. If lung cancer is assessed using observer reported, or self-reported scales, the risk of over- or under-reporting is increased, and objectivity is compromised. Importantly, determine if the measurement tools used were validated instruments as this has a significant impact on outcome assessment validity.

Having established the objectivity of the outcome measurement (e.g. lung cancer) instrument, it's important to establish how the measurement was conducted. Were those involved in collecting data trained or educated in the use of the instrument/s? (e.g. radiographers). If there was more than one data collector, were they similar in terms of level of education, clinical or research experience, or level of responsibility in the piece of research being appraised?

#### 8. Was appropriate statistical analysis used?

As with any consideration of statistical analysis, consideration should be given to whether there was a more appropriate alternate statistical method that could have been used. The methods section should be detailed enough for reviewers to identify which analytical techniques were used (in particular, regression or stratification) and how specific confounders were measured.

For studies utilizing regression analysis, it is useful to identify if the study identified which variables were included and how they related to the outcome. If stratification was the analytical approach used, were the strata of analysis defined by the specified variables? Additionally, it is also important to assess the appropriateness of the analytical strategy in terms of the assumptions associated with the approach as differing methods of analysis are based on differing assumptions about the data and how it will respond.

© JBI, 2020. All rights reserved. JBI grants use of these tools for research purposes only. All other enquiries should be sent to.

### JBI CRITICAL APPRAISAL CHECKLIST FOR TEXT AND OPINION PAPERS

Reviewer Marco Antonio Delaye Martinez Date 07/04/20

Author Falahi S., Kenarkoohi A. Year 2020 Record Number 141_33200033

| | Yes | No | Unclear | Not applicable |
|---|---|---|---|---|
| 1. Is the source of the opinion clearly identified? | ☑ | ☐ | ☐ | ☐ |
| 2. Does the source of opinion have standing in the field of expertise? | ☑ | ☐ | ☐ | ☐ |
| 3. Are the interests of the relevant population the central focus of the opinion? | ☑ | ☐ | ☐ | ☐ |
| 4. Is the stated position the result of an analytical process, and is there logic in the opinion expressed? | ☑ | ☐ | ☐ | ☐ |
| 5. Is there reference to the extant literature? | ☑ | ☐ | ☐ | ☐ |
| 6. Is any incongruence with the literature/sources logically defended? | ☑ | ☐ | ☐ | ☐ |

Overall appraisal: Include ☐ Exclude ☑ Seek further info ☐

Comments (Including reason for exclusion)

El artículo no aporta informacion util para los fines de la presente revisión sistemática.

© JBI, 2020. All rights reserved. JBI grants use of these tools for research purposes only. All other enquiries should be sent to.

### CHECKLIST FOR ANALYTICAL CROSS SECTIONAL STUDIES

Critical Appraisal tools for use in JBI Systematic Reviews

### INTRODUCTION

JBI is an JBI is an international research organisation based in the Faculty of Health and Medical Sciences at the University of Adelaide, South Australia. JBI develops and delivers unique evidence-based information, software, education and training designed to improve healthcare practice and health outcomes. With over 70 Collaborating Entities, servicing over 90 countries, JBI is a recognised global leader in evidence-based healthcare.

#### JBI Systematic Reviews

The core of evidence synthesis is the systematic review of literature of a particular intervention, condition or issue. The systematic review is essentially an analysis of the available literature (that is, evidence) and a judgment of the effectiveness or otherwise of a practice, involving a series of complex steps. JBI takes a particular view on what counts as evidence and the methods utilised to synthesise those different types of evidence. In line with this broader view of evidence, JBI has developed theories, methodologies and rigorous processes for the critical appraisal and synthesis of these diverse forms of evidence in order to aid in clinical decision-making in healthcare. There now exists JBI guidance for conducting reviews of effectiveness research, qualitative research, prevalence/incidence, etiology/risk, economic evaluations, text/opinion, diagnostic test accuracy, mixed-methods, umbrella reviews and scoping reviews. Further information regarding JBI systematic reviews can be found in the JBI Evidence Synthesis Manual.

#### JBI Critical Appraisal Tools

All systematic reviews incorporate a process of critique or appraisal of the research evidence. The purpose of this appraisal is to assess the methodological quality of a study and to determine the extent to which a study has addressed the possibility of bias in its design, conduct and analysis. All papers selected for inclusion in the systematic review (that is – those that meet the inclusion criteria described in the protocol) need to be subjected to rigorous appraisal by two critical appraisers. The results of this appraisal can then be used to inform synthesis and interpretation of the results of the study. JBI Critical appraisal tools have been developed by the JBI and collaborators and approved by the JBI Scientific Committee following extensive peer review. Although designed for use in systematic reviews, JBI critical appraisal tools can also be used when creating Critically Appraised Topics (CAT), in journal clubs and as an educational tool.

© JBI, 2020. All rights reserved. JBI grants use of these tools for research purposes only. All other enquiries should be sent to.

### JBI CRITICAL APPRAISAL CHECKLIST FOR ANALYTICAL CROSS SECTIONAL STUDIES

Reviewer: Daniel de la Rosa Martinez. Date: Martes 13 de Abril 2021

Author: Beatriz López-Barbeito. Year: 2020 Record Number: 142_33275358_DDM

| | Yes | No | Unclear | Not applicable |
|---|---|---|---|---|
| 1. Were the criteria for inclusion in the sample clearly defined? | ☐ | ■ | ☐ | ☐ |
| 2. Were the study subjects and the setting described in detail? | ☐ | ☐ | ■ | ☐ |
| 3. Was the exposure measured in a valid and reliable way? | ☐ | ■ | ☐ | ☐ |
| 4. Were objective, standard criteria used for measurement of the condition? | ☐ | ■ | ☐ | ☐ |
| 5. Were confounding factors identified? | ■ | ☐ | ☐ | ☐ |
| 6. Were strategies to deal with confounding factors stated? | ☐ | ■ | ☐ | ☐ |
| 7. Were the outcomes measured in a valid and reliable way? | ☐ | ■ | ☐ | ☐ |
| 8. Was appropriate statistical analysis used? | ☐ | ☐ | ■ | ☐ |

Overall appraisal: Include ☐ Exclude ■ Seek further info ☐

Comments (Including reason for exclusion)

El diagnostico no fue uniforme para todos y no queda claro el tiempo de seguimiento o si los sintomas son al momento de la revaloracion

© JBI, 2020. All rights reserved. JBI grants use of these tools for research purposes only. All other enquiries should be sent to.

**Analytical cross sectional studies Critical Appraisal Tool**

Answers: Yes, No, Unclear or Not/Applicable

#### 1. Were the criteria for inclusion in the sample clearly defined?

The authors should provide clear inclusion and exclusion criteria that they developed prior to recruitment of the study participants. The inclusion/exclusion criteria should be specified (e.g., risk, stage of disease progression) with sufficient detail and all the necessary information critical to the study.

#### 2. Were the study subjects and the setting described in detail?

The study sample should be described in sufficient detail so that other researchers can determine if it is comparable to the population of interest to them. The authors should provide a clear description of the population from which the study participants were selected or recruited, including demographics, location, and time period.

#### 3. Was the exposure measured in a valid and reliable way?

The study should clearly describe the method of measurement of exposure. Assessing validity requires that a 'gold standard' is available to which the measure can be compared. The validity of exposure measurement usually relates to whether a current measure is appropriate or whether a measure of past exposure is needed.

Reliability refers to the processes included in an epidemiological study to check repeatability of measurements of the exposures. These usually include intra-observer reliability and inter-observer reliability.

#### 4. Were objective, standard criteria used for measurement of the condition?

It is useful to determine if patients were included in the study based on either a specified diagnosis or definition. This is more likely to decrease the risk of bias. Characteristics are another useful approach to matching groups, and studies that did not use specified diagnostic methods or definitions should provide evidence on matching by key characteristics

#### 5. Were confounding factors identified?

Confounding has occurred where the estimated intervention exposure effect is biased by the presence of some difference between the comparison groups (apart from the exposure investigated/of interest). Typical confounders include baseline characteristics, prognostic factors, or concomitant exposures (e.g. smoking). A confounder is a difference between the comparison groups and it influences the direction of the study results. A high quality study at the level of cohort design will identify the potential confounders and measure them (where possible). This is difficult for studies where behavioral, attitudinal or lifestyle factors may impact on the results.

#### 6. Were strategies to deal with confounding factors stated?

Strategies to deal with effects of confounding factors may be dealt within the study design or in data analysis. By matching or stratifying sampling of participants, effects of confounding factors can be adjusted for. When dealing with adjustment in data analysis, assess the statistics used in the study. Most will be some form of multivariate regression analysis to account for the confounding factors measured.

#### 7. Were the outcomes measured in a valid and reliable way?

Read the methods section of the paper. If for e.g. lung cancer is assessed based on existing definitions or diagnostic criteria, then the answer to this question is likely to be yes. If lung cancer is assessed using observer reported, or self-reported scales, the risk of over- or under-reporting is increased, and objectivity is compromised. Importantly, determine if the measurement tools used were validated instruments as this has a significant impact on outcome assessment validity.

© JBI, 2020. All rights reserved. JBI grants use of these tools for research purposes only. All other enquiries should be sent to.

Having established the objectivity of the outcome measurement (e.g. lung cancer) instrument, it's important to establish how the measurement was conducted. Were those involved in collecting data trained or educated in the use of the instrument/s? (e.g. radiographers). If there was more than one data collector, were they similar in terms of level of education, clinical or research experience, or level of responsibility in the piece of research being appraised?

#### 8. Was appropriate statistical analysis used?

As with any consideration of statistical analysis, consideration should be given to whether there was a more appropriate alternate statistical method that could have been used. The methods section should be detailed enough for reviewers to identify which analytical techniques were used (in particular, regression or stratification) and how specific confounders were measured.

For studies utilizing regression analysis, it is useful to identify if the study identified which variables were included and how they related to the outcome. If stratification was the analytical approach used, were the strata of analysis defined by the specified variables? Additionally, it is also important to assess the appropriateness of the analytical strategy in terms of the assumptions associated with the approach as differing methods of analysis are based on differing assumptions about the data and how it will respond.

© JBI, 2020. All rights reserved. JBI grants use of these tools for research purposes only. All other enquiries should be sent to.

### JBI CRITICAL APPRAISAL CHECKLIST FOR TEXT AND OPINION PAPERS

Reviewer Daniel Eduardo Sandoval Colin Date 31/03/2021

Author Jonathan Wei Ting Wen LIU, Renata D'alloé DE LUCA, Heraldo Oliveira MELLO NETO and Igor BARCELLOS Year 2020 Record Number 143_33331469_DESC

| | Yes | No | Unclear | Not applicable |
|---|---|---|---|---|
| 1. Is the source of the opinion clearly identified? | ☒ | ☐ | ☐ | ☐ |
| 2. Does the source of opinion have standing in the field of expertise? | ☒ | ☐ | ☐ | ☐ |
| 3. Are the interests of the relevant population the central focus of the opinion? | ☒ | ☐ | ☐ | ☐ |
| 4. Is the stated position the result of an analytical process, and is there logic in the opinion expressed? | ☒ | ☐ | ☐ | ☐ |
| 5. Is there reference to the extant literature? | ☒ | ☐ | ☐ | ☐ |
| 6. Is any incongruence with the literature/sources logically defended? | ☐ | ☐ | ☐ | ☒ |

Overall appraisal: Include ☐ Exclude ☒ Seek further info ☐

Comments (Including reason for exclusion)

No cumple los criterios de inclusión. Sin embargo, puede ser de utilidad para la discusión.

### JBI CRITICAL APPRAISAL CHECKLIST FOR TEXT AND OPINION PAPERS

Reviewer Daniel Eduardo Sandoval Colin Date 31/03/2021

Author Ana P. Bouças, Jakeline Rheinheimer and Jim Lagopoulos Year 2020 Record Number 144_33135582_DESC

| | Yes | No | Unclear | Not applicable |
|---|---|---|---|---|
| 1. Is the source of the opinion clearly identified? | ☒ | ☐ | ☐ | ☐ |
| 2. Does the source of opinion have standing in the field of expertise? | ☒ | ☐ | ☐ | ☐ |
| 3. Are the interests of the relevant population the central focus of the opinion? | ☒ | ☐ | ☐ | ☐ |
| 4. Is the stated position the result of an analytical process, and is there logic in the opinion expressed? | ☒ | ☐ | ☐ | ☐ |
| 5. Is there reference to the extant literature? | ☒ | ☐ | ☐ | ☐ |
| 6. Is any incongruence with the literature/sources logically defended? | ☐ | ☐ | ☐ | ☒ |

Overall appraisal: Include ☐ Exclude ☒ Seek further info ☐

Comments (Including reason for exclusion)

No cumple los criterios de inclusión. Sin embargo, puede ser de utilidad para la discusión.

### JBI CRITICAL APPRAISAL CHECKLIST FOR TEXT AND OPINION PAPERS

Reviewer Daniel Eduardo Sandoval Colin Date 31/03/2021

Author Oliver O'Sullivan Year 2020 Record Number 145_33144403_DESC

| | Yes | No | Unclear | Not applicable |
|---|---|---|---|---|
| 1. Is the source of the opinion clearly identified? | ☒ | ☐ | ☐ | ☐ |
| 2. Does the source of opinion have standing in the field of expertise? | ☐ | ☐ | ☒ | ☐ |
| 3. Are the interests of the relevant population the central focus of the opinion? | ☒ | ☐ | ☐ | ☐ |
| 4. Is the stated position the result of an analytical process, and is there logic in the opinion expressed? | ☒ | ☐ | ☐ | ☐ |
| 5. Is there reference to the extant literature? | ☒ | ☐ | ☐ | ☐ |
| 6. Is any incongruence with the literature/sources logically defended? | ☐ | ☐ | ☐ | ☒ |

Overall appraisal: Include ☐ Exclude ☒ Seek further info ☐

Comments (including reason for exclusion)

No cumple los criterios de inclusión.

No es relevante para la discusión de este proyecto.

Concientiza sobre la necesidad de rehabilitación y evaluación multidisiplinaria después de la fase aguda.

### JBI Critical Appraisal Checklist for Case Series

Reviewer Daniel Eduardo Sandoval Colin Date 31/03/2021

Author Luigi Urciuoli and Elvira Guerriero Year 2020 Record Number 146_33152991_DESC

| | Yes | No | Unclear | Not applicable |
|---|---|---|---|---|
| Were there clear criteria for inclusion in the case series? | ☐ | ☐ | ☐ | ☐ |
| Was the condition measured in a standard, reliable way for all participants included in the case series? | ☐ | ☐ | ☐ | ☐ |
| Were valid methods used for identification of the condition for all participants included in the case series? | ☐ | ☐ | ☐ | ☐ |
| Did the case series have consecutive inclusion of participants? | ☐ | ☐ | ☐ | ☐ |
| Did the case series have complete inclusion of participants? | ☐ | ☐ | ☐ | ☐ |
| Was there clear reporting of the demographics of the participants in the study? | ☐ | ☐ | ☐ | ☐ |
| Was there clear reporting of clinical information of the participants? | ☐ | ☐ | ☐ | ☐ |
| Were the outcomes or follow up results of cases clearly reported? | ☐ | ☐ | ☐ | ☐ |
| Was there clear reporting of the presenting site(s)/clinic(s) demographic information? | ☐ | ☐ | ☐ | ☐ |
| Was statistical analysis appropriate? | ☐ | ☐ | ☐ | ☐ |

Overall appraisal: Include ☐ Exclude ☒ Seek further info ☐

Comments (Including reason for exclusion)

No se realiza cuestionario porque no cumple con los criterios de inclusión. No reportan persistencia de sintomas.

### JBI CRITICAL APPRAISAL CHECKLIST FOR TEXT AND OPINION PAPERS

Reviewer: Daniel Eduardo Sandoval Colin  Date: 31/03/2021

Author: Denyse D. Lutchmansingh, Melissa P. Knauert, Danielle E. Antin-Ozerkis et al.  Year: 2020  Record Number: 147_33159907_DESC

| | Yes | No | Unclear | Not applicable |
|---|---|---|---|---|
| 1. Is the source of the opinion clearly identified? | ☒ | ☐ | ☐ | ☐ |
| 2. Does the source of opinion have standing in the field of expertise? | ☒ | ☐ | ☐ | ☐ |
| 3. Are the interests of the relevant population the central focus of the opinion? | ☒ | ☐ | ☐ | ☐ |
| 4. Is the stated position the result of an analytical process, and is there logic in the opinion expressed? | ☒ | ☐ | ☐ | ☐ |
| 5. Is there reference to the extant literature? | ☒ | ☐ | ☐ | ☐ |
| 6. Is any incongruence with the literature/sources logically defended? | ☐ | ☐ | ☐ | ☒ |

Overall appraisal: Include ☐ Exclude ☒ Seek further info ☐

Comments (including reason for exclusion)

No cumple los criterios de inclusión.

Habla principalmente sobre manifestaciones agudas pulmonares y extrapulmonares y los posibles síntomas persistentes.

Aborda principalmente las seculas de la función ventilatoria y cambios radiográficos.

### JBI CRITICAL APPRAISAL CHECKLIST FOR ANALYTICAL CROSS SECTIONAL STUDIES

Reviewer Daniel Eduardo Sandoval Colin Date 31/03/2021

Author Mao Peng, Li Wang, Qing Xue et al. Year 2020 Record Number 148_33152729_DESC

| | Yes | No | Unclear | Not applicable |
|---|---|---|---|---|
| 1. Were the criteria for inclusion in the sample clearly defined? | ☐ | ☐ | ☐ | ☐ |
| 2. Were the study subjects and the setting described in detail? | ☐ | ☐ | ☐ | ☐ |
| 3. Was the exposure measured in a valid and reliable way? | ☐ | ☐ | ☐ | ☐ |
| 4. Were objective, standard criteria used for measurement of the condition? | ☐ | ☐ | ☐ | ☐ |
| 5. Were confounding factors identified? | ☐ | ☐ | ☐ | ☐ |
| 6. Were strategies to deal with confounding factors stated? | ☐ | ☐ | ☐ | ☐ |
| 7. Were the outcomes measured in a valid and reliable way? | ☐ | ☐ | ☐ | ☐ |
| 8. Was appropriate statistical analysis used? | ☐ | ☐ | ☐ | ☐ |

Overall appraisal: Include ☐ Exclude ☒ Seek further info ☐

Comments (Including reason for exclusion)

No se realiza cuestionario porque no cumple con los criterios de inclusión.

No aborda síntomas persistentes.

Encuesta en linea para estudiar la prevalencia de carga alostatica en trabajadores médicos vs trabajadores no médicos.

### JBI Critical Appraisal Checklist for Case Series

Reviewer Daniel Eduardo Sandoval Colin Date 31/03/2021

Author Yale Tung-Chen, Milagros Martí de Gracia, María Luz Parra-Gordo et al Year 2020 Record Number 149_33159704_DESC

| | Yes | No | Unclear | Not applicable |
|---|---|---|---|---|
| Were there clear criteria for inclusion in the case series? | ☐ | ☐ | ☐ | ☐ |
| Was the condition measured in a standard, reliable way for all participants included in the case series? | ☐ | ☐ | ☐ | ☐ |
| Were valid methods used for identification of the condition for all participants included in the case series? | ☐ | ☐ | ☐ | ☐ |
| Did the case series have consecutive inclusion of participants? | ☐ | ☐ | ☐ | ☐ |
| Did the case series have complete inclusion of participants? | ☐ | ☐ | ☐ | ☐ |
| Was there clear reporting of the demographics of the participants in the study? | ☐ | ☐ | ☐ | ☐ |
| Was there clear reporting of clinical information of the participants? | ☐ | ☐ | ☐ | ☐ |
| Were the outcomes or follow up results of cases clearly reported? | ☐ | ☐ | ☐ | ☐ |
| Was there clear reporting of the presenting site(s)/clinic(s) demographic information? | ☐ | ☐ | ☐ | ☐ |
| Was statistical analysis appropriate? | ☐ | ☐ | ☐ | ☐ |

Overall appraisal: Include ☐ Exclude ☒ Seek further info ☐

Comments (Including reason for exclusion)

No cumple los criterios de inclusión. Sin embargo, puede ser de utilidad para la discusión.

No se abordan síntomas persistentes. Este estudio reporta la posible utilidad del ultrasonido para el seguimiento de lesiones pulmonares.

### JBI CRITICAL APPRAISAL CHECKLIST FOR ANALYTICAL CROSS SECTIONAL STUDIES

Reviewer Daniel Eduardo Sandoval Colin Date 1/04/2021

Author Liam Townsend, Adam H. Dyer, Karen Jones et al. Year 2020 Record Number 150_33166287_DESC

| | Yes | No | Unclear | Not applicable |
|---|---|---|---|---|
| 1. Were the criteria for inclusion in the sample clearly defined? | ☒ | ☐ | ☐ | ☐ |
| 2. Were the study subjects and the setting described in detail? | ☒ | ☐ | ☐ | ☐ |
| 3. Was the exposure measured in a valid and reliable way? | ☒ | ☐ | ☐ | ☐ |
| 4. Were objective, standard criteria used for measurement of the condition? | ☒ | ☐ | ☐ | ☐ |
| 5. Were confounding factors identified? | ☒ | ☐ | ☐ | ☐ |
| 6. Were strategies to deal with confounding factors stated? | ☐ | ☐ | ☒ | ☐ |
| 7. Were the outcomes measured in a valid and reliable way? | ☒ | ☐ | ☐ | ☐ |
| 8. Was appropriate statistical analysis used? | ☒ | ☐ | ☐ | ☐ |

Overall appraisal: Include ☒ Exclude ☐ Seek further info ☐

Comments (Including reason for exclusion)

### JBI CRITICAL APPRAISAL CHECKLIST FOR TEXT AND OPINION PAPERS

Reviewer Daniel Eduardo Sandoval Colin Date 1/04/2020

Author Jose Vergara, Camila Lirani-Silva, Martin B. Brodsky et al. Year 2020 Record Number 151_33167752_DESC

| | Yes | No | Unclear | Not applicable |
|---|---|---|---|---|
| 1. Is the source of the opinion clearly identified? | ☒ | ☐ | ☐ | ☐ |
| 2. Does the source of opinion have standing in the field of expertise? | ☒ | ☐ | ☐ | ☐ |
| 3. Are the interests of the relevant population the central focus of the opinion? | ☒ | ☐ | ☐ | ☐ |
| 4. Is the stated position the result of an analytical process, and is there logic in the opinion expressed? | ☒ | ☐ | ☐ | ☐ |
| 5. Is there reference to the extant literature? | ☒ | ☐ | ☐ | ☐ |
| 6. Is any incongruence with the literature/sources logically defended? | ☐ | ☐ | ☐ | ☒ |

Overall appraisal: Include ☐ Exclude ☒ Seek further info ☐

Comments (Including reason for exclusion)

No cumple los criterios de inclusión.

Habla sobre las posibles alteraciones neurológicas que pueden interferir en la deglución, con efecto no solo en el gusto y el olfato, sino también en la función sensoriomotora de la faringe y la laringe, que pueden comprometer la protección de las vías respiratorias y la seguridad de la deglución.

### JBI CRITICAL APPRAISAL CHECKLIST FOR ANALYTICAL CROSS SECTIONAL STUDIES

Reviewer: Daniel Eduardo Sandoval Colin    Date: 29/03/2021

Author: Swapna Mandal, Joseph Barnett, Simon E Brill et al.    Year: 2020    Record Number: 152_33172844_DESC

| | Yes | No | Unclear | Not applicable |
|---|---|---|---|---|
| 1. Were the criteria for inclusion in the sample clearly defined? | X | | | |
| 2. Were the study subjects and the setting described in detail? | X | | | |
| 3. Was the exposure measured in a valid and reliable way? | X | | | |
| 4. Were objective, standard criteria used for measurement of the condition? | X | | | |
| 5. Were confounding factors identified? | X | | | |
| 6. Were strategies to deal with confounding factors stated? | | X | | |
| 7. Were the outcomes measured in a valid and reliable way? | X | | | |
| 8. Was appropriate statistical analysis used? | | X | | |

Overall appraisal: Include ☐ Exclude ☒ Seek further info ☐

Comments (Including reason for exclusion)

### JBI CRITICAL APPRAISAL CHECKLIST FOR TEXT AND OPINION PAPERS

Reviewer Daniel Eduardo Sandoval Colin Date 29/03/2021

Author Peter V. Dicpinigaitis and Brendan J. Canning Year 2020 Record Number 153_33188436_DESC

| | Yes | No | Unclear | Not applicable |
|---|---|---|---|---|
| 1. Is the source of the opinion clearly identified? | ☒ | ☐ | ☐ | ☐ |
| 2. Does the source of opinion have standing in the field of expertise? | ☒ | ☐ | ☐ | ☐ |
| 3. Are the interests of the relevant population the central focus of the opinion? | ☒ | ☐ | ☐ | ☐ |
| 4. Is the stated position the result of an analytical process, and is there logic in the opinion expressed? | ☒ | ☐ | ☐ | ☐ |
| 5. Is there reference to the extant literature? | ☒ | ☐ | ☐ | ☐ |
| 6. Is any incongruence with the literature/sources logically defended? | ☒ | ☐ | ☐ | ☐ |

Overall appraisal: Include ☐ Exclude ☒ Seek further info ☐

Comments (including reason for exclusion)

No posee datos para los propósitos de este trabajo, puede ser de utilidad para la discusión del manuscrito.

### JBI CRITICAL APPRAISAL CHECKLIST FOR ANALYTICAL CROSS SECTIONAL STUDIES

Reviewer: Daniel Eduardo Sandoval Colin    Date: 29/03/2021

Author: Tadayuki Oshima, Kewin Tien Ho Siah, Takanori Yoshimoto et al.    Year: 2020    Record Number: 154_33197076_DESC

| | Yes | No | Unclear | Not applicable |
|---|---|---|---|---|
| 1. Were the criteria for inclusion in the sample clearly defined? | X | | | |
| 2. Were the study subjects and the setting described in detail? | X | | | |
| 3. Was the exposure measured in a valid and reliable way? | X | | | |
| 4. Were objective, standard criteria used for measurement of the condition? | X | | | |
| 5. Were confounding factors identified? | X | | | |
| 6. Were strategies to deal with confounding factors stated? | X | | | |
| 7. Were the outcomes measured in a valid and reliable way? | | X | | |
| 8. Was appropriate statistical analysis used? | X | | | |

Overall appraisal: Include ☐ Exclude ☒ Seek further info ☐

Comments (Including reason for exclusion)

No se estudia a sujetos con diagnóstico previo de COVID-19

### JBI CRITICAL APPRAISAL CHECKLIST FOR COHORT STUDIES

Reviewer: Daniel Eduardo Sandoval Colin  Date: 30/03/2021

Author: Mariarosa Carta, Luciana Bragagnolo, Andrea Tramarin et al.  Year: 2020  Record Number: 155_32827394_DESC

| | Yes | No | Unclear | Not applicable |
|---|---|---|---|---|
| 1. Were the two groups similar and recruited from the same population? | ☐ | ☐ | ☐ | ☐ |
| 2. Were the exposures measured similarly to assign people<br>3. to both exposed and unexposed groups? | ☐ | ☐ | ☐ | ☐ |
| 4. Was the exposure measured in a valid and reliable way? | ☐ | ☐ | ☐ | ☐ |
| 5. Were confounding factors identified? | ☐ | ☐ | ☐ | ☐ |
| 6. Were strategies to deal with confounding factors stated? | ☐ | ☐ | ☐ | ☐ |
| 7. Were the groups/participants free of the outcome at the start of the study (or at the moment of exposure)? | ☐ | ☐ | ☐ | ☐ |
| 8. Were the outcomes measured in a valid and reliable way? | ☐ | ☐ | ☐ | ☐ |
| 9. Was the follow up time reported and sufficient to be long enough for outcomes to occur? | ☐ | ☐ | ☐ | ☐ |
| 10. Was follow up complete, and if not, were the reasons to loss to follow up described and explored? | ☐ | ☐ | ☐ | ☐ |
| 11. Were strategies to address incomplete follow up utilized? | ☐ | ☐ | ☐ | ☐ |
| 12. Was appropriate statistical analysis used? | ☐ | ☐ | ☐ | ☐ |

Overall appraisal: Include ☐ Exclude ☒ Seek further info ☐

Comments (Including reason for exclusion)

No se realiza cuestionario porque no cumple con los criterios de inclusión.

No investiga síntomas postdiagnóstico de COVID-19. Se evalúa positividad a 20, 32 y 49 días con RT-PCR, además de IgM-Ab e IgG-Ab en un asilo para adultos mayores.

### JBI CRITICAL APPRAISAL CHECKLIST FOR COHORT STUDIES

Reviewer Daniel Eduardo Sandoval Colin Date 30/03/2021

Author Gloria Gambini, Maria Cristina Savastano, Alfonso Savastano et al. Year 2021 Record Number 156_33214412_DESC

| | Yes | No | Unclear | Not applicable |
|---|---|---|---|---|
| 1. Were the two groups similar and recruited from the same population? | ☒ | ☐ | ☐ | ☐ |
| 2. Were the exposures measured similarly to assign people<br>3. to both exposed and unexposed groups? | ☒ | ☐ | ☐ | ☐ |
| 4. Was the exposure measured in a valid and reliable way? | ☒ | ☐ | ☐ | ☐ |
| 5. Were confounding factors identified? | ☒ | ☐ | ☐ | ☐ |
| 6. Were strategies to deal with confounding factors stated? | ☐ | ☒ | ☐ | ☐ |
| 7. Were the groups/participants free of the outcome at the start of the study (or at the moment of exposure)? | ☒ | ☐ | ☐ | ☐ |
| 8. Were the outcomes measured in a valid and reliable way? | ☒ | ☐ | ☐ | ☐ |
| 9. Was the follow up time reported and sufficient to be long enough for outcomes to occur? | ☒ | ☐ | ☐ | ☐ |
| 10. Was follow up complete, and if not, were the reasons to loss to follow up described and explored? | ☒ | ☐ | ☐ | ☐ |
| 11. Were strategies to address incomplete follow up utilized? | ☐ | ☐ | ☐ | ☒ |
| 12. Was appropriate statistical analysis used? | ☐ | ☒ | ☐ | ☐ |

Overall appraisal: Include ☒ Exclude ☐ Seek further info ☐

Comments (Including reason for exclusion)

______________________________________________

______________________________________________

### JBI CRITICAL APPRAISAL CHECKLIST FOR COHORT STUDIES

Reviewer Daniel Eduardo Sandoval Colin Date 30/03/2021

Author Fran A. Ganz-Lord, Kathryn R. Segal BA and Michael L. Rinke Year 2020 Record Number 157_33213542_DESC

| | Yes | No | Unclear | Not applicable |
|---|---|---|---|---|
| 1. Were the two groups similar and recruited from the same population? | ☐ | ☐ | ☐ | ☐ |
| 2. Were the exposures measured similarly to assign people<br>3. to both exposed and unexposed groups? | ☐ | ☐ | ☐ | ☐ |
| 4. Was the exposure measured in a valid and reliable way? | ☐ | ☐ | ☐ | ☐ |
| 5. Were confounding factors identified? | ☐ | ☐ | ☐ | ☐ |
| 6. Were strategies to deal with confounding factors stated? | ☐ | ☐ | ☐ | ☐ |
| 7. Were the groups/participants free of the outcome at the start of the study (or at the moment of exposure)? | ☐ | ☐ | ☐ | ☐ |
| 8. Were the outcomes measured in a valid and reliable way? | ☐ | ☐ | ☐ | ☐ |
| 9. Was the follow up time reported and sufficient to be long enough for outcomes to occur? | ☐ | ☐ | ☐ | ☐ |
| 10. Was follow up complete, and if not, were the reasons to loss to follow up described and explored? | ☐ | ☐ | ☐ | ☐ |
| 11. Were strategies to address incomplete follow up utilized? | ☐ | ☐ | ☐ | ☐ |
| 12. Was appropriate statistical analysis used? | ☐ | ☐ | ☐ | ☐ |

Overall appraisal: Include ☐ Exclude ☒ Seek further info ☐

Comments (Including reason for exclusion)

No se realiza cuestionario porque no cumple con los criterios de inclusión.

Las pruebas de PCR fueron inicialmente limitadas debido a la disponibilidad de hisopos y reactivos

En primera instancia la captura de sintomas estaba limitada a 4, posteriormente a 12. La captura de los sintomas esta limitada a la fase aguda.

### JBI CRITICAL APPRAISAL CHECKLIST FOR COHORT STUDIES

Reviewer Daniel Eduardo Sandoval Colin Date 30/03/2021

Author Bram van der Borst, Jeannette B Peters, Monique Brink et al. Year 2020 Record Number 158_33220049_DESC

| | Yes | No | Unclear | Not applicable |
|---|---|---|---|---|
| 1. Were the two groups similar and recruited from the same population? | ☒ | ☐ | ☐ | ☐ |
| 2. Were the exposures measured similarly to assign people<br>3. to both exposed and unexposed groups? | ☒ | ☐ | ☐ | ☐ |
| 4. Was the exposure measured in a valid and reliable way? | ☒ | ☐ | ☐ | ☐ |
| 5. Were confounding factors identified? | ☒ | ☐ | ☐ | ☐ |
| 6. Were strategies to deal with confounding factors stated? | ☐ | ☐ | ☒ | ☐ |
| 7. Were the groups/participants free of the outcome at the start of the study (or at the moment of exposure)? | ☐ | ☐ | ☒ | ☐ |
| 8. Were the outcomes measured in a valid and reliable way? | ☒ | ☐ | ☐ | ☐ |
| 9. Was the follow up time reported and sufficient to be long enough for outcomes to occur? | ☒ | ☐ | ☐ | ☐ |
| 10. Was follow up complete, and if not, were the reasons to loss to follow up described and explored? | ☒ | ☐ | ☐ | ☐ |
| 11. Were strategies to address incomplete follow up utilized? | ☐ | ☒ | ☐ | ☐ |
| 12. Was appropriate statistical analysis used? | ☒ | ☐ | ☐ | ☐ |

Overall appraisal: Include ☐ Exclude ☒ Seek further info ☐

Comments (Including reason for exclusion)

_______________________________________________

_______________________________________________

### JBI CRITICAL APPRAISAL CHECKLIST FOR ANALYTICAL CROSS SECTIONAL STUDIES

Reviewer Daniel Eduardo Sandoval Colin Date 30/03/2021

Author Marcel S.Woo, Jakob Malsy, Jana Pöttgen et al. Year 2020 Record Number 159_33376990_DESC

| | Yes | No | Unclear | Not applicable |
|---|---|---|---|---|
| 1. Were the criteria for inclusion in the sample clearly defined? | X | | | |
| 2. Were the study subjects and the setting described in detail? | X | | | |
| 3. Was the exposure measured in a valid and reliable way? | X | | | |
| 4. Were objective, standard criteria used for measurement of the condition? | X | | | |
| 5. Were confounding factors identified? | X | | | |
| 6. Were strategies to deal with confounding factors stated? | X | | | |
| 7. Were the outcomes measured in a valid and reliable way? | X | | | |
| 8. Was appropriate statistical analysis used? | X | | | |

Overall appraisal: Include ☐ Exclude ☒ Seek further info ☐

Comments (Including reason for exclusion)

### JBI CRITICAL APPRAISAL CHECKLIST FOR TEXT AND OPINION PAPERS

Reviewer Daniel Eduardo Sandoval Colin Date 30/03/2021

Author M Mendelson, J Nel, L Blumberg et al. Year 2020 Record Number 160_33403997_DESC

| | Yes | No | Unclear | Not applicable |
|---|---|---|---|---|
| 1. Is the source of the opinion clearly identified? | X | | | |
| 2. Does the source of opinion have standing in the field of expertise? | X | | | |
| 3. Are the interests of the relevant population the central focus of the opinion? | X | | | |
| 4. Is the stated position the result of an analytical process, and is there logic in the opinion expressed? | X | | | |
| 5. Is there reference to the extant literature? | X | | | |
| 6. Is any incongruence with the literature/sources logically defended? | X | | | |

Overall appraisal: Include ☐ Exclude ☒ Seek further info ☐

Comments (Including reason for exclusion)

No posee información para los propósitos de este trabajo. No cumple con los criterios de inclusión.

### JBI CRITICAL APPRAISAL CHECKLIST FOR COHORT STUDIES

Reviewer Daniel Eduardo Sandoval Colin Date 30/03/2021

Author Jerome R. Lechien, Fabrice Journe, Stephane Hans et al. Year 2020 Record Number 161_33330539_DESC

| | Yes | No | Unclear | Not applicable |
|---|---|---|---|---|
| 1. Were the two groups similar and recruited from the same population? | ☐ | ☐ | ☐ | ☒ |
| 2. Were the exposures measured similarly to assign people<br>3. to both exposed and unexposed groups? | ☐ | ☐ | ☐ | ☒ |
| 4. Was the exposure measured in a valid and reliable way? | ☒ | ☐ | ☐ | ☐ |
| 5. Were confounding factors identified? | ☒ | ☐ | ☐ | ☐ |
| 6. Were strategies to deal with confounding factors stated? | ☒ | ☐ | ☐ | ☐ |
| 7. Were the groups/participants free of the outcome at the start of the study (or at the moment of exposure)? | ☐ | ☐ | ☐ | ☒ |
| 8. Were the outcomes measured in a valid and reliable way? | ☒ | ☐ | ☐ | ☐ |
| 9. Was the follow up time reported and sufficient to be long enough for outcomes to occur? | ☒ | ☐ | ☐ | ☐ |
| 10. Was follow up complete, and if not, were the reasons to loss to follow up described and explored? | ☐ | ☐ | ☐ | ☒ |
| 11. Were strategies to address incomplete follow up utilized? | ☐ | ☐ | ☐ | ☒ |
| 12. Was appropriate statistical analysis used? | ☒ | ☐ | ☐ | ☐ |

Overall appraisal: Include ☐ Exclude ☒ Seek further info ☐

Comments (Including reason for exclusion)

### JBI CRITICAL APPRAISAL CHECKLIST FOR SYSTEMATIC REVIEWS AND RESEARCH SYNTHESES

Reviewer: Daniel de la Rosa  Date: 21-11-20

Author: R. Torres-Castroa et al  Year: 2020  Record Number: 162_33262076

| | | Yes | No | Unclear | Not applicable |
|---|---|---|---|---|---|
| 1. | Is the review question clearly and explicitly stated? | ☒ | ☐ | ☐ | ☐ |
| 2. | Were the inclusion criteria appropriate for the review question? | ☐ | ☐ | ☐ | ☒ |
| 3. | Was the search strategy appropriate? | ☐ | ☐ | ☐ | ☒ |
| 4. | Were the sources and resources used to search for studies adequate? | ☐ | ☐ | ☐ | ☒ |
| 5. | Were the criteria for appraising studies appropriate? | ☐ | ☐ | ☐ | ☒ |
| 6. | Was critical appraisal conducted by two or more reviewers independently? | ☐ | ☐ | ☐ | ☒ |
| 7. | Were there methods to minimize errors in data extraction? | ☐ | ☐ | ☐ | ☒ |
| 8. | Were the methods used to combine studies appropriate? | ☐ | ☐ | ☐ | ☒ |
| 9. | Was the likelihood of publication bias assessed? | ☐ | ☐ | ☐ | ☒ |
| 10. | Were recommendations for policy and/or practice supported by the reported data? | ☒ | ☐ | ☐ | ☐ |
| 11. | Were the specific directives for new research appropriate? | ☒ | ☐ | ☐ | ☐ |

Overall appraisal: Include ☐ Exclude ☒ Seek further info ☐

Comments (Including reason for exclusion)

Review

#### JBI Critical Appraisal Checklist for Case Series

Reviewer: Daniel de la Rosa Date: 02-02-21

Author: Melanie Dani et al Year: 2020 Record Number: 163_33243837

| | Yes | No | Unclear | Not applicable |
|---|---|---|---|---|
| Were there clear criteria for inclusion in the case series? | ☑ | ☐ | ☐ | ☐ |
| Was the condition measured in a standard, reliable way for all participants included in the case series? | ☑ | ☐ | ☐ | ☐ |
| Were valid methods used for identification of the condition for all participants included in the case series? | ☑ | ☐ | ☐ | ☐ |
| Did the case series have consecutive inclusion of participants? | ☐ | ☐ | ☑ | ☐ |
| Did the case series have complete inclusion of participants? | ☐ | ☑ | ☐ | ☐ |
| Was there clear reporting of the demographics of the participants in the study? | ☐ | ☑ | ☐ | ☐ |
| Was there clear reporting of clinical information of the participants? | ☑ | ☐ | ☐ | ☐ |
| Were the outcomes or follow up results of cases clearly reported? | ☑ | ☐ | ☐ | ☐ |
| Was there clear reporting of the presenting site(s)/clinic(s) demographic information? | ☑ | ☐ | ☐ | ☐ |
| Was statistical analysis appropriate? | ☐ | ☐ | ☐ | ☑ |

Overall appraisal: Include ☐ Exclude ☑ Seek further info ☐

Comments (Including reason for exclusion)

Cases report

© JBI, 2020. All rights reserved. JBI grants use of these tools for research purposes only. All other enquiries should be sent to.

#### JBI Critical Appraisal Checklist for Case Series

Reviewer: Daniel de la Rosa    Date: 02-02-21

Author: Edward Needham et all    Year: 2020    Record Number: 164_33244712

| | Yes | No | Unclear | Not applicable |
|---|---|---|---|---|
| • Were there clear criteria for inclusion in the case series? | ☑ | ☐ | ☐ | ☐ |
| • Was the condition measured in a standard, reliable way for all participants included in the case series? | ☑ | ☐ | ☐ | ☐ |
| • Were valid methods used for identification of the condition for all participants included in the case series? | ☑ | ☐ | ☐ | ☐ |
| • Did the case series have consecutive inclusion of participants? | ☐ | ☐ | ☑ | ☐ |
| • Did the case series have complete inclusion of participants? | ☑ | ☐ | ☐ | ☐ |
| • Was there clear reporting of the demographics of the participants in the study? | ☐ | ☑ | ☐ | ☐ |
| • Was there clear reporting of clinical information of the participants? | ☐ | ☑ | ☐ | ☐ |
| • Were the outcomes or follow up results of cases clearly reported? | ☑ | ☐ | ☐ | ☐ |
| • Was there clear reporting of the presenting site(s)/clinic(s) demographic information? | ☐ | ☑ | ☐ | ☐ |
| • Was statistical analysis appropriate? | ☐ | ☐ | ☐ | ☑ |

Overall appraisal: Include ☐ Exclude ☑ Seek further info ☐

Comments (Including reason for exclusion)

Cases report

© JBI, 2020. All rights reserved. JBI grants use of these tools for research purposes only. All other enquiries should be sent to.

### JBI CRITICAL APPRAISAL CHECKLIST FOR SYSTEMATIC REVIEWS AND RESEARCH SYNTHESES

Reviewer Daniel de la Rosa Date 02-02-21

Author Elham Atabati, et al. Year 2020 Record Number 165_33274259

| | Yes | No | Unclear | Not applicable |
|---|---|---|---|---|
| 1. Is the review question clearly and explicitly stated? | ☐ | ☐ | X | ☐ |
| 2. Were the inclusion criteria appropriate for the review question? | ☐ | ☐ | ☐ | X |
| 3. Was the search strategy appropriate? | ☐ | ☐ | ☐ | X |
| 4. Were the sources and resources used to search for studies adequate? | ☐ | ☐ | ☐ | X |
| 5. Were the criteria for appraising studies appropriate? | ☐ | ☐ | ☐ | X |
| 6. Was critical appraisal conducted by two or more reviewers independently? | ☐ | ☐ | ☐ | X |
| 7. Were there methods to minimize errors in data extraction? | ☐ | ☐ | ☐ | X |
| 8. Were the methods used to combine studies appropriate? | ☐ | ☐ | ☐ | X |
| 9. Was the likelihood of publication bias assessed? | ☐ | ☐ | ☐ | X |
| 10. Were recommendations for policy and/or practice supported by the reported data? | X | ☐ | ☐ | ☐ |
| 11. Were the specific directives for new research appropriate? | X | ☐ | ☐ | ☐ |

Overall appraisal: Include ☐ Exclude X Seek further info ☐

Comments (Including reason for exclusion)

### JBI CRITICAL APPRAISAL CHECKLIST FOR SYSTEMATIC REVIEWS AND RESEARCH SYNTHESES

Reviewer Daniel de la Rosa Date 21-11-20

Author Scott Rooney, et al Year 2020 Record Number 21_32737507

| | | Yes | No | Unclear | Not applicable |
|---|---|---|---|---|---|
| 1. | Is the review question clearly and explicitly stated? | ☒ | ☐ | ☐ | ☐ |
| 2. | Were the inclusion criteria appropriate for the review question? | ☐ | ☐ | ☐ | ☒ |
| 3. | Was the search strategy appropriate? | ☐ | ☐ | ☐ | ☒ |
| 4. | Were the sources and resources used to search for studies adequate? | ☐ | ☐ | ☐ | ☒ |
| 5. | Were the criteria for appraising studies appropriate? | ☐ | ☐ | ☐ | ☒ |
| 6. | Was critical appraisal conducted by two or more reviewers independently? | ☐ | ☐ | ☐ | ☒ |
| 7. | Were there methods to minimize errors in data extraction? | ☐ | ☐ | ☐ | ☒ |
| 8. | Were the methods used to combine studies appropriate? | ☐ | ☐ | ☐ | ☒ |
| 9. | Was the likelihood of publication bias assessed? | ☐ | ☐ | ☐ | ☒ |
| 10. | Were recommendations for policy and/or practice supported by the reported data? | ☒ | ☐ | ☐ | ☐ |
| 11. | Were the specific directives for new research appropriate? | ☒ | ☐ | ☐ | ☐ |

Overall appraisal: Include ☐ Exclude ☒ Seek further info ☐

Comments (Including reason for exclusion)

Review

### JBI CRITICAL APPRAISAL CHECKLIST FOR TEXT AND OPINION PAPERS

ARTICLE IN FRENCH - IT WAS NOT EVALUATED

Reviewer Daniel de la Rosa Date 14/11/20

Author C. Andrejaka et al Year 2020 Record Number 167_33280941

| | Yes | No | Unclear | Not applicable |
|---|---|---|---|---|
| 1. Is the source of the opinion clearly identified? | ☐ | ☐ | ☐ | ☐ |
| 2. Does the source of opinion have standing in the field of expertise? | ☐ | ☐ | ☐ | ☐ |
| 3. Are the interests of the relevant population the central focus of the opinion? | ☐ | ☐ | ☐ | ☐ |
| 4. Is the stated position the result of an analytical process, and is there logic in the opinion expressed? | ☐ | ☐ | ☐ | ☐ |
| 5. Is there reference to the extant literature? | ☐ | ☐ | ☐ | ☐ |
| 6. Is any incongruence with the literature/sources logically defended? | ☐ | ☐ | ☐ | ☐ |

Overall appraisal: Include ☐ Exclude ☐ Seek further info ☐

Comments (Including reason for exclusion)

ARTICLE IN FRENCH - IT WAS NOT EVALUATED

© JBI, 2020. All rights reserved. JBI grants use of these tools for research purposes only. All other enquiries should be sent to.

### JBI CRITICAL APPRAISAL CHECKLIST FOR ANALYTICAL CROSS SECTIONAL STUDIES

Reviewer Carlos Alberto Fermín Martínez Date 11/04/2021

Author Walsh-Messinger J, et al. Year 2020 Record Number 168_33269366_CAFM

| | Yes | No | Unclear | Not applicable |
|---|---|---|---|---|
| 1. Were the criteria for inclusion in the sample clearly defined? | ☑ | ☐ | ☐ | ☐ |
| 2. Were the study subjects and the setting described in detail? | ☑ | ☐ | ☐ | ☐ |
| 3. Was the exposure measured in a valid and reliable way? | ☐ | ☐ | ☑ | ☐ |
| 4. Were objective, standard criteria used for measurement of the condition? | ☑ | ☐ | ☐ | ☐ |
| 5. Were confounding factors identified? | ☐ | ☑ | ☐ | ☐ |
| 6. Were strategies to deal with confounding factors stated? | ☐ | ☑ | ☐ | ☐ |
| 7. Were the outcomes measured in a valid and reliable way? | ☑ | ☐ | ☐ | ☐ |
| 8. Was appropriate statistical analysis used? | ☑ | ☐ | ☐ | ☐ |

Overall appraisal: Include ☐ Exclude ☐ Seek further info ☑

Comments (Including reason for exclusion)

La metodología de este artículo es buena, el único inconveniente es que el diagnóstico de COVID-19 se hizo con PCR o con juicio clínico de un médico (25% de los pacientes). Se podría contactar a los autores para que provean información solo de los pacientes que tuvieron PCR positiva.

© JBI, 2020. All rights reserved. JBI grants use of these tools for research purposes only. All other enquiries should be sent to.

### JBI CRITICAL APPRAISAL CHECKLIST FOR COHORT STUDIES

Reviewer Carlos Alberto Fermin Martinez Date 11/04/2021

Author Petersen MS, et al. Year 2020 Record Number 169_33252665_CAFM

| | Yes | No | Unclear | Not applicable |
|---|---|---|---|---|
| 1. Were the two groups similar and recruited from the same population? | ☑ | ☐ | ☐ | ☐ |
| 2. Were the exposures measured similarly to assign people<br>3. to both exposed and unexposed groups? | ☑ | ☐ | ☐ | ☐ |
| 4. Was the exposure measured in a valid and reliable way? | ☑ | ☐ | ☐ | ☐ |
| 5. Were confounding factors identified? | ☐ | ☑ | ☐ | ☐ |
| 6. Were strategies to deal with confounding factors stated? | ☐ | ☑ | ☐ | ☐ |
| 7. Were the groups/participants free of the outcome at the start of the study (or at the moment of exposure)? | ☑ | ☐ | ☐ | ☐ |
| 8. Were the outcomes measured in a valid and reliable way? | ☑ | ☐ | ☐ | ☐ |
| 9. Was the follow up time reported and sufficient to be long enough for outcomes to occur? | ☑ | ☐ | ☐ | ☐ |
| 10. Was follow up complete, and if not, were the reasons to loss to follow up described and explored? | ☑ | ☐ | ☐ | ☐ |
| 11. Were strategies to address incomplete follow up utilized? | ☐ | ☐ | ☑ | ☐ |
| 12. Was appropriate statistical analysis used? | ☑ | ☐ | ☐ | ☐ |

Overall appraisal: Include ☐ Exclude ☐ Seek further info ☑

Comments (Including reason for exclusion)

En el estudio se incluyeron menores de edad y pacientes asintomáticos en la infección aguda, por lo que no cumple con los criterios de inclusión establecidos en nuestro protocolo. Se podría contactar a los autores para excluir a estos grupos de pacientes.

© JBI, 2020. All rights reserved. JBI grants use of these tools for research purposes only. All other enquiries should be sent to.

### JBI CRITICAL APPRAISAL CHECKLIST FOR COHORT STUDIES

Reviewer Carlos Alberto Fermin Martinez Date 11/04/2021

Author Deisenhammer F, et al. Year 2020 Record Number 170_33315138_CAFM

| | Yes | No | Unclear | Not applicable |
|---|---|---|---|---|
| 1. Were the two groups similar and recruited from the same population? | ☑ | ☐ | ☐ | ☐ |
| 2. Were the exposures measured similarly to assign people<br>3. to both exposed and unexposed groups? | ☑ | ☐ | ☐ | ☐ |
| 4. Was the exposure measured in a valid and reliable way? | ☑ | ☐ | ☐ | ☐ |
| 5. Were confounding factors identified? | ☑ | ☐ | ☐ | ☐ |
| 6. Were strategies to deal with confounding factors stated? | ☐ | ☐ | ☑ | ☐ |
| 7. Were the groups/participants free of the outcome at the start of the study (or at the moment of exposure)? | ☑ | ☐ | ☐ | ☐ |
| 8. Were the outcomes measured in a valid and reliable way? | ☑ | ☐ | ☐ | ☐ |
| 9. Was the follow up time reported and sufficient to be long enough for outcomes to occur? | ☑ | ☐ | ☐ | ☐ |
| 10. Was follow up complete, and if not, were the reasons to loss to follow up described and explored? | ☐ | ☐ | ☑ | ☐ |
| 11. Were strategies to address incomplete follow up utilized? | ☐ | ☐ | ☑ | ☐ |
| 12. Was appropriate statistical analysis used? | ☑ | ☐ | ☐ | ☐ |

Overall appraisal: Include ☐ Exclude ☑ Seek further info ☐

Comments (Including reason for exclusion)

El artículo incluye menores de 18 años, no se hace el diagnóstico exclusivamente con PCR o prueba de antígenos y no reporta síntomas persistentes

© JBI, 2020. All rights reserved. JBI grants use of these tools for research purposes only. All other enquiries should be sent to.

### JBI CRITICAL APPRAISAL CHECKLIST FOR COHORT STUDIES

Reviewer Carlos Alberto Fermin Martinez Date 11/04/2021

Author Chevrier S, et al. Year 2021 Record Number 171_33521697_CAFM

| | Yes | No | Unclear | Not applicable |
|---|---|---|---|---|
| 1. Were the two groups similar and recruited from the same population? | ☐ | ☐ | ☑ | ☐ |
| 2. Were the exposures measured similarly to assign people<br>3. to both exposed and unexposed groups? | ☑ | ☐ | ☐ | ☐ |
| 4. Was the exposure measured in a valid and reliable way? | ☑ | ☐ | ☐ | ☐ |
| 5. Were confounding factors identified? | ☑ | ☐ | ☐ | ☐ |
| 6. Were strategies to deal with confounding factors stated? | ☑ | ☐ | ☐ | ☐ |
| 7. Were the groups/participants free of the outcome at the start of the study (or at the moment of exposure)? | ☑ | ☐ | ☐ | ☐ |
| 8. Were the outcomes measured in a valid and reliable way? | ☑ | ☐ | ☐ | ☐ |
| 9. Was the follow up time reported and sufficient to be long enough for outcomes to occur? | ☑ | ☐ | ☐ | ☐ |
| 10. Was follow up complete, and if not, were the reasons to loss to follow up described and explored? | ☑ | ☐ | ☐ | ☐ |
| 11. Were strategies to address incomplete follow up utilized? | ☑ | ☐ | ☐ | ☐ |
| 12. Was appropriate statistical analysis used? | ☑ | ☐ | ☐ | ☐ |

Overall appraisal: Include ☐ Exclude ☑ Seek further info ☐

Comments (Including reason for exclusion)

El articulo no presenta síntomas persistentes, por lo que aunque la metodología sea robusta, no se justifica su inclusión en nuestro análisis.

© JBI, 2020. All rights reserved. JBI grants use of these tools for research purposes only. All other enquiries should be sent to.

### JBI CRITICAL APPRAISAL CHECKLIST FOR COHORT STUDIES

Reviewer Alejandro Sicilia Andrade Date 30 / 04 / 2021

Author Aditi S. Shah, Alyson W. Wong, Cameron J Hague et al. Year 2020 Record Number 172_33273023_ASA

| | Yes | No | Unclear | Not applicable |
|---|---|---|---|---|
| 1. Were the two groups similar and recruited from the same population? | X | | | |
| 2. Were the exposures measured similarly to assign people<br>3. to both exposed and unexposed groups? | X | | | |
| 4. Was the exposure measured in a valid and reliable way? | X | | | |
| 5. Were confounding factors identified? | X | | | |
| 6. Were strategies to deal with confounding factors stated? | | X | | |
| 7. Were the groups/participants free of the outcome at the start of the study (or at the moment of exposure)? | | X | | |
| 8. Were the outcomes measured in a valid and reliable way? | X | | | |
| 9. Was the follow up time reported and sufficient to be long enough for outcomes to occur? | X | | | |
| 10. Was follow up complete, and if not, were the reasons to loss to follow up described and explored? | X | | | |
| 11. Were strategies to address incomplete follow up utilized? | | X | | |
| 12. Was appropriate statistical analysis used? | | | X | |

Overall appraisal: Include ☐ Exclude ☒ Seek further info ☐

Comments (Including reason for exclusion)

© JBI, 2020. All rights reserved. JBI grants use of these tools for research purposes only. All other enquiries should be sent to.

### JBI CRITICAL APPRAISAL CHECKLIST FOR COHORT STUDIES

Reviewer Alejandro Sicilia Andrade Date 30/04/2021

Author Thomas Sonnweber, Sabina Sahanic, Alex Pizzini et al. Year 2020 Record Number 173_33303539_ASA

| | Yes | No | Unclear | Not applicable |
|---|---|---|---|---|
| 1. Were the two groups similar and recruited from the same population? | ☑ | ☐ | ☐ | ☐ |
| 2. Were the exposures measured similarly to assign people<br>3. to both exposed and unexposed groups? | ☑ | ☐ | ☐ | ☐ |
| 4. Was the exposure measured in a valid and reliable way? | ☑ | ☐ | ☐ | ☐ |
| 5. Were confounding factors identified? | ☑ | ☐ | ☐ | ☐ |
| 6. Were strategies to deal with confounding factors stated? | ☑ | ☐ | ☐ | ☐ |
| 7. Were the groups/participants free of the outcome at the start of the study (or at the moment of exposure)? | ☑ | ☐ | ☐ | ☐ |
| 8. Were the outcomes measured in a valid and reliable way? | ☑ | ☐ | ☐ | ☐ |
| 9. Was the follow up time reported and sufficient to be long enough for outcomes to occur? | ☑ | ☐ | ☐ | ☐ |
| 10. Was follow up complete, and if not, were the reasons to loss to follow up described and explored? | ☑ | ☐ | ☐ | ☐ |
| 11. Were strategies to address incomplete follow up utilized? | ☑ | ☐ | ☐ | ☐ |
| 12. Was appropriate statistical analysis used? | ☑ | ☐ | ☐ | ☐ |

Overall appraisal: Include ☑ Exclude ☐ Seek further info ☐

Comments (Including reason for exclusion)

This is a study of excellent quality.

© JBI, 2020. All rights reserved. JBI grants use of these tools for research purposes only. All other enquiries should be sent to.

### JBI CRITICAL APPRAISAL CHECKLIST FOR COHORT STUDIES

Reviewer Alejandro Sicilia Andrade Date 30 / 04 / 2021

Author C. Huart, C. Philpott, I. Konstantinidis, A. Altundag et al. Year 2020 Record Number 175_32812014_ASA

| | Yes | No | Unclear | Not applicable |
|---|---|---|---|---|
| 1. Were the two groups similar and recruited from the same population? | ☐ | ☐ | ☑ | ☐ |
| 2. Were the exposures measured similarly to assign people 3. to both exposed and unexposed groups? | ☐ | ☐ | ☑ | ☐ |
| 4. Was the exposure measured in a valid and reliable way? | ☐ | ☐ | ☑ | ☐ |
| 5. Were confounding factors identified? | ☐ | ☑ | ☐ | ☐ |
| 6. Were strategies to deal with confounding factors stated? | ☐ | ☑ | ☐ | ☐ |
| 7. Were the groups/participants free of the outcome at the start of the study (or at the moment of exposure)? | ☐ | ☐ | ☑ | ☐ |
| 8. Were the outcomes measured in a valid and reliable way? | ☑ | ☐ | ☐ | ☐ |
| 9. Was the follow up time reported and sufficient to be long enough for outcomes to occur? | ☐ | ☐ | ☑ | ☐ |
| 10. Was follow up complete, and if not, were the reasons to loss to follow up described and explored? | ☐ | ☐ | ☑ | ☐ |
| 11. Were strategies to address incomplete follow up utilized? | ☐ | ☐ | ☑ | ☐ |
| 12. Was appropriate statistical analysis used? | ☐ | ☐ | ☑ | ☐ |

Overall appraisal: Include ☐ Exclude ☑ Seek further info ☐

Comments (Including reason for exclusion)

Esta carta al editor menciona tener una pequeña cohorte, pero no profundiza en nada sobre la misma, no se incluye por falta de información.

© JBI, 2020. All rights reserved. JBI grants use of these tools for research purposes only. All other enquiries should be sent to.

### JBI CRITICAL APPRAISAL CHECKLIST FOR TEXT AND OPINION PAPERS

Reviewer Alejandro Sicilia Andrade Date 30/04/2021

Author Mark Vink, Alexandra Vink-Niese Year 2020 Record Number 176_33322316_ASA

| | Yes | No | Unclear | Not applicable |
|---|---|---|---|---|
| 1. Is the source of the opinion clearly identified? | ☑ | ☐ | ☐ | ☐ |
| 2. Does the source of opinion have standing in the field of expertise? | ☑ | ☐ | ☐ | ☐ |
| 3. Are the interests of the relevant population the central focus of the opinion? | ☑ | ☐ | ☐ | ☐ |
| 4. Is the stated position the result of an analytical process, and is there logic in the opinion expressed? | ☑ | ☐ | ☐ | ☐ |
| 5. Is there reference to the extant literature? | ☑ | ☐ | ☐ | ☐ |
| 6. Is any incongruence with the literature/sources logically defended? | ☑ | ☐ | ☐ | ☐ |

Overall appraisal: Include ☐ Exclude ☑ Seek further info ☐

Comments (Including reason for exclusion)

No se incluyen revisiones narrativas en el estudio.

© JBI, 2020. All rights reserved. JBI grants use of these tools for research purposes only. All other enquiries should be sent to.

### JBI CRITICAL APPRAISAL CHECKLIST FOR COHORT STUDIES

Reviewer: Alejandro Sicilia Andrade Date: 30 / 04 / 2021

Author: Mayssam Nehme, Olivia Braillard et al. Year: 2020 Record Number: 177_33284676

| | Yes | No | Unclear | Not applicable |
|---|---|---|---|---|
| 1. Were the two groups similar and recruited from the same population? | ☒ | ☐ | ☐ | ☐ |
| 2. Were the exposures measured similarly to assign people<br>3. to both exposed and unexposed groups? | ☒ | ☐ | ☐ | ☐ |
| 4. Was the exposure measured in a valid and reliable way? | ☒ | ☐ | ☐ | ☐ |
| 5. Were confounding factors identified? | ☐ | ☒ | ☐ | ☐ |
| 6. Were strategies to deal with confounding factors stated? | ☐ | ☒ | ☐ | ☐ |
| 7. Were the groups/participants free of the outcome at the start of the study (or at the moment of exposure)? | ☒ | ☐ | ☐ | ☐ |
| 8. Were the outcomes measured in a valid and reliable way? | ☒ | ☐ | ☐ | ☐ |
| 9. Was the follow up time reported and sufficient to be long enough for outcomes to occur? | ☒ | ☐ | ☐ | ☐ |
| 10. Was follow up complete, and if not, were the reasons to loss to follow up described and explored? | ☒ | ☐ | ☐ | ☐ |
| 11. Were strategies to address incomplete follow up utilized? | ☐ | ☒ | ☐ | ☐ |
| 12. Was appropriate statistical analysis used? | ☐ | ☐ | ☒ | ☐ |

Overall appraisal: Include ☐ Exclude ☒ Seek further info ☐

Comments (Including reason for exclusion)

© JBI, 2020. All rights reserved. JBI grants use of these tools for research purposes only. All other enquiries should be sent to.

### JBI CRITICAL APPRAISAL CHECKLIST FOR COHORT STUDIES

Reviewer: Alejandro Sicilia Andrade  Date: 30 / 04 / 2021

Author: Bin Liang, Lingi Xie, Fan Yang, Joyman Makamure, et al.  Year: 2020  Record Number: 178_33318560_ASA

| | Yes | No | Unclear | Not applicable |
|---|---|---|---|---|
| 1. Were the two groups similar and recruited from the same population? | ☑ | ☐ | ☐ | ☐ |
| 2. Were the exposures measured similarly to assign people<br>3. to both exposed and unexposed groups? | ☑ | ☐ | ☐ | ☐ |
| 4. Was the exposure measured in a valid and reliable way? | ☑ | ☐ | ☐ | ☐ |
| 5. Were confounding factors identified? | ☑ | ☐ | ☐ | ☐ |
| 6. Were strategies to deal with confounding factors stated? | ☐ | ☑ | ☐ | ☐ |
| 7. Were the groups/participants free of the outcome at the start of the study (or at the moment of exposure)? | ☑ | ☐ | ☐ | ☐ |
| 8. Were the outcomes measured in a valid and reliable way? | ☑ | ☐ | ☐ | ☐ |
| 9. Was the follow up time reported and sufficient to be long enough for outcomes to occur? | ☑ | ☐ | ☐ | ☐ |
| 10. Was follow up complete, and if not, were the reasons to loss to follow up described and explored? | ☑ | ☐ | ☐ | ☐ |
| 11. Were strategies to address incomplete follow up utilized? | ☐ | ☐ | ☑ | ☐ |
| 12. Was appropriate statistical analysis used? | ☑ | ☐ | ☐ | ☐ |

Overall appraisal: Include ☐ Exclude ☑ Seek further info ☐

Comments (Including reason for exclusion)

El articulo no menciona los sintomas del seguimiento.

© JBI, 2020. All rights reserved. JBI grants use of these tools for research purposes only. All other enquiries should be sent to.

JBI Critical Appraisal Checklist for
Qualitative Research

Reviewer Daniel de la Rosa. ________________ Date 03-02-21

Author Tom Kingstone et al ______________ Year: 2020 Record Number: 179_33051223

| | | Yes | No | Unclear | Not applicable |
|---|---|---|---|---|---|
| 1. | Is there congruity between the stated philosophical perspective and the research methodology? | ☒ | ☐ | ☐ | ☐ |
| 2. | Is there congruity between the research methodology and the research question or objectives? | ☒ | ☐ | ☐ | ☐ |
| 3. | Is there congruity between the research methodology and the methods used to collect data? | ☒ | ☐ | ☐ | ☐ |
| 4. | Is there congruity between the research methodology and the representation and analysis of data? | ☒ | ☐ | ☐ | ☐ |
| 5. | Is there congruity between the research methodology and the interpretation of results? | ☒ | ☐ | ☐ | ☐ |
| 6. | Is there a statement locating the researcher culturally or theoretically? | ☒ | ☐ | ☐ | ☐ |
| 7. | Is the influence of the researcher on the research, and vice- versa, addressed? | ☐ | ☐ | ☒ | ☐ |
| 8. | Are participants, and their voices, adequately represented? | ☐ | ☐ | ☒ | ☐ |
| 9. | Is the research ethical according to current criteria or, for recent studies, and is there evidence of ethical approval by an appropriate body? | ☐ | ☐ | ☒ | ☐ |
| 10. | Do the conclusions drawn in the research report flow from the analysis, or interpretation, of the data? | ☒ | ☐ | ☐ | ☐ |

Overall appraisal: Include ☐ Exclude ☒ Seek further info ☐

Comments (Including reason for exclusion)

__________ Qualitative Research articles are not included as inclusion criteria

### JBI CRITICAL APPRAISAL CHECKLIST FOR ANALYTICAL CROSS SECTIONAL STUDIES

Reviewer: Daniel de la Rosa  Date: 04-02-21

Author: Roy Meys, et al  Year: 2020  Record Number: 180_33317214

| | Yes | No | Unclear | Not applicable |
|---|---|---|---|---|
| 1. Were the criteria for inclusion in the sample clearly defined? | ☒ | ☐ | ☐ | ☐ |
| 2. Were the study subjects and the setting described in detail? | ☒ | ☐ | ☐ | ☐ |
| 3. Was the exposure measured in a valid and reliable way? | ☒ | ☐ | ☐ | ☐ |
| 4. Were objective, standard criteria used for measurement of the condition? | ☒ | ☐ | ☐ | ☐ |
| 5. Were confounding factors identified? | ☐ | ☒ | ☐ | ☐ |
| 6. Were strategies to deal with confounding factors stated? | ☐ | ☒ | ☐ | ☐ |
| 7. Were the outcomes measured in a valid and reliable way? | ☒ | ☐ | ☐ | ☐ |
| 8. Was appropriate statistical analysis used? | ☒ | ☐ | ☐ | ☐ |

Overall appraisal: Include ☐ Exclude ☒ Seek further info ☐

Comments (Including reason for exclusion)

El único síntoma persistente que se reporta es disnea en 64% de los pacientes, sin embargo, solo un pequeño porcentaje de los pacientes (23%, n=49) tuvieron COVID-19 confirmado por PCR. Se intenta mostrar la información de síntomas persistentes desagregada para los pacientes con PCR positiva, pero al parecer hay un error en la presentación de estos resultados. Se podría contactar a los autores para obtener solo la información de los pacientes PCR positivos, sin embargo, la baja calidad del estudio, aunado a el bajo porcentaje de pacientes con prueba de PCR, considero que no se justifica contactar a los autores y es mejor rechazar el artículo.

#### JBI Critical Appraisal Checklist for Case Series

Reviewer: Daniel de la Rosa Martinez Date: 03-02-21

Author: David Garcia-Azorin, et al Year: 2020 Record Number: 24_32998840

| | Yes | No | Unclear | Not applicable |
|---|---|---|---|---|
| • Were there clear criteria for inclusion in the case series? | ☑ | ☐ | ☐ | ☐ |
| • Was the condition measured in a standard, reliable way for all participants included in the case series? | ☐ | ☐ | ☑ | ☐ |
| • Were valid methods used for identification of the condition for all participants included in the case series? | ☑ | ☐ | ☐ | ☐ |
| • Did the case series have consecutive inclusion of participants? | ☐ | ☐ | ☑ | ☐ |
| • Did the case series have complete inclusion of participants? | ☐ | ☐ | ☑ | ☐ |
| • Was there clear reporting of the demographics of the participants in the study? | ☑ | ☐ | ☐ | ☐ |
| • Was there clear reporting of clinical information of the participants? | ☑ | ☐ | ☐ | ☐ |
| • Were the outcomes or follow up results of cases clearly reported? | ☑ | ☐ | ☐ | ☐ |
| • Was there clear reporting of the presenting site(s)/clinic(s) demographic information? | ☑ | ☐ | ☐ | ☐ |
| • Was statistical analysis appropriate? | ☐ | ☐ | ☐ | ☑ |

Overall appraisal: Include ☐ Exclude ☑ Seek further info ☐

Comments (Including reason for exclusion)

© JBI, 2020. All rights reserved. JBI grants use of these tools for research purposes only. All other enquiries should be sent to.

### JBI CRITICAL APPRAISAL CHECKLIST FOR COHORT STUDIES

Reviewer Enrique Cañedo Guerra Date 4/04/2021

Author Ortelli, P. O., et al. Year 2020 Record Number 182_33359928_ECG

| | Yes | No | Unclear | Not applicable |
|---|---|---|---|---|
| 1. Were the two groups similar and recruited from the same population? | ☐ | ☐ | ☐ | ☒ |
| 2. Were the exposures measured similarly to assign people to both exposed and unexposed groups? | ☐ | ☐ | ☐ | ☒ |
| 3. Was the exposure measured in a valid and reliable way? | ☐ | ☒ | ☐ | ☐ |
| 4. Were confounding factors identified? | ☐ | ☒ | ☐ | ☐ |
| 5. Were strategies to deal with confounding factors stated? | ☐ | ☒ | ☐ | ☐ |
| 6. Were the groups/participants free of the outcome at the start of the study (or at the moment of exposure)? | ☒ | ☐ | ☐ | ☐ |
| 7. Were the outcomes measured in a valid and reliable way? | ☒ | ☐ | ☐ | ☐ |
| 8. Was the follow up time reported and sufficient to be long enough for outcomes to occur? | ☒ | ☐ | ☐ | ☐ |
| 9. Was follow up complete, and if not, were the reasons to loss to follow up described and explored? | ☐ | ☐ | ☐ | ☒ |
| 10. Were strategies to address incomplete follow up utilized? | ☐ | ☐ | ☐ | ☒ |
| 11. Was appropriate statistical analysis used? | ☒ | ☐ | ☐ | ☐ |

Overall appraisal: Include ☐ Exclude ☒ Seek further info ☐

Comments (including reason for exclusion)

Estudio no especifica si se les hizo prueba de PCR. Fueron hospitalizados, pero no se especifica.

© JBI, 2020. All rights reserved. JBI grants use of these tools for research purposes only. All other enquiries should be sent to.

### JBI CRITICAL APPRAISAL CHECKLIST FOR COHORT STUDIES

Reviewer Enrique Cañedo Guerra Date 4/04/2021

Author Arnold, D. T., et al. Year 2020 Record Number 183_33273026_ECG

| | Yes | No | Unclear | Not applicable |
|---|---|---|---|---|
| 1. Were the two groups similar and recruited from the same population? | ☐ | ☐ | ☐ | ☒ |
| 2. Were the exposures measured similarly to assign people to both exposed and unexposed groups? | ☐ | ☐ | ☐ | ☒ |
| 3. Was the exposure measured in a valid and reliable way? | ☒ | ☐ | ☐ | ☐ |
| 4. Were confounding factors identified? | ☒ | ☐ | ☐ | ☐ |
| 5. Were strategies to deal with confounding factors stated? | ☒ | ☐ | ☐ | ☐ |
| 6. Were the groups/participants free of the outcome at the start of the study (or at the moment of exposure)? | ☐ | ☐ | ☒ | ☐ |
| 7. Were the outcomes measured in a valid and reliable way? | ☒ | ☐ | ☐ | ☐ |
| 8. Was the follow up time reported and sufficient to be long enough for outcomes to occur? | ☒ | ☐ | ☐ | ☐ |
| 9. Was follow up complete, and if not, were the reasons to loss to follow up described and explored? | ☒ | ☐ | ☐ | ☐ |
| 10. Were strategies to address incomplete follow up utilized? | ☐ | ☐ | ☐ | ☒ |
| 11. Was appropriate statistical analysis used? | ☒ | ☐ | ☐ | ☐ |

Overall appraisal: Include ☐ Exclude ☒ Seek further info ☐

Comments (including reason for exclusion)

Algunos valores de proporciones de sintomas no se pueden medir de la base de datos.

© JBI, 2020. All rights reserved. JBI grants use of these tools for research purposes only. All other enquiries should be sent to.

### CHECKLIST FOR CASE CONTROL STUDIES

Critical Appraisal tools for use in JBI Systematic Reviews

jbi.global

CRICOS Provider Number [illegible]

### INTRODUCTION

JBI is an international research organisation based in the Faculty of Health and Medical Sciences at the University of Adelaide, South Australia. JBI develops and delivers unique evidence-based information, software, education and training designed to improve healthcare practice and health outcomes. With over 70 Collaborating Entities, servicing over 90 countries, JBI is a recognised global leader in evidence-based healthcare.

#### JBI Systematic Reviews

The core of evidence synthesis is the systematic review of literature of a particular intervention, condition or issue. The systematic review is essentially an analysis of the available literature (that is, evidence) and a judgment of the effectiveness or otherwise of a practice, involving a series of complex steps. JBI takes a particular view on what counts as evidence and the methods utilised to synthesise those different types of evidence. In line with this broader view of evidence, JBI has developed theories, methodologies and rigorous processes for the critical appraisal and synthesis of these diverse forms of evidence in order to aid in clinical decision-making in healthcare. There now exists JBI guidance for conducting reviews of effectiveness research, qualitative research, prevalence/incidence, etiology/risk, economic evaluations, text/opinion, diagnostic test accuracy, mixed-methods, umbrella reviews and scoping reviews. Further information regarding JBI systematic reviews can be found in the JBI Evidence Synthesis Manual.

#### JBI Critical Appraisal Tools

All systematic reviews incorporate a process of critique or appraisal of the research evidence. The purpose of this appraisal is to assess the methodological quality of a study and to determine the extent to which a study has addressed the possibility of bias in its design, conduct and analysis. All papers selected for inclusion in the systematic review (that is – those that meet the inclusion criteria described in the protocol) need to be subjected to rigorous appraisal by two critical appraisers. The results of this appraisal can then be used to inform synthesis and interpretation of the results of the study. JBI Critical appraisal tools have been developed by the JBI and collaborators and approved by the JBI Scientific Committee following extensive peer review. Although designed for use in systematic reviews, JBI critical appraisal tools can also be used when creating Critically Appraised Topics (CAT), in journal clubs and as an educational tool.

© JBI, 2020. All rights reserved. JBI grants use of these tools for research purposes only. All other enquiries should be sent to.

### JBI CRITICAL APPRAISAL CHECKLIST FOR CASE CONTROL STUDIES

Reviewer Enrique Cañedo Guerra Date 4/04/2021

Author Galván-Tejada, C. E., et al. Year 2020 Record Number 184_33327641_ECG

| | Yes | No | Unclear | Not applicable |
|---|---|---|---|---|
| 1. Were the groups comparable other than the presence of disease in cases or the absence of disease in controls? | [x] | [ ] | [ ] | [ ] |
| 2. Were cases and controls matched appropriately? | [x] | [ ] | [ ] | [ ] |
| 3. Were the same criteria used for identification of cases and controls? | [x] | [ ] | [ ] | [ ] |
| 4. Was exposure measured in a standard, valid and reliable way? | [ ] | [ ] | [x] | [ ] |
| 5. Was exposure measured in the same way for cases and controls? | [x] | [ ] | [ ] | [ ] |
| 6. Were confounding factors identified? | [x] | [ ] | [ ] | [ ] |
| 7. Were strategies to deal with confounding factors stated? | [x] | [ ] | [ ] | [ ] |
| 8. Were outcomes assessed in a standard, valid and reliable way for cases and controls? | [x] | [ ] | [ ] | [ ] |
| 9. Was the exposure period of interest long enough to be meaningful? | [x] | [ ] | [ ] | [ ] |
| 10. Was appropriate statistical analysis used? | [x] | [ ] | [ ] | [ ] |

Overall appraisal: Include [ ] Exclude [x] Seek further info [ ]

Comments (Including reason for exclusion)

Dicen que incluyeron a sujetos con diagnóstico confirmado por laboratorio de acuerdo a los lineamientos de la OMS. Sin embargo, esto implicaría que podrían estar sujetos sin PCR dado que la definición operacional toma en cuenta prueba de antígenos.

© JBI, 2020. All rights reserved. JBI grants use of these tools for research purposes only. All other enquiries should be sent to.

### EXPLANATION OF CASE CONTROL STUDIES CRITICAL APPRAISAL

How to cite: *Moola S, Munn Z, Tufanaru C, Aromataris E, Sears K, Sfetcu R, Currie M, Qureshi R, Mattis P, Lisy K, Mu P-F. Chapter 7: Systematic reviews of etiology and risk . In: Aromataris E, Munn Z (Editors). JBI Manual for Evidence Synthesis.* JBI, 2020. Available from https://synthesismanual.jbi.global

**Case Control Studies Critical Appraisal Tool**

Answers: Yes, No, Unclear or Not/Applicable

1. **Were the groups comparable other than presence of disease in cases or absence of disease in controls?**

   The control group should be representative of the source population that produced the cases. This is usually done by individual matching; wherein controls are selected for each case on the basis of similarity with respect to certain characteristics other than the exposure of interest. Frequency or group matching is an alternative method. Selection bias may result if the groups are not comparable.

2. **Were cases and controls matched appropriately?**

   As in item 1, the study should include clear definitions of the source population. Sources from which cases and controls were recruited should be carefully looked at. For example, cancer registries may be used to recruit participants in a study examining risk factors for lung cancer, which typify population-based case control studies. Study participants may be selected from the target population, the source population, or from a pool of eligible participants (such as in hospital-based case control studies).

3. **Were the same criteria used for identification of cases and controls?**

   It is useful to determine if patients were included in the study based on either a specified diagnosis or definition. This is more likely to decrease the risk of bias. Characteristics are another useful approach to matching groups, and studies that did not use specified diagnostic methods or definitions should provide evidence on matching by key characteristics. A case should be defined clearly. It is also important that controls must fulfil all the eligibility criteria defined for the cases except for those relating to diagnosis of the disease.

4. **Was exposure measured in a standard, valid and reliable way?**

   The study should clearly describe the method of measurement of exposure. Assessing validity requires that a 'gold standard' is available to which the measure can be compared. The validity of exposure measurement usually relates to whether a current measure is appropriate or whether a measure of past exposure is needed.

   Case control studies may investigate many different 'exposures' that may or may not be associated with the condition. In these cases, reviewers should use the main exposure of interest for their review to answer this question when using this tool at the study level.

   Reliability refers to the processes included in an epidemiological study to check repeatability of measurements of the exposures. These usually include intra-observer reliability and inter-observer reliability.

5. **Was exposure measured in the same way for cases and controls?**

   As in item 4, the study should clearly describe the method of measurement of exposure. The exposure measures should be clearly defined and described in detail. Assessment of exposure or

© JBI, 2020. All rights reserved. JBI grants use of these tools for research purposes only. All other enquiries should be sent to.

risk factors should have been carried out according to same procedures or protocols for both cases and controls.

#### 6. Were confounding factors identified?

Confounding has occurred where the estimated intervention exposure effect is biased by the presence of some difference between the comparison groups (apart from the exposure investigated/of interest). Typical confounders include baseline characteristics, prognostic factors, or concomitant exposures (e.g. smoking). A confounder is a difference between the comparison groups and it influences the direction of the study results. A high quality study at the level of case control design will identify the potential confounders and measure them (where possible). This is difficult for studies where behavioral, attitudinal or lifestyle factors may impact on the results.

#### 7. Were strategies to deal with confounding factors stated?

Strategies to deal with effects of confounding factors may be dealt within the study design or in data analysis. By matching or stratifying sampling of participants, effects of confounding factors can be adjusted for. When dealing with adjustment in data analysis, assess the statistics used in the study. Most will be some form of multivariate regression analysis to account for the confounding factors measured. Look out for a description of statistical methods as regression methods such as logistic regression are usually employed to deal with confounding factors/ variables of interest.

#### 8. Were outcomes assessed in a standard, valid and reliable way for cases and controls?

Read the methods section of the paper. If for e.g. lung cancer is assessed based on existing definitions or diagnostic criteria, then the answer to this question is likely to be yes. If lung cancer is assessed using observer reported, or self-reported scales, the risk of over- or under-reporting is increased, and objectivity is compromised. Importantly, determine if the measurement tools used were validated instruments as this has a significant impact on outcome assessment validity.

Having established the objectivity of the outcome measurement (e.g. lung cancer) instrument, it's important to establish how the measurement was conducted. Were those involved in collecting data trained or educated in the use of the instrument/s? (e.g. radiographers). If there was more than one data collector, were they similar in terms of level of education, clinical or research experience, or level of responsibility in the piece of research being appraised?

#### 9. Was the exposure period of interest long enough to be meaningful?

It is particularly important in a case control study that the exposure time was sufficient enough to show an association between the exposure and the outcome. It may be that the exposure period may be too short or too long to influence the outcome.

#### 10. Was appropriate statistical analysis used?

As with any consideration of statistical analysis, consideration should be given to whether there was a more appropriate alternate statistical method that could have been used. The methods section should be detailed enough for reviewers to identify which analytical techniques were used (in particular, regression or stratification) and how specific confounders were measured.

For studies utilizing regression analysis, it is useful to identify if the study identified which variables were included and how they related to the outcome. If stratification was the analytical approach used, were the strata of analysis defined by the specified variables? Additionally, it is also important to assess the appropriateness of the analytical strategy in terms of the assumptions associated with the approach as differing methods of analysis are based on differing assumptions about the data and how it will respond.

© JBI, 2020. All rights reserved. JBI grants use of these tools for research purposes only. All other enquiries should be sent to.

© JBI, 2020. All rights reserved. JBI grants use of these tools for research purposes only. All other enquiries should be sent to.

### JBI CRITICAL APPRAISAL CHECKLIST FOR COHORT STUDIES

Reviewer Enrique Cañedo Guerra Date 4/04/2021

Author Ikegami, S, et al. Year 2020 Record Number 185_32840002_ECG

| | Yes | No | Unclear | Not applicable |
|---|---|---|---|---|
| 1. Were the two groups similar and recruited from the same population? | ☐ | ☐ | ☐ | ☒ |
| 2. Were the exposures measured similarly to assign people to both exposed and unexposed groups? | ☐ | ☐ | ☐ | ☒ |
| 3. Was the exposure measured in a valid and reliable way? | ☒ | ☐ | ☐ | ☐ |
| 4. Were confounding factors identified? | ☒ | ☐ | ☐ | ☐ |
| 5. Were strategies to deal with confounding factors stated? | ☒ | ☐ | ☐ | ☐ |
| 6. Were the groups/participants free of the outcome at the start of the study (or at the moment of exposure)? | ☒ | ☐ | ☐ | ☐ |
| 7. Were the outcomes measured in a valid and reliable way? | ☒ | ☐ | ☐ | ☐ |
| 8. Was the follow up time reported and sufficient to be long enough for outcomes to occur? | ☒ | ☐ | ☐ | ☐ |
| 9. Was follow up complete, and if not, were the reasons to loss to follow up described and explored? | ☒ | ☐ | ☐ | ☐ |
| 10. Were strategies to address incomplete follow up utilized? | ☐ | ☐ | ☐ | ☒ |
| 11. Was appropriate statistical analysis used? | ☒ | ☐ | ☐ | ☐ |

Overall appraisal: Include ☐ Exclude ☒ Seek further info ☐

Comments (including reason for exclusion)

No trata de síntomas persistentes, si no de persistencia de PCR positiva.

© JBI, 2020. All rights reserved. JBI grants use of these tools for research purposes only. All other enquiries should be sent to.

### JBI CRITICAL APPRAISAL CHECKLIST FOR COHORT STUDIES

Reviewer Enrique Cañedo Guerra Date 4/04/2021

Author Ikegami, S, et al. Year 2020 Record Number 186_33310664_ECG

| | Yes | No | Unclear | Not applicable |
|---|---|---|---|---|
| 1. Were the two groups similar and recruited from the same population? | ☐ | ☐ | ☐ | ☑ |
| 2. Were the exposures measured similarly to assign people to both exposed and unexposed groups? | ☐ | ☐ | ☐ | ☑ |
| 3. Was the exposure measured in a valid and reliable way? | ☑ | ☐ | ☐ | ☐ |
| 4. Were confounding factors identified? | ☑ | ☐ | ☐ | ☐ |
| 5. Were strategies to deal with confounding factors stated? | ☑ | ☐ | ☐ | ☐ |
| 6. Were the groups/participants free of the outcome at the start of the study (or at the moment of exposure)? | ☑ | ☐ | ☐ | ☐ |
| 7. Were the outcomes measured in a valid and reliable way? | ☑ | ☐ | ☐ | ☐ |
| 8. Was the follow up time reported and sufficient to be long enough for outcomes to occur? | ☑ | ☐ | ☐ | ☐ |
| 9. Was follow up complete, and if not, were the reasons to loss to follow up described and explored? | ☑ | ☐ | ☐ | ☐ |
| 10. Were strategies to address incomplete follow up utilized? | ☐ | ☐ | ☐ | ☑ |
| 11. Was appropriate statistical analysis used? | ☑ | ☐ | ☐ | ☐ |

Overall appraisal: Include ☐ Exclude ☑ Seek further info ☐

Comments (including reason for exclusion)

El articulo trata de persistencia de anticuerpos especificos contra SARS-CoV-2, no de sintomas.

© JBI, 2020. All rights reserved. JBI grants use of these tools for research purposes only. All other enquiries should be sent to.

### JBI CRITICAL APPRAISAL CHECKLIST FOR COHORT STUDIES

Reviewer Enrique Cañedo Guerra Date 4/04/2021

Author Jacobs, LG, et al. Year 2020 Record Number 187_33306721_ECG

| | Yes | No | Unclear | Not applicable |
|---|---|---|---|---|
| 1. Were the two groups similar and recruited from the same population? | ☐ | ☐ | ☐ | ☑ |
| 2. Were the exposures measured similarly to assign people to both exposed and unexposed groups? | ☐ | ☐ | ☐ | ☑ |
| 3. Was the exposure measured in a valid and reliable way? | ☑ | ☐ | ☐ | ☐ |
| 4. Were confounding factors identified? | ☑ | ☐ | ☐ | ☐ |
| 5. Were strategies to deal with confounding factors stated? | ☑ | ☐ | ☐ | ☐ |
| 6. Were the groups/participants free of the outcome at the start of the study (or at the moment of exposure)? | ☑ | ☐ | ☐ | ☐ |
| 7. Were the outcomes measured in a valid and reliable way? | ☐ | ☑ | ☐ | ☐ |
| 8. Was the follow up time reported and sufficient to be long enough for outcomes to occur? | ☑ | ☐ | ☐ | ☐ |
| 9. Was follow up complete, and if not, were the reasons to loss to follow up described and explored? | ☑ | ☐ | ☐ | ☐ |
| 10. Were strategies to address incomplete follow up utilized? | ☐ | ☐ | ☐ | ☑ |
| 11. Was appropriate statistical analysis used? | ☑ | ☐ | ☐ | ☐ |

Overall appraisal: Include ☑ Exclude ☐ Seek further info ☐

Comments (including reason for exclusion)

Autorreporte de sintomas por los pacientes, pero de todas formas creo que es información útil.

© JBI, 2020. All rights reserved. JBI grants use of these tools for research purposes only. All other enquiries should be sent to.

### JBI CRITICAL APPRAISAL CHECKLIST FOR COHORT STUDIES

Reviewer Enrique Cañedo Guerra Date 4/04/2021

Author Stavem, K, et al. Year 2020 Record Number 188 33273028 ECG

| | Yes | No | Unclear | Not applicable |
|---|---|---|---|---|
| 1. Were the two groups similar and recruited from the same population? | ☐ | ☐ | ☐ | ☒ |
| 2. Were the exposures measured similarly to assign people to both exposed and unexposed groups? | ☐ | ☐ | ☐ | ☒ |
| 3. Was the exposure measured in a valid and reliable way? | ☒ | ☐ | ☐ | ☐ |
| 4. Were confounding factors identified? | ☒ | ☐ | ☐ | ☐ |
| 5. Were strategies to deal with confounding factors stated? | ☒ | ☐ | ☐ | ☐ |
| 6. Were the groups/participants free of the outcome at the start of the study (or at the moment of exposure)? | ☐ | ☒ | ☐ | ☐ |
| 7. Were the outcomes measured in a valid and reliable way? | ☐ | ☐ | ☒ | ☐ |
| 8. Was the follow up time reported and sufficient to be long enough for outcomes to occur? | ☒ | ☐ | ☐ | ☐ |
| 9. Was follow up complete, and if not, were the reasons to loss to follow up described and explored? | ☒ | ☐ | ☐ | ☐ |
| 10. Were strategies to address incomplete follow up utilized? | ☒ | ☐ | ☐ | ☐ |
| 11. Was appropriate statistical analysis used? | ☒ | ☐ | ☐ | ☐ |

Overall appraisal: Include ☐ Exclude ☒ Seek further info ☐

Comments (including reason for exclusion)

No queda claro como se hizo la recolección de datos de los sintomas. Algunas suposiciones se hicieron para determinar que los pacientes eran elegibles en el estudio.

© JBI, 2020. All rights reserved. JBI grants use of these tools for research purposes only. All other enquiries should be sent to.

### JBI CRITICAL APPRAISAL CHECKLIST FOR ANALYTICAL CROSS SECTIONAL STUDIES

Reviewer Enrique Cañedo Guerra Date 14/04/2021

Author El Sayed, S, et al. Year 2020 Record Number 189_33332756_ECG

| | Yes | No | Unclear | Not applicable |
|---|---|---|---|---|
| 1. Were the criteria for inclusion in the sample clearly defined? | ☐ | ☒ | ☐ | ☐ |
| 2. Were the study subjects and the setting described in detail? | ☐ | ☒ | ☐ | ☐ |
| 3. Was the exposure measured in a valid and reliable way? | ☒ | ☐ | ☐ | ☐ |
| 4. Were objective, standard criteria used for measurement of the condition? | ☒ | ☐ | ☐ | ☐ |
| 5. Were confounding factors identified? | ☒ | ☐ | ☐ | ☐ |
| 6. Were strategies to deal with confounding factors stated? | ☒ | ☐ | ☐ | ☐ |
| 7. Were the outcomes measured in a valid and reliable way? | ☒ | ☐ | ☐ | ☐ |
| 8. Was appropriate statistical analysis used? | ☒ | ☐ | ☐ | ☐ |

Overall appraisal: Include ☐ Exclude ☐ Seek further info ☒

Comments (Including reason for exclusion)

Se comenta información de anhedonia y fatiga medida con dos escalas diferentes en pacientes recuperados con COVID-19 que hayan tenido dos pruebas negativas. Se podría haber caracterizado mejor a la población. Hicieron un muestreo por conveniencia. Valdría la pena preguntarle a los autores si en su estudio seleccionaron a pacientes con prueba de PCR positivo en el diagnóstico.

© JBI, 2020. All rights reserved. JBI grants use of these tools for research purposes only. All other enquiries should be sent to.

### JBI CRITICAL APPRAISAL CHECKLIST FOR QUALITATIVE RESEARCH

Reviewer Enrique Cañedo Guerra Date 14/04/2021

Author Ladds, E, et al. Year 2020 Record Number 190_33342437_ECG

| | Yes | No | Unclear | Not applicable |
|---|---|---|---|---|
| 1. Is there congruity between the stated philosophical perspective and the research methodology? | ☒ | ☐ | ☐ | ☐ |
| 2. Is there congruity between the research methodology and the research question or objectives? | ☒ | ☐ | ☐ | ☐ |
| 3. Is there congruity between the research methodology and the methods used to collect data? | ☒ | ☐ | ☐ | ☐ |
| 4. Is there congruity between the research methodology and the representation and analysis of data? | ☒ | ☐ | ☐ | ☐ |
| 5. Is there congruity between the research methodology and the interpretation of results? | ☒ | ☐ | ☐ | ☐ |
| 6. Is there a statement locating the researcher culturally or theoretically? | ☒ | ☐ | ☐ | ☐ |
| 7. Is the influence of the researcher on the research, and vice- versa, addressed? | ☒ | ☐ | ☐ | ☐ |
| 8. Are participants, and their voices, adequately represented? | ☒ | ☐ | ☐ | ☐ |
| 9. Is the research ethical according to current criteria or, for recent studies, and is there evidence of ethical approval by an appropriate body? | ☒ | ☐ | ☐ | ☐ |
| 10. Do the conclusions drawn in the research report flow from the analysis, or interpretation, of the data? | ☒ | ☐ | ☐ | ☐ |

Overall appraisal: Include ☐ Exclude ☒ Seek further info ☐

Comments (Including reason for exclusion)

Es un estudio cualitativo donde no todos los pacientes fueron diagnósticados con prueba de PCR aparentemente. Al parecer algunos sujetos solo fueron incluidos por presencia de síntomas en un periodo de tiempo.

© JBI, 2020. All rights reserved. JBI grants use of these tools for research purposes only. All other enquiries should be sent to.

### JBI CRITICAL APPRAISAL CHECKLIST FOR COHORT STUDIES

Reviewer Enrique Cañedo Guerra Date________________________

Author Chun Gao, et. al. Year 2020 Record Number 192 33068796 ECG

| | Yes | No | Unclear | Not applicable |
|---|---|---|---|---|
| 1. Were the two groups similar and recruited from the same population? | ☒ | ☐ | ☐ | ☐ |
| 2. Were the exposures measured similarly to assign people to both exposed and unexposed groups? | ☒ | ☐ | ☐ | ☐ |
| 3. Was the exposure measured in a valid and reliable way? | ☒ | ☐ | ☐ | ☐ |
| 4. Were confounding factors identified? | ☒ | ☐ | ☐ | ☐ |
| 5. Were strategies to deal with confounding factors stated? | ☒ | ☐ | ☐ | ☐ |
| 6. Were the groups/participants free of the outcome at the start of the study (or at the moment of exposure)? | ☐ | ☒ | ☐ | ☐ |
| 7. Were the outcomes measured in a valid and reliable way? | ☒ | ☐ | ☐ | ☐ |
| 8. Was the follow up time reported and sufficient to be long enough for outcomes to occur? | ☒ | ☐ | ☐ | ☐ |
| 9. Was follow up complete, and if not, were the reasons to loss to follow up described and explored? | ☒ | ☐ | ☐ | ☐ |
| 10. Were strategies to address incomplete follow up utilized? | ☐ | ☐ | ☐ | ☒ |
| 11. Was appropriate statistical analysis used? | ☒ | ☐ | ☐ | ☐ |

Overall appraisal: Include ☐ Exclude ☒ Seek further info ☐

Comments (including reason for exclusion)

The clinical status of patients is unclear throughout the study. It is implied that their symptoms were recollected from hospital admission. The primary objective of the study is to explore factors associated with prolonged viral shedding, not long COVID.

© JBI, 2020. All rights reserved. JBI grants use of these tools for research purposes only. All other enquiries should be sent to.

### JBI CRITICAL APPRAISAL CHECKLIST FOR COHORT STUDIES

Reviewer Enrique Cañedo Guerra Date 04/04/2021

Author Taboada M, et al. Year 2020 Record Number 193 33413976 ECG

| | Yes | No | Unclear | Not applicable |
|---|---|---|---|---|
| 1. Were the two groups similar and recruited from the same population? | ☐ | ☐ | ☐ | ☒ |
| 2. Were the exposures measured similarly to assign people<br>3. to both exposed and unexposed groups? | ☐ | ☐ | ☐ | ☒ |
| 4. Was the exposure measured in a valid and reliable way? | ☒ | ☐ | ☐ | ☐ |
| 5. Were confounding factors identified? | ☐ | ☒ | ☐ | ☐ |
| 6. Were strategies to deal with confounding factors stated? | ☐ | ☒ | ☐ | ☐ |
| 7. Were the groups/participants free of the outcome at the start of the study (or at the moment of exposure)? | ☐ | ☒ | ☐ | ☐ |
| 8. Were the outcomes measured in a valid and reliable way? | ☐ | ☒ | ☐ | ☐ |
| 9. Was the follow up time reported and sufficient to be long enough for outcomes to occur? | ☒ | ☐ | ☐ | ☐ |
| 10. Was follow up complete, and if not, were the reasons to loss to follow up described and explored? | ☒ | ☐ | ☐ | ☐ |
| 11. Were strategies to address incomplete follow up utilized? | ☐ | ☐ | ☐ | ☒ |
| 12. Was appropriate statistical analysis used? | ☒ | ☐ | ☐ | ☐ |

Overall appraisal: Include ☒ Exclude ☐ Seek further info ☐

Comments (including reason for exclusion)

© JBI, 2020. All rights reserved. JBI grants use of these tools for research purposes only. All other enquiries should be sent to.

### JBI CRITICAL APPRAISAL CHECKLIST FOR COHORT STUDIES

Reviewer Mónica Itzel Martínez Gutiérrez Date 01/04/2021

Author Chen, Y., Zuiani, A., Fischinger, S., Mullur, J., Atyeo, C. et.al. Year 2020 Record Number 194 33171099 MIMG

| | Yes | No | Unclear | Not applicable |
|---|---|---|---|---|
| 1. Were the two groups similar and recruited from the same population? | ☐ | ☐ | ☐ | ☑ |
| 2. Were the exposures measured similarly to assign people to both exposed and unexposed groups? | ☐ | ☐ | ☐ | ☑ |
| 3. Was the exposure measured in a valid and reliable way? | ☑ | ☐ | ☐ | ☐ |
| 4. Were confounding factors identified? | ☐ | ☑ | ☐ | ☐ |
| 5. Were strategies to deal with confounding factors stated? | ☐ | ☑ | ☐ | ☐ |
| 6. Were the groups/participants free of the outcome at the start of the study (or at the moment of exposure)? | ☐ | ☑ | ☐ | ☐ |
| 7. Were the outcomes measured in a valid and reliable way? | ☐ | ☐ | ☐ | ☐ |
| 8. Was the follow up time reported and sufficient to be long enough for outcomes to occur? | ☑ | ☐ | ☐ | ☐ |
| 9. Was follow up complete, and if not, were the reasons to loss to follow up described and explored? | ☑ | ☐ | ☐ | ☐ |
| 10. Were strategies to address incomplete follow up utilized? | ☐ | ☐ | ☐ | ☑ |
| 11. Was appropriate statistical analysis used? | ☑ | ☐ | ☐ | ☐ |

Overall appraisal: Include ☐ Exclude ☑ Seek further info ☐

Comments (Including reason for exclusion)

### JBI CRITICAL APPRAISAL CHECKLIST FOR CASE CONTROL STUDIES

Reviewer Mónica Itzel Martínez Gutiérrez Date 01/04/2021

Author Hartley, G. E., Edwards, E., Aui, P. M., Varese, N., Stojanovic, S., McMahon, J. et al. Year 2020 Record Number 195 33443036 MIMG

| | Yes | No | Unclear | Not applicable |
|---|---|---|---|---|
| 1. Were the groups comparable other than the presence of disease in cases or the absence of disease in controls? | ☑ | ☐ | ☐ | ☐ |
| 2. Were cases and controls matched appropriately? | ☐ | ☐ | ☑ | ☐ |
| 3. Were the same criteria used for identification of cases and controls? | ☑ | ☐ | ☐ | ☐ |
| 4. Was exposure measured in a standard, valid and reliable way? | ☑ | ☐ | ☐ | ☐ |
| 5. Was exposure measured in the same way for cases and controls? | ☑ | ☐ | ☐ | ☐ |
| 6. Were confounding factors identified? | ☐ | ☑ | ☐ | ☐ |
| 7. Were strategies to deal with confounding factors stated? | ☐ | ☑ | ☐ | ☐ |
| 8. Were outcomes assessed in a standard, valid and reliable way for cases and controls? | ☑ | ☐ | ☐ | ☐ |
| 9. Was the exposure period of interest long enough to be meaningful? | ☑ | ☐ | ☐ | ☐ |
| 10. Was appropriate statistical analysis used? | ☑ | ☐ | ☐ | ☐ |

Overall appraisal: Include ☐ Exclude ☑ Seek further info ☐

Comments (Including reason for exclusion)

© JBI, 2020. All rights reserved. JBI grants use of these tools for research purposes only. All other enquiries should be sent to.

### JBI CRITICAL APPRAISAL CHECKLIST FOR ANALYTICAL CROSS SECTIONAL STUDIES

Reviewer Enrique Cañedo Guerra Date 03/04/2021

Author Richard C. Gerkin, Kathrin Ohla, et al. Year 2020 Record Number 196_33367502_ECG

| | Yes | No | Unclear | Not applicable |
|---|---|---|---|---|
| 1. Were the criteria for inclusion in the sample clearly defined? | ☒ | ☐ | ☐ | ☐ |
| 2. Were the study subjects and the setting described in detail? | ☒ | ☐ | ☐ | ☐ |
| 3. Was the exposure measured in a valid and reliable way? | ☒ | ☐ | ☐ | ☐ |
| 4. Were objective, standard criteria used for measurement of the condition? | ☒ | ☐ | ☐ | ☐ |
| 5. Were confounding factors identified? | ☐ | ☐ | ☒ | ☐ |
| 6. Were strategies to deal with confounding factors stated? | ☐ | ☐ | ☒ | ☐ |
| 7. Were the outcomes measured in a valid and reliable way? | ☒ | ☐ | ☐ | ☐ |
| 8. Was appropriate statistical analysis used? | ☒ | ☐ | ☐ | ☐ |

Overall appraisal: Include ☒ Exclude ☐ Seek further info ☐

Comments (including reason for exclusion)

Sólo se comenta en una sección sobre la recuperación de la función olfatoria en los primeros 40 días de seguimiento después del episodio agudo. 50.7% de los 3147 pacientes reportaron pérdida del olfato persistente, pero no se tiene información de ningún síntoma adicional.

© JBI, 2020. All rights reserved. JBI grants use of these tools for research purposes only. All other enquiries should be sent to.

### JBI CRITICAL APPRAISAL CHECKLIST FOR COHORT STUDIES

Reviewer Mónica Itzel Martínez Gutiérrez Date 01/04/2021

Author Tien, J., Zhang, M., Jin, M., Zhang, F., Chu, Q., et al. Year 2020 Record Number 197 33298617 MIMG

| | Yes | No | Unclear | Not applicable |
|---|---|---|---|---|
| 1. Were the two groups similar and recruited from the same population? | X | | | |
| 2. Were the exposures measured similarly to assign people to both exposed and unexposed groups? | X | | | |
| 3. Was the exposure measured in a valid and reliable way? | X | | | |
| 4. Were confounding factors identified? | X | | | |
| 5. Were strategies to deal with confounding factors stated? | X | | | |
| 6. Were the groups/participants free of the outcome at the start of the study (or at the moment of exposure)? | X | | | |
| 7. Were the outcomes measured in a valid and reliable way? | X | | | |
| 8. Was the follow up time reported and sufficient to be long enough for outcomes to occur? | X | | | |
| 9. Was follow up complete, and if not, were the reasons to loss to follow up described and explored? | X | | | |
| 10. Were strategies to address incomplete follow up utilized? | | | | X |
| 11. Was appropriate statistical analysis used? | X | | | |

Overall appraisal: Include ☐ Exclude ☑ Seek further info ☐

Comments (Including reason for exclusion)

Es un estudio centrado en el resultado del uso de tocilizumab como tratamiento en pacientes con cuadro severo de COVID, donde se reporta mejoría clínica y diminución de la mortalidad.

### JBI CRITICAL APPRAISAL CHECKLIST FOR COHORT STUDIES

Reviewer Enrique Cañedo Guerra Date 4/04/2021

Author Zhang, X, et al. Year 2020 Record Number 198 33388574 ECG

| | Yes | No | Unclear | Not applicable |
|---|---|---|---|---|
| 1. Were the two groups similar and recruited from the same population? | ☐ | ☐ | ☒ | ☐ |
| 2. Were the exposures measured similarly to assign people to both exposed and unexposed groups? | ☐ | ☐ | ☐ | ☒ |
| 3. Was the exposure measured in a valid and reliable way? | ☒ | ☐ | ☐ | ☐ |
| 4. Were confounding factors identified? | ☐ | ☒ | ☐ | ☐ |
| 5. Were strategies to deal with confounding factors stated? | ☐ | ☒ | ☐ | ☐ |
| 6. Were the groups/participants free of the outcome at the start of the study (or at the moment of exposure)? | ☐ | ☒ | ☐ | ☐ |
| 7. Were the outcomes measured in a valid and reliable way? | ☒ | ☐ | ☐ | ☐ |
| 8. Was the follow up time reported and sufficient to be long enough for outcomes to occur? | ☒ | ☐ | ☐ | ☐ |
| 9. Was follow up complete, and if not, were the reasons to loss to follow up described and explored? | ☐ | ☐ | ☒ | ☐ |
| 10. Were strategies to address incomplete follow up utilized? | ☐ | ☒ | ☐ | ☐ |
| 11. Was appropriate statistical analysis used? | ☒ | ☐ | ☐ | ☐ |

Overall appraisal: Include ☐ Exclude ☒ Seek further info ☐

Comments (including reason for exclusion)

No se mencionan datos de sintomas ni caracteristicas de los pacientes.

© JBI, 2020. All rights reserved. JBI grants use of these tools for research purposes only. All other enquiries should be sent to.

### JBI CRITICAL APPRAISAL CHECKLIST FOR COHORT STUDIES

Reviewer Mónica Itzel Martínez Gutiérrez Date 02/04/2021

Author Xiaoyong Zhang, Suwen Lu, Hui Li, Yi Wang, Zhen Lu, et al. Year 2020 Record Number 199 33351168 MIMG

| | Yes | No | Unclear | Not applicable |
|---|---|---|---|---|
| 1. Were the two groups similar and recruited from the same population? | ☐ | ☐ | ☑ | ☐ |
| 2. Were the exposures measured similarly to assign people to both exposed and unexposed groups? | ☐ | ☐ | ☐ | ☑ |
| 3. Was the exposure measured in a valid and reliable way? | ☑ | ☐ | ☐ | ☐ |
| 4. Were confounding factors identified? | ☐ | ☑ | ☐ | ☐ |
| 5. Were strategies to deal with confounding factors stated? | ☐ | ☑ | ☐ | ☐ |
| 6. Were the groups/participants free of the outcome at the start of the study (or at the moment of exposure) ? | ☐ | ☑ | ☐ | ☐ |
| 7. Were the outcomes measured in a valid and reliable way? | ☑ | ☐ | ☐ | ☐ |
| 8. Was the follow up time reported and sufficient to be long enough for outcomes to occur? | ☑ | ☐ | ☐ | ☐ |
| 9. Was follow up complete, and if not, were the reasons to loss to follow up described and explored? | ☐ | ☐ | ☑ | ☐ |
| 10. Were strategies to address incomplete follow up utilized? | ☐ | ☑ | ☐ | ☐ |
| 11. Was appropriate statistical analysis used? | ☑ | ☐ | ☐ | ☐ |

© JBI, 2020. All rights reserved. JBI grants use of these tools for research purposes only. All other enquiries should be sent to.

Overall appraisal: Include ☐ Exclude ☑ Seek further info ☐

Comments (Including reason for exclusion)

El seguimiento de pacientes convalecientes solo conforma una sección en el artículo, no se mencionan las características clínicas o demográficas de estos pacientes.

© JBI, 2020. All rights reserved. JBI grants use of these tools for research purposes only. All other enquiries should be sent to.

### JBI CRITICAL APPRAISAL CHECKLIST FOR COHORT STUDIES

Reviewer Enrique Cañedo Guerra Date 4/16/2021

Author Blair, PW, et al. Year 2021 Record Number 200_33614816_ECG

| | Yes | No | Unclear | Not applicable |
|---|---|---|---|---|
| 1. Were the two groups similar and recruited from the same population? | ☐ | ☐ | ☐ | ☑ |
| 2. Were the exposures measured similarly to assign people to both exposed and unexposed groups? | ☐ | ☐ | ☐ | ☑ |
| 3. Was the exposure measured in a valid and reliable way? | ☑ | ☐ | ☐ | ☐ |
| 4. Were confounding factors identified? | ☐ | ☑ | ☐ | ☐ |
| 5. Were strategies to deal with confounding factors stated? | ☐ | ☑ | ☐ | ☐ |
| 6. Were the groups/participants free of the outcome at the start of the study (or at the moment of exposure)? | ☐ | ☑ | ☐ | ☐ |
| 7. Were the outcomes measured in a valid and reliable way? | ☑ | ☐ | ☐ | ☐ |
| 8. Was the follow up time reported and sufficient to be long enough for outcomes to occur? | ☑ | ☐ | ☐ | ☐ |
| 9. Was follow up complete, and if not, were the reasons to loss to follow up described and explored? | ☑ | ☐ | ☐ | ☐ |
| 10. Were strategies to address incomplete follow up utilized? | ☑ | ☐ | ☐ | ☐ |
| 11. Was appropriate statistical analysis used? | ☑ | ☐ | ☐ | ☐ |

Overall appraisal: Include ☑ Exclude ☐ Seek further info ☐

Comments (including reason for exclusion)

© JBI, 2020. All rights reserved. JBI grants use of these tools for research purposes only. All other enquiries should be sent to.

### JBI CRITICAL APPRAISAL CHECKLIST FOR CASE REPORTS

Reviewer Mónica Itzel Martínez Gutiérrez Date 02/04/2021

Author Dadhwal, R., Sharma, M., & Surani, S. Year 2021 Record Number 201_33564509_MIMG

| | Yes | No | Unclear | Not applicable |
|---|---|---|---|---|
| 1. Were patient's demographic characteristics clearly described? | ☐ | ☑ | ☐ | ☐ |
| 2. Was the patient's history clearly described and presented as a timeline? | ☑ | ☐ | ☐ | ☐ |
| 3. Was the current clinical condition of the patient on presentation clearly described? | ☑ | ☐ | ☐ | ☐ |
| 4. Were diagnostic tests or assessment methods and the results clearly described? | ☑ | ☐ | ☐ | ☐ |
| 5. Was the intervention(s) or treatment procedure(s) clearly described? | ☑ | ☐ | ☐ | ☐ |
| 6. Was the post-intervention clinical condition clearly described? | ☐ | ☑ | ☐ | ☐ |
| 7. Were adverse events (harms) or unanticipated events identified and described? | ☐ | ☑ | ☐ | ☐ |
| 8. Does the case report provide takeaway lessons? | ☑ | ☐ | ☐ | ☐ |

Overall appraisal: Include ☐ Exclude ☑ Seek further info ☐

Comments (Including reason for exclusion)

© JBI, 2020. All rights reserved. JBI grants use of these tools for research purposes only. All other enquiries should be sent to.

### JBI CRITICAL APPRAISAL CHECKLIST FOR CASE CONTROL STUDIES

Reviewer: Mónica Itzel Martínez Gutiérrez Date 29/03/21

Author Kiva A. Fisher Samantha M. Olson Mark W. Tenforde Wesley H. Self Michael Wu et.al.

Year 2021 Record Number 202 33405338 MIMG

| | Yes | No | Unclear | Not applicable |
|---|---|---|---|---|
| 1. Were the groups comparable other than the presence of disease in cases or the absence of disease in controls? | ☑ | ☐ | ☐ | ☐ |
| 2. Were cases and controls matched appropriately? | ☑ | ☐ | ☐ | ☐ |
| 3. Were the same criteria used for identification of cases and controls? | ☑ | ☐ | ☐ | ☐ |
| 4. Was exposure measured in a standard, valid and reliable way? | ☐ | ☑ | ☐ | ☐ |
| 5. Was exposure measured in the same way for cases and controls? | ☑ | ☐ | ☐ | ☐ |
| 6. Were confounding factors identified? | ☑ | ☐ | ☐ | ☐ |
| 7. Were strategies to deal with confounding factors stated? | ☑ | ☐ | ☐ | ☐ |
| 8. Were outcomes assessed in a standard, valid and reliable way for cases and controls? | ☑ | ☐ | ☐ | ☐ |
| 9. Was the exposure period of interest long enough to be meaningful? | ☐ | ☐ | ☑ | ☐ |
| 10. Was appropriate statistical analysis used? | ☑ | ☐ | ☐ | ☐ |

Overall appraisal: Include ☐ Exclude ☐ Seek further info ☑

Comments (Including reason for exclusion)

Las entrevistas realizadas para el reporte de la salud física y mental de los participantes, fueron realizadas con preguntas modificadas del CDC Healthy Days Measures y éste no ha sido validado para abordar la calidad de vida relacionada con la salud durante la fase de convalecencia de la enfermedad aguda y los días en el cuestionario fueron ajustados a 14 días previos al diagnóstico en vez del original, de 30 días.

© JBI, 2020. All rights reserved. JBI grants use of these

tools for research purposes only. All other enquiries should be sent to.

### JBI CRITICAL APPRAISAL CHECKLIST FOR COHORT STUDIES

Reviewer Fernández Chirino Luisa Date 15/04/2021

Author Kandermirli, S., Altundag A., et. al. Year 2021 Record Number 203_33132007_LFC

| | Yes | No | Unclear | Not applicable |
|---|---|---|---|---|
| 1. Were the two groups similar and recruited from the same population? | X | ☐ | ☐ | ☐ |
| 2. Were the exposures measured similarly to assign people<br>3. to both exposed and unexposed groups? | X | ☐ | ☐ | ☐ |
| 4. Was the exposure measured in a valid and reliable way? | X | ☐ | ☐ | ☐ |
| 5. Were confounding factors identified? | X | ☐ | ☐ | ☐ |
| 6. Were strategies to deal with confounding factors stated? | X | ☐ | ☐ | ☐ |
| 7. Were the groups/participants free of the outcome at the start of the study (or at the moment of exposure)? | X | ☐ | ☐ | ☐ |
| 8. Were the outcomes measured in a valid and reliable way? | X | ☐ | ☐ | ☐ |
| 9. Was the follow up time reported and sufficient to be long enough for outcomes to occur? | X | ☐ | ☐ | ☐ |
| 10. Was follow up complete, and if not, were the reasons to loss to follow up described and explored? | X | ☐ | ☐ | ☐ |
| 11. Were strategies to address incomplete follow up utilized? | X | ☐ | ☐ | ☐ |
| 12. Was appropriate statistical analysis used? | X | ☐ | ☐ | ☐ |

Overall appraisal: Include ☐ Exclude X Seek further info ☐

Comments (including reason for exclusion)

### JBI CRITICAL APPRAISAL CHECKLIST FOR COHORT STUDIES

Reviewer Fernández Chirino Luisa Date 15/04/2021

Author Kavaz, E., Tahir, E., et. al. Year 2021 Record Number 204_33386439_LFC

| | Yes | No | Unclear | Not applicable |
|---|---|---|---|---|
| 1. Were the two groups similar and recruited from the same population? | ☒ | ☐ | ☐ | ☐ |
| 2. Were the exposures measured similarly to assign people<br>3. to both exposed and unexposed groups? | ☒ | ☐ | ☐ | ☐ |
| 4. Was the exposure measured in a valid and reliable way? | ☒ | ☐ | ☐ | ☐ |
| 5. Were confounding factors identified? | ☐ | ☐ | ☒ | ☐ |
| 6. Were strategies to deal with confounding factors stated? | ☐ | ☐ | ☒ | ☐ |
| 7. Were the groups/participants free of the outcome at the start of the study (or at the moment of exposure)? | ☒ | ☐ | ☐ | ☐ |
| 8. Were the outcomes measured in a valid and reliable way? | ☒ | ☐ | ☐ | ☐ |
| 9. Was the follow up time reported and sufficient to be long enough for outcomes to occur? | ☒ | ☐ | ☐ | ☐ |
| 10. Was follow up complete, and if not, were the reasons to loss to follow up described and explored? | ☐ | ☒ | ☐ | ☐ |
| 11. Were strategies to address incomplete follow up utilized? | ☒ | ☐ | ☐ | ☐ |
| 12. Was appropriate statistical analysis used? | ☒ | ☐ | ☐ | ☐ |

Overall appraisal: Include ☐ Exclude ☒ Seek further info ☐

Comments (Including reason for exclusion)

### JBI CRITICAL APPRAISAL CHECKLIST FOR QUALITATIVE RESEARCH

Reviewer Mónica Itzel Martínez Gutiérrez Date 30/03/2021

Author Emma Ladds, Alex Rushforth, Sietse Wieringa, Sharon Taylor, Clare Rayner, Laiba Husain and Trisha Greenhalgh Year 2021 Record Number 205_33479069_MIMG

| | | Yes | No | Unclear | Not applicable |
|---|---|---|---|---|---|
| 1. | Is there congruity between the stated philosophical perspective and the research methodology? | ☑ | ☐ | ☐ | ☐ |
| 2. | Is there congruity between the research methodology and the research question or objectives? | ☑ | ☐ | ☐ | ☐ |
| 3. | Is there congruity between the research methodology and the methods used to collect data? | ☑ | ☐ | ☐ | ☐ |
| 4. | Is there congruity between the research methodology and the representation and analysis of data? | ☑ | ☐ | ☐ | ☐ |
| 5. | Is there congruity between the research methodology and the interpretation of results? | ☑ | ☐ | ☐ | ☐ |
| 6. | Is there a statement locating the researcher culturally or theoretically? | ☐ | ☑ | ☐ | ☐ |
| 7. | Is the influence of the researcher on the research, and vice- versa, addressed? | ☐ | ☑ | ☐ | ☐ |
| 8. | Are participants, and their voices, adequately represented? | ☐ | ☑ | ☐ | ☐ |
| 9. | Is the research ethical according to current criteria or, for recent studies, and is there evidence of ethical approval by an appropriate body? | ☑ | ☐ | ☐ | ☐ |
| 10. | Do the conclusions drawn in the research report flow from the analysis, or interpretation, of the data? | ☑ | ☐ | ☐ | ☐ |

© JBI, 2020. All rights reserved. JBI grants use of these tools for research purposes only. All other enquiries should be sent to

Overall appraisal: Include ☐ Exclude ☑ Seek further info ☐

Comments (Including reason for exclusion)

Es un estudio qualitativo que no cumple con los criterios de inclusión, no se hace seguimiento de los síntomas posteriores a la enfermedad aguda sino que más bien reporta las experiencias del personal de salud (que fueron a su vez pacientes) con el denominado "long COVID", centrándose en temas como facilidad de acceso a los servicios, atención al paciente, responsabilidad clínica y la propuesta de un modelo para mejorar la atención a los pacientes con long COVID

© JBI, 2020. All rights reserved. JBI grants use of these tools for research purposes only. All other enquiries should be sent to

### JBI CRITICAL APPRAISAL CHECKLIST FOR COHORT STUDIES

Reviewer Fernández Chirino Luisa Date 15/04/2021

Author Langham, D., Loe, J., et. al. Year 2021 Record Number 206_33419864_LFC

| | Yes | No | Unclear | Not applicable |
|---|---|---|---|---|
| 1. Were the two groups similar and recruited from the same population? | ☒ | ☐ | ☐ | ☐ |
| 2. Were the exposures measured similarly to assign people<br>3. to both exposed and unexposed groups? | ☒ | ☐ | ☐ | ☐ |
| 4. Was the exposure measured in a valid and reliable way? | ☒ | ☐ | ☐ | ☐ |
| 5. Were confounding factors identified? | ☐ | ☐ | ☒ | ☐ |
| 6. Were strategies to deal with confounding factors stated? | ☐ | ☐ | ☒ | ☐ |
| 7. Were the groups/participants free of the outcome at the start of the study (or at the moment of exposure)? | ☒ | ☐ | ☐ | ☐ |
| 8. Were the outcomes measured in a valid and reliable way? | ☒ | ☐ | ☐ | ☐ |
| 9. Was the follow up time reported and sufficient to be long enough for outcomes to occur? | ☒ | ☐ | ☐ | ☐ |
| 10. Was follow up complete, and if not, were the reasons to loss to follow up described and explored? | ☐ | ☒ | ☐ | ☐ |
| 11. Were strategies to address incomplete follow up utilized? | ☒ | ☐ | ☐ | ☐ |
| 12. Was appropriate statistical analysis used? | ☒ | ☐ | ☐ | ☐ |

Overall appraisal: Include ☐ Exclude ☐ Seek further info ☒

Comments (including reason for exclusion)

Useful for discussion

### JBI CRITICAL APPRAISAL CHECKLIST FOR COHORT STUDIES

Reviewer Fernández Chirino Luisa Date 15/04/2021

Author Niklassen, A. S., Draf, J., et. al. Year 2021 Record Number 207_33404079_LFC

| | Yes | No | Unclear | Not applicable |
|---|---|---|---|---|
| 1. Were the two groups similar and recruited from the same population? | ☒ | ☐ | ☐ | ☐ |
| 2. Were the exposures measured similarly to assign people<br>3. to both exposed and unexposed groups? | ☒ | ☐ | ☐ | ☐ |
| 4. Was the exposure measured in a valid and reliable way? | ☒ | ☐ | ☐ | ☐ |
| 5. Were confounding factors identified? | ☒ | ☐ | ☐ | ☐ |
| 6. Were strategies to deal with confounding factors stated? | ☒ | ☐ | ☐ | ☐ |
| 7. Were the groups/participants free of the outcome at the start of the study (or at the moment of exposure)? | ☒ | ☐ | ☐ | ☐ |
| 8. Were the outcomes measured in a valid and reliable way? | ☒ | ☐ | ☐ | ☐ |
| 9. Was the follow up time reported and sufficient to be long enough for outcomes to occur? | ☒ | ☐ | ☐ | ☐ |
| 10. Was follow up complete, and if not, were the reasons to loss to follow up described and explored? | ☐ | ☒ | ☐ | ☐ |
| 11. Were strategies to address incomplete follow up utilized? | ☒ | ☐ | ☐ | ☐ |
| 12. Was appropriate statistical analysis used? | ☒ | ☐ | ☐ | ☐ |

Overall appraisal: Include ☒ Exclude ☐ Seek further info ☐

Comments (including reason for exclusion)

### JBI CRITICAL APPRAISAL CHECKLIST FOR CASE CONTROL STUDIES

Reviewer Fernández Chirino Luisa Date 15/04/2021

Author Ramman, B., Cassar, M.P., et. al Year 2021 Record Number 208_33490928_LFC

| | Yes | No | Unclear | Not applicable |
|---|---|---|---|---|
| 1. Were the groups comparable other than the presence of disease in cases or the absence of disease in controls? | ☒ | ☐ | ☐ | ☐ |
| 2. Were cases and controls matched appropriately? | ☒ | ☐ | ☐ | ☐ |
| 3. Were the same criteria used for identification of cases and controls? | ☒ | ☐ | ☐ | ☐ |
| 4. Was exposure measured in a standard, valid and reliable way? | ☒ | ☐ | ☐ | ☐ |
| 5. Was exposure measured in the same way for cases and controls? | ☒ | ☐ | ☐ | ☐ |
| 6. Were confounding factors identified? | ☒ | ☐ | ☐ | ☐ |
| 7. Were strategies to deal with confounding factors stated? | ☒ | ☐ | ☐ | ☐ |
| 8. Were outcomes assessed in a standard, valid and reliable way for cases and controls? | ☒ | ☐ | ☐ | ☐ |
| 9. Was the exposure period of interest long enough to be meaningful? | ☒ | ☐ | ☐ | ☐ |
| 10. Was appropriate statistical analysis used? | ☒ | ☐ | ☐ | ☐ |

Overall appraisal: Include ☒ Exclude ☐ Seek further info ☐

Comments (Including reason for exclusion)

### EXPLANATION OF CASE CONTROL STUDIES CRITICAL APPRAISAL

How to cite: *Moola S, Munn Z, Tufanaru C, Aromataris E, Sears K, Sfetcu R, Currie M, Qureshi R, Mattis P, Lisy K, Mu P-F. Chapter 7: Systematic reviews of etiology and risk . In: Aromataris E, Munn Z (Editors). JBI Manual for Evidence Synthesis.* JBI, 2020. Available from https://synthesismanual.jbi.global

**Case Control Studies Critical Appraisal Tool**

Answers: Yes, No, Unclear or Not/Applicable

1. **Were the groups comparable other than presence of disease in cases or absence of disease in controls?**

   The control group should be representative of the source population that produced the cases. This is usually done by individual matching; wherein controls are selected for each case on the basis of similarity with respect to certain characteristics other than the exposure of interest. Frequency or group matching is an alternative method. Selection bias may result if the groups are not comparable.

2. **Were cases and controls matched appropriately?**

   As in item 1, the study should include clear definitions of the source population. Sources from which cases and controls were recruited should be carefully looked at. For example, cancer registries may be used to recruit participants in a study examining risk factors for lung cancer, which typify population-based case control studies. Study participants may be selected from the target population, the source population, or from a pool of eligible participants (such as in hospital-based case control studies).

3. **Were the same criteria used for identification of cases and controls?**

   It is useful to determine if patients were included in the study based on either a specified diagnosis or definition. This is more likely to decrease the risk of bias. Characteristics are another useful approach to matching groups, and studies that did not use specified diagnostic methods or definitions should provide evidence on matching by key characteristics. A case should be defined clearly. It is also important that controls must fulfil all the eligibility criteria defined for the cases except for those relating to diagnosis of the disease.

4. **Was exposure measured in a standard, valid and reliable way?**

   The study should clearly describe the method of measurement of exposure. Assessing validity requires that a 'gold standard' is available to which the measure can be compared. The validity of exposure measurement usually relates to whether a current measure is appropriate or whether a measure of past exposure is needed.

   Case control studies may investigate many different 'exposures' that may or may not be associated with the condition. In these cases, reviewers should use the main exposure of interest for their review to answer this question when using this tool at the study level.

   Reliability refers to the processes included in an epidemiological study to check repeatability of measurements of the exposures. These usually include intra-observer reliability and inter-observer reliability.

5. **Was exposure measured in the same way for cases and controls?**

   As in item 4, the study should clearly describe the method of measurement of exposure. The exposure measures should be clearly defined and described in detail. Assessment of exposure or

© JBI, 2020. All rights reserved. JBI grants use of these tools for research purposes only. All other enquiries should be sent to.

risk factors should have been carried out according to same procedures or protocols for both cases and controls.

#### 6. Were confounding factors identified?

Confounding has occurred where the estimated intervention exposure effect is biased by the presence of some difference between the comparison groups (apart from the exposure investigated/of interest). Typical confounders include baseline characteristics, prognostic factors, or concomitant exposures (e.g. smoking). A confounder is a difference between the comparison groups and it influences the direction of the study results. A high quality study at the level of case control design will identify the potential confounders and measure them (where possible). This is difficult for studies where behavioral, attitudinal or lifestyle factors may impact on the results.

#### 7. Were strategies to deal with confounding factors stated?

Strategies to deal with effects of confounding factors may be dealt within the study design or in data analysis. By matching or stratifying sampling of participants, effects of confounding factors can be adjusted for. When dealing with adjustment in data analysis, assess the statistics used in the study. Most will be some form of multivariate regression analysis to account for the confounding factors measured. Look out for a description of statistical methods as regression methods such as logistic regression are usually employed to deal with confounding factors/ variables of interest.

#### 8. Were outcomes assessed in a standard, valid and reliable way for cases and controls?

Read the methods section of the paper. If for e.g. lung cancer is assessed based on existing definitions or diagnostic criteria, then the answer to this question is likely to be yes. If lung cancer is assessed using observer reported, or self-reported scales, the risk of over- or under-reporting is increased, and objectivity is compromised. Importantly, determine if the measurement tools used were validated instruments as this has a significant impact on outcome assessment validity.

Having established the objectivity of the outcome measurement (e.g. lung cancer) instrument, it's important to establish how the measurement was conducted. Were those involved in collecting data trained or educated in the use of the instrument/s? (e.g. radiographers). If there was more than one data collector, were they similar in terms of level of education, clinical or research experience, or level of responsibility in the piece of research being appraised?

#### 9. Was the exposure period of interest long enough to be meaningful?

It is particularly important in a case control study that the exposure time was sufficient enough to show an association between the exposure and the outcome. It may be that the exposure period may be too short or too long to influence the outcome.

#### 10. Was appropriate statistical analysis used?

As with any consideration of statistical analysis, consideration should be given to whether there was a more appropriate alternate statistical method that could have been used. The methods section should be detailed enough for reviewers to identify which analytical techniques were used (in particular, regression or stratification) and how specific confounders were measured.

For studies utilizing regression analysis, it is useful to identify if the study identified which variables were included and how they related to the outcome. If stratification was the analytical approach used, were the strata of analysis defined by the specified variables? Additionally, it is also important to assess the appropriateness of the analytical strategy in terms of the assumptions associated with the approach as differing methods of analysis are based on differing assumptions about the data and how it will respond.

© JBI, 2020. All rights reserved. JBI grants use of these tools for research purposes only. All other enquiries should be sent to.

© JBI, 2020. All rights reserved. JBI grants use of these tools for research purposes only. All other enquiries should be sent to.

### JBI CRITICAL APPRAISAL CHECKLIST FOR CASE CONTROL STUDIES

Reviewer Fernández Chirino Luisa Date 15/04/2021

Author Ratchford, S., M., Stickford, J. L., et. al Year 2021 Record Number 209_33306450_LFC

| | Yes | No | Unclear | Not applicable |
|---|---|---|---|---|
| 1. Were the groups comparable other than the presence of disease in cases or the absence of disease in controls? | ☒ | ☐ | ☐ | ☐ |
| 2. Were cases and controls matched appropriately? | ☒ | ☐ | ☐ | ☐ |
| 3. Were the same criteria used for identification of cases and controls? | ☒ | ☐ | ☐ | ☐ |
| 4. Was exposure measured in a standard, valid and reliable way? | ☒ | ☐ | ☐ | ☐ |
| 5. Was exposure measured in the same way for cases and controls? | ☒ | ☐ | ☐ | ☐ |
| 6. Were confounding factors identified? | ☒ | ☐ | ☐ | ☐ |
| 7. Were strategies to deal with confounding factors stated? | ☒ | ☐ | ☐ | ☐ |
| 8. Were outcomes assessed in a standard, valid and reliable way for cases and controls? | ☒ | ☐ | ☐ | ☐ |
| 9. Was the exposure period of interest long enough to be meaningful? | ☒ | ☐ | ☐ | ☐ |
| 10. Was appropriate statistical analysis used? | ☒ | ☐ | ☐ | ☐ |

Overall appraisal: Include ☐ Exclude ☒ Seek further info ☐

Comments (Including reason for exclusion)

Not relevant to Long COVID.

### EXPLANATION OF CASE CONTROL STUDIES CRITICAL APPRAISAL

How to cite: *Moola S, Munn Z, Tufanaru C, Aromataris E, Sears K, Sfetcu R, Currie M, Qureshi R, Mattis P, Lisy K, Mu P-F. Chapter 7: Systematic reviews of etiology and risk . In: Aromataris E, Munn Z (Editors). JBI Manual for Evidence Synthesis.* JBI, 2020. Available from https://synthesismanual.jbi.global

**Case Control Studies Critical Appraisal Tool**

Answers: Yes, No, Unclear or Not/Applicable

1. **Were the groups comparable other than presence of disease in cases or absence of disease in controls?**

   The control group should be representative of the source population that produced the cases. This is usually done by individual matching; wherein controls are selected for each case on the basis of similarity with respect to certain characteristics other than the exposure of interest. Frequency or group matching is an alternative method. Selection bias may result if the groups are not comparable.

2. **Were cases and controls matched appropriately?**

   As in item 1, the study should include clear definitions of the source population. Sources from which cases and controls were recruited should be carefully looked at. For example, cancer registries may be used to recruit participants in a study examining risk factors for lung cancer, which typify population-based case control studies. Study participants may be selected from the target population, the source population, or from a pool of eligible participants (such as in hospital-based case control studies).

3. **Were the same criteria used for identification of cases and controls?**

   It is useful to determine if patients were included in the study based on either a specified diagnosis or definition. This is more likely to decrease the risk of bias. Characteristics are another useful approach to matching groups, and studies that did not use specified diagnostic methods or definitions should provide evidence on matching by key characteristics. A case should be defined clearly. It is also important that controls must fulfil all the eligibility criteria defined for the cases except for those relating to diagnosis of the disease.

4. **Was exposure measured in a standard, valid and reliable way?**

   The study should clearly describe the method of measurement of exposure. Assessing validity requires that a 'gold standard' is available to which the measure can be compared. The validity of exposure measurement usually relates to whether a current measure is appropriate or whether a measure of past exposure is needed.

   Case control studies may investigate many different 'exposures' that may or may not be associated with the condition. In these cases, reviewers should use the main exposure of interest for their review to answer this question when using this tool at the study level.

   Reliability refers to the processes included in an epidemiological study to check repeatability of measurements of the exposures. These usually include intra-observer reliability and inter-observer reliability.

5. **Was exposure measured in the same way for cases and controls?**

   As in item 4, the study should clearly describe the method of measurement of exposure. The exposure measures should be clearly defined and described in detail. Assessment of exposure or

© JBI, 2020. All rights reserved. JBI grants use of these tools for research purposes only. All other enquiries should be sent to.

risk factors should have been carried out according to same procedures or protocols for both cases and controls.

##### 6. Were confounding factors identified?

Confounding has occurred where the estimated intervention exposure effect is biased by the presence of some difference between the comparison groups (apart from the exposure investigated/of interest). Typical confounders include baseline characteristics, prognostic factors, or concomitant exposures (e.g. smoking). A confounder is a difference between the comparison groups and it influences the direction of the study results. A high quality study at the level of case control design will identify the potential confounders and measure them (where possible). This is difficult for studies where behavioral, attitudinal or lifestyle factors may impact on the results.

##### 7. Were strategies to deal with confounding factors stated?

Strategies to deal with effects of confounding factors may be dealt within the study design or in data analysis. By matching or stratifying sampling of participants, effects of confounding factors can be adjusted for. When dealing with adjustment in data analysis, assess the statistics used in the study. Most will be some form of multivariate regression analysis to account for the confounding factors measured. Look out for a description of statistical methods as regression methods such as logistic regression are usually employed to deal with confounding factors/ variables of interest.

##### 8. Were outcomes assessed in a standard, valid and reliable way for cases and controls?

Read the methods section of the paper. If for e.g. lung cancer is assessed based on existing definitions or diagnostic criteria, then the answer to this question is likely to be yes. If lung cancer is assessed using observer reported, or self-reported scales, the risk of over- or under-reporting is increased, and objectivity is compromised. Importantly, determine if the measurement tools used were validated instruments as this has a significant impact on outcome assessment validity.

Having established the objectivity of the outcome measurement (e.g. lung cancer) instrument, it's important to establish how the measurement was conducted. Were those involved in collecting data trained or educated in the use of the instrument/s? (e.g. radiographers). If there was more than one data collector, were they similar in terms of level of education, clinical or research experience, or level of responsibility in the piece of research being appraised?

##### 9. Was the exposure period of interest long enough to be meaningful?

It is particularly important in a case control study that the exposure time was sufficient enough to show an association between the exposure and the outcome. It may be that the exposure period may be too short or too long to influence the outcome.

##### 10. Was appropriate statistical analysis used?

As with any consideration of statistical analysis, consideration should be given to whether there was a more appropriate alternate statistical method that could have been used. The methods section should be detailed enough for reviewers to identify which analytical techniques were used (in particular, regression or stratification) and how specific confounders were measured.

For studies utilizing regression analysis, it is useful to identify if the study identified which variables were included and how they related to the outcome. If stratification was the analytical approach used, were the strata of analysis defined by the specified variables? Additionally, it is also important to assess the appropriateness of the analytical strategy in terms of the assumptions associated with the approach as differing methods of analysis are based on differing assumptions about the data and how it will respond.

© JBI, 2020. All rights reserved. JBI grants use of these tools for research purposes only. All other enquiries should be sent to.

© JBI, 2020. All rights reserved. JBI grants use of these tools for research purposes only. All other enquiries should be sent to.

### JBI CRITICAL APPRAISAL CHECKLIST FOR COHORT STUDIES

Reviewer Mónica Itzel Martinez Gutiérrez Date 31/03/2021

Author Rosales-Castillo A, García de Los Ríos C, Mediavilla García Year 2021 Record Number 210 33521308 MIMG

| | Yes | No | Unclear | Not applicable |
|---|---|---|---|---|
| 1. Were the two groups similar and recruited from the same population? | ☐ | ☐ | ☐ | ☑ |
| 2. Were the exposures measured similarly to assign people to both exposed and unexposed groups? | ☐ | ☐ | ☐ | ☑ |
| 3. Was the exposure measured in a valid and reliable way? | ☐ | ☐ | ☐ | ☑ |
| 4. Were confounding factors identified? | ☐ | ☑ | ☐ | ☐ |
| 5. Were strategies to deal with confounding factors stated? | ☐ | ☑ | ☐ | ☐ |
| 6. Were the groups/participants free of the outcome at the start of the study (or at the moment of exposure)? | ☐ | ☑ | ☐ | ☐ |
| 7. Were the outcomes measured in a valid and reliable way? | ☑ | ☐ | ☐ | ☐ |
| 8. Was the follow up time reported and sufficient to be long enough for outcomes to occur? | ☑ | ☐ | ☐ | ☐ |
| 9. Was follow up complete, and if not, were the reasons to loss to follow up described and explored? | ☑ | ☐ | ☐ | ☐ |
| 10. Were strategies to address incomplete follow up utilized? | ☐ | ☐ | ☑ | ☐ |
| 11. Was appropriate statistical analysis used? | ☐ | ☐ | ☑ | ☐ |

Overall appraisal: Include ☐ Exclude ☐ Seek further info ☑

Comments (Including reason for exclusion)

### JBI CRITICAL APPRAISAL CHECKLIST FOR CASE CONTROL STUDIES

Reviewer Fernández Chirino Luisa Date 15/04/2021

Author Sheng, W.H., Lui, W.D., et. al Year 2021 Record Number 211_33139151_LFC

| | Yes | No | Unclear | Not applicable |
|---|---|---|---|---|
| 1. Were the groups comparable other than the presence of disease in cases or the absence of disease in controls? | ☒ | ☐ | ☐ | ☐ |
| 2. Were cases and controls matched appropriately? | ☒ | ☐ | ☐ | ☐ |
| 3. Were the same criteria used for identification of cases and controls? | ☒ | ☐ | ☐ | ☐ |
| 4. Was exposure measured in a standard, valid and reliable way? | ☒ | ☐ | ☐ | ☐ |
| 5. Was exposure measured in the same way for cases and controls? | ☒ | ☐ | ☐ | ☐ |
| 6. Were confounding factors identified? | ☒ | ☐ | ☐ | ☐ |
| 7. Were strategies to deal with confounding factors stated? | ☒ | ☐ | ☐ | ☐ |
| 8. Were outcomes assessed in a standard, valid and reliable way for cases and controls? | ☒ | ☐ | ☐ | ☐ |
| 9. Was the exposure period of interest long enough to be meaningful? | ☒ | ☐ | ☐ | ☐ |
| 10. Was appropriate statistical analysis used? | ☒ | ☐ | ☐ | ☐ |

Overall appraisal: Include ☐ Exclude ☒ Seek further info ☐

Comments (Including reason for exclusion)

Does not speak about Long COVID.

### EXPLANATION OF CASE CONTROL STUDIES CRITICAL APPRAISAL

How to cite: *Moola S, Munn Z, Tufanaru C, Aromataris E, Sears K, Sfetcu R, Currie M, Qureshi R, Mattis P, Lisy K, Mu P-F. Chapter 7: Systematic reviews of etiology and risk . In: Aromataris E, Munn Z (Editors). JBI Manual for Evidence Synthesis.* JBI, 2020. Available from https://synthesismanual.jbi.global

**Case Control Studies Critical Appraisal Tool**

Answers: Yes, No, Unclear or Not/Applicable

1. **Were the groups comparable other than presence of disease in cases or absence of disease in controls?**

   The control group should be representative of the source population that produced the cases. This is usually done by individual matching; wherein controls are selected for each case on the basis of similarity with respect to certain characteristics other than the exposure of interest. Frequency or group matching is an alternative method. Selection bias may result if the groups are not comparable.

2. **Were cases and controls matched appropriately?**

   As in item 1, the study should include clear definitions of the source population. Sources from which cases and controls were recruited should be carefully looked at. For example, cancer registries may be used to recruit participants in a study examining risk factors for lung cancer, which typify population-based case control studies. Study participants may be selected from the target population, the source population, or from a pool of eligible participants (such as in hospital-based case control studies).

3. **Were the same criteria used for identification of cases and controls?**

   It is useful to determine if patients were included in the study based on either a specified diagnosis or definition. This is more likely to decrease the risk of bias. Characteristics are another useful approach to matching groups, and studies that did not use specified diagnostic methods or definitions should provide evidence on matching by key characteristics. A case should be defined clearly. It is also important that controls must fulfil all the eligibility criteria defined for the cases except for those relating to diagnosis of the disease.

4. **Was exposure measured in a standard, valid and reliable way?**

   The study should clearly describe the method of measurement of exposure. Assessing validity requires that a 'gold standard' is available to which the measure can be compared. The validity of exposure measurement usually relates to whether a current measure is appropriate or whether a measure of past exposure is needed.

   Case control studies may investigate many different 'exposures' that may or may not be associated with the condition. In these cases, reviewers should use the main exposure of interest for their review to answer this question when using this tool at the study level.

   Reliability refers to the processes included in an epidemiological study to check repeatability of measurements of the exposures. These usually include intra-observer reliability and inter-observer reliability.

5. **Was exposure measured in the same way for cases and controls?**

   As in item 4, the study should clearly describe the method of measurement of exposure. The exposure measures should be clearly defined and described in detail. Assessment of exposure or

© JBI, 2020. All rights reserved. JBI grants use of these tools for research purposes only. All other enquiries should be sent to.

risk factors should have been carried out according to same procedures or protocols for both cases and controls.

##### 6. Were confounding factors identified?

Confounding has occurred where the estimated intervention exposure effect is biased by the presence of some difference between the comparison groups (apart from the exposure investigated/of interest). Typical confounders include baseline characteristics, prognostic factors, or concomitant exposures (e.g. smoking). A confounder is a difference between the comparison groups and it influences the direction of the study results. A high quality study at the level of case control design will identify the potential confounders and measure them (where possible). This is difficult for studies where behavioral, attitudinal or lifestyle factors may impact on the results.

##### 7. Were strategies to deal with confounding factors stated?

Strategies to deal with effects of confounding factors may be dealt within the study design or in data analysis. By matching or stratifying sampling of participants, effects of confounding factors can be adjusted for. When dealing with adjustment in data analysis, assess the statistics used in the study. Most will be some form of multivariate regression analysis to account for the confounding factors measured. Look out for a description of statistical methods as regression methods such as logistic regression are usually employed to deal with confounding factors/ variables of interest.

##### 8. Were outcomes assessed in a standard, valid and reliable way for cases and controls?

Read the methods section of the paper. If for e.g. lung cancer is assessed based on existing definitions or diagnostic criteria, then the answer to this question is likely to be yes. If lung cancer is assessed using observer reported, or self-reported scales, the risk of over- or under-reporting is increased, and objectivity is compromised. Importantly, determine if the measurement tools used were validated instruments as this has a significant impact on outcome assessment validity.

Having established the objectivity of the outcome measurement (e.g. lung cancer) instrument, it's important to establish how the measurement was conducted. Were those involved in collecting data trained or educated in the use of the instrument/s? (e.g. radiographers). If there was more than one data collector, were they similar in terms of level of education, clinical or research experience, or level of responsibility in the piece of research being appraised?

##### 9. Was the exposure period of interest long enough to be meaningful?

It is particularly important in a case control study that the exposure time was sufficient enough to show an association between the exposure and the outcome. It may be that the exposure period may be too short or too long to influence the outcome.

##### 10. Was appropriate statistical analysis used?

As with any consideration of statistical analysis, consideration should be given to whether there was a more appropriate alternate statistical method that could have been used. The methods section should be detailed enough for reviewers to identify which analytical techniques were used (in particular, regression or stratification) and how specific confounders were measured.

For studies utilizing regression analysis, it is useful to identify if the study identified which variables were included and how they related to the outcome. If stratification was the analytical approach used, were the strata of analysis defined by the specified variables? Additionally, it is also important to assess the appropriateness of the analytical strategy in terms of the assumptions associated with the approach as differing methods of analysis are based on differing assumptions about the data and how it will respond.

© JBI, 2020. All rights reserved. JBI grants use of these tools for research purposes only. All other enquiries should be sent to.

© JBI, 2020. All rights reserved. JBI grants use of these tools for research purposes only. All other enquiries should be sent to.

### JBI CRITICAL APPRAISAL CHECKLIST FOR ANALYTICAL CROSS SECTIONAL STUDIES

Reviewer Marco Antonio Delaye Martínez Date 20/04/21

Author Townsend L., Dowds J., O'Brien K., et al. Year 2020 Record Number 212_33413026

| | Yes | No | Unclear | Not applicable |
|---|---|---|---|---|
| 1. Were the criteria for inclusion in the sample clearly defined? | ☑ | ☐ | ☐ | ☐ |
| 2. Were the study subjects and the setting described in detail? | ☑ | ☐ | ☐ | ☐ |
| 3. Was the exposure measured in a valid and reliable way? | ☑ | ☐ | ☐ | ☐ |
| 4. Were objective, standard criteria used for measurement of the condition? | ☑ | ☐ | ☐ | ☐ |
| 5. Were confounding factors identified? | ☑ | ☐ | ☐ | ☐ |
| 6. Were strategies to deal with confounding factors stated? | ☑ | ☐ | ☐ | ☐ |
| 7. Were the outcomes measured in a valid and reliable way? | ☑ | ☐ | ☐ | ☐ |
| 8. Was appropriate statistical analysis used? | ☑ | ☐ | ☐ | ☐ |

Overall appraisal: Include ☑ Exclude ☐ Seek further info ☐

Comments (Including reason for exclusion)

No comments.

© JBI, 2020. All rights reserved. JBI grants use of these tools for research purposes only. All other enquiries should be sent to.

### JBI CRITICAL APPRAISAL CHECKLIST FOR COHORT STUDIES

Reviewer Fernández Chirino Luisa Date 15/04/2021

Author Myall, K.J., Mukherjee, B., et. al. Year 2021 Record Number 213_33433263_LFC

| | Yes | No | Unclear | Not applicable |
|---|---|---|---|---|
| 1. Were the two groups similar and recruited from the same population? | ☒ | ☐ | ☐ | ☐ |
| 2. Were the exposures measured similarly to assign people<br>3. to both exposed and unexposed groups? | ☒ | ☐ | ☐ | ☐ |
| 4. Was the exposure measured in a valid and reliable way? | ☒ | ☐ | ☐ | ☐ |
| 5. Were confounding factors identified? | ☒ | ☐ | ☐ | ☐ |
| 6. Were strategies to deal with confounding factors stated? | ☒ | ☐ | ☐ | ☐ |
| 7. Were the groups/participants free of the outcome at the start of the study (or at the moment of exposure)? | ☒ | ☐ | ☐ | ☐ |
| 8. Were the outcomes measured in a valid and reliable way? | ☒ | ☐ | ☐ | ☐ |
| 9. Was the follow up time reported and sufficient to be long enough for outcomes to occur? | ☒ | ☐ | ☐ | ☐ |
| 10. Was follow up complete, and if not, were the reasons to loss to follow up described and explored? | ☒ | ☐ | ☐ | ☐ |
| 11. Were strategies to address incomplete follow up utilized? | ☒ | ☐ | ☐ | ☐ |
| 12. Was appropriate statistical analysis used? | ☒ | ☐ | ☐ | ☐ |

Overall appraisal: Include ☒ Exclude ☐ Seek further info ☐

Comments (including reason for exclusion)

### JBI CRITICAL APPRAISAL CHECKLIST FOR TEXT AND OPINION PAPERS

Reviewer Marco Antonio Delaye Martinez Date 19/04/21

Author Wijeratne G., Crewther S. Year 2020 Record Number 214_33543150

| | Yes | No | Unclear | Not applicable |
|---|---|---|---|---|
| 1. Is the source of the opinion clearly identified? | ✔ | ☐ | ☐ | ☐ |
| 2. Does the source of opinion have standing in the field of expertise? | ✔ | ☐ | ☐ | ☐ |
| 3. Are the interests of the relevant population the central focus of the opinion? | ✔ | ☐ | ☐ | ☐ |
| 4. Is the stated position the result of an analytical process, and is there logic in the opinion expressed? | ✔ | ☐ | ☐ | ☐ |
| 5. Is there reference to the extant literature? | ✔ | ☐ | ☐ | ☐ |
| 6. Is any incongruence with the literature/sources logically defended? | ☐ | ☐ | ✔ | ☐ |

Overall appraisal: Include ☐ Exclude ✔ Seek further info ☐

Comments (Including reason for exclusion)

Se presenta una revision de una guía publicada por 'The Royal Australian Collegue for General Practioners" para la identificacion y manejo del Sindrome post-COVID 19 agudo. No se acepta por la naturaleza del articulo, sin embargo es util para discusión.

© JBI, 2020. All rights reserved. JBI grants use of these tools for research purposes only. All other enquiries should be sent to.

### JBI CRITICAL APPRAISAL CHECKLIST FOR COHORT STUDIES

Reviewer: Marco Antonio Delaye Martínez Date: ____

Author: Moreno-Perez et al Year: 2021 Record Number: 215_33450302

| | Yes | No | Unclear | Not applicable |
|---|---|---|---|---|
| 1. Were the two groups similar and recruited from the same population? | ✔ | ☐ | ☐ | ☐ |
| 2. Were the exposures measured similarly to assign people to both exposed and unexposed groups? | ✔ | ☐ | ☐ | ☐ |
| 3. Was the exposure measured in a valid and reliable way? | ✔ | ☐ | ☐ | ☐ |
| 4. Were confounding factors identified? | ☐ | ☐ | ☐ | ☐ |
| 5. Were strategies to deal with confounding factors stated? | ✔ | ☐ | ☐ | ☐ |
| 6. Were the groups/participants free of the outcome at the start of the study (or at the moment of exposure)? | ✔ | ☐ | ☐ | ☐ |
| 7. Were the outcomes measured in a valid and reliable way? | ✔ | ☐ | ☐ | ☐ |
| 8. Was the follow up time reported and sufficient to be long enough for outcomes to occur? | ✔ | ☐ | ☐ | ☐ |
| 9. Was follow up complete, and if not, were the reasons to loss to follow up described and explored? | ✔ | ☐ | ☐ | ☐ |
| 10. Were strategies to address incomplete follow up utilized? | ✔ | ☐ | ☐ | ☐ |
| 11. Was appropriate statistical analysis used? | ✔ | ☐ | ☐ | ☐ |

Overall appraisal: Include ☐ Exclude ✔ Seek further info ☐

Comments (Including reason for exclusion)

© JBI, 2020. All rights reserved. JBI grants use of these tools for research purposes only. All other enquiries should be sent to.

### JBI CRITICAL APPRAISAL CHECKLIST FOR COHORT STUDIES

Reviewer Fernández Chirino Luisa Date 15/04/2021

Author Weerahandi, H., Hochman, K., et. al. Year 2021 Record Number 216_33443703_LFC

| | Yes | No | Unclear | Not applicable |
|---|---|---|---|---|
| 1. Were the two groups similar and recruited from the same population? | ☒ | ☐ | ☐ | ☐ |
| 2. Were the exposures measured similarly to assign people<br>3. to both exposed and unexposed groups? | ☒ | ☐ | ☐ | ☐ |
| 4. Was the exposure measured in a valid and reliable way? | ☒ | ☐ | ☐ | ☐ |
| 5. Were confounding factors identified? | ☒ | ☐ | ☐ | ☐ |
| 6. Were strategies to deal with confounding factors stated? | ☒ | ☐ | ☐ | ☐ |
| 7. Were the groups/participants free of the outcome at the start of the study (or at the moment of exposure)? | ☒ | ☐ | ☐ | ☐ |
| 8. Were the outcomes measured in a valid and reliable way? | ☒ | ☐ | ☐ | ☐ |
| 9. Was the follow up time reported and sufficient to be long enough for outcomes to occur? | ☒ | ☐ | ☐ | ☐ |
| 10. Was follow up complete, and if not, were the reasons to loss to follow up described and explored? | ☒ | ☐ | ☐ | ☐ |
| 11. Were strategies to address incomplete follow up utilized? | ☒ | ☐ | ☐ | ☐ |
| 12. Was appropriate statistical analysis used? | ☒ | ☐ | ☐ | ☐ |

Overall appraisal: Include ☒ Exclude ☐ Seek further info ☐

Comments (including reason for exclusion)

### JBI CRITICAL APPRAISAL CHECKLIST FOR CASE REPORTS

Reviewer Marco Antonio Delaye Martínez Date 20/04/21

Author Iqbal P., Laswi B., Jamshaid M., et al. Year 2020 Record Number 217_33446625

| | Yes | No | Unclear | Not applicable |
|---|---|---|---|---|
| 1. Were patient's demographic characteristics clearly described? | ☑ | ☐ | ☐ | ☐ |
| 2. Was the patient's history clearly described and presented as a timeline? | ☑ | ☐ | ☐ | ☐ |
| 3. Was the current clinical condition of the patient on presentation clearly described? | ☑ | ☐ | ☐ | ☐ |
| 4. Were diagnostic tests or assessment methods and the results clearly described? | ☑ | ☐ | ☐ | ☐ |
| 5. Was the intervention(s) or treatment procedure(s) clearly described? | ☑ | ☐ | ☐ | ☐ |
| 6. Was the post-intervention clinical condition clearly described? | ☑ | ☐ | ☐ | ☐ |
| 7. Were adverse events (harms) or unanticipated events identified and described? | ☑ | ☐ | ☐ | ☐ |
| 8. Does the case report provide takeaway lessons? | ☐ | ☑ | ☐ | ☐ |

Overall appraisal: Include ☐ Exclude ☑ Seek further info ☐

Comments (Including reason for exclusion)

No se reporta suficiente evidencia para atribuir estas características a PACS.

© JBI, 2020. All rights reserved. JBI grants use of these tools for research purposes only. All other enquiries should be sent to.

### JBI Critical Appraisal Checklist for case control studies

Reviewer Marco Antonio Delaye Martínez Date 20/04/21

Author Ortelkl P., Ferrazzoli D., Sebastianelli L., et al. Year 2020 Record Number 218_33359928

| | Yes | No | Unclear | Not applicable |
|---|---|---|---|---|
| 1. Were the groups comparable other than the presence of disease in cases or the absence of disease in controls? | ☑ | ☐ | ☐ | ☐ |
| 2. Were cases and controls matched appropriately? | ☑ | ☐ | ☐ | ☐ |
| 3. Were the same criteria used for identification of cases and controls? | ☑ | ☐ | ☐ | ☐ |
| 4. Was exposure measured in a standard, valid and reliable way? | ☑ | ☐ | ☐ | ☐ |
| 5. Was exposure measured in the same way for cases and controls? | ☑ | ☐ | ☐ | ☐ |
| 6. Were confounding factors identified? | ☑ | ☐ | ☐ | ☐ |
| 7. Were strategies to deal with confounding factors stated? | ☑ | ☐ | ☐ | ☐ |
| 8. Were outcomes assessed in a standard, valid and reliable way for cases and controls? | ☑ | ☐ | ☐ | ☐ |
| 9. Was the exposure period of interest long enough to be meaningful? | ☑ | ☐ | ☐ | ☐ |

© JBI, 2020. All rights reserved. JBI grants use of these tools for research purposes only. All other enquiries should be sent to.

10. Was appropriate statistical analysis used? ☑ ☐ ☐ ☐

Overall appraisal: Include ☐ Exclude ☑ Seek further info ☐

Comments (including reason for exclusion)

No aporta informacion útil para los fines de la presente revisión sistemática.

© JBI, 2020. All rights reserved. JBI grants use of these tools for research purposes only. All other enquiries should be sent to.

### JBI CRITICAL APPRAISAL CHECKLIST FOR SYSTEMATIC REVIEWS AND RESEARCH SYNTHESES

Reviewer Fernández Chirino Luisa Date 15/04/2020

Author Tjan, L. H., Nagano, T., et. al. Year 2020 Record Number 219_33575497_LFC

| | Yes | No | Unclear | Not applicable |
|---|---|---|---|---|
| 1. Is the review question clearly and explicitly stated? | ☒ | ☐ | ☐ | ☐ |
| 2. Were the inclusion criteria appropriate for the review question? | ☒ | ☐ | ☐ | ☐ |
| 3. Was the search strategy appropriate? | ☒ | ☐ | ☐ | ☐ |
| 4. Were the sources and resources used to search for studies adequate? | ☒ | ☐ | ☐ | ☐ |
| 5. Were the criteria for appraising studies appropriate? | ☒ | ☐ | ☐ | ☐ |
| 6. Was critical appraisal conducted by two or more reviewers independently? | ☒ | ☐ | ☐ | ☐ |
| 7. Were there methods to minimize errors in data extraction? | ☒ | ☐ | ☐ | ☐ |
| 8. Were the methods used to combine studies appropriate? | ☒ | ☐ | ☐ | ☐ |
| 9. Was the likelihood of publication bias assessed? | ☒ | ☐ | ☐ | ☐ |
| 10. Were recommendations for policy and/or practice supported by the reported data? | ☐ | ☐ | ☒ | ☐ |
| 11. Were the specific directives for new research appropriate? | ☐ | ☐ | ☒ | ☐ |

Overall appraisal: Include ☐ Exclude ☒ Seek further info ☐

Comments (Including reason for exclusion)

Too general.

### JBI CRITICAL APPRAISAL CHECKLIST FOR COHORT STUDIES

Reviewer Marco Antonio Delaye Martínez Date 20/04/21

Author Brant-Zawadski M., Fridman D., Robinson P., et al. Year 2020 Record Number 220_33604403

| | Yes | No | Unclear | Not applicable |
|---|---|---|---|---|
| 1. Were the two groups similar and recruited from the same population? | ☑ | ☐ | ☐ | ☐ |
| 2. Were the exposures measured similarly to assign people to both exposed and unexposed groups? | ☑ | ☐ | ☐ | ☐ |
| 3. Was the exposure measured in a valid and reliable way? | ☑ | ☐ | ☐ | ☐ |
| 4. Were confounding factors identified? | ☑ | ☐ | ☐ | ☐ |
| 5. Were strategies to deal with confounding factors stated? | ☑ | ☐ | ☐ | ☐ |
| 6. Were the groups/participants free of the outcome at the start of the study (or at the moment of exposure)? | ☑ | ☐ | ☐ | ☐ |
| 7. Were the outcomes measured in a valid and reliable way? | ☑ | ☐ | ☐ | ☐ |
| 8. Was the follow up time reported and sufficient to be long enough for outcomes to occur? | ☑ | ☐ | ☐ | ☐ |
| 9. Was follow up complete, and if not, were the reasons to loss to follow up described and explored? | ☑ | ☐ | ☐ | ☐ |
| 10. Were strategies to address incomplete follow up utilized? | ☑ | ☐ | ☐ | ☐ |
| 11. Was appropriate statistical analysis used? | ☑ | ☐ | ☐ | ☐ |

Overall appraisal: Include ☐ Exclude ☑ Seek further info ☐

Comments (Including reason for exclusion)

El artículo no aporta información útil para los fines de la presemte revisión sistemática.

© JBI, 2020. All rights reserved. JBI grants use of these tools for research purposes only. All other enquiries should be sent to.
